# The genomic landscape of recurrent ovarian high grade serous carcinoma: the BriTROC-1 study

**DOI:** 10.1101/2022.10.21.22280992

**Authors:** P-S. Smith, T. Bradley, L. Morrill Gavarró, T. Goranova, D. Ennis, H. Mirza, D. De Silva, A.M. Piskorz, S. Al-Khalidi, C. Sauer, I-G. Funingana, M. Reinius, G. Giannone, L-A. Lewsley, J. Stobo, J. McQueen, G. Bryson, M. Eldridge, The BriTROC Investigators, G. Macintyre, F. Markowetz, J.D. Brenton, I.A. McNeish

## Abstract

The drivers of recurrence and resistance in ovarian high grade serous carcinoma (HGSC) remain unclear. We established BriTROC-1 to investigate the acquisition of resistance by collecting tumour biopsies from women with recurrent ovarian HGSC that had relapsed following at least one line of platinum-based chemotherapy. Patients underwent biopsy or secondary debulking surgery, with tumour samples fixed in methanol-based fixative. Normal and tumour DNA samples underwent tagged-amplicon panel sequencing. Tumour DNA underwent shallow whole genome sequencing (sWGS). Tissue microarrays (TMA), created from diagnosis samples, were stained for CD3, CD8, CD20 and FoxP3. 276 patients were recruited (209 platinum-sensitive, 67 platinum-resistant). Panel sequencing showed close concordance between diagnosis and relapse, with only 4 discordant cases, and no revertant mutations in *BRCA1* or *BRCA2* were identified in relapse samples. CN signatures were strongly correlated with immune cell infiltration. There was very strong concordance in copy number (CN) between diagnosis and relapse, with no significant difference in purity, ploidy or focal somatic CN alterations, even when stratified by platinum sensitivity or prior chemotherapy lines. Small increases in CN signature 3 and 7 exposure were observed between diagnosis and relapse across the whole cohort but were not present in matched sample pairs. Diagnosis samples from patients with primary platinum resistance had increased rates of *CCNE1* and *KRAS* amplification and CN signature 6 exposure. The HGSC genome is remarkably stable between diagnosis and relapse and acquired chemotherapy resistance does not select for common copy number drivers. We have identified new genomic events at diagnosis, including *KRAS* amplification and CN signature 6 exposure, that are associated with primary platinum resistance.

## Introduction

Ovarian high grade serous carcinoma (HGSC) is marked by ubiquitous TP53 mutation^1^, chromosomal instability (CIN), extensive copy number alterations^2-4^ and marked inter- and intra-patient genomic heterogeneity. This complexity has prevented effective precision medicine strategies. Consequently, the only molecular classification utilised in clinical practice is identification of tumours with defective homologous recombination (HR). The recent development of copy number signatures provides a robust analysis framework to quantify different types of CIN and to assign its extent and origins^5,6^.

Clinically, response rates to first line platinum-taxane therapy are high (65% by imaging criteria, 85% by CA125 criteria^7^). However, nearly all patients subsequently relapse despite the addition of maintenance therapy with PARP inhibition and/or bevacizumab^8-11^. The probability of response to further platinum-based chemotherapy decreases with each exposure and the vast majority of patients eventually acquire fatal chemotherapy resistance. Understanding how divergent evolution could underlie acquired resistance in HGSC is, therefore, of great importance.

Beyond rare revertant mutations in *BRCA1/2*^12^, and loss of *BRCA1* promoter methylation^13^, the drivers of recurrence and resistance remain unclear, and time interval since last platinum chemotherapy still remains the most useful predictor of subsequent treatment response in clinical practice. Assessing the prevalence of temporal heterogeneity and evolution in HGSC requires access to tumour material obtained at both diagnosis and relapse.

We established BriTROC-1, a UK-based ovarian cancer translational study, to investigate the acquisition of resistance in women with HGSC by collecting tumour biopsies from women with relapsed HGSC. We previously demonstrated that obtaining tumour biopsies in relapsed HGSC is safe and feasible, and that these biopsies yield sufficient DNA for genomic analyses^14^. We have used this well-annotated cohort to investigate whether recurrence and evolution of treatment resistance in HGSC is accompanied by clonal selection and enrichment in common driver copy number alterations. We have also utilised BriTROC-1 samples to identify markers present at time of diagnosis that predict early, platinum-resistant relapse.

## Patients and methods

### Study conduct

Details of the BriTROC-1 study and the first 220 patients were reported previously^14^. Briefly, BriTROC-1 was funded by Ovarian Cancer Action (grant number OCA_006) and sponsored by NHS Greater Glasgow and Clyde. Ethics/IRB approval was given by Cambridge Central Research Ethics Committee (Reference 12/EE/0349). All patients provided written informed consent – this included specific consent to biopsy, access to archival material, use of biopsy and archival material (and ascites if present) for genomic studies and testing of germline DNA for *BRCA1*/2 and other mutations. In addition, patients could optionally consent to a second biopsy upon disease progression and to be informed of germline *BRCA1*/2 analysis results. Researchers had no access to any patient-identifying data.

### Patients

Between 16/JAN/2013 and 05/SEP/2017, the study enrolled patients with recurrent ovarian high grade serous or grade 3 endometrioid carcinoma who had relapsed following at least one line of platinum-based chemotherapy. Other histological subtypes were only allowed in patients with known deleterious germline *BRCA1* or *BRCA2* mutations. Patients were classified as platinum sensitive (relapse ≥6 months since last platinum-based chemotherapy) or platinum resistant (relapse <6 months since last platinum-based chemotherapy) by recruiting sites at the time of study registration. All patients had to have disease amenable either to image-guided or other interventional (*e*.*g*. endoscopy, bronchoscopy) biopsy, or secondary debulking surgery. Access to archival diagnostic formalin-fixed tissue samples, or snap frozen tumour material if available, was also required for patient registration. Overall survival was calculated from the date of enrolment to the date of death or the last clinical assessment, with data cut-off at 01/APR/2018. Full inclusion and exclusion criteria are listed in **supplementary methods**.

Patients underwent biopsy (at least two cores, 14–16G biopsy needle) or secondary debulking surgery, with tumour samples fixed in methanol (UMFIX, TissueTek Xpress, Sakura)^15^. For patients undergoing secondary debulking or other interventional biopsies, 14–16G cores or a 1 cm^3^ piece of macroscopically identified tumour tissue were taken. All samples were shipped within 24 hours at ambient temperature to the University of Glasgow. There were no study-mandated therapies and all treatment after study entry was at the discretion of the treating oncologist.

### Tagged-amplicon sequencing

Normal and tumour DNA samples were assessed for single nucleotide variants and short indels using tagged-amplicon sequencing^16^. Paired-end sequencing was performed on either MiSeq and HiSeq 4000 Illumina platforms at 125 and 150nt respectively. A full set of amplicons is provided in **Table S1**.

### Amplicon read alignment

Alignment and post-alignment processing methods for sequenced amplicon reads are described in the **supplementary methods**.

### Germline variant calling

Germline short variants were called using CRUK-CI’s ampliconseq pipeline (https://github.com/crukci-bioinformatics/ampliconseq; v0.7.2)^5^ using GATK’s HaplotypeCaller (GATK version 3.8-0-ge9d806836) as the core variant calling algorithm. Further details are in **supplementary methods**.

### Tumour sample variant calling

Variant calling on tumour samples was performed using the cancer calling mode of Octopus (v0.7.2)^17^ with the exception of TP53 variants, which were as reported previously^5,14,15^. Further details are in **supplementary methods**.

### Short variant functional annotation

All non-TP53 variants were functionally annotated using Ensembl’s variant effect prediction (VEP) pipeline18 (v102.0). Variants were further refined using the molecular tumour board portal (MTBP)19. Variants labelled as ‘benign’ or ‘likely benign’ by MBTP were discarded. Further details are in **supplementary methods**.

### Shallow whole genome sequencing and alignment

Single-end 50bp read length sequencing was performed with 0.1X coverage target. Raw sequencing reads were aligned to GRCh37 (g1kp2) using bwa samse (version 0.7.17-r1188) and duplicates were marked using picard MarkDuplicates (version 2.9.5).

### Absolute copy number profile fitting

Relative copy number profiles for each sample were fitted using a bespoke absolute copy number pipeline (**code availability**). Initial relative copy number profiles were generated using a modified version of QDNAseq (**supplemental methods**)^20^. Relative copy number profiles were then fitted to an optimal ploidy and purity combination after GC and mappability correction, including a quantitative and qualitative quality control, to generate absolute copy number profiles using 30 kilobase bins (**supplemental methods**). A REMARK diagram is provided in **Figure S1**. Copy number event calling thresholds and quality metric assignment of ploidy change patients are detailed in the **supplemental methods**.

### Intra-tumour heterogeneity

Intra-tumour heterogeneity was estimated for fitted copy number profiles using a methodology analogous to that published previously^21^ with alterations described (**supplemental methods**).

### Copy number signature analysis

Copy number signatures were derived from absolute copy number profiles using the methodology described previously^5^, utilising the pre-computed signature-feature matrix to generate the signature-sample exposures for all samples with available absolute copy number profiles. Copy number signatures were compared using established methodologies, as well as a statistical framework to determine global shifts in signature abundance between groupings described herein (**supplemental methods**).

### Immunohistochemistry

Quantitative immunohistochemistry (IHC) data were available for a subset of diagnosis samples. Methodology for the generation and normalisation of IHC data is described in detail in the **supplemental methods**.

## Results

### Patients and samples

276 patients with relapsed ovarian high grade serous carcinoma (HGSC) were recruited between January 2013 and September 2017 from 14 UK centres. Clinical characteristics are summarised in **Table 1** and **Tables S2-4**, and the study scheme in **Figure 1A**. 209 patients were classified as platinum-sensitive and 67 as platinum-resistant and the median time from diagnosis to enrolment was 31.5 months (range 10–284, IQR 21–56). The median number of lines of prior chemotherapy was 1 (IQR 1–2) for platinum-sensitive patients and 2 (IQR 1–2) for platinum-resistant patients. Germline *BRCA1*/2 status was known for only 98/276 patients at time of enrolment: 22 had known pathogenic *BRCA1* mutations and 14 pathogenic *BRCA2* mutations. All treatment before and following study enrolment was at the discretion of treating oncologists (**Tables S5-6**), and median overall survival following enrolment was 35.6 months for the platinum-sensitive patients and 11.5 months for the platinum-resistant cohort (**Figure 1)**.

**Table 1:** BriTROC-1 patient demographic and disease characteristics.

| Characteristic | Platinum-sensitive<br>(N=209) | Platinum-resistant<br>(N=67) | Total<br>(N=276) |
| --- | --- | --- | --- |
| Median age at study entry (range), years | 66 (31–85) | 63 (24–81) | 65 (24–85) |
| Median time since diagnosis (range), months | 32·5 (10·2–284·2) | 23·8 (5·1–184·2) | 31·5 (5·1–284·2) |
| Histology, N (%) |  |  |  |
| High grade serous | 200 (95·7) | 66 (98·5) | 266 (96·4) |
| Grade 3 Endometrioid | 6 (2·9) | 0 | 6 (2·3) |
| Carcinosarcoma | 1 (0·6) | 0 | 1 (0·4) |
| Missing | 2 (1·0) | 1 (1·5) | 3 (1·1) |
| Prior treatment |  |  |  |
| Median number of regimens (range) | 1 (1–5) | 2 (1–12) | 1 (1–12) |
| 1, N (%) | 148 (70·8) | 19 (28·4) | 167 (60·5) |
| 2, N (%) | 39 (18·7) | 29 (43·3) | 68 (24·6) |
| 3, N (%) | 12 (5·7) | 8 (11·9) | 20 (7·2) |
| 4, N (%) | 5 (2·4) | 3 (4·5) | 8 (2·9) |
| >4, N (%) | 3 (1·4) | 7 (10·4) | 10 (3·6) |
| Data missing | 2 (1·0) | 1 (1·5) | 3 (1·1) |

**Figure 1.**
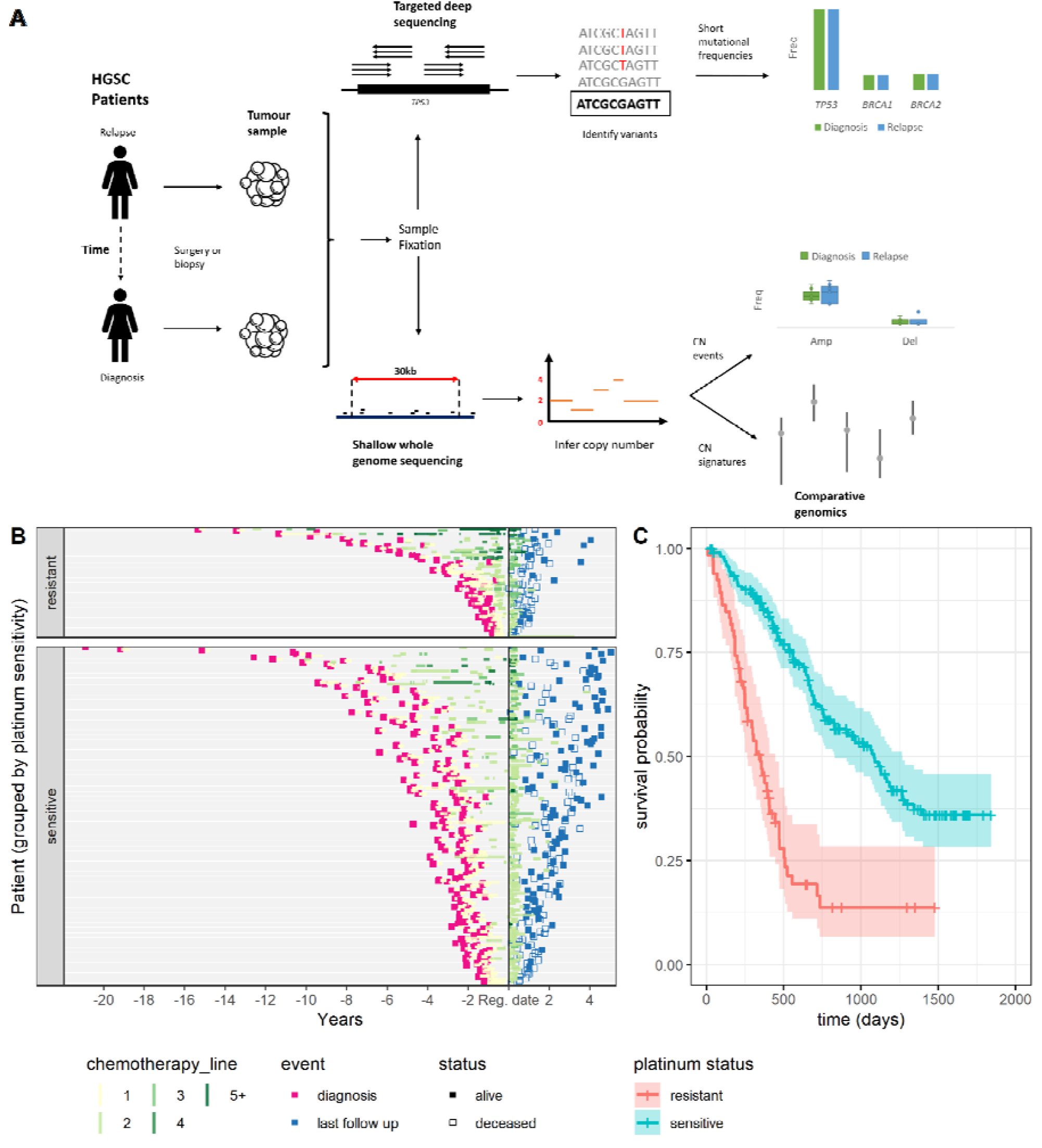
*BriTROC-1* Study scheme, recruitment timelines and overall survival. **A** - Visual representation of study recruitment and data analysis pipeline. A workflow for the procedures, experiments and analyses conducted as part of this study. Patient samples were processed in parallel for short variant and copy number analysis. **B** - Timeline representing the clinical course and outcomes of BriTROC-1 patients (n = 269). Each row represents a study participant; Black vertical line represents the time of recruitment. Initial primary tumour diagnosis date is shown in pink and last follow up date at study end is shown in blue. Participants who died are shown as an unfilled point. Orange segments represent chemotherapy treatments and timescale of treatments, each subsequent treatment course is shown up to four. Treatment courses of 5 or more are aggregated. **C** - Kaplan-Meier survival analysis for all patients enrolled into BriTROC-1 from the point of study entry stratified by platinum status. Crosses indicate right-censored data. The shaded area indicates a 95% confidence interval.

### Germline and somatic short variant analyses

Germline variants in key genes in the homologous recombination repair pathway were assessed in 228/276 patients (**Figure S2**). Pathogenic mutations in *BRCA1* and *BRCA2* were identified in 25 (11%) and 27 patients (12%) respectively. Pathogenic mutations in non-BRCA HRD genes were identified in four other patients, one each of RAD51B, *RAD51C, RAD51D, PALB2* and *BRIP1* (0.5% mutation rate for all). Somatic sequencing on samples from 264 patients identified a pathogenic TP53 mutation in 252 (95%) (**Figure S3**), whilst the somatic mutation rate for *BRCA1* and *BRCA2* was estimated as 1% and 5% respectively. Comparison of matched tumour sample pairs (n=134 pairs) showed very close concordance between diagnosis and relapse, with new mutation events being rare (**Figure 2** and **Figure S4, S5**). No revertant mutations were identified in *BRCA1* or *BRCA2* in relapse samples.

**Figure 2.**
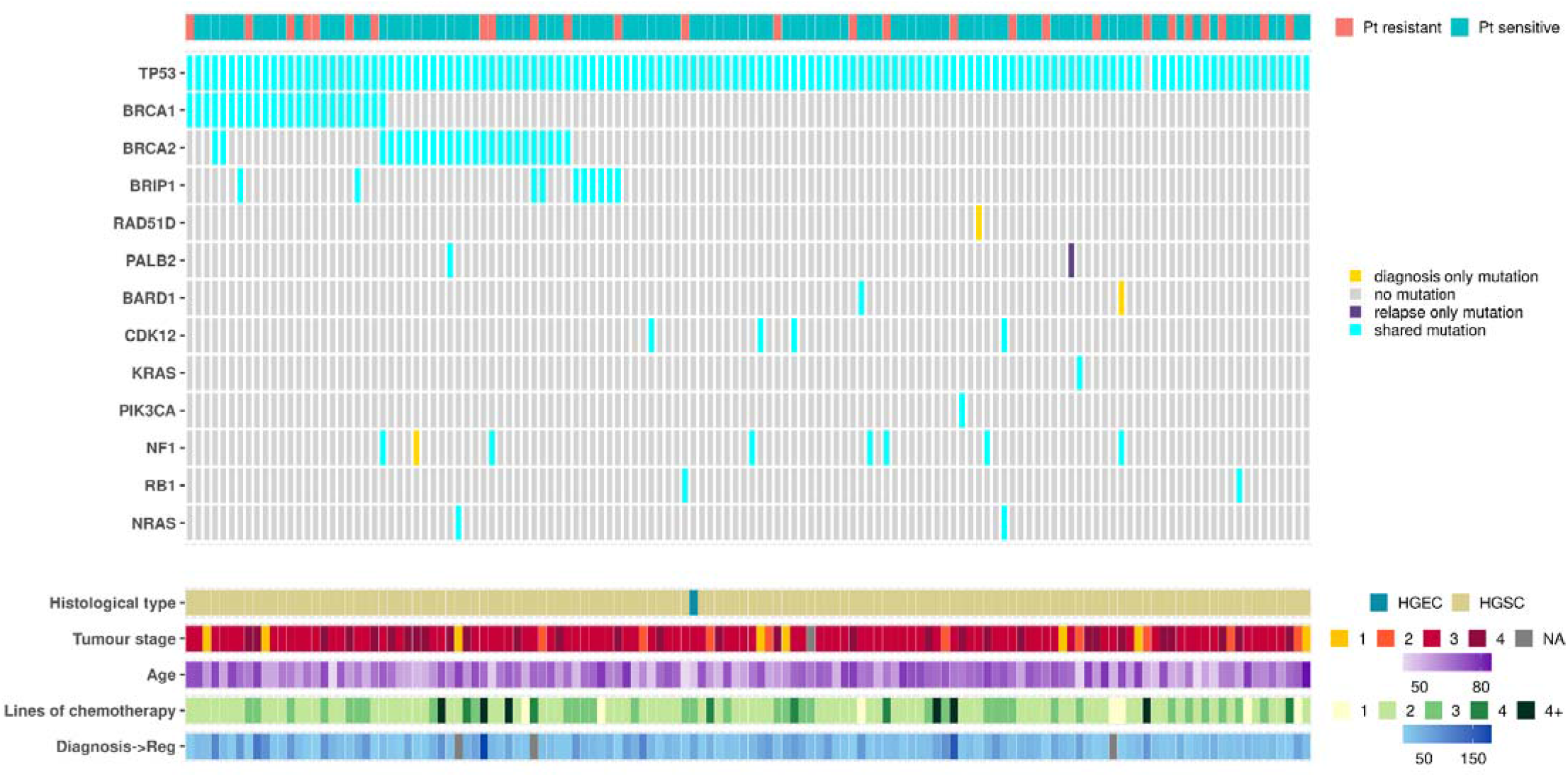
Paired short-variant analysis from 134 diagnosis/relapse sample pairs. Each column represents one patient and variants are not classified as either somatic or germline. Three cases had diagnosis-only mutations: patient 51 (NF1 c.6643-3del), patient 101 (RAD51 p.Gln175Ter) and patient 163 (BARD1 p.Val767Ala). Patient 139 had a relapse-only mutation in PALB2 (p.Lys1163Glu). Abbreviations: Pt - Platinum. HGSC - High grade serous carcinoma, HGEC - High grade endometrioid carcinoma. Tumour stage refers to stage at the time of diagnosis; Age refers to age at study entry; Lines of chemotherapy refers to the number of lines of chemotherapy prior to enrolment into BriTROC-1. Diagnosis-Reg denotes the interval (in months) between diagnosis and registration into BriTROC-1.

### Copy number alterations between diagnosis and relapse cohorts

We assessed genome-wide copy number and found strong concordance between diagnosis and relapse (**Figure S6**). Subtracting the median copy number profile at diagnosis from that at relapse generated a flat profile with only 6.1% (5075/78532) bins showing significant alteration (**Figure 3A**). Moreover, the median gain was only 0.06 copies across these regions (IQR 0.05-0.10). The significantly altered bins overlapped with 322 protein coding genes, of which 59 were identified as cancer-related22. Gene ontology enrichment23 analysis identified 111 significantly overrepresented biological processes after false discovery rate correction, one of which was regulation of MAP kinase activity (*q*=0.036). However, reactome^24^ analysis did not identify an overrepresentation of any given pathway or biological process in these genes.

**Figure 3.**
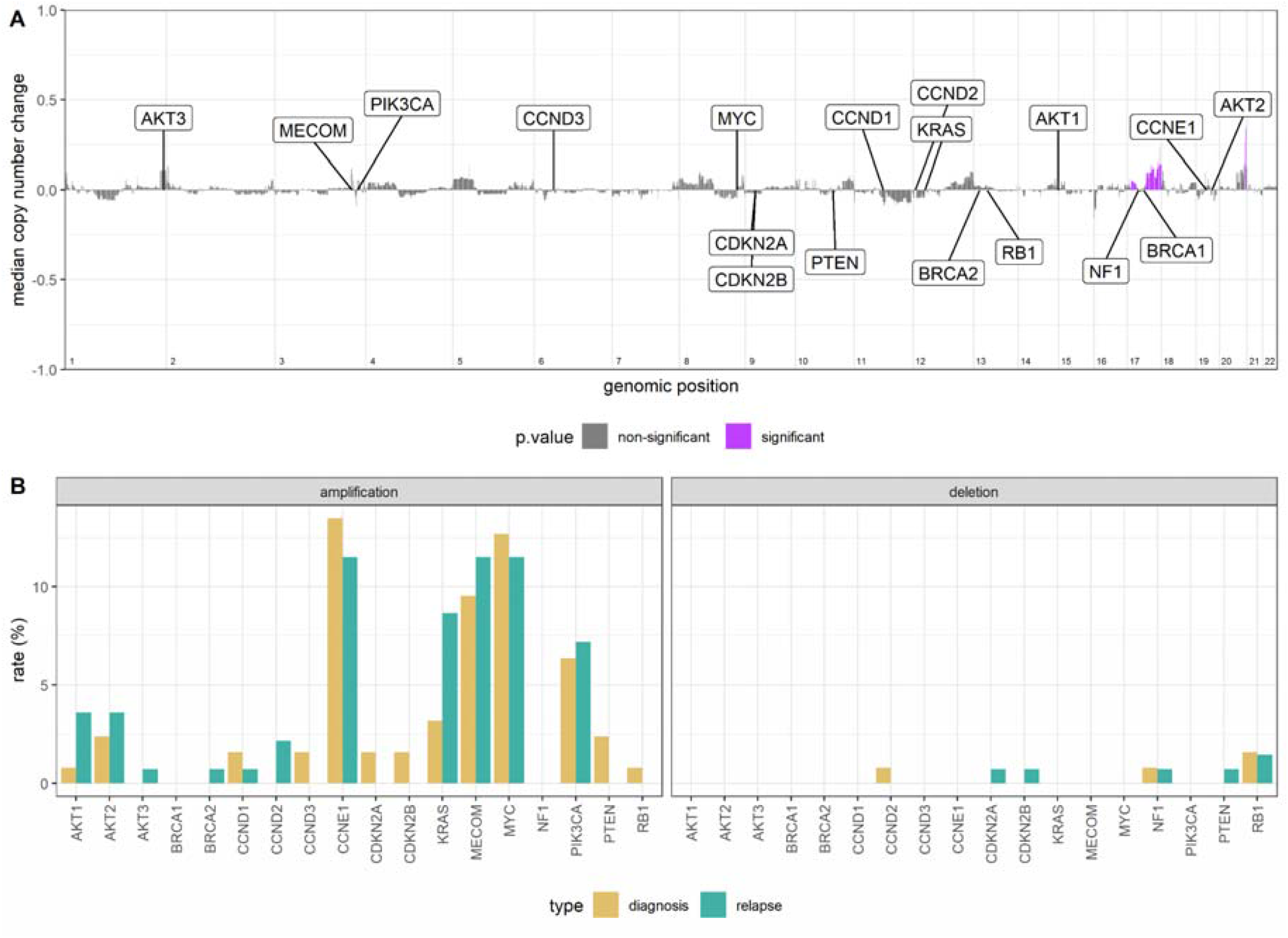
Copy number alterations. **A** - Genome-wide subtraction plot to visualise copy number differences between diagnosis and relapse in 47 matched pairs. Positive values indicate an increase in copy number state and a negative value indicates a decrease in copy number state at relapse. Purple peaks represent genomic regions (30kb bins) with significantly altered copy number between the diagnosis and relapse tumours (chromosome-specific FDR-corrected Mann-Whitney test). Eighteen key and frequently altered genes in HGSC are highlighted. **B** - Rates of focal amplification and deletion for the same eighteen genes (n = 126 and n = 139, diagnosis and relapse samples, respectively).

We did not identify a co-ordinated difference in purity, ploidy, or copy number segments between diagnosis and relapse, including when stratifying by platinum sensitivity (**Figure S7-S9**). The total number of copy number events and features5 were also broadly consistent between diagnosis and relapse (**Figure S10-11**). We identified nine patients who showed ploidy changes between diagnosis and relapse (**Figure S8**; increased in 7, decreased in 2). However, these nine patients did not differ from the remainder of the cohort in age, platinum sensitivity or lines of prior chemotherapy (*p*=0.052; Mann-Whitney U test, *p*=0.69; Fisher’s exact test, and *p*=0.86; Mann-Whitney U test, respectively). We also observed no significant differences in chromosomal arm-level events. A higher amplification rate at only two cytobands, 11q13.5 and 5p15.2, was identified at diagnosis compared to relapse, although neither of these remained significant after multiple testing correction (**Figure S12**).

We next assessed focal amplification and deletion of 18 genes that are frequently altered in HGSC^3,25^ (**Table S7, Figure S13**) and found no significant changes between diagnosis and relapse. The rate of *KRAS* amplification increased from 3.3% (4/122) at diagnosis to 9.4% (12/127) at relapse, although this did not reach statistical significance (p=0.07; Fisher’s exact test) (**Figure 3B, Figure S13**). When paired samples were analysed, we also found no significant difference in absolute copy number counts of these genes, including *CCNE1* (**Figure S14**). Rates of amplification or deletion of these genes at relapse did not increase with greater lines of prior therapy, in either paired or unpaired analyses (**Figure S15**), nor when stratified by platinum status (sensitive vs resistant). Absolute *CCNE1* gene copy number was marginally lower in relapse samples from platinum-resistant patients and *BRCA1* marginally higher in relapse sensitive patients (p=0.03 and p=0.02, Mann-Whitney) (**Figure S16-S17**), though not after multiple testing corrections.

### Tumour heterogeneity

Intra-tumour heterogeneity (ITH) was estimated from the copy number profiles using the average segment distance from integer state across a given genome (**Figure S18**). There was no statistically significant difference in ITH between diagnosis and relapse in either paired (*p*=0.27, Wilcoxon signed-rank) and unpaired (*p*=0.18, Mann-Whitney) analyses (**Figure S19**). From paired samples, we found no difference in ΔITH, an ITH change metric, by platinum sensitivity (*p*=0.11, Mann-Whitney U test), patient age at diagnosis (*p*=0.32, Kendall’s rank test), time between diagnosis and study entry (*p*=0.71, Kendall’s rank test), or number of prior lines of chemotherapy (*p*=0.85, Mann-Whitney U) (**Figure S20**).

### Copy number signatures

Copy number signatures for all diagnosis and relapse samples^5^ were highly consistent with the initial analysis of 118 BriTROC-1 samples (**Figure S21**) and stable when compared to The Cancer Genome Atlas (TCGA)26 and Pan-Cancer Analysis of Whole Genomes study (PCAWG)27 cohorts (**Figure S22**).

We observed small increases in exposure to signatures 3 (s3) and 7 (s7) between diagnosis and relapse across the whole cohort (**Figure 4, Figure S23**), but were not present in matched sample pairs (**Figure 5A, B**). Given the compositional nature of copy number signature data (*i*.*e*., they add to 1 in all samples), we designed a model to test for global differential abundance of copy number signature exposures between matched diagnosis and relapse groups (**supplementary methods**). Using a partial isometric Log-ratio (ILR) model with correlated random effects, we identified a coordinated shift in s5 exposure, indicating differential abundance between diagnosis and relapse (*p*=0.003, Wald test) (**Figure 5C**). However, s5 is associated with chromothriptic-like events and subclonal changes that may partially reflect different fixation methods^15^. Most diagnosis samples were formalin-fixed, whilst relapse/study-entry samples were processed in a methanol-based fixative. We hypothesised that s5 was capturing differential fixation artefact rather than a true difference in mutational processes. Repeating the analysis whilst excluding s5 indicated no significant difference in signature abundances between diagnosis and relapse samples (p=0.052, Wald test).

**Figure 4.**
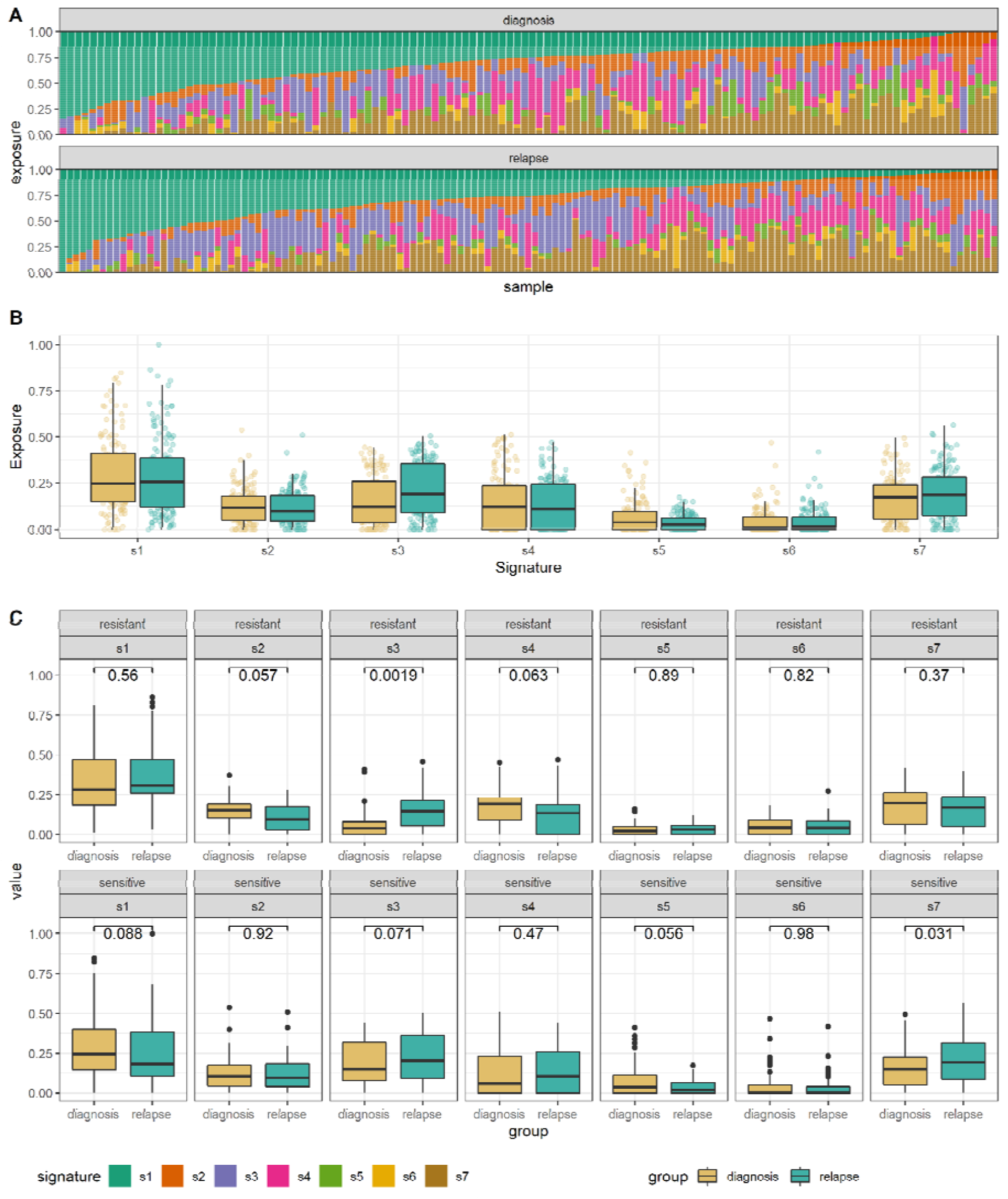
Unpaired copy number signatures. **A** - Copy number signature spectrum across all samples. Stacked bar plots do not align between diagnosis and relapse groups due to different sample numbers (n = 126 and n = 139, diagnosis and relapse, respectively). **B** - Box plot demonstrating the unpaired copy number signature exposures between the diagnosis and relapse tumours (n = 126 and n = 139, diagnosis and relapse, respectively). **C** - Boxplot demonstrating copy number signature distributions in unpaired copy number between diagnosis and relapse tumours, stratified by platinum status (n = 30, n = 36, n = 94, and n = 101, diagnosis resistant, relapse resistant, diagnosis sensitive, and relapse sensitive, respectively).

**Figure 5.**
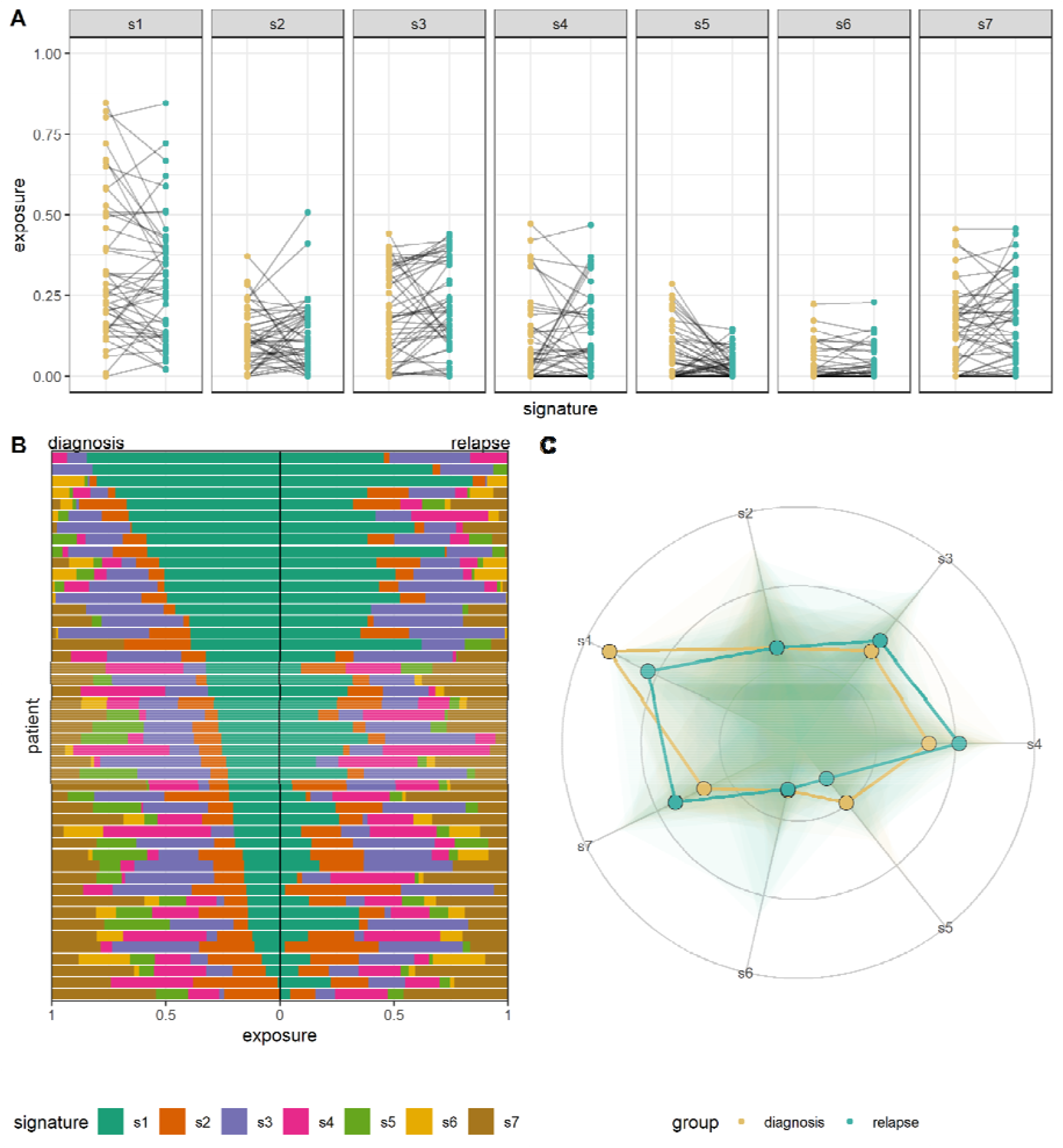
Paired copy number signatures. **A** - Paired copy number signature exposures between the diagnosis and relapse tumours in 47 paired samples. **B** - Copy number signature spectrum of paired copy samples. Each horizontal bar represents one patient, ranked by signature 1 exposure in the diagnosis sample. **C** - Radar plot of diagnosis and relapse copy number signature exposures. The points represent beta values calculated for the ILR of each copy number signature after inverse partial ILR transformation. The lines show the sample-level distribution of copy number signature exposure across the seven signatures.

### Copy number changes in different tissue sites in HGSC

To address whether metastasis to specific anatomical locations was associated with discrete genomic alterations, we first assessed the copy number of the eighteen key genes in different tissue sites across all samples (both diagnosis and relapse). Absolute copy number counts of *AKT1, BRCA1* and *PIK3CA* were significantly different between tissues (*p*=0.004, *p*=0.02, and *p*=0.03, respectively, one-way ANOVA). However, after post-hoc testing and multiple testing correction, the only significant difference was *AKT1* copies between pelvic deposits and tissues classified as ‘other’ (**Figure S24A**). Rates of amplification and deletion were also consistent across tissues, with the exception of *PIK3CA* amplification (*p*=0.02, Fisher’s exact), which was more frequent in lymph nodes compared to peritoneal deposits (**Figure S24B**). Differences in ITH were not statistically significant across tissue sites (**Figure S24C**), and copy number signatures were also broadly consistent, although s1 exposures in pelvic and lymph node deposits were statistically significant after correction (*p*=0.01 and *q*=0.02, one-way ANOVA and Bonferroni-corrected Tukey’s test, respectively) (**Figure S24D, Table S8**).

When comparing diagnosis and relapse, we were restricted to abdominal, pelvic, and peritoneal deposits due to sample number limitations. Absolute copy number counts in the eighteen key genes were consistent between diagnosis and relapse across different tissue sites and no significant differences in copy number counts were found. Similarly, copy number signatures between diagnosis and relapse samples were also broadly consistent across different tissues, with the exception of s2 in abdominal deposits (p=0.002, Mann-Whitney U). Lastly, ITH, when stratified by diagnosis and relapse, was not statistically different across differing tissues of origin (**Figure S25**).

### Patient specific alterations between diagnosis and relapse HGSC

We assessed patient-specific gene alterations between diagnosis and relapse and determined if patient subgroups existed within the larger cohort-level analysis. Hierarchical clustering of the top 20% gene loci with the greatest variance in copy number between diagnosis and relapse identified a heterogeneous pattern of copy number alterations between patients. No obvious groupings were identified on the basis of specific genes or concordant changes with clinical features (**Figure 6A**). Focal genes that are frequently altered or of clinical relevance did not determine any clustering of patients. However, we did observe significantly correlated shifts in copy number changes between diagnosis and relapse in a subset of these genes, although the magnitude of copy number change was low (**Figure 6B, Figure S26**). Moreover, attempts to identify patient clusters using k-means clustering failed to identify any meaningful clusters (**Figure S27**).

**Figure 6:**
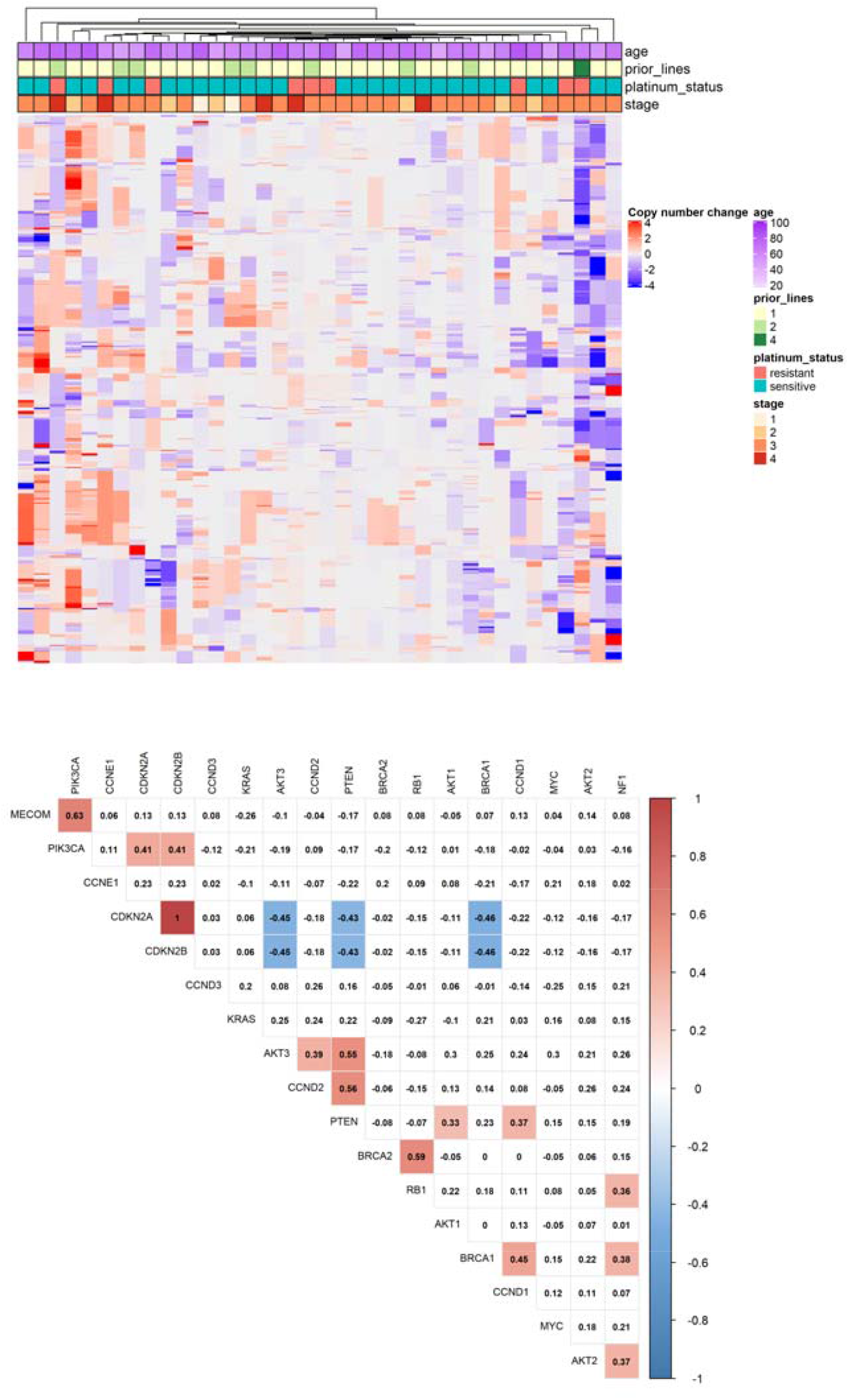
Gene change heatmap and correlation matrix. **A** - Heatmap showing the direction of copy number change for gene loci between 47 paired diagnosis and relapse samples at patient level. Rows correspond to the top 20% most variable loci (corresponding to 3623 genes) and columns correspond to patients. Cell colouration represents the change in copy number for a given locus (mean value in the relapse sample minus mean value in the diagnosis sample). B - Correlation matrix of 18 frequently altered genes. Correlation plot displays any Spearman’s rank correlation between copy number changes (across diagnosis and relapse) in different genes, where a positive value indicates a positive correlation and negative value indicates a negative correlation.

Looking specifically at the 18 frequently altered genes, we observed minimal differences between diagnosis and relapse across all paired samples when normalised for ploidy (**Figure S28**). However, patient-specific analysis did demonstrate biologically interesting genomic alterations. For example, BRITROC-65 and BRITROC-37 demonstrated marked gains of *KRAS* at relapse; BRITROC-36 had a 16 copy loss of *AKT2* and 6 copy loss of *NF1* at relapse, and BRITROC-242 and BRITROC-246 lost >10 copies of *CCNE1* between diagnosis and relapse (**Figure S29 - patient vignettes**). Cases with extreme copy gains and losses present across all gene loci were more frequent in platinum sensitive patients than resistant (**Figure S30**).

### Primary platinum resistance

Given the overall stability of genomic changes between diagnosis and relapse, we investigated genomes at diagnosis for features that might be associated with poor outcome, specifically focussing on the 21 patients with primary platinum resistance (defined as relapse <6 months after completion of first-line treatment). We found higher rates of *CCNE1* and *KRAS* amplification at diagnosis in these patients compared to all other patients (58.3 % vs. 8.77%, *p*=0.002; 25% vs. 0.8%, *p*=0.05, respectively, FDR-adjusted fisher’s exact) as well as higher absolute *CCNE1* copy number counts (median 9.92 vs. 2.73 copies, p=0.03 FDR-adjusted Mann-Whitney) (**Figure 7A, Figure S31**). Intriguingly, diagnosis samples from these 21 patients also had higher absolute *BRCA2* copy number, although the magnitude of change was small (median 2.57 vs 1.98 copies, *p*=0.02, (**Figure S31**). These differences were also evident at relapse (**Figure S31**). The patients with primary platinum resistance also had significantly different CN signature exposures at diagnosis with significantly lower exposure to s3 and higher s6 exposure than samples from all other patients (*p*=0.003, Wald test) (**Figure 7B-E**).

**Figure 7.**
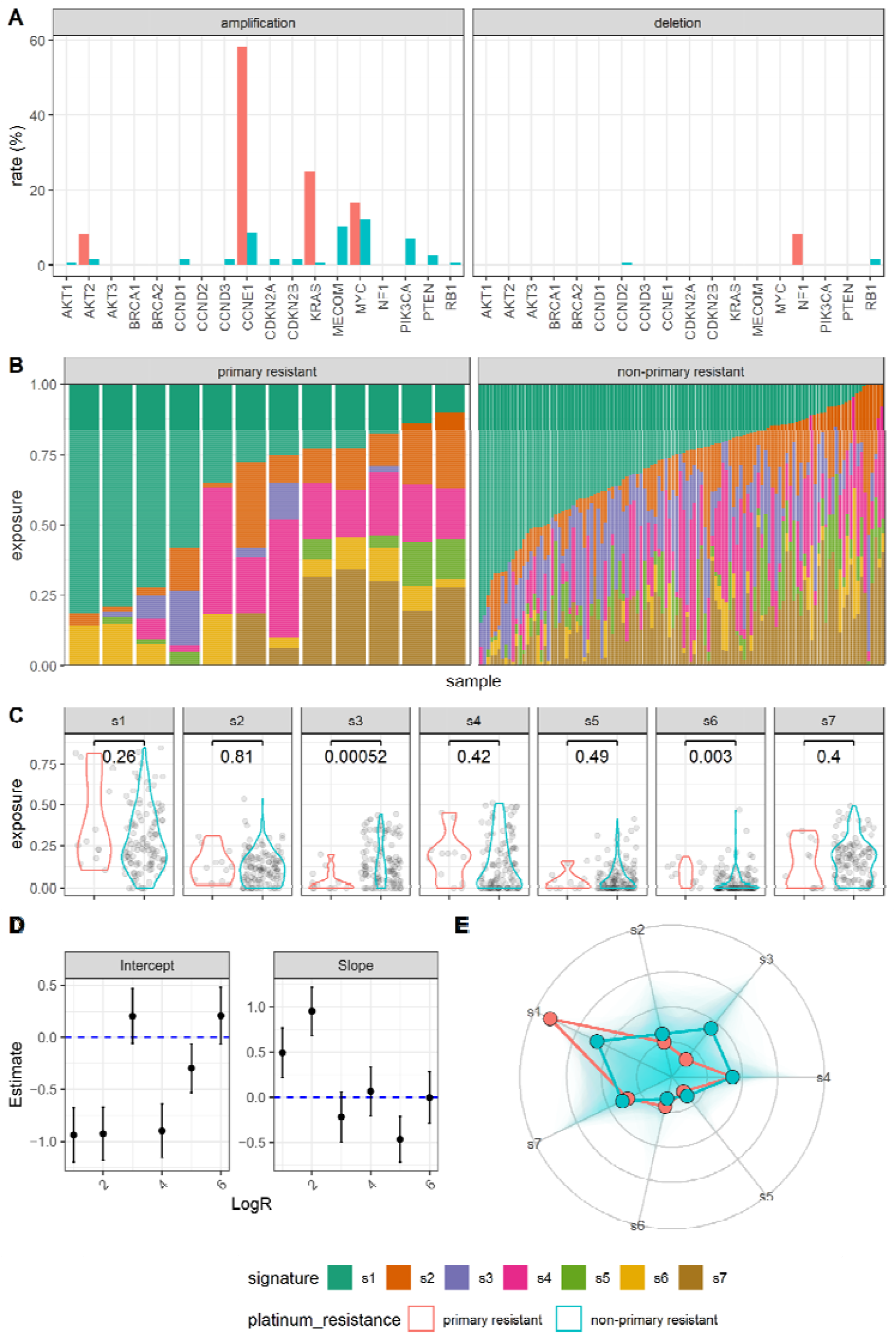
copy number signatures stratified by primary platinum-based treatment resistance. **A** - Copy number alteration rates for 18 frequently altered genes in diagnosis samples from BriTROC-1 patients with primary platinum resistance (n=12) compared to all others (non-primary platinum resistance, n=114). **B** - Copy number signature exposures and **C** - Copy number signature distributions in diagnosis samples from BriTROC-1 patients with primary platinum resistance (n = 12), compared to non-primary platinum resistance (n=114). **D** - Beta slope and intercept for primary platinum resistant diagnosis samples versus all other diagnosis samples. **E -** Radar plot showing distribution of ratio between copy number signature exposures between primary resistant and non-primary resistant patients. The points represent beta values calculated for the ILR of each copy number signature after inverse partial ILR transformation.

### Immune correlations

Finally, we investigated whether copy number alterations and chromosomal instability were associated with features of the tumour immune environment, using quantitative IHC for CD3 and CD8. There was statistically significant inter-marker correlation between CD3 and CD8 across tumour and stromal tissue independently (✉. = 0.73, 0.73, and 0.72, Spearman’s rank; all, stromal, and tumour tissue, respectively) (**Figure S32**). CD3 and CD8 were shown to correlate strongly with copy number signatures. Specifically, s1 was negatively correlated with CD3 and CD8, s3 and s7 were both positively correlated with CD3 and CD8, and s6 was negatively correlated with CD3 (**Figure 8**; Spearman’s rank). These correlations were present in both stromal and tumour tissues, with a stronger signal identified in the stromal tissue.

**Figure 8.**
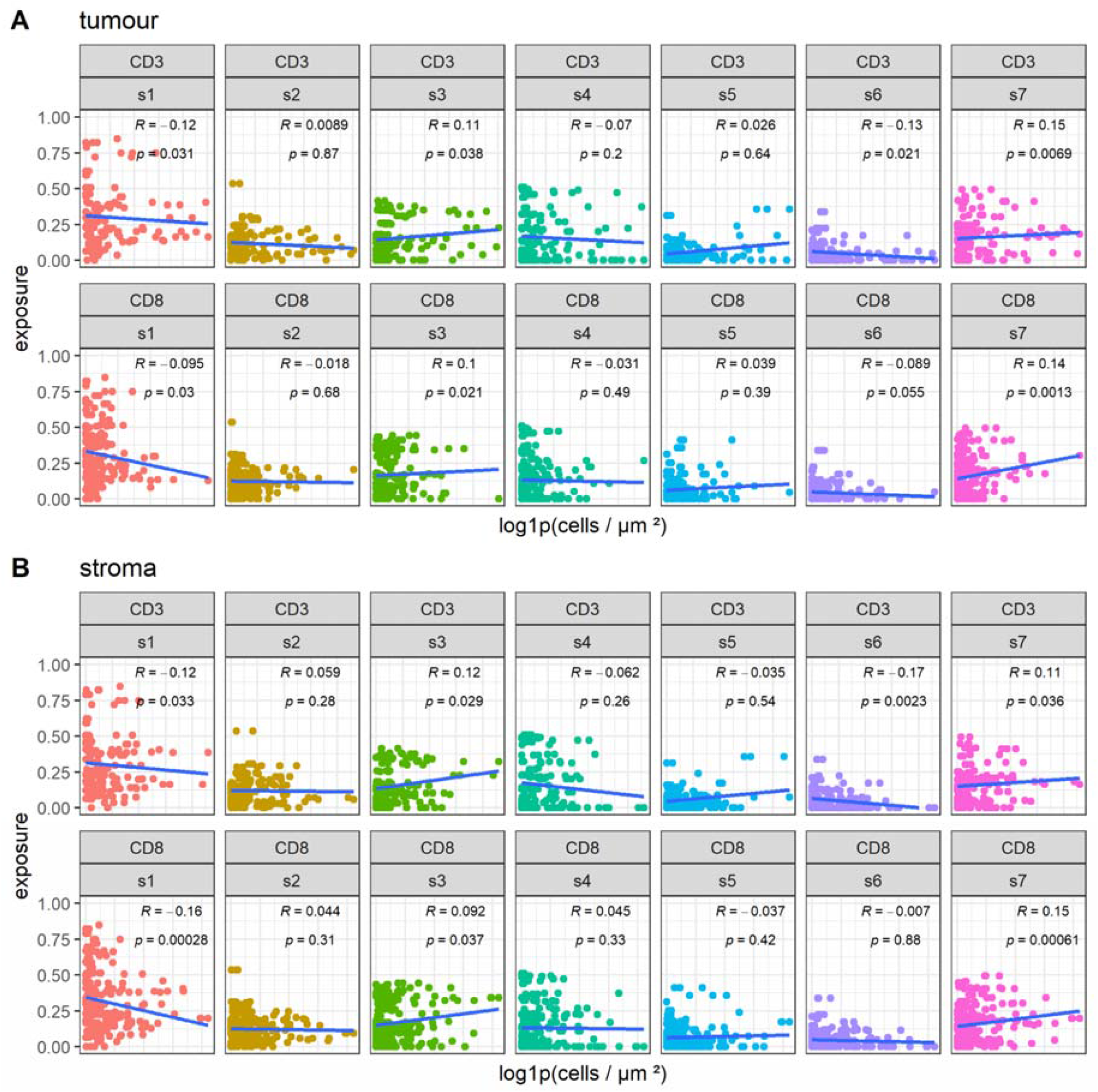
IHC-derived Immune marker signature correlation plots. Stratified correlations plot for IHC-derived CD3+ and CD8+ cell densities against copy number signatures in **A** - tumour and **B** - stroma. Solid blue lines are fitted linear regressions, R values are the Kendall tau correlation coefficient and associated p values for the significance of the given correlation. Calculated p values may not accurately estimate the strength of the correlation for each immune marker versus copy number signature due to the compositional nature of copy number signatures and intra-patient dependencies associated with the immune marker image data.

## Discussion

We used shallow whole genome sequencing and deep sequencing of a targeted gene panel to analyse samples from BriTROC-1, the largest prospective study yet of relapsed HGSC genomes. In a cohort of 276 cases, there are strikingly few recurrent changes between diagnosis and relapse. We identified only four cases with changes in single nucleotide variants/indels between diagnosis and study entry in a targeted panel of relevant HGSC genes, and no revertant mutations in *BRCA1* or *BRCA2* were detected in our population. Copy number profiles also showed minimal changes – we did observe selection for *KRAS* amplifications at relapse, although this did not reach statistical significance, suggesting that coordinated changes in driver CNA are infrequent. Copy number (CN) signatures did not show any statistically significant shift in exposures, suggesting that the mutational processes in HGSC either remain consistent or do not drive divergent patterns of CN alterations between diagnosis and relapse. We also did not identify recurrent changes in ploidy or intra-tumoural heterogeneity. These data strongly indicate that the major copy number features of HGSC, as determined by our assays, is stable between diagnosis and relapse, and does not explain recurrence and acquired chemotherapy resistance.

The large size of the BriTROC-1 cohort and the stability of genomic changes allowed us to identify prognostic markers at diagnosis. In the patients with primary platinum-resistance, we found significantly higher rates of *CCNE1* and *KRAS* amplification at diagnosis than the remainder of the cohort. Although *CCNE1* amplification has been identified previously as a poor prognostic feature in HGSC^28,29^, this is the first identification of amplification of non-mutated *KRAS* as a marker of primary platinum resistance. In addition, CN signature 3 (s3), associated with defective HR^5^, was significantly lower and s6 significantly higher in the primary resistant population at diagnosis. The absence of defective HR is associated with inferior outcome following platinum-based chemotherapy^10,30^. Exposure to s6, marked by high copy number states, is greater in cases with *CCNE1* amplification and mutations in the PI3K/AKT pathway5, which reinforces the negative prognostic implications of *CCNE1* amplification.

We also show that CN signatures have strong associations with specific immune microenvironment at diagnosis. The presence of intra-tumoural T cells^31^, in particular CD8+ cells^32^, is strongly positively prognostic in HGSC. However, the tumour-autonomous drivers of immune cell infiltration remain elusive. Here, we found positive correlations between both CD3 and CD8 cell infiltration with increasing exposure to s3 and s7, and negative correlations with increasing s1. Previous studies have certainly identified an association between *BRCA1* loss and higher intra-epithelial CD8+ numbers33,34, whilst tumours marked by fold-back inversions contain fewer CD8+ cells35. In primary platinum-resistant patients, we observed higher signature exposure to s1, which is strongly associated with breakage-fusion bridge mutational processes and overlaps with cases with foldback inversions. Thus, our data extend known links between tumour genotype and immune phenotype and support the hypothesis that tumour autonomous features strongly shape the immune microenvironment. However, detailed analyses of the interplay between immune cell populations and tumour genotype in the BriTROC-1 cohort are ongoing.

Previous studies of HGSC evolution have examined multiple samples taken at primary surgery, revealing significant intra-patient heterogeneity with evidence of diverse metastatic processes and patterns of clonal expansion^36-38^. Analyses before and after neoadjuvant chemotherapy have also failed to observe recurrent chemotherapy-induced mutations, but did suggest copy number alterations, including SIK2 amplification^39^. We did not identify any cases with SIK2 amplification or any difference in SIK2 absolute copy number (SIK2 median copies; diagnosis = 2.04 versus relapse = 1.99, p=0.41, Mann-Whitney U). The OCTIPS consortium investigated 31 matched HGSC sample pairs (diagnosis and relapse, with 24/31 cases analysed at first relapse) with whole exome sequencing and SNP profiling40. Again, there were no consistent changes across pairs and the changes observed did not correlate with clinical characteristics. Similarly, there were no recurrent primary-or relapse-only unique somatic CNA in these pairs.

Broader studies examining the progression of tumours from diagnosis to relapse and/or metastasis have identified patterns of genomic alterations that are cancer-type dependent. Comprehensive analysis of genomic alterations in unpaired HGSC samples in the MSK-MET study also found no statistically different copy number alterations41. This is in stark contrast to other cancer types, including melanoma, which demonstrate distinct changes in SNV mutational signatures and large increases in copy number events, aneuploidy and whole genome duplications between diagnosis and end stage or metastatic disease^42,43^. Similarly, renal clear cell carcinoma demonstrates an evolutionary bottleneck followed by expansive increase in copy number alterations between primary and metastatic sites^44,45^.

Our unpaired analyses suggested small but significant increases in s3 and s7 at relapse. The apparent increase of two signatures at relapse is consistent with our previous quantification of genome-wide LOH as a marker of defective HR46. In the ARIEL2 study, we observed a general increase in LOH between diagnosis and relapse that was sufficient to change classification from HR-proficient at diagnosis to HR-defective at relapse in approximately 15% cases^46^. Together, these data suggest an overall increase in CN damage as HGSC progresses, which may reflect both time-dependent and platinum-induced change.

There are several important caveats to our data. Firstly, BriTROC-1 enrolled women who had relapsed following prior therapy and who were well enough to undergo surgery or an image-guided biopsy, potentially biassing the study towards those with good prognosis. Nonetheless, overall survival following study enrolment was broadly consistent with previous clinical studies of chemotherapy in these populations47-50, suggesting that our population responded as expected.

Secondly, we had to utilise routinely-collected formalin-fixed, paraffin-embedded diagnostic samples. This, combined with small biopsies at relapse, meant that the number of high quality matched sample pairs was limited, thereby reducing our statistical power and our ability to observe potentially important events. This limitation was most apparent in the analysis of CN signatures, where we observed a significant difference in global abundance of s5 between diagnosis and relapse: s5 is more prevalent in samples from FFPE sources15 and we hypothesise that its differential abundance reflects a degree of fixation artefact.

Thirdly, we deliberately restricted our targeted sequencing panel given the low number of recurrent mutations seen in HGSC^3^ and so were unable to identify mutations in other genes, quantify mutational burden or comment on mutational signatures. Additionally, shallow WGS cannot assess genome-wide LOH and is subject to noise that is dependent on read depth and bin size^20^. It is thus possible we failed to observe recurrent rare events, such as patient-specific translocations in ABC transporter genes25 or other structural variants51 that can only be detected by deep WGS. Similarly, although we found no coordinated copy number changes between diagnosis and relapse, some individual genomes showed marked changes. It is possible that these diverse patient-specific changes ultimately converge on a common phenotype of recurrence and resistance, but analysis of larger cohorts will be required to ascertain whether these changes are more frequent at relapse. Certainly, spatial transcriptomic analysis suggests the existence of discrete subclones with unique CN alterations within individual tumour sections that may be critical drivers of resistance52.

Finally, a key potential driver of resistance is epigenetic change, which we did not examine here. Loss of *BRCA1* and *RAD51C* promoter methylation can drive platinum and PARP inhibitor resistance^13,53,54^ whilst acquired methylation, in particular of MLH1, is seen in acquired platinum resistance models^55^. More broadly, RNAseq of matched pairs has previously identified changes in immune-related genes^56^. However, transcriptional subtypes of HGSC^57,58^ derived from bulk analyses largely reflect immune cell composition and abundance of fibroblasts rather than intrinsic difference in malignant cells^59,60^. Overall, detailed single-cell analyses of matched pairs will be required to elucidate critical changes in copy number and gene transcription at relapse.

In summary, BriTROC-1 has allowed interrogation of genomes in relapsed HGSC and revealed remarkable stability of copy number changes across time, despite the extreme complexity of the HGSC genome. These data suggest that common short variants and copy number alterations cannot explain the pattern of relapse and acquired resistance that is the major hallmark of this disease. Importantly, we identified new genomic events at diagnosis, including *KRAS* amplification and CN signature 6 exposure, that are associated with primary platinum resistance and may have predictive utility for patients receiving neoadjuvant chemotherapy.

## Data Availability

All data produced in the present study are available upon reasonable request to the authors

## Code availability

Primary git repository - [https://github.com/BRITROC/britroc-HGSOC-landscape]

- Clinical database code [https://github.com/TBradley27/britroc1_db]
- Copy number pipeline [https://github.com/Phil9S/swgs-absolutecn]
- Copy number analysis [https://github.com/Phil9S/britroc-cn-analysis]
- Signature comparison methodology [https://github.com/lm687/BriTROC-1-paired_analysis]
- SNV analysis and pipeline [https://github.com/TBradley27/tamseq_somatic_variant_calling]

## Data availability

- Clinical data (anonymised)
- EGA submission for BAM files

## Supplementary Materials

<u>Supplemental data</u>

<u>Supplemental methods</u>

<u>Supplemental data - patient vignettes</u>

<u>Supplemental tables</u>

