## Supplementary methods for "The genomic landscape of recurrent ovarian high grade serous carcinoma: the BriTROC-1 study"

Supplemental methods and patient material

### Patient inclusion and exclusion criteria

#### Inclusion criteria

1. Patients with recurrent histologically-proven ovarian cancer, primary peritoneal carcinoma or fallopian tube cancer of high grade serous and high grade endometrioid subtypes. Patients who have a diagnosis of ovarian cancer with a known germline mutation in *BRCA1* or *BRCA2* will also be eligible for inclusion regardless of histological subtype. Patients who are having a diagnostic image-guided biopsy may be consented and study biopsy taken while awaiting pathological review. Eligible patients who have had samples collected under generic research consent may be registered retrospectively only after full discussion between the site, Chief Investigator and Cancer Research UK Clinical Trials Unit (and BriTROC-1 specific consent obtained).
2. Patients must have received at least one line of platinum-containing chemotherapy
3. Availability of formalin-fixed, paraffin-embedded tissue taken at the time of original diagnosis of high grade serous ovarian cancer. This may be primary surgical debulking specimen OR core biopsy. For those with only a core biopsy from time of diagnosis, availability of specimens taken at interval debulking surgery is desirable, but not essential.
4. Patients must have disease deemed suitable for imaging-guided biopsy (ultrasound or CT) by an experienced radiologist or suitable for intra-operative biopsy during secondary debulking surgery as determined by an experienced gynaecological oncology surgeon. Other biopsies, such as skin deposits, are also acceptable. However, this must be confirmed with the Cancer Research UK Clinical Trials Unit prior to patient registration.
5. Age ≥ 18 years.
6. Written informed consent.
7. Able to apply with study procedures.
8. Life expectancy > 3 months
9. No contraindication to biopsy as appropriate.

#### Exclusion criteria

1. Ovarian, primary peritoneal or fallopian tube cancer of low grade serous, grades 1 or 2 endometrioid, clear cell or carcinosarcoma/MMMT subtypes unless associated with known germline mutation in BRCA1 or BRCA2.
2. Borderline/low malignant potential tumours
3. Any non-epithelial ovarian malignancy
4. Patients with asymptomatic rising CA125 with no radiological evidence of recurrent ovarian cancer.
5. Original diagnosis of high grade serous cancer made on cytology only

### Tagged Amplicon Sequencing

#### Read alignment

Sequenced reads were aligned to the human reference genome (GRCh37 - g1kp2 *i.e.* ‘hs37d5’), using the bwa-mem algorithm (version: 0.7.17-r1188) in paired-end mode. Duplicate reads (*i.e.* paired-end reads with the same orientation position and start and end positions) were left unmarked and were not removed during the alignment process.

### Read alignment post-processing and QC

Samtools (v1.10) was used to fill in mate co-ordinates and insert sizes fields using the *fixmate* utility after the reads had been sorted by name using samtools *sort*. The same utility was used to resort the aligned reads this time by position so that the bam files could be cleaned by the Picard (2.25.7) *CleanSam* utility. Mate information in the alignments were further cleaned using the Picard *FixMateInformtion* utility. Picard’s *AddOrReplaceReadGroup* was then used to annotate bam headers with information relating to library, barcode and sample identifiers as well as sequencing platform and centre information. Bam files were then indexed using samtools *index* and finally validated using Picard’s *ValidateSamFile* utility to ensure that bam files were valid before any downstream processing occurred.

The following steps were implemented using code developed by CRUK Cambridge Institue’s bioinformatics core (v0.7.2; <https://github.com/crukci-bioinformatics/ampliconseq>):

Alignments were further cleaned to retain only those aligned reads whose alignment began within 1 base pair of the start or end position of any pre-specified amplicon genomic interval. Reads in which the corresponding mate pair read did also not align to the corresponding end of the amplicon were also removed. Each alignment file was split into a minimal set of non-overlapping amplicon alignment files, such that no subsequent alignment file contained any overlapping amplicons. This precludes the opportunity of errors in which the primer regions of some amplicons overlap with the targeted region of other amplicons, and therefore creating erroneous mutant allele fractions.

### Germline Variant Calling

Germline short variants were called on non-tumour samples with the aid of CRUK-CI’s amplicon seq pipeline (<https://github.com/crukci-bioinformatics/ampliconseq>; v0.7.2). Relevant pipeline steps were as followed:

All variant calling was performed using GATK’s HaplotypeCaller (GATK version 3.8-0-ge9d806836)^1, 2^. Variant calling was performed on individual library read alignments. Within each library, variant calling was performed on the targeted regions of individual amplicons, and therefore excluded amplicon primer and non-amplicon genomic regions.

In instances in which the same variant was called from multiple amplicons covering the same locus, the variant record with highest variant quality score was selected, whilst the remaining variant record was discarded. For instances in which paired end reads overlapped, both reads were discarded if the base calls were discordant, otherwise the read with the highest mapping quality was selected for the computation of downstream read count and SNV metrics.

HaplotypeCaller hard filters implemented were as followed for SNVs:

*QD < 2.0; FS > 60.0; MQ < 40.0; MQRankSum < -12.5*

And for short indels:
*QD < 2.0; FS > 200.0*

In addition to HaplotypeCaller filters, further filtering was performed by two unique functions within the ampliconseq pipeline which models dataset noise: The first models substitution specific noise at a specific locus for all libraries within a single sequencing run. The second models noise within individual libraries. Thresholds are determined based on modelled beta distributions using quantiles corresponding to a probability of 0.9999. All called variants below these two library and position specific noise thresholds are discarded. Variants which were not detected in both technical replicate libraries were also excluded.

### Tumour Sample Variant Calling

All variant calling on tumour samples were performed using the *cancer* calling mode of Octopus (v0.7.2)^3^ with the exception of *TP53* variants and their associated mutant allele fractions used to guide copy number calling, which were as reported previously^4-6^.

Variant calling was performed on tumour samples in two primary modes which will be described here individually - a *TP53* and a non-*TP53* calling mode:

*TP53* variants are called separately due to their unique role as necessary and ubiquitous drivers of tumourigenesis in high grade serous carcinoma^7^, and the presence therefore of an identical *TP53* mutation being expected in all tumour samples from the same patient. For *TP53,* variants were called on each tumour sample individually with the often integration of multiple libraries deriving from the same tumour sample.

Octopus was called with the following options: Downsampling of reads was disabled using the *–disable-downsampling* flag and both aligner and Octopus recognised read duplicates were not removed using the *–allow-marked-duplicates* and *–allow-octopus-duplicates* flags respectively. Expected somatic mutation frequencies were set at 0.03 and 0.01 using the *–min-expected-somatic-frequency* and *–min-credible-somatic-frequency* flags respectively. The maximum number of somatic haplotypes modelled (*--max-somatic-haplotype*) was set to 2.

Octopus will attempt to classify called variants as germline or somatic, and only variants classified as somatic were retained using the *–somatics-only* command line flag. Somatic variants were further filtered using the following set of hard filters: *--somatic-filter-expression "QUAL < 2 | GQ < 20 | MQ < 30 | SMQ < 40 | SB > 0.90 | SD > 0.90 | FRF > 0.5 | BQ < 20 | DP < 3 | ADP < 1 | MF > 0.2 | NC > 1 | AD < 1 | AF < 0.03"*

The set of amplicon regions for *TP53* variant calling were set to the union of genomic ranges specified in amplicon panels 1, 10 and 28 (as described in **Table S1**)*.*

Due to their often being multiple *TP53* variants detected per patient, all *TP53* mutations identified per patient were classified as suspected *driver* or *non-driver* mutations. A combined ranking and scoring process was conducted for each patient’s set of identified *TP53* variants. The mutation designated as the likely driver mutation for each patient was selected as being the representative *TP53* mutation for the construction of oncoprints reported in this study (*i.e.* **Figure 2**, **Figure S4** and **Figure S5**).

More precisely and formally, the scoring process for each patients set of *TP53* variants were followed as followed:

$v_{i}^{score}=v_{i}^{maf}+3v_{i}^{nsample}+v_{i}^{qual}{+v_{i}^{penality}}$

Where $v$ is a unique list of *TP53* mutations identified per patient indexed from $1,...,n$ for $n$ unique identified *TP53* mutations per patient. $v^{maf}, v^{nsample}$ and $v^{qual}$ are all descending rankings of $v$ by the mutant allele fraction, number of tumour samples in which the variant was located, and the variant quality score respectively such that the set of elements of $v^{maf}, v^{nsample}$ and $v^{qual}$ are all equal to $\left\{ 1,. . ., n \right\}$. $v^{penalty}$ accounts for potential batch effects by penalising variants which do not appear in both the diagnosis and relapse tumour classes such that $v^{penalty} \in\left\{ 0,12 \right\}.$ The suspected *TP53* driver mutation is labelled using the variant corresponding to $min(v^{score}).$

Non-*TP53* variants were called using three different methods: *Paired, paired and matched* and *cohort* analysis modes. *Paired* analyses (*cf* **Figure 2**) refers to analyses performed for patients with paired diagnosis and relapse tumour samples with the aim of identifying variants which are shared and not shared between these two tumour classes. *Paired and matched* methodology is used for patients with a diagnosis, relapse and matching non-tumour sample, and is used to confidently classify identified variants as germline or somatic (cf **Figure S4** and **Figure S5**). *Cohort* level analyses are neither *paired* nor *matched* but provide the greatest coverage of patients in this study.

All three non-*TP53* analysis modes were completed jointly for all relevant samples (depending on analysis mode) within a patient.

For the *paired and matched* analysis, octopus was ran as described previously for *TP53* mutations with the following additions/modifications:

Non-tumour bam files were supplied via the -N flag. Called somatic variants were additionally filtered using the supplied somatic random forest model supplied as part of the octopus software. In addition, the AF hard filter was set to ‘AF < 0.0001’. The genomic intervals used for this analysis was the intersection of amplicons used in amplicon panels 6 and 28 described in **Table S9**.

Variants which only appear in one of two technical duplicates are discarded as suspected artefacts. Additional filtering of C>T/G>A SNVs is implemented as described in the supplementary methods.

The *cohort* analysis was similarly performed to the *TP53* analysis with the addition that this analysis did not attempt to filter specifically for somatic variants (due to the lack of matched non-tumour samples). Both somatic and germline random forest models supplied with octopus were used for filtering using the *--somatic-forest* and *--forest* flags respectively. The set of hard filters for filtering putative germline variants were as followed:

*QUAL < 10 | MQ < 10 | MP < 10 | AD < 1 | AF < 0.01 | AFB > 0.25 | SB > 0.98 | BQ < 15 | DP < 1 | ADP < 1*

Variants were similarly filtered for those not appearing in both technical duplicates, and with additional thresholds for C>T/G>A mutations as described in the previous section.

*Paired* variant calling was performed as described for the *cohort* analysis method for a subset of patients possessing both diagnostic and relapse tumour samples, though with an additional step:

A second round of targeted variant calling was performed for all loci identified in the first round of variant calling, though this time using a different set of variant hard filters:

Changes in putative germline variant filtering: AF<0.0001, AFB>0.50
Changes in putative somatic variant filtering: AF<0.001

The aim of this step was to overcome an issue in the algorithm software in which suspected germline variants having experienced loss of heterozygosity were erroneously filtered by either the AF or AFB filters for unexpected allele fractions given an expected copy number of two for germline variants. Relaxing these threshold parameters at targeted calling only prevented an otherwise large increase in false positive calls.

### Fixation artefact correction

In order to identify any potential substitution-specific artefacts, suspected artefacts in which a mutation appeared in only one of two technical duplicates were counted, and MAF density estimates were produced. As previously reported^8^, a large enrichment of C>T transitions were identified in formalin fixed tissue compared to tissues preserved using different methods (UMFIX fixation in this study). Additionally, MAF density estimates for C>T substitutions from formalin fixed samples were shifted to the right compared to those not fixed with formalin.

An additional MAF threshold of 0.23 was implemented for C>T substitutions (and correspondingly cognate G>A substitutions) as a result. In DNA samples with particularly poor quality DNA due to formalin fixation, artefact MAFs are inflated^9^ leading to highly discordant MAFs between technical replicates. As a result, an additional C>T/G>A filter was also implemented for variants in which MAFs differed by more than 0.30 in order to remove further artefacts.

### Variant annotation

### All non-*TP53* variants were functionally annotated using Ensembl’s *variant effect prediction* (VEP) pipeline^10^ (v102.0). VEP was executed using the *–everything* and –*check_existing* flags. Only annotations for a gene’s canonical/representative (as determined by VEP) were considered. Variants with an allele frequency > 0.001 reported by the 1000 genomes project (The 1000 genomes project consortium 2015) or gnomAD^11^ were discarded. Variants with a labelled biotype of ‘non-stop decay’, or ‘processed pseudogene’ were removed from the variant set. All variants labelled as ‘benign’ by ClinVar^12^ were also discarded. Nonsynonymous and inframe indel variants without a ‘clinical significance’ annotation or an annotation of ‘uncertain_significance’ (*i.e.* VUS) were also discarded. Variants were also discarded if there was no clinical significance annotation and either SIFT^13^ or PolyPhen^14^ labelled the variant as ‘tolerated’ or ‘benign’ respectively. All synonymous, ‘intronic’, ‘upstream gene’, ‘downstream gene’, ‘non_coding_transcript_exon_variant’, ’intron_variant,non_coding_transcript_variant’ and 3’ and 5’ UTR labelled variants were also removed from the variant set. Variants were further refined using the molecular tumour board portal (MTBP)^15^. More specifically, variants labelled as benign or likely benign by MBTP were discarded.

### Inference of sample mislabelling events

Two different approaches were used and combined in order to identify putative sample mislabelling events. Firstly, the concordance between libraries belonging to normal and tumour samples deriving from the same patient were tested using a modified version of the HaveYouSwappedYourSamples method (Schröder *et al.* 2017). Namely, pairwise concordance scores were calculated for all normal-tumour library pairs by determining the proportion of high MAF variants (as determined by HaplotypeCaller) which were shared between each library pair.

A threshold was then applied in order to classify library pairs as being either concordant or discordant. Normal-tumour sample pairs were classified as potentially discordant if they did not contain any expected concordant library pairs. Secondly, a sample was identified as potentially being mislabelled if it contained what appeared to be a high MAF TP53 mutation that was discordant for other *TP53* mutations classified as ‘driver’ for that patient.

### Copy number fitting

#### Modified QDNAseq implementation

QDNAseq was modified to allow for read counts to be corrected for GC and mappability whilst being able to transform data back into read count space. To do this, several modifications were made to QDNAseq. In the correctBins() function the following line was altered.

After bin correction is performed using QDNAseq correctBins() function, the following transformation is applied to the binned copy number data to correct by the estimated bin correction generated by the estimateCorrection() function.

readCountsFiltered<-applyFilters(readCounts,residual=TRUE,blacklist=TRUE)
readCountsFiltered <- estimateCorrection(readCountsFiltered)
copyNumbers <- correctBins(readCountsFiltered)

assayDataElement(copyNumbers,"copynumber") <- sweep(assayDataElement( copyNumbers,"copynumber"),2,apply(assayDataElement(readCountsFiltered,"fit"),2,Median,na.rm=T),FUN='*')

copyNumbersSmooth <- smoothOutlierBins(copyNumbers)
copyNumbersSegmented <- segmentBins(copyNumbersSmooth,transformFun="sqrt")

Additionally, the sqrtadhoc() function which is utilised in the segmentBins() function was modified to adjust the anscombe transformation to be utilised with non-log data, in accordance with the previous alterations to the bin correction.

\## Original

sqrtadhoc <- function(x, factor=sqrtfactor(), offset=sqrtoffset(), inv=FALSE){
 if (!inv){
 x <- x + offset
 sqrt(x * factor)
 } else{
 x <- x^2 * (factor^-1)
 x - offset
 }
}

\## Modified
sqrtadhoc <- function(x, factor=sqrtfactor(), offset=sqrtoffset(), inv=FALSE){
 if (!inv){
 x <- x + offset
 2*sqrt(x + factor)
 } else{
 x <- (x/2)^2 - (factor)
 x - offset
 }
}

This modified version of QDNAseq is available [here](https://github.com/markowetzlab/QDNAseqmod).

#### Profile fitting

A grid search was performed across ploidy and purity ranges to estimate absolute copy number profile fitting. A range of quantitative and qualitative metrics were used to select the correct ploidy-purity combination from a single or set of best absolute fits for a given copy number profile. Samples with multiple equally likely fits were assessed independently by two investigators to select the best fit or exclude a sample from downstream analysis. Discordant assessments were discussed until a consensus was reached.

Sample fits were also subject to fit ‘power’ calculations, in which fits with insufficient reads to support the selected ploidy-purity combination were excluded from selection. Sufficiently ‘powered’ fits after quality control underwent downsampling to a fixed read depth of 15 reads per bin per tumour copy which acts to normalise inter-sample and intra-patient variance caused by varying coverage between samples. This manifests as an increased or decreased standard deviation in the bins associated with a given segment. Downsampled absolute copy number profiles were then generated and selected in the same manner as described previously, with selected fits used for downstream analysis.

Lastly, selected downsampled fits were subject to profile variance filtering. While the downsampling process seeks to reduce the amount of intra-patient and inter-sample bin variance, some absolute copy number profiles still retain a high degree of bin variance across their selected copy number fit. The standard deviation for bin values across copy number states 1, 2, 3, and 4 where calculated and samples with standard deviations of all bin values across all copy number states exceeding 3 standard deviations above the mean were removed, excluding four samples from downstream analysis (IM_159, IM_181, JBLAB-19324, JBLAB-4121).

### Copy number analysis

#### Copy number event calling

Copy number events were defined as a given segment in a copy number profile, under the assumption that each segment called as a CN event is independent of other neighbouring segment changes. For the analysis of CNA (both focal/gene-level and broad) copy number thresholds were used as defined by COSMIC and Allele-specific copy number analysis of tumours^16^.

Average genome ploidy ≤2.7

- Amplification: total copy number ≥5
- Deletion: total copy number = 0

Average genome ploidy >2.7

- Amplification: total copy number ≥9
- Deletion: total copy number < (average genome ploidy - 2.7)

For gene-level extending across more than one 30 kb genome bin, a mean was taken of all intersected bins. Broad events are defined on the basis of the proportion of affected cytoband with a threshold of 80% called as either amplified or deleted. For arm-level events a threshold of 50% of a chromosome arm (as a proportion of supporting bins) was selected. Copy number events which were equal to plus or minus one from a sample ploidy, but not called as amplification or deletions, were termed gains and losses, respectively.

#### Ploidy changes

Ploidy changes are difficult to assess due to the nature of absolute copy number fitting, where an incorrectly selected ploidy-purity combination would appear as change in ploidy between the diagnosis and relapse samples. Here, the patients with suspected ploidy changes underwent a scoring methodology to assess the likelihood of a true ploidy change versus a technical error during absolute profile fitting. Change in ploidy between samples was defined as;

$$ploidy change = | median(r_{i}) -median(d_{i}) | \geq1$$

Where $i$ is the patient, $r$ is the ploidy of relapse samples for patient $i$, and $d$ is the ploidy of diagnosis samples. Where multiple samples occur for a given sample group, a median value is taken. An absolute value of one or greater is defined as a patient with a ploidy change.

Ploidy change patients were assessed on the basis of three criteria to determine the confidence to which a ploidy change is likely to be true rather than as a consequence of erroneous or poor quality copy number fitting. These criteria are;

1) Selected fits have the highest scoring quantitative quality metrics compared to other sufficiently powered copy number fits (clonality error, TP53 estimate).
2) No underpowered fits with otherwise acceptable quality metrics are available which would contradict the selected copy number fit.
3) Additional samples, attributed to either diagnosis or relapse groups, support the ploidy change by also conforming to criteria 1 and/or criteria 2.

Meeting any of these criteria provides a given patient ploidy change with one star, with a maximum of three stars for a ploidy change with the maximum confidence. Patients with ploidy change and the assigned rating are detailed in **supplemental data - Table P9**.

For patient-specific analyses (patient loci clustering & gene correlation and heatmap), these ploidy change samples were excluded from the analysis due to the impact on patient clustering. This can be visualised below, where ploidy change patients constitute a large proportion of the more extreme copy number changes between diagnosis and relapse.

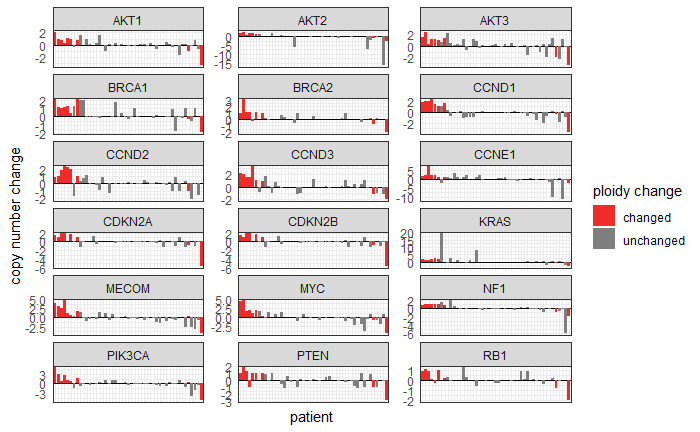

#### *Supplemental methods Figure 1 -* ***Gene change bar plot***

A faceted bar plot showing the copy number changes at frequently altered or clinically relevant genes (n = 47). Plots show the net gain or loss of gene copies between diagnosis and relapse tumours, where a median was taken for time points with multiple samples. A positive value indicates a net gain of gene copies and a negative value a net loss of gene copies, between diagnosis and relapse. Patients with identified ploidy change between diagnosis and relapse are coloured in red.

#### Purity differences

As expected, we observed differences in tumour purity values for absolute fitted copy number profiles across different biopsy sites and fixation methods, but purity was still consistent between diagnosis and relapse (p=0.61; Mann-Whitney U test), including stratification for platinum-based treatment response (resistant p=0.57, sensitive p=0.80; Mann-Whitney U test) (**supplemental data - Figures S7A & S7C**). Sample purity remained stable at the cohort-level when only including paired patients between diagnosis and relapse (p=0.73; Wilcoxon signed-rank test), including stratification for platinum-based treatment response (resistant p=1.0, sensitive p=0.62; Wilcoxon signed-rank test) (**supplemental data - Figures S7B & S7D**).

### Intra-tumour heterogeneity

As implemented by van Dijk *et al.*^17^, copy number heterogeneity (CNH) is calculated as the minimisation of segment distance from integer state using a ploidy-purity grid search over segment $i$, where;

$CNH = min_{\alpha,\tau}\left( \frac{\sum_{i} d_{i}w_{i}}{\sum_{i} w_{i}} \right)$	(1)

Where d$i$ is the absolute distance of a segment from an integer defined as;

$d_{i}=|q_{i}-round(q_{i}) |$	(2)

Where q$i$ is the absolute copy number of a segment, α is the sample purity, τ is the average sample ploidy, and w$i$ is the segment width.

Our implementation forgoes performing a ploidy-purity grid search to determine the lowest chromosomal copy number heterogeneity across as ploidy and purity values for a given sample have already been determined during absolute fitting. As such we calculate CNH (hereto referred to as intra-tumour heterogeneity; ITH) as;

$ITH = \frac{\sum_{i} d_{i}w_{i}}{\sum_{i} w_{i}}$	(3)

Where d$i$ is the absolute distance of a segment from an integer defined in equation 2. Noisy segments were excluded as described by van Dijk *et al.^17^* using the standard deviation of the mean (*σ_μ_*) bin distributions across each segment.

Noise thresholds were set at 2 standard deviations greater than the mean noise, where cutoffs were set to a threshold of *σ_μ_* > 1.48, and *σ_μ_* > 0.875, for segments and samples, respectively. Noise thresholds removed nine samples (3.4%) and 685 segments (1.37%, mean and median of 2.59 and 1.00 segments per sample, respectively). After sample exclusion, segment filtering removed 467 segments (0.97%, mean and median of 1.82 and 1.00 segments per sample, respectively). (**Supplemental data - Figure S16**).

### Copy number signature abundance modelling

#### Partial ILR-Bernoulli model

Compositional data are defined by their sum constraint (exposures add up to one) and positivity (exposures are equal or larger than zero), therefore, any regression methods used to analyse them have to be appropriate for a multivariate compositional response. The basis of compositional data analysis^18^ is that a compositional vector of length d can be transformed to an unconstrained vector in R^{d-1} without loss of information, and removing the sum-constraint. Here, as we have described previously^19^, we use the Isometric Log-Ratio (ILR) transformation^20^, in which we use an orthonormal basis to transform the data.

A further challenge in copy number exposure data is the presence of zero values. We address it by using a variant of the ILR transformation, the partial ILR, in which only non-zero values are taken into account. The presence or absence of signatures is analysed using a Bernoulli model. We introduce mixed effects in both models to capture the information about paired archival-relapse samples. The models are implemented in Template Model Builder^21^ and run through R.

#### Model interpretability

The transformation of compositional data is adequately explained as follows; instead of analysing signatures S1 through S7, we analyse the following signature comparisons after ILR transformation of S1 vs S2, S3 vs the mean of S1 and S2, S4 vs the mean of S1-S3, and so forth. This leads to a total of six pairwise comparisons (**supplemental methods fig 2**). The means of comparison is by taking the log-ratio of signatures (or groups of signatures), and we use the geometric mean to group signatures.

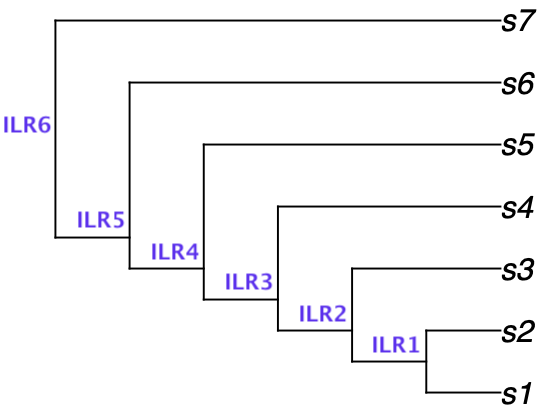

**Supplemental methods Figure 2 - Visualisation of IRL pairwise comparisons within partial ILR model** - Tree structure visualisation represents the pairwise comparisons of ratios between various subgroupings of transformed signatures. For example, ILR1 is the ratio of S1 to S2, ILR4 is the ratio of the geometric mean of S1-S4 to S5.

The two parameters of interest in the model are the intercept and the slope. Both are vectors of length 6 (i.e. as many as comparisons). The intercept indicates, in transformed space, the abundance of signatures in the first group of samples. This intercept can be transformed back to compositional data (using the inverse ILR transformation) to get the mean abundance of signatures in the first group. The slope is the difference in signature abundance between the groups, in transformed space. Therefore, the sum of the intercept and the slope gives us the mean abundance of signatures in the second group, in transformed space. A slope of zero indicates that the exposures are not different between groups.

As an example, **supplemental methods Figure 3** shows the intercept and slope for a scenario in which there are three mutational signatures (therefore, they are both vectors of length two). For the intercept, the first ILR is close to zero, indicating that the log-ratio between S1 and S2 is close to zero, and that therefore the mean abundance of S1 and S2 in the first group is roughly the same. ILR2 is negative, indicating that the abundance of S3 is lower than the geometric mean of S1 and S2. For the beta slopes, that of ILR1 is slightly negative, indicating that the ratio between S1 and S2 is a bit lower in the second group than in the first group. The slope for ILR2 is positive, indicating that S3 is more prevalent in the second group.

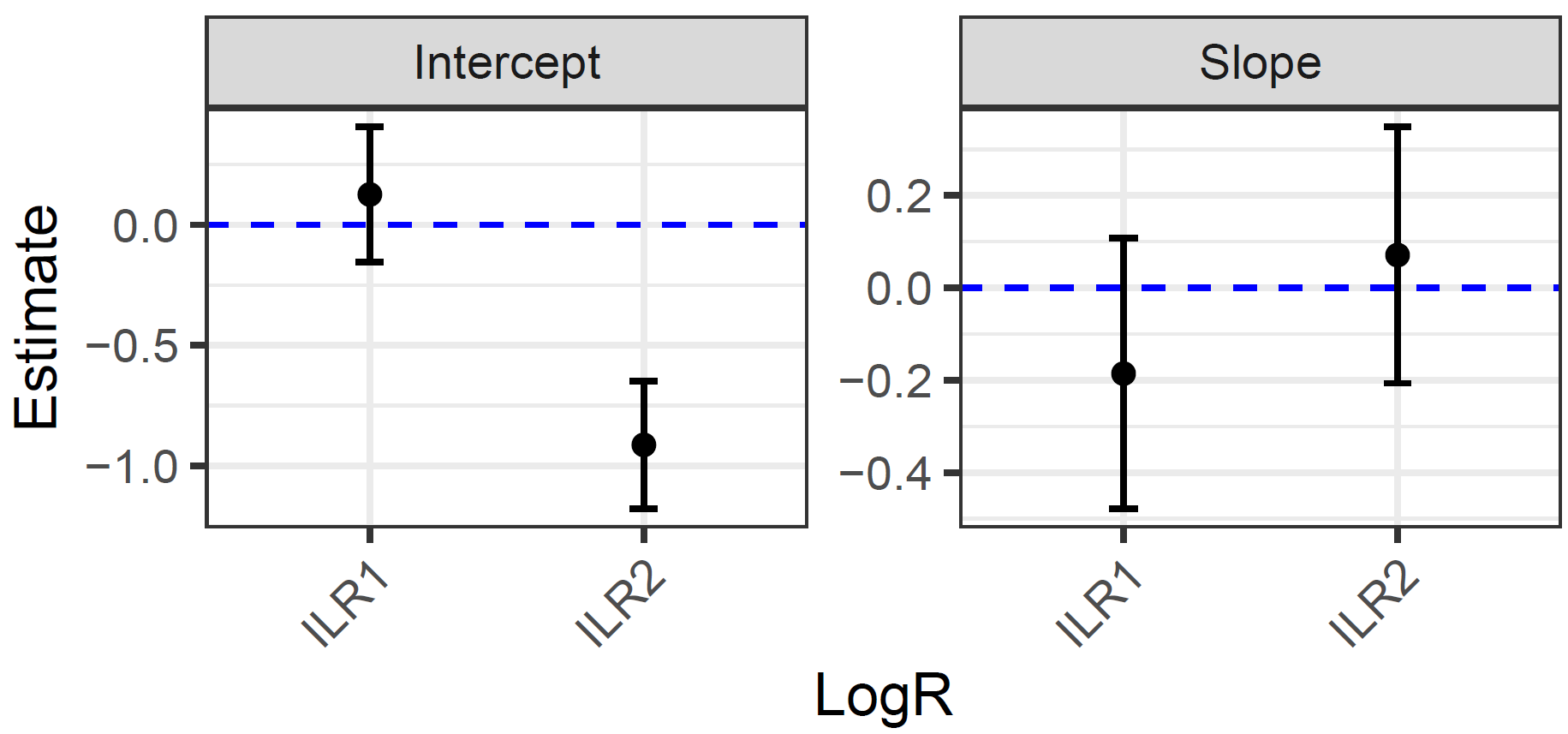

**Supplemental methods Figure 3 - Example beta slope and beta intercept plot** - Plot demonstrates an example outcome from a set of 3 signatures modelled using the described partial ILR transformation. Left plot is the beta intercept for ILR1 and ILR2 which are the transformed ratios of signatures S1-S3. Right plot is the beta slope for ILR1 and ILR2 which are the transformed ratios of signatures S1-S3.

### Immune environment from copy number signatures

#### Sample preparation

Tissue microarrays (TMA) were created using 1mm cores from viable archival formalin-fixed paraffin-embedded blocks of archival samples of the BriTROC study. Three representative cores were taken from each block. 3μm sections of the TMA blocks were cut using a Leica microtome, transferred to a water bath pre-heated to 60°C and collected on a SuperFrost Plus glass slides. The sections were dried overnight at 37°C then baked at 60°C for 1h to remove excess wax.

#### Automated Staining

Automated staining was carried out using the Ventana Discovery Ultra platform. All bulk reagents used were purchased from Roche. Sections were rehydrated by incubating them in EZ prep solution for 32min at 69°C, then antigen retrieval was performed by incubating the sections for 1h in Ventana Cell Conditioning buffer 1 (CC1) at 96°C (pH 8.5). After 4min incubation in hydrogen peroxide anti-human primary antibodies were applied and incubated for 1h at 37°C (CD3, Roche 790-4341, prediluted; CD8, Spring Bioscience M5394, 0.5ug/ml; CD20 Dako M0755, 0.1ug/ml; Foxp3, Spring Bioscience M3974, 0.25ug/ml; Pan-Keratin, Roche 760-2135, prediluted; Mouse IgG, Roche 760-2014, prediluted; Rabbit IgG, Roche 760-1029, prediluted; Goat IgG, Novus Biologicals AB-108, 0.1ug/ml). After a 16 min incubation with an HRP-conjugated secondary antibody, the chromogenic signal was developed in either 3, 3’-Diaminobenzidine (DAB) chromogen for 8 min, in Purple chromogen for 40 min, in Yellow chromogen for 28 min or in Teal chromogen for 8 mins. For chromogenic counter-staining, the slides were incubated for 4 min in Copper, 8 min in haematoxylin and 4 min in Bluing Reagent. Stained slides were dehydrated using the Leica Autostainer ST020, manually cover-slipped and digitally scanned by Aperio Scanscope XT.

#### Quantification of tumour immunohistochemistry markers

After quantification of marker positive cells across fixed image areas, grouped by stromal, tumour, and all tissue, cell counts were normalised to a marker-positive cells per micrometre squared (cells/μm^2^) and were further rescaled into log(1+ X) where X is cells/μm^2^ to account for extreme positive counts and zero counts for each image during modelling.

### Supplementary Methods References

1	McKenna A, Hanna M, Banks E et al. The Genome Analysis Toolkit: a MapReduce framework for analyzing next-generation DNA sequencing data. Genome research 2010; 20 (9): 1297-1303.

2	Poplin R, Chang PC, Alexander D et al. A universal SNP and small-indel variant caller using deep neural networks. Nature biotechnology 2018; 36 (10): 983-987.

3	Cooke DP, Wedge DC, Lunter G. A unified haplotype-based method for accurate and comprehensive variant calling. Nature biotechnology 2021; 39 (7): 885-892.

4	Piskorz AM, Ennis D, Macintyre G et al. Methanol-based fixation is superior to buffered formalin for next-generation sequencing of DNA from clinical cancer samples. Ann Oncol 2016; 27 (3): 532-539.

5	Goranova T, Ennis D, Piskorz AM et al. Safety and utility of image-guided research biopsies in relapsed high-grade serous ovarian carcinoma-experience of the BriTROC consortium. Br J Cancer 2017; 116 (10): 1294-1301.

6	Macintyre G, Goranova T, De Silva D et al. Copy-number signatures and mutational processes in ovarian carcinoma. Nat Genet 2018; 50 (9): 1262-1270.

7	Ahmed AA, Etemadmoghadam D, Temple J et al. Driver mutations in TP53 are ubiquitous in high grade serous carcinoma of the ovary. J Pathol 2010; 221 (1): 49-56.

8	Williams C, Pontén F, Moberg C et al. A high frequency of sequence alterations is due to formalin fixation of archival specimens. Am J Pathol 1999; 155 (5): 1467-1471.

9	Wong SQ, Li J, Tan AY et al. Sequence artefacts in a prospective series of formalin-fixed tumours tested for mutations in hotspot regions by massively parallel sequencing. BMC medical genomics 2014; 7: 23.

10	McLaren W, Gil L, Hunt SE et al. The Ensembl Variant Effect Predictor. Genome biology 2016; 17 (1): 122.

11	Karczewski KJ, Francioli LC, Tiao G et al. The mutational constraint spectrum quantified from variation in 141,456 humans. Nature 2020; 581 (7809): 434-443.

12	Landrum MJ, Lee JM, Benson M et al. ClinVar: improving access to variant interpretations and supporting evidence. Nucleic Acids Res 2018; 46 (D1): D1062-d1067.

13	Vaser R, Adusumalli S, Leng SN et al. SIFT missense predictions for genomes. Nature protocols 2016; 11 (1): 1-9.

14	Ramensky V, Bork P, Sunyaev S. Human non-synonymous SNPs: server and survey. Nucleic Acids Res 2002; 30 (17): 3894-3900.

15	Tamborero D, Dienstmann R, Rachid MH et al. The Molecular Tumor Board Portal supports clinical decisions and automated reporting for precision oncology. Nat Cancer 2022; 3 (2): 251-261.

16	Van Loo P, Nordgard SH, Lingjaerde OC et al. Allele-specific copy number analysis of tumors. Proc Natl Acad Sci U S A 2010; 107 (39): 16910-16915.

17	van Dijk E, van den Bosch T, Lenos KJ et al. Chromosomal copy number heterogeneity predicts survival rates across cancers. Nature Communications 2021; 12 (1): 3188.

18	Aitchison J. The Statistical Analysis of Compositional Data. Journal of the Royal Statistical Society Series B (Methodological) 1982; 44 (2): 139-177.

19	Cheng Z, Mirza H, Ennis DP et al. The Genomic Landscape of Early-Stage Ovarian High-Grade Serous Carcinoma. Clin Cancer Res 2022; 28 (13): 2911-2922.

20	Egozcue JJ, Pawlowsky-Glahn V, Mateu-Figueras G et al. Isometric Logratio Transformations for Compositional Data Analysis. Mathematical Geology 2003; 35 (3): 279-300.

21	Kristensen K, Nielsen A, Berg CW et al. TMB: Automatic Differentiation and Laplace Approximation. Journal of Statistical Software 2016; 70 (5): 1 - 21.
