## Supplementary figures and tables for "The genomic landscape of recurrent ovarian high grade serous carcinoma: the BriTROC-1 study"

**A**

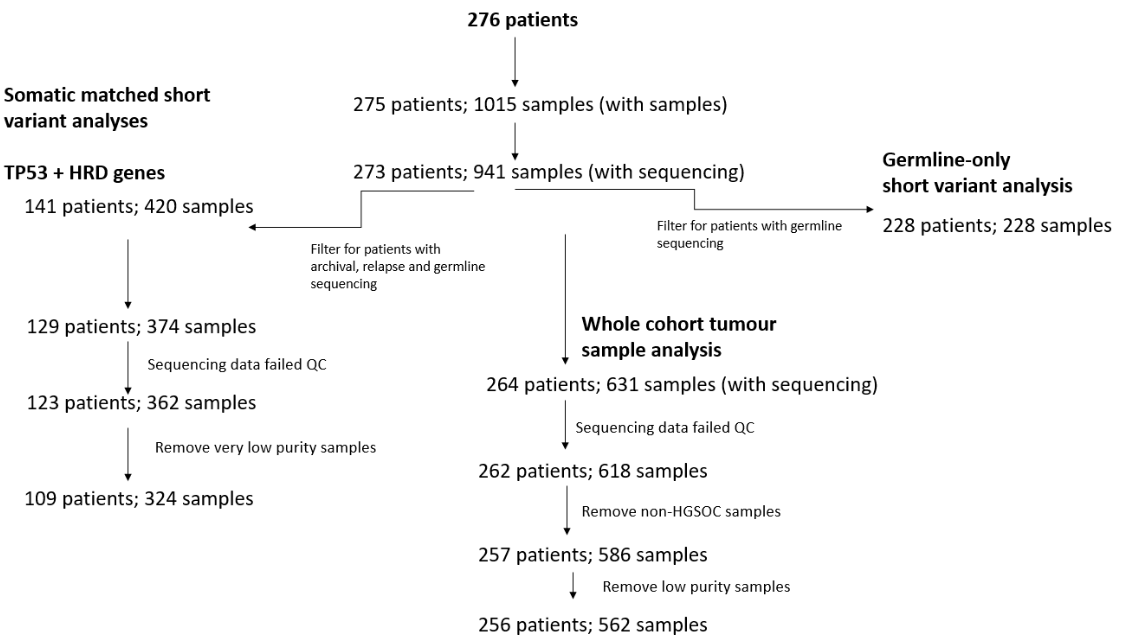

**B**

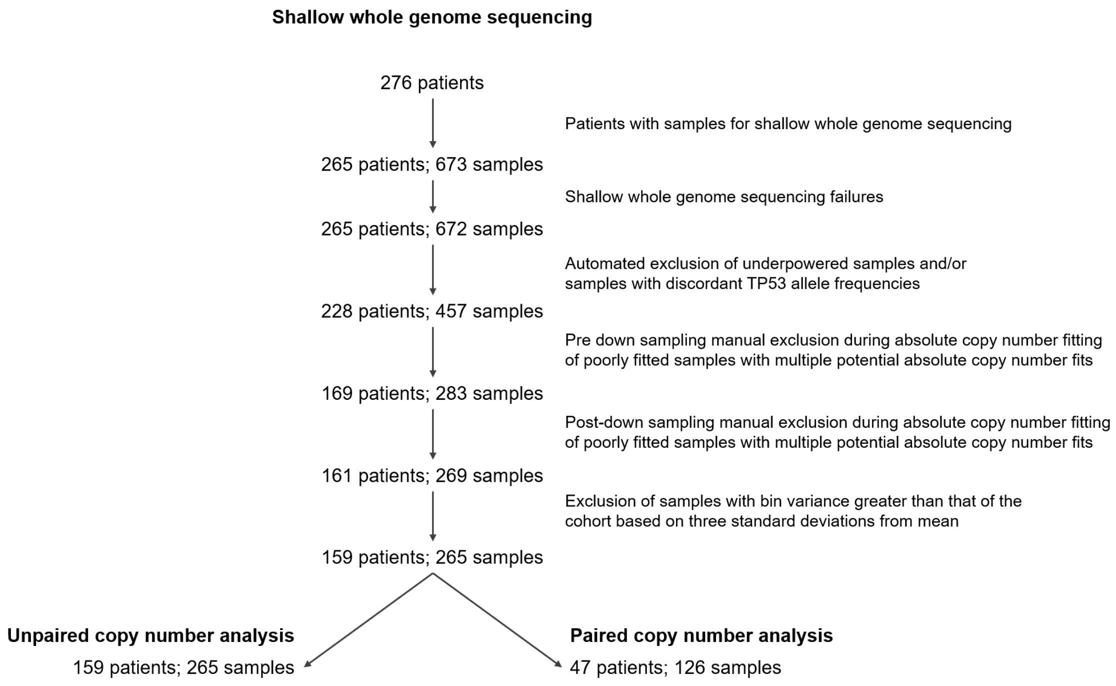

***Figure S1. BriTROC study REMARK diagrams***

***A*** *- Patient/sample flow through the single nucleotide variant analysis pipeline, including patient/sample exclusion rationale and pipeline end points.*

***B*** *- Patient/sample flow through the shallow whole genome sequencing pipeline, including patient/sample exclusion rationale and pipeline end*

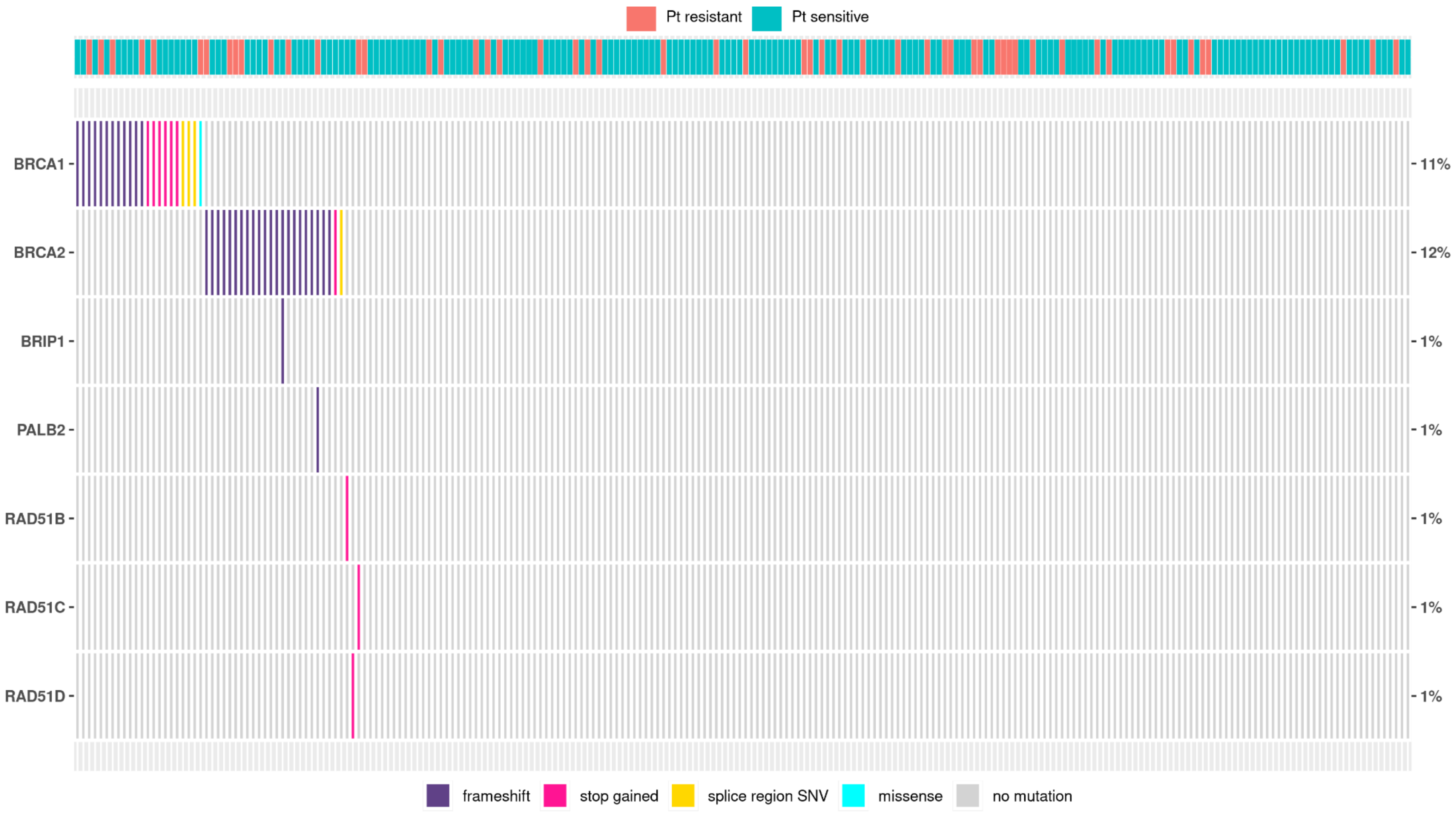

***Figure S2. Germline SNVs and short indels identified in key homologous recombination pathway genes***

Germline DNA extracted from whole blood samples from 228 BriTROC-1 patients was tested for short variants in key HR genes. Each column represents one patient, colour coded to denote patient platinum sensitivity status at study entry. The bottom legend denotes variant type. *FANCM* and *BARD1* were also tested, but no mutations were identified for any patient

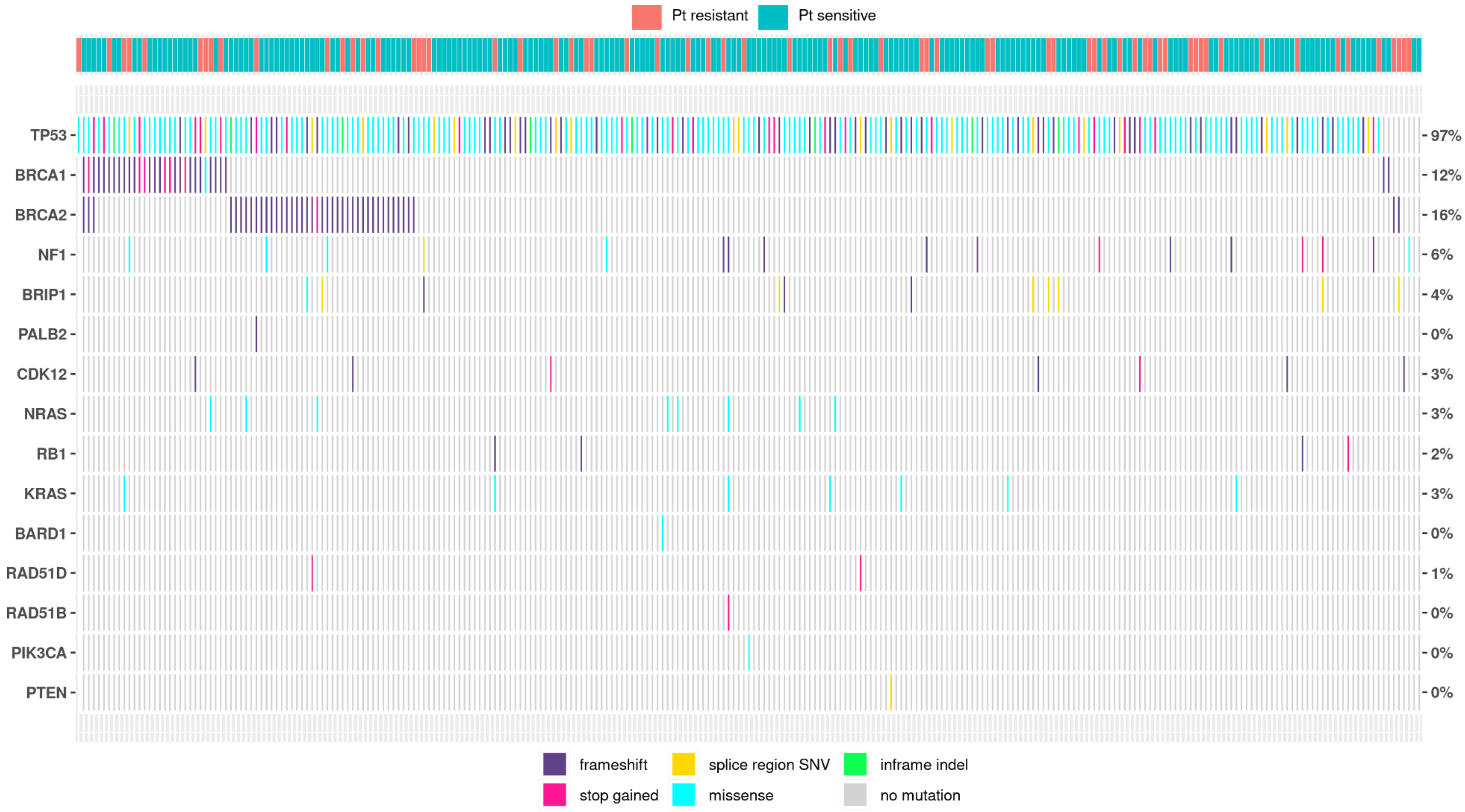

***Figure S3. Whole cohort-level detection of SNVs and short indels in key cancer related genes (unpaired)***
*DNA samples extracted from all tumour samples (both diagnostic and relapse) from 265 patients were tested for short variants in 20 relevant cancer genes. Mutations were not classified as somatic or germline in this analysis nor classified by relapse status (diagnosis vs relapse). Samples were not matched with corresponding normal DNA for each patient. The bottom legend denotes variant type. EGFR, FANCM, RAD51C, PALB2, BRAF and CTNNB1 were also targeted, but no mutations were identified.*

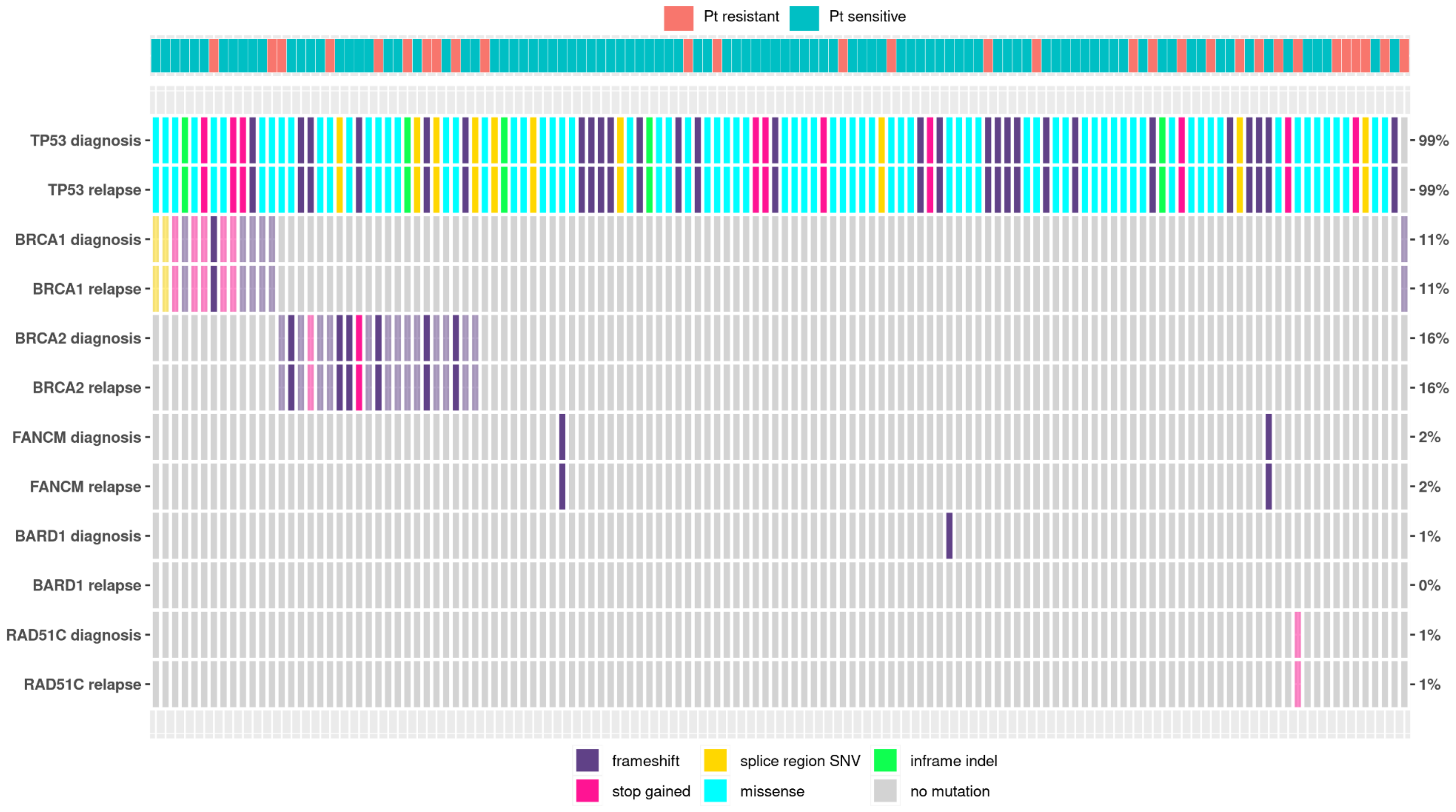

***Figure S4. Changes in detected somatic mutations from diagnosis to relapse (matched and paired analysis)***
*Patients with both diagnosis and relapse tumour samples and also non-tumour samples were examined for the gain or loss of variants from diagnosis to relapse. Inclusion of matched non-tumour samples allowed the confident classification of variants as germline or somatic. TP53 was not matched for the non-tumour sample. Variants with full opacity represent somatic variants whereas variants with reduced opacity represent germline variants. RAD51B, RAD51D, BRIP1 and PALB2 were tested but no mutations were identiified.*

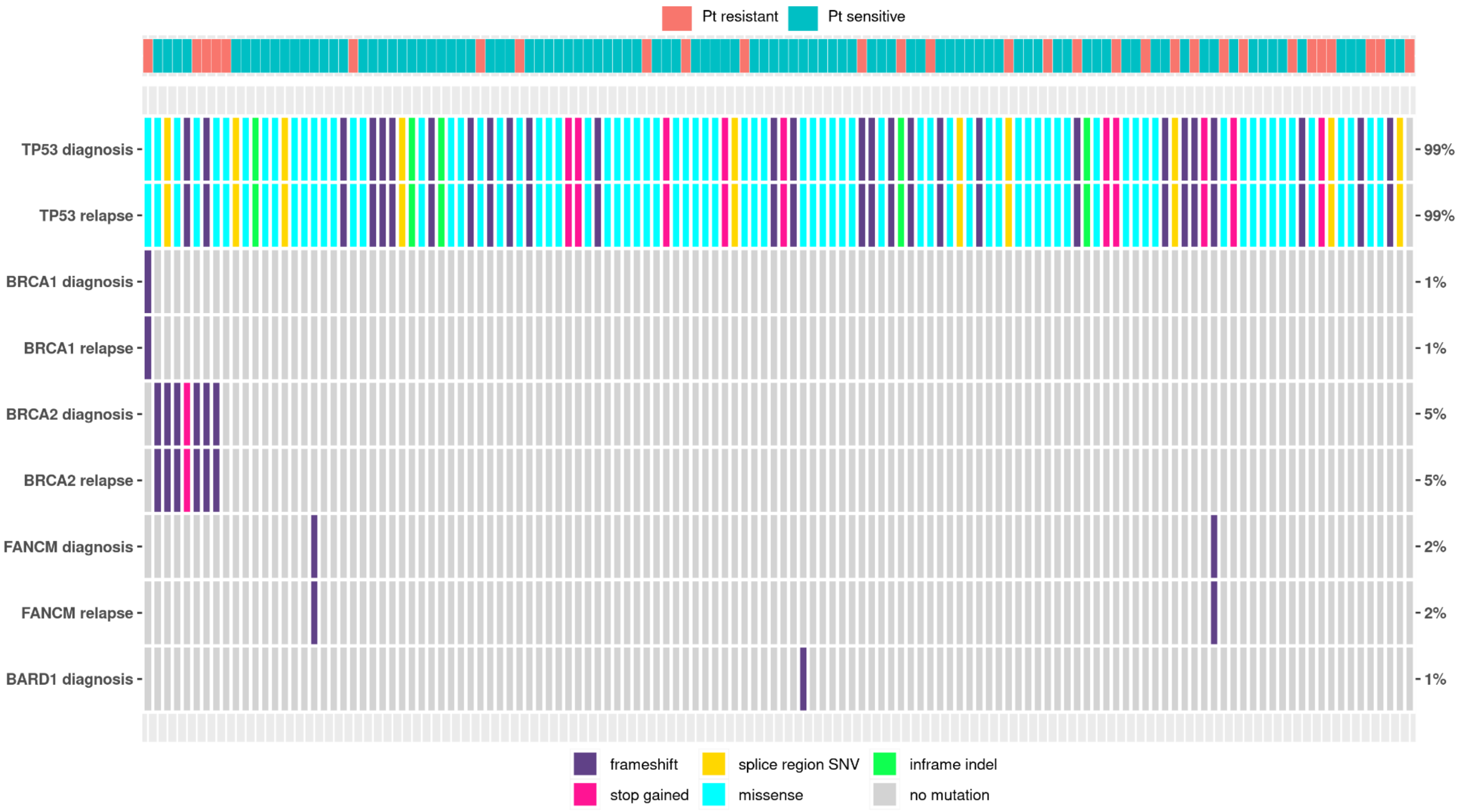

***Figure S5. Changes in detected somatic mutations from diagnosis to relapse (matched and paired analysis)***
*As in figure S4 with somatic mutations only displayed for this figure*

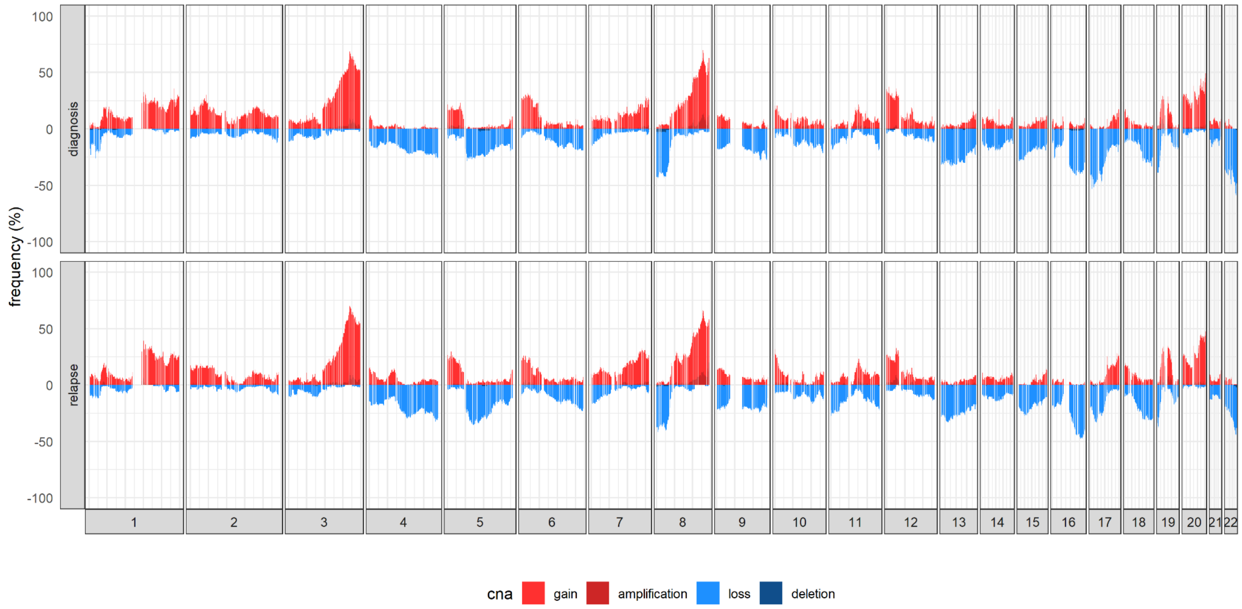

***Figure S6. Genome-wide copy number alteration frequency plot***

*A summary plot of the genome-wide frequency of absolute copy number alterations across diagnosis and relapse samples. Red indicates an increase in genomic copies (defined as either gains or amplifications) and blue indicates a decrease in genomic copies (defined as either losses or deletions). This plot demonstrates the genomic similarities between the alteration frequency of diagnosis and relapse cohorts.*

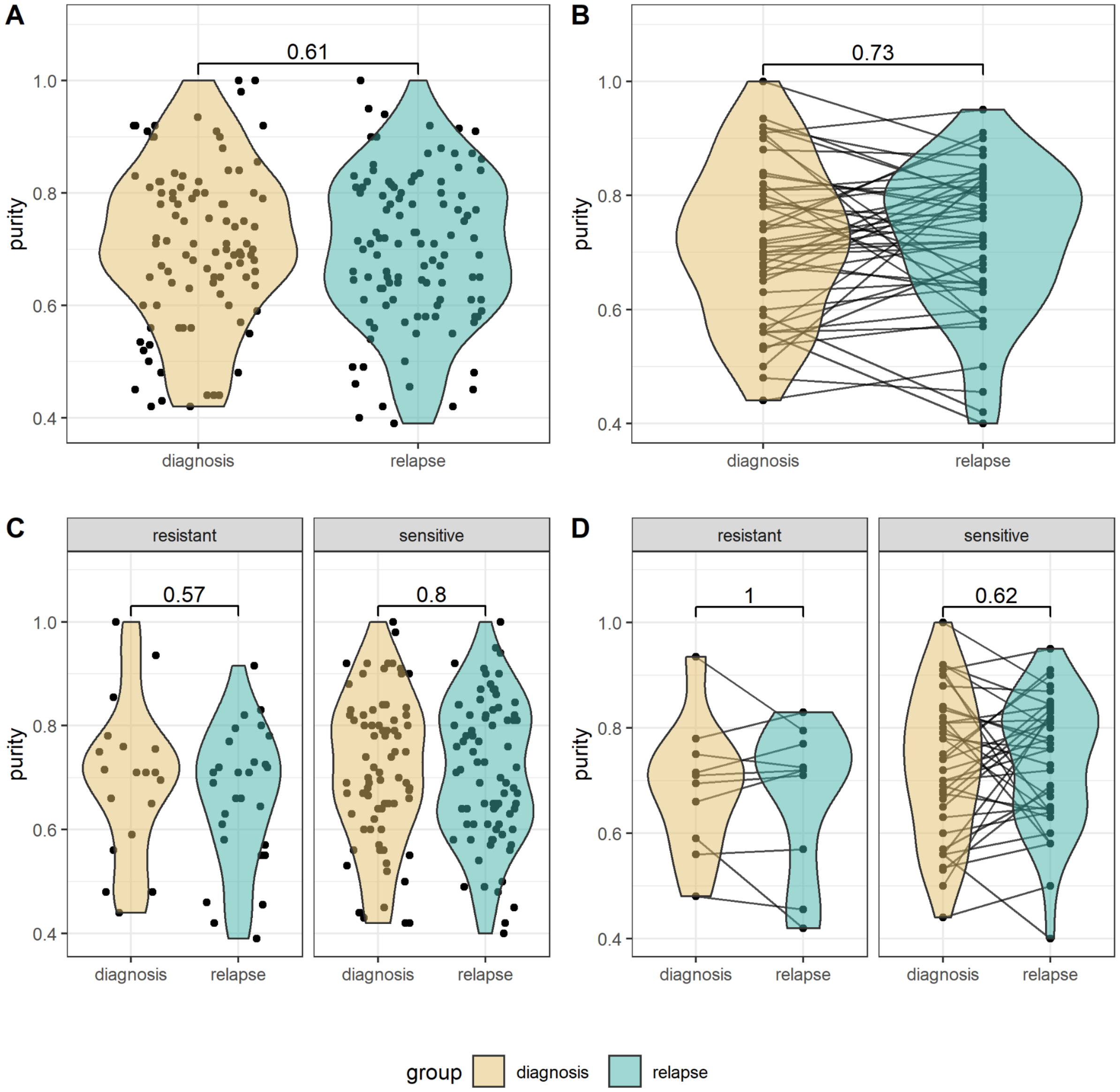

***Figure S7: Purity distributions for primary and relapse tumours***

*Distribution of fitted purity values for each sample identified during absolute copy number fitting (tested using Mann-Whitney U and Wilcoxon signed-rank for unpaired and paired groupings, respectively).*

***A** - All samples between diagnosis and relapse (n = 98 & n = 108, respectively).*

***B** - Paired samples between diagnosis and relapse (n = 47 & n = 47, respectively).*

***C** - All samples between diagnosis and relapse, stratified by platinum sensitivity (n = 19, n = 28, n = 77, n = 79, resistant-diagnosis, resistant-relapse, sensitive-diagnosis, sensitive-relapse).*

***D** - Paired samples between diagnosis and relapse, stratified by platinum sensitivity (n = 10 & n = 37, resistant and sensitive, respectively).*

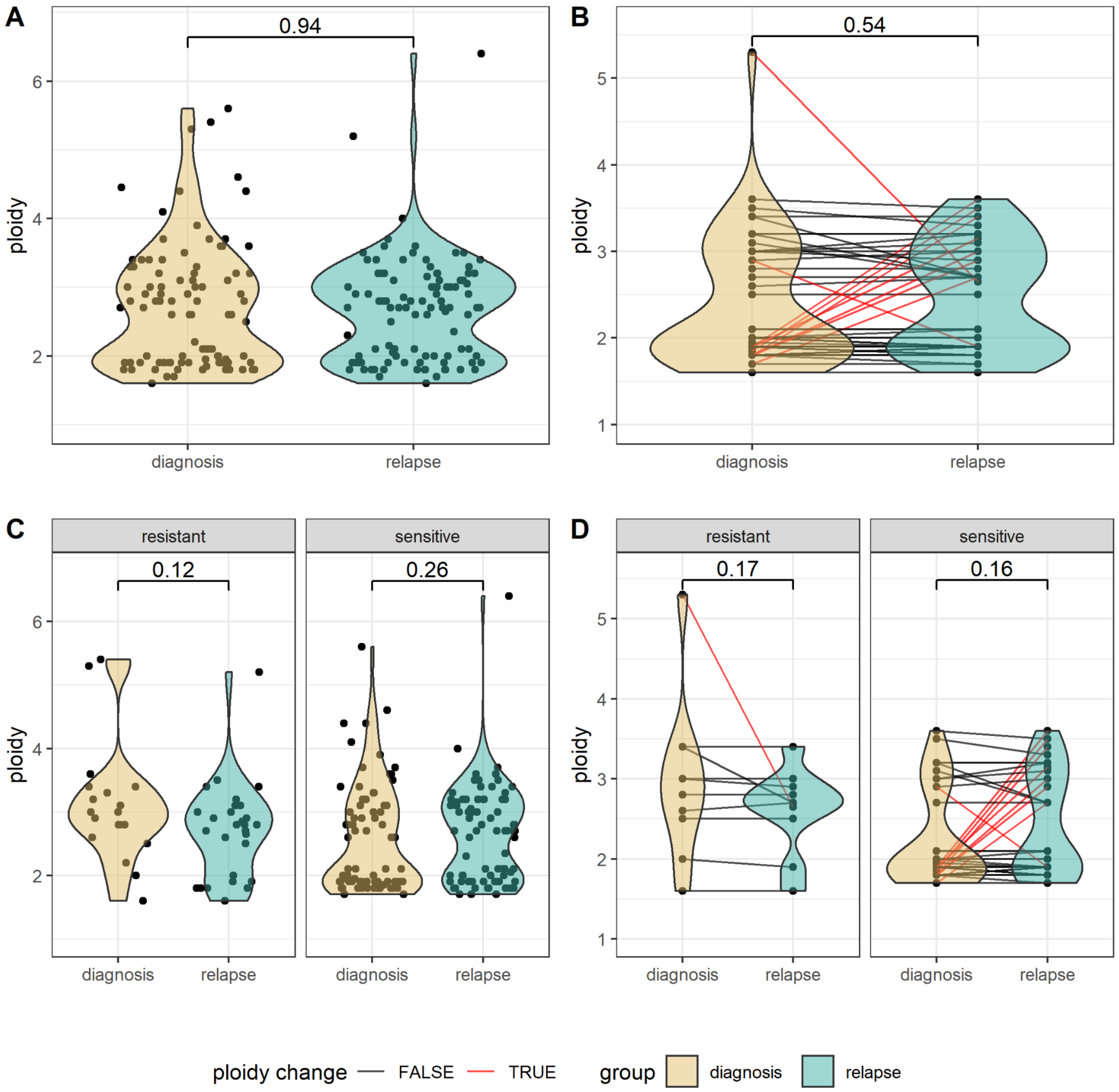

***Figure S8. Ploidy distributions for primary and relapse tumours***

*Distribution of fitted ploidy values for each sample identified during absolute copy number fitting (tested using Mann-Whitney U and Wilcoxon signed-rank for unpaired and paired groupings, respectively). Samples with ploidy change are marked in red.*

***A** - All diagnosis and relapse relapse (n = 98 & n = 108, respectively).*

***B** - Paired (diagnosis and relapse) samples (n = 47 & n = 47, respectively).*

***C** - All samples stratified by platinum sensitivity (n = 19, n = 28, n = 77, n = 79, resistant-diagnosis, resistant-relapse, sensitive-diagnosis, sensitive-relapse).*

***D** - Paired (diagnosis and relapse samples), stratified by platinum sensitivity (n = 10 & n = 37, resistant and sensitive, respectively).*

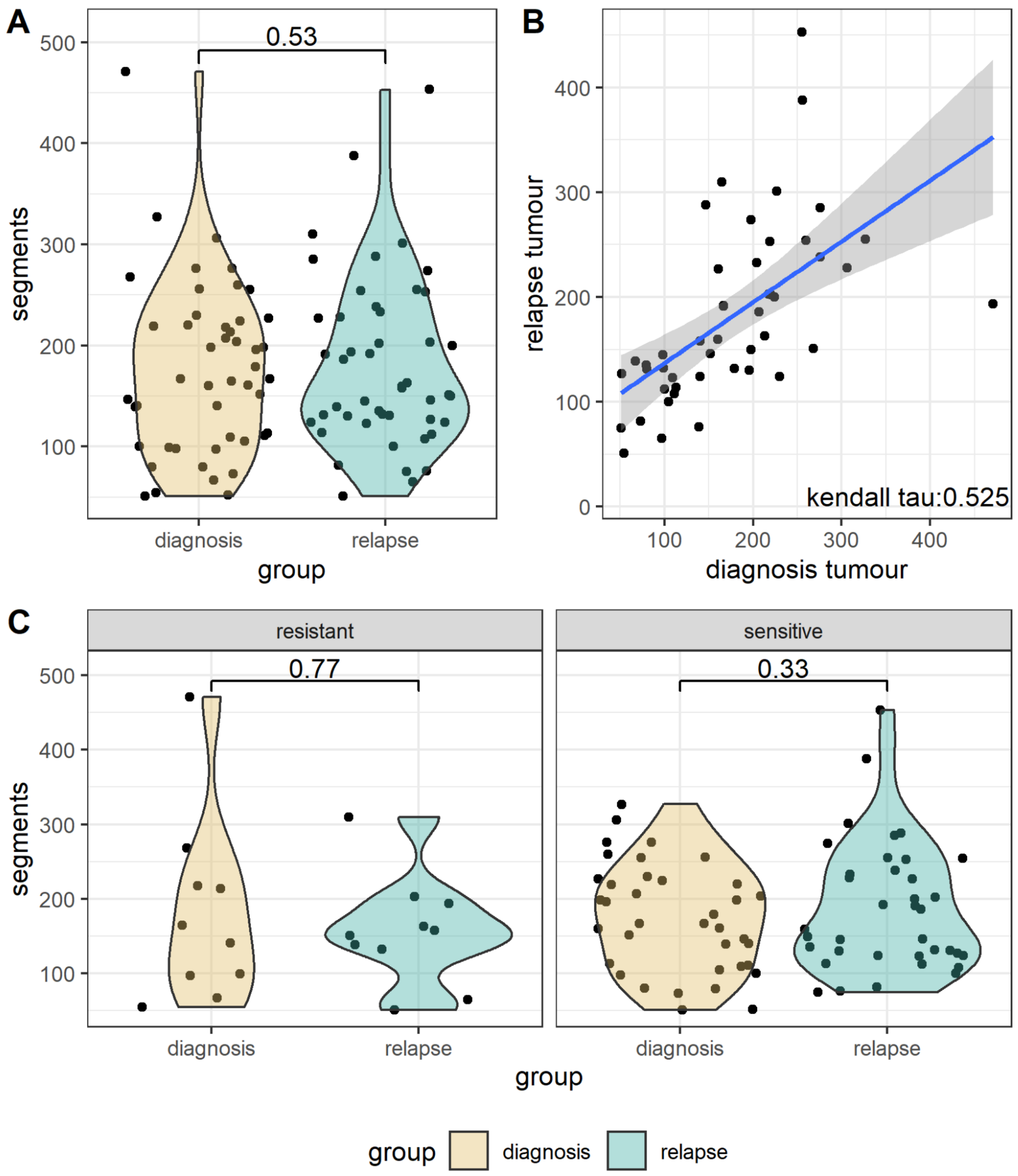

***Figure S9. Segment distributions for primary and relapse tumours***

***A** - Comparison of segment counts per sample between primary and relapse samples across paired samples. No statistically significant difference was found between the number of segments in paired primary and relapse samples (n = 47) (p-values are calculated using a wilcoxon signed rank test).*

***B** - Scatter plot of segment counts for each sample in primary and relapse tumours in paired patients. Segments were averaged across samples where multiple samples were available for either the primary or relapse group. Blue line indicates the linear regression line and the shaded portion is the 95% confidence interval. Correlation was calculated using kendall rank correlation.*

***C** - Comparison of segment counts per sample between paired primary and relapse samples stratified by patient platinum-based treatment sensitivity (resistant; n = 10 & sensitive; n = 37) (p-values are calculated using a wilcoxon signed rank test).*

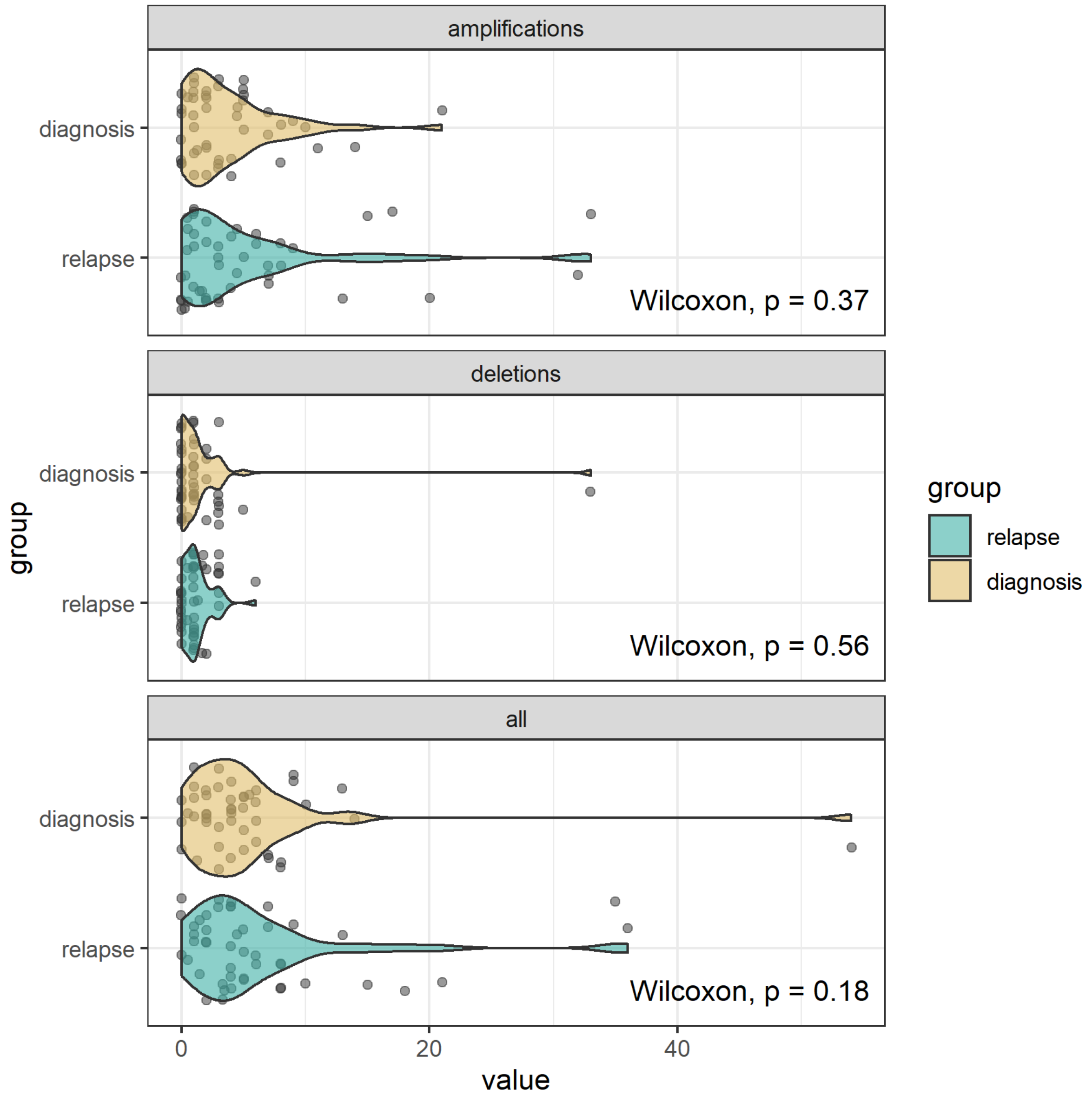

***Figure S10. Copy number events***

*Copy number event distributions calculated using segments as a proxy for a copy number event change between diagnosis and relapse sample groups. Summary plots of copy number events are stratified by event type (all, amplification, and deletion; diagnosis and relapse n = 47 paired samples). Differences in event distributions were tested using a wilcoxon ranked-sign.*

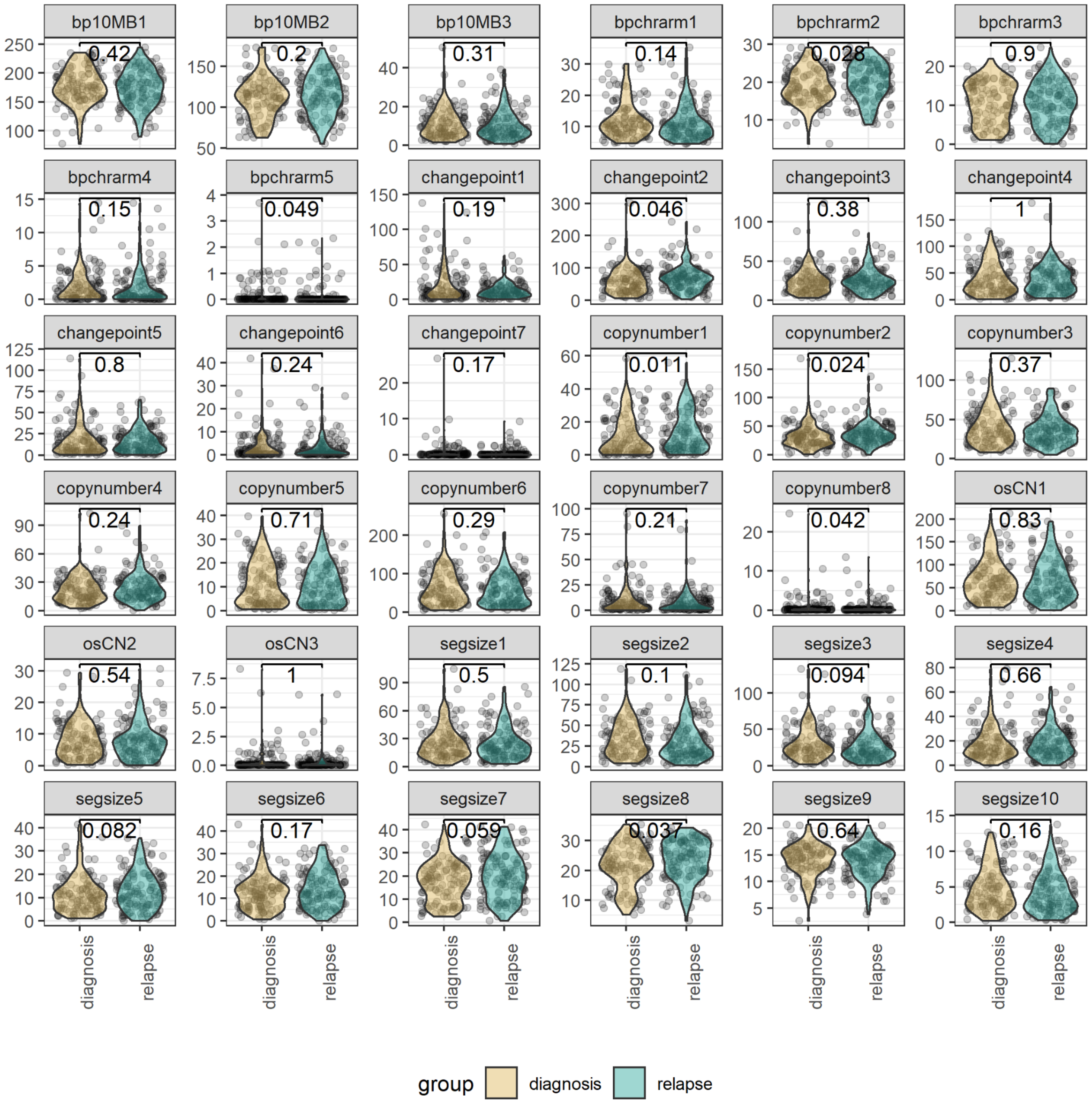

***Figure S11. Copy number features***

*Faceted plot of copy number feature distributions calculated during copy number signature extraction. These features are the same copy number features utilised in the derivation of copy number signatures {Macintyre, 2018 #6710} and should therefore provide a robust comparison of the differing copy number processes between diagnosis and relapse samples (n = 126 & n = 139, respectively). Distributions were tested using a Mann-Whitney U test . Of all tested, 7/36 copy number features were determined to be significantly different between diagnosis and relapse but none were statistically different after false discovery rate testing correction.*

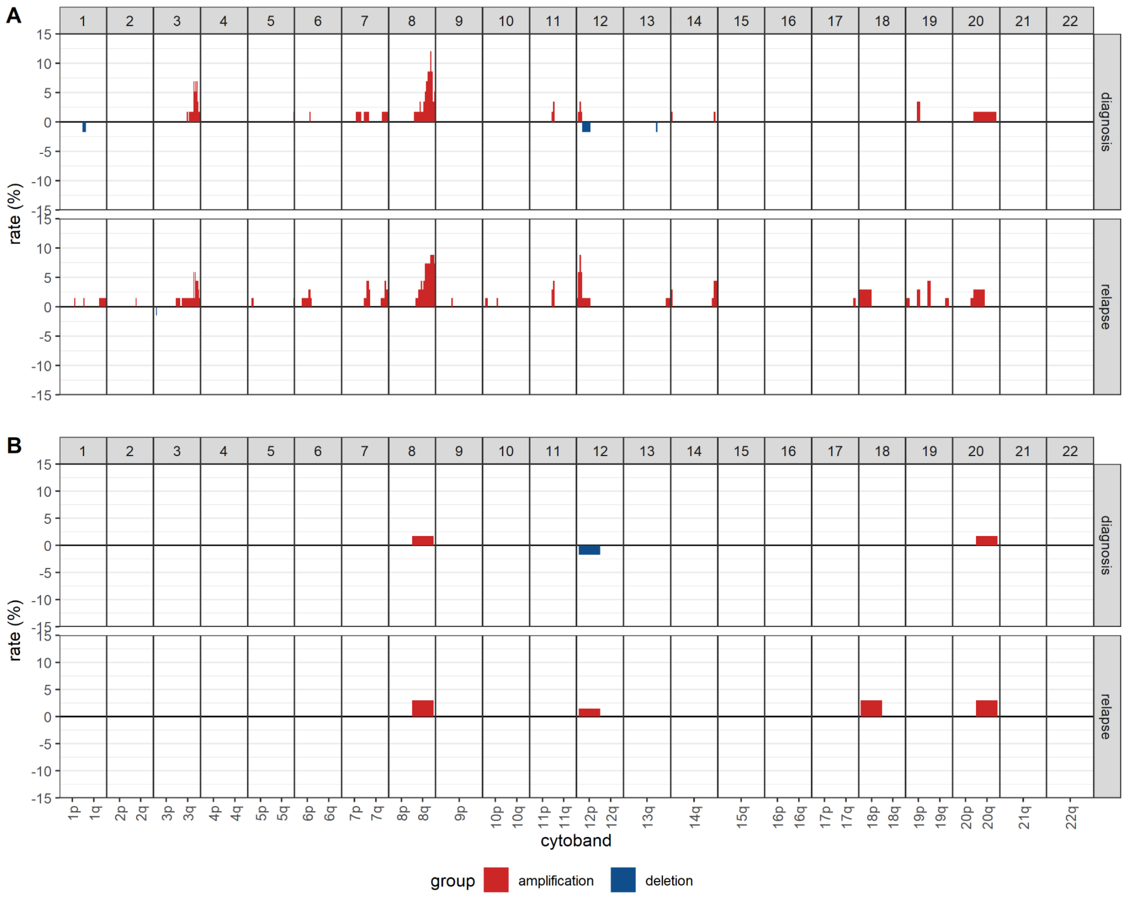

***FIgure S12: Cytoband and chromosome arm alteration rates***

***A*** *- Copy number alteration rates for cytoband-resolution where each cytoband is assessed independently between diagnosis and relapse tumour groups. X-axis denotes each cytoband across each chromosome (noted by axis facets) where 80% of bins supported a CNA call.*

***B*** *- Copy number alteration rates for arm-resolution where each arm is assessed independently between diagnosis and relapse tumour groups. X-axis denotes each cytoband across each chromosome (noted by axis facets) where 50% of bins supported a CNA call.*

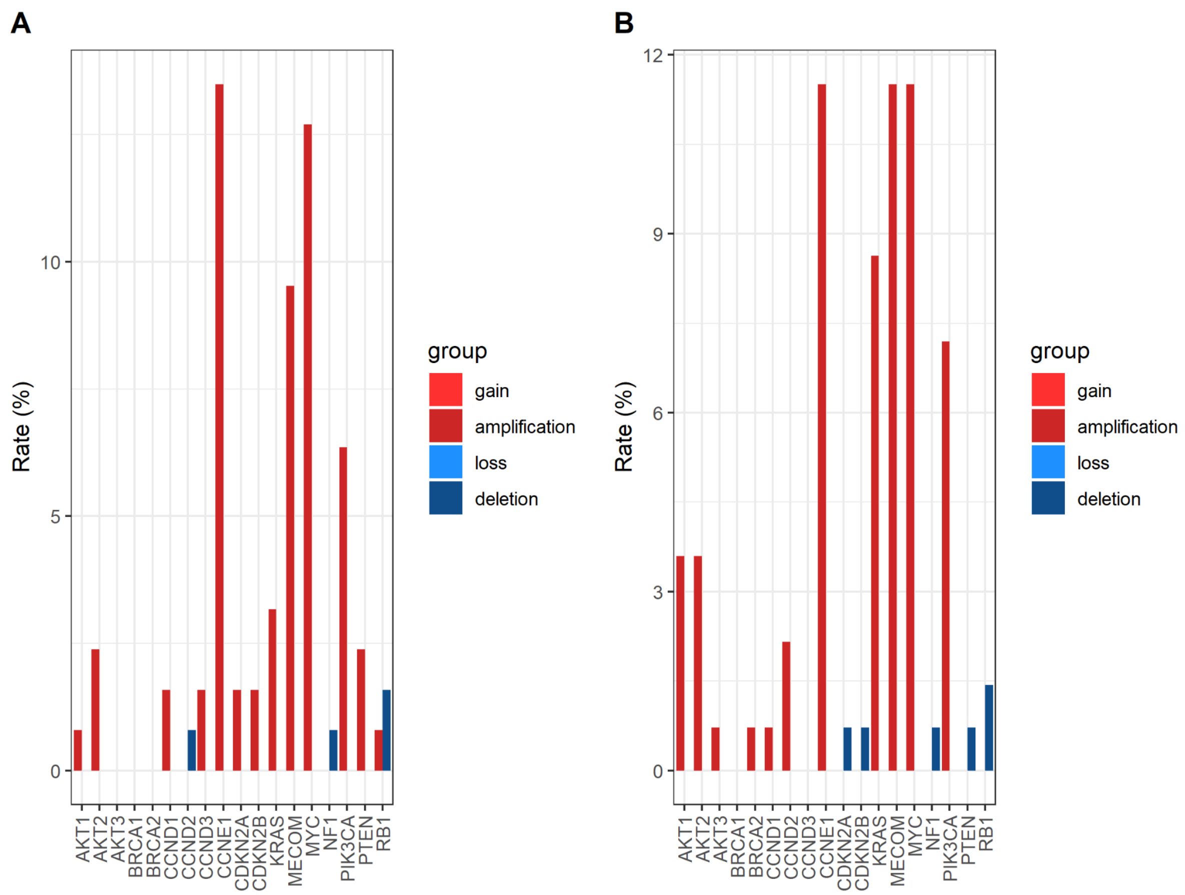

***Figure S13. Amplification and deletion rates across BriTROC-1 samples***

***A** - Copy number alteration rates for eighteen recurrently altered genes across diagnosis samples, stratified by copy number event type (amplification/deletion).*

***B** - Copy number alteration rates for eighteen recurrently altered genes across relapse samples, stratified by copy number event type (amplification/deletion).*

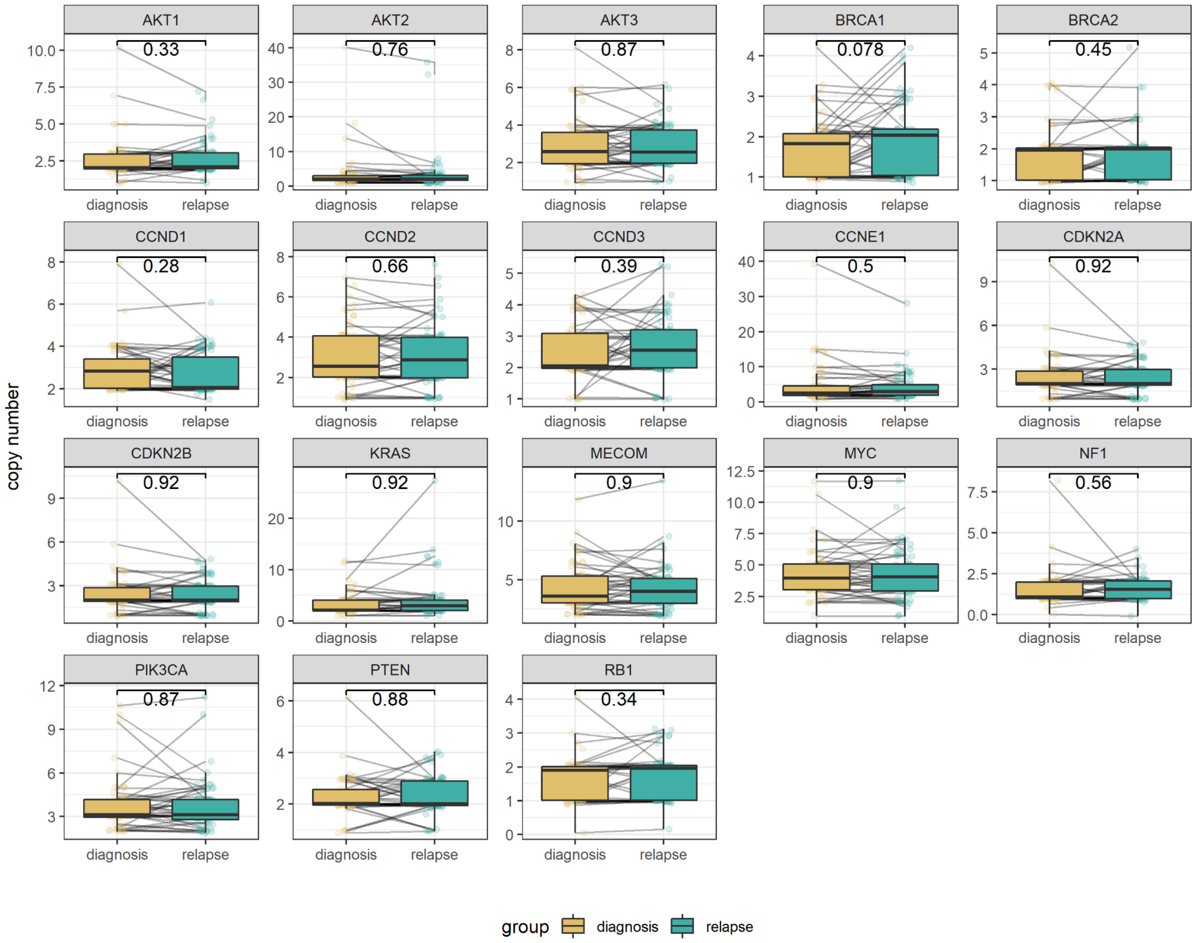

***Figure S14. Total gene copies***

*Absolute copy number state distributions for the list of 18 frequently altered genes between paired diagnosis and relapse samples (n = 58 & n = 68, diagnosis and relapse, respectively). No statistically significant difference was found between the diagnosis and relapse group when comparing the distributions of copy number states over each gene locus (Mann-Whitney U test).*

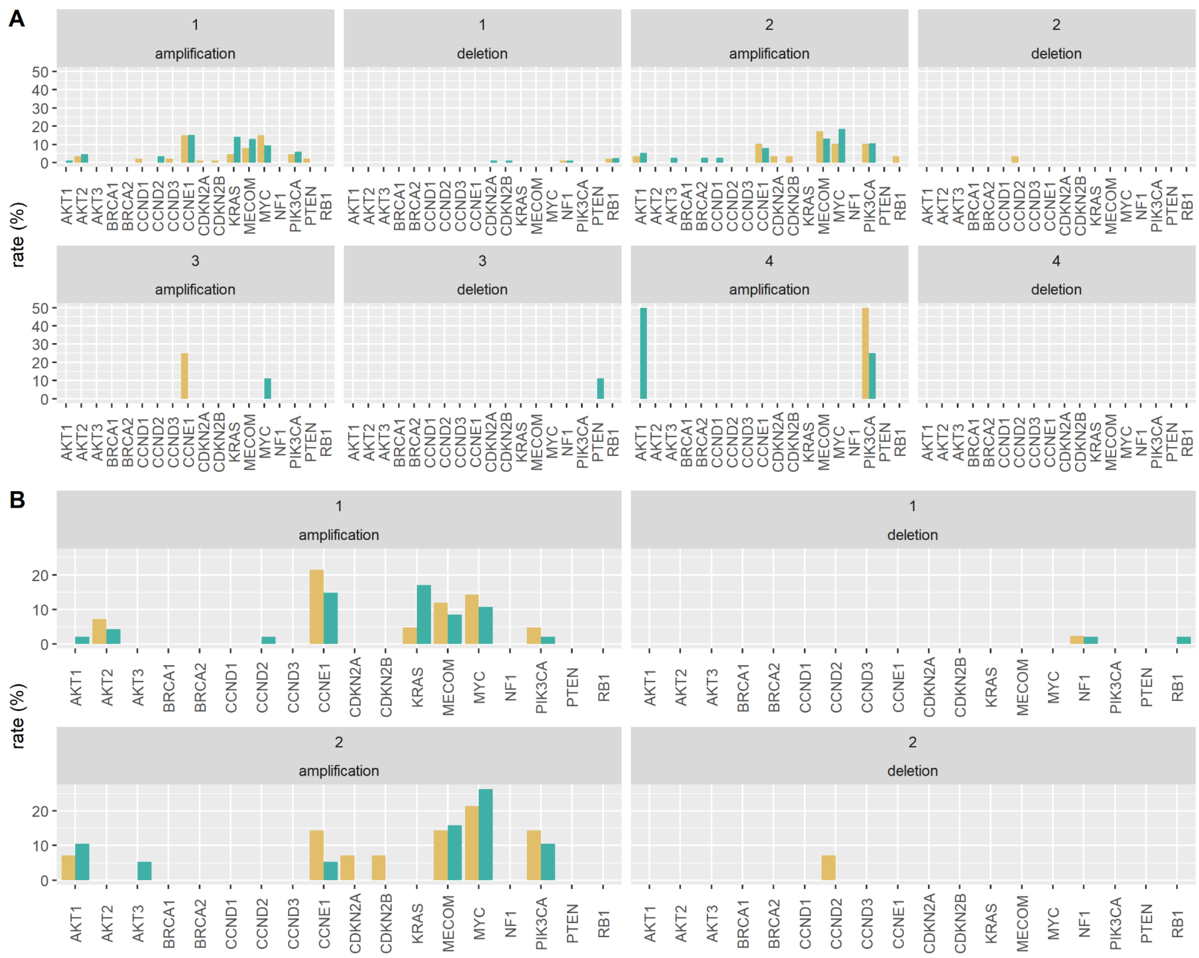

***Figure S15: Copy number focal changes in frequently altered genes stratified by prior lines of therapy***

***A** - Frequency of focal amplification and deletions in frequently altered genes stratified by diagnosis or relapse and number of prior lines of chemotherapy at study registration in all samples.*

***B** - Frequency of focal amplification and deletions in frequently altered genes stratified by diagnosis or relapse and number of prior lines of chemotherapy at study registration in paired samples.*

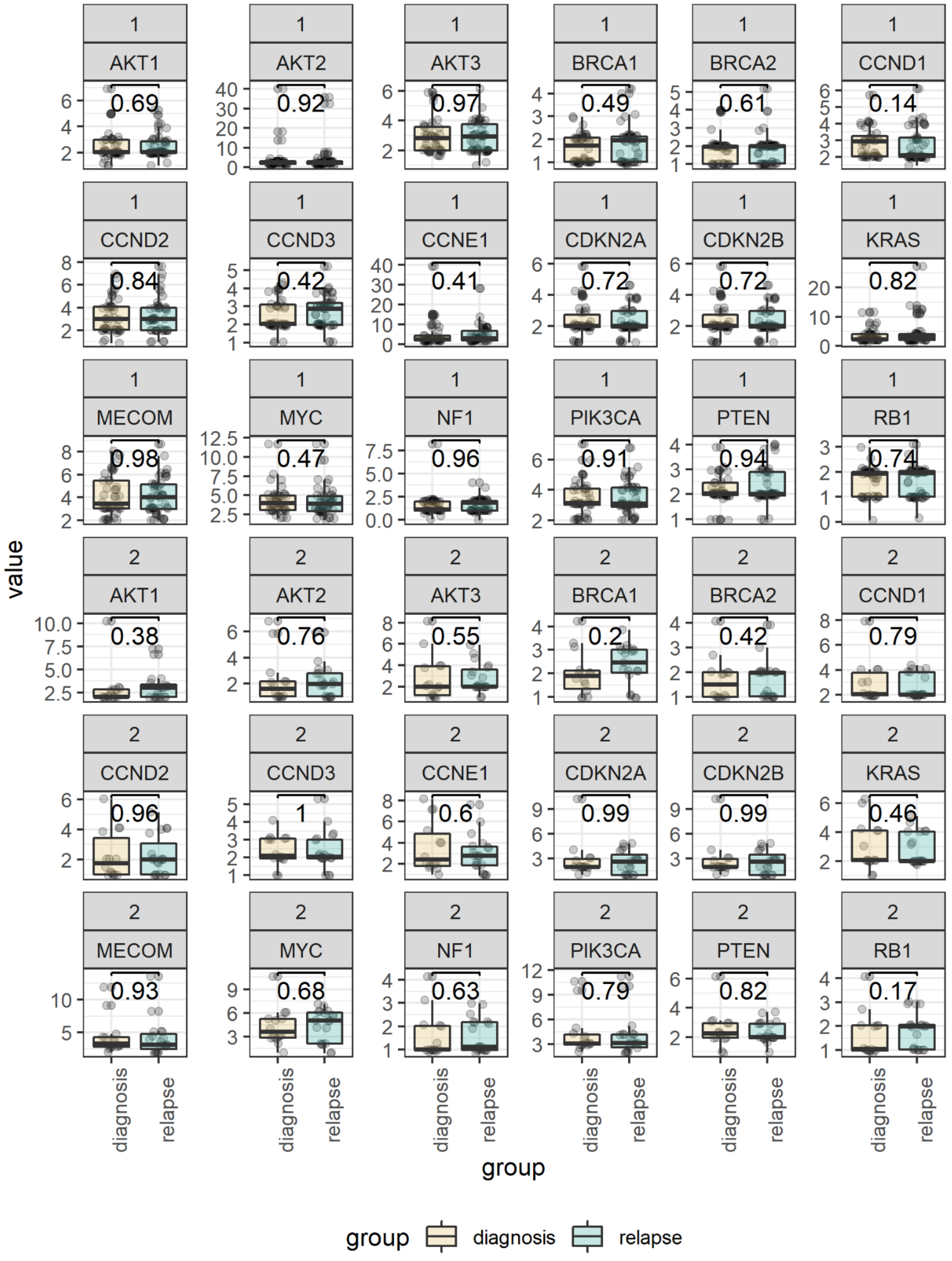

***Figure S16 Stratified copy number count by prior lines and tumour timepoint***

*Faceted boxplot of copy number state distributions for 18 clinically relevant / frequently altered genes comparing diagnosis and relapse tumours, stratified by either one or two prior lines of therapy. Displayed p-values are the uncorrected statistical outcome of a Mann-Whitney U test (n = 42, n = 14, n = 47, n = 19, diagnosis-1 prior, diagnosis-2 prior, relapse-1 prior, relapse-2 prior, respectively)*

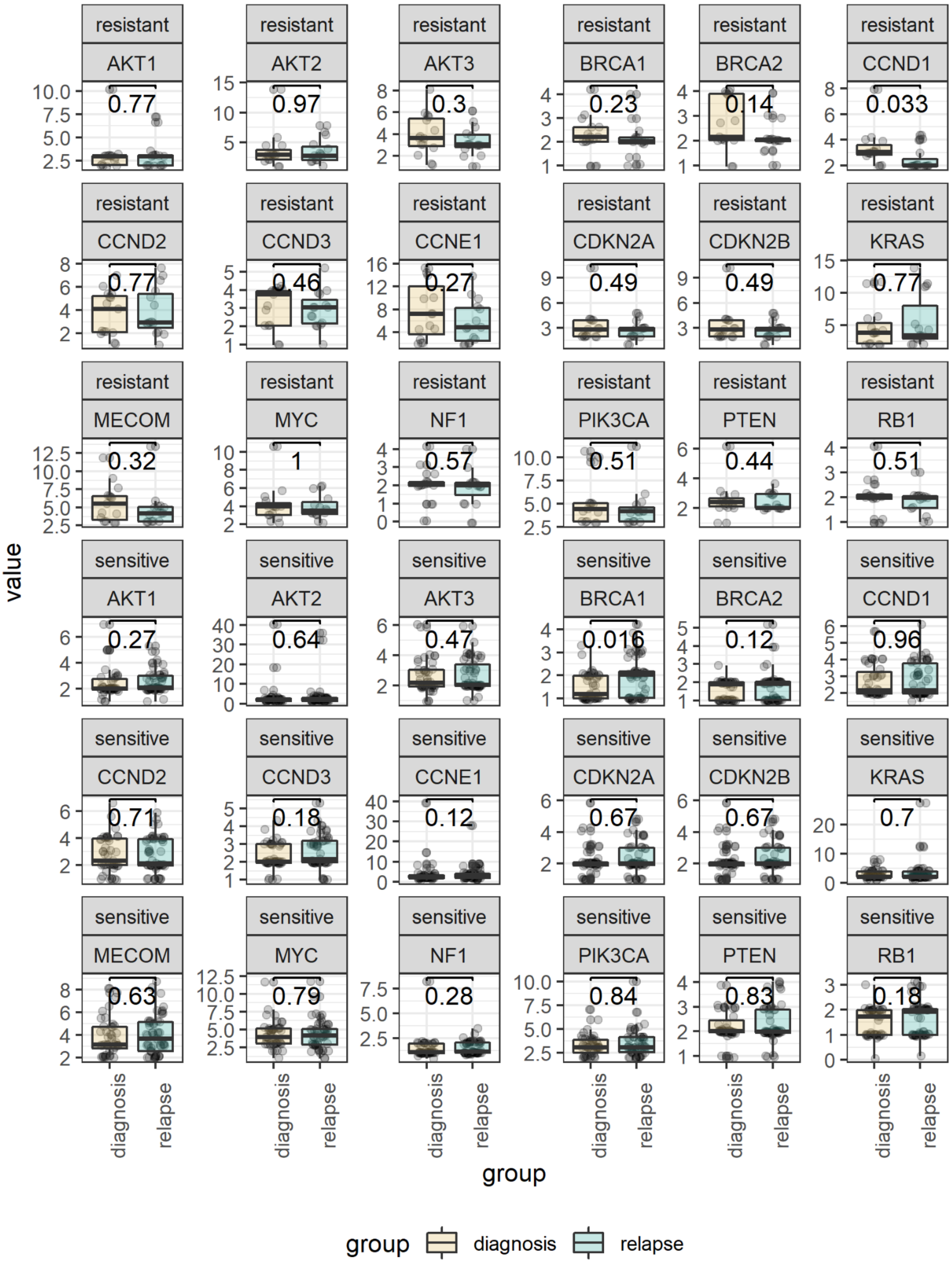

***Figure S17 Stratified copy number count by platinum status and tumour timepoint***

*Faceted boxplot of copy number state distributions for 18 clinically relevant / frequently altered genes comparing diagnosis and relapse tumours, stratified by resistance or sensitivity to platinum-based chemotherapy. Displayed p-values are the uncorrected statistical outcome of a Mann-Whitney U test (n = 15, n = 43, n = 15, n = 53, diagnosis-resistant, diagnosis-sensitive, relapse-resistant, relapse-sensitive, respectively).*

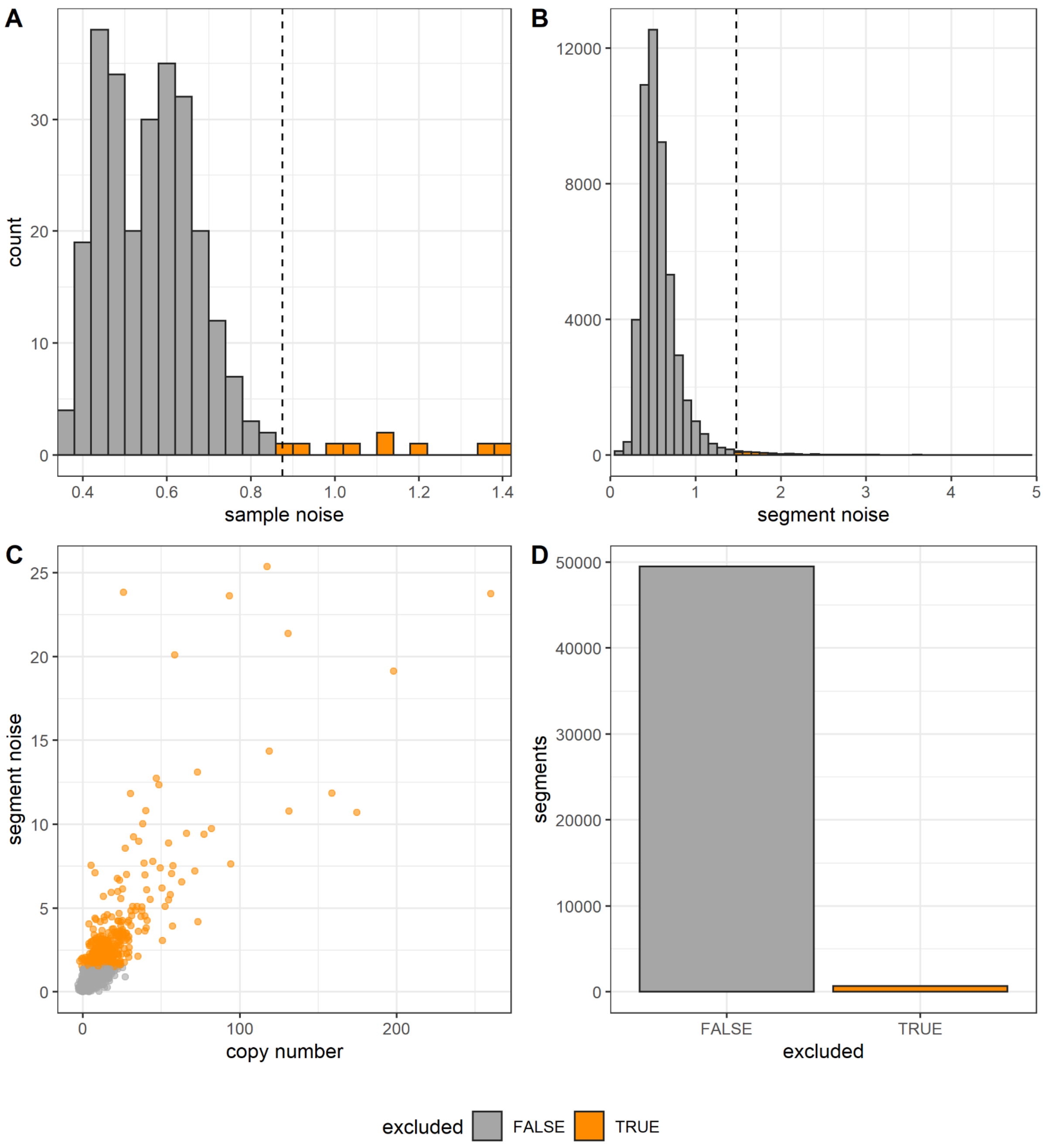

***Figure S18. Intra-tumoural heterogeneity filtering***

***A*** *- Histogram of mean segment noise across each sample. Vertical dashed line indicates a value of two standard deviations above the mean. Bars highlighted in orange (right of dashed line) were removed from further ITH analysis.*

***B*** *- Histogram of segment noise. Vertical dashed line indicates a value of two standard deviations above the mean. Bars highlighted in orange (right of dashed line) were removed from further ITH analysis.*

***C*** *- Plot visualising the distribution of copy number value against segment noise. Points with orange colouration were those exceeding the segment noise cutoff.*

***D*** *- Bar plot visualising the proportion of total segments that were excluded from ITH by segment noise filtering.*

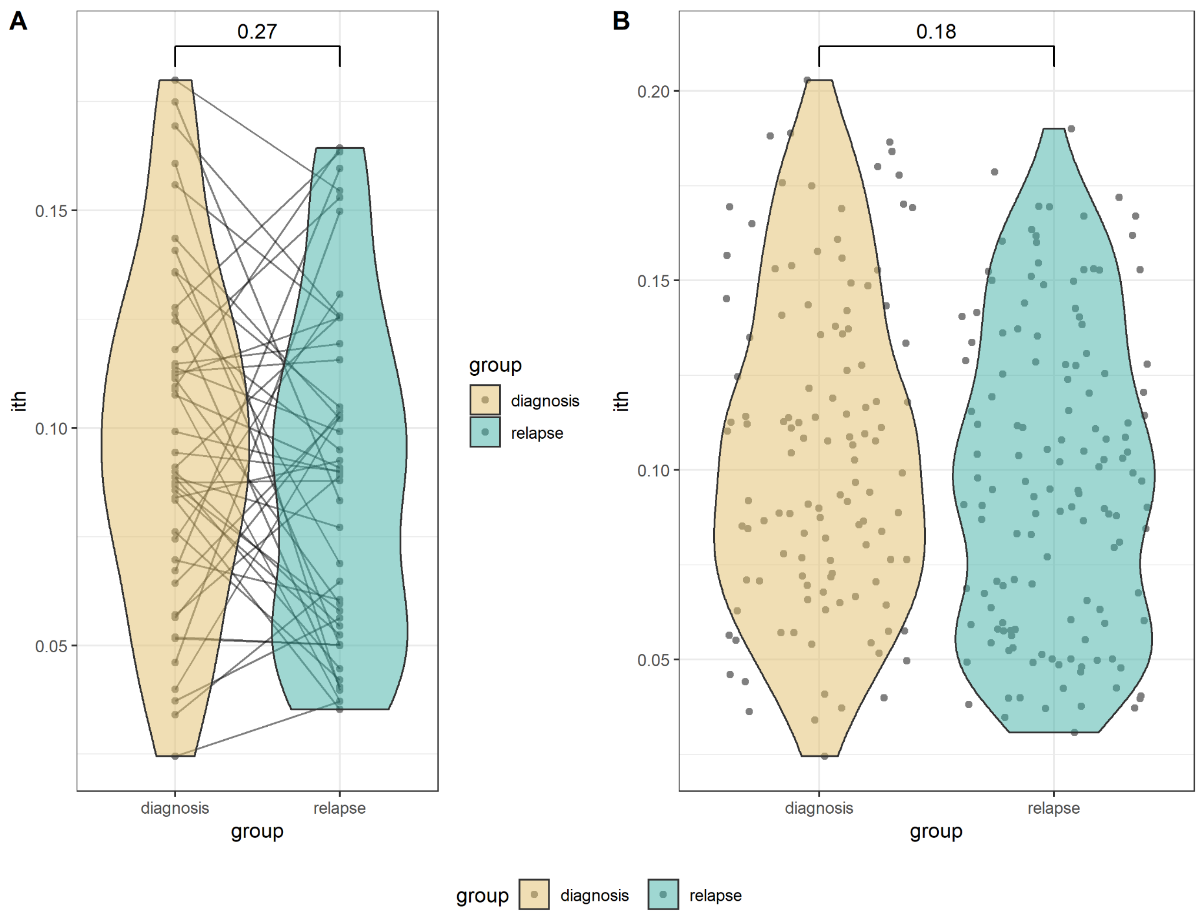

***Figure S19. Intra-tumoural heterogeneity***

***A** - Violin plot comparing the patient-level comparison of estimated ITH as calculated from integer copy number segment deviations stratified by diagnosis and relapse samples (n = 46). Distributions were not shown to be non-statistically significant by Wilcoxon signed-rank test utilising paired comparison.*

***B** - Violin plot comparing the unpaired sample-level comparison of estimated ITH as calculated from integer copy number segment deviations stratified by diagnosis and relapse samples (n = 119 & 137, diagnosis and relapse, respectively). Distributions were not shown to be non-statistically significant by Wilcoxon signed-rank test utilising paired comparison.*

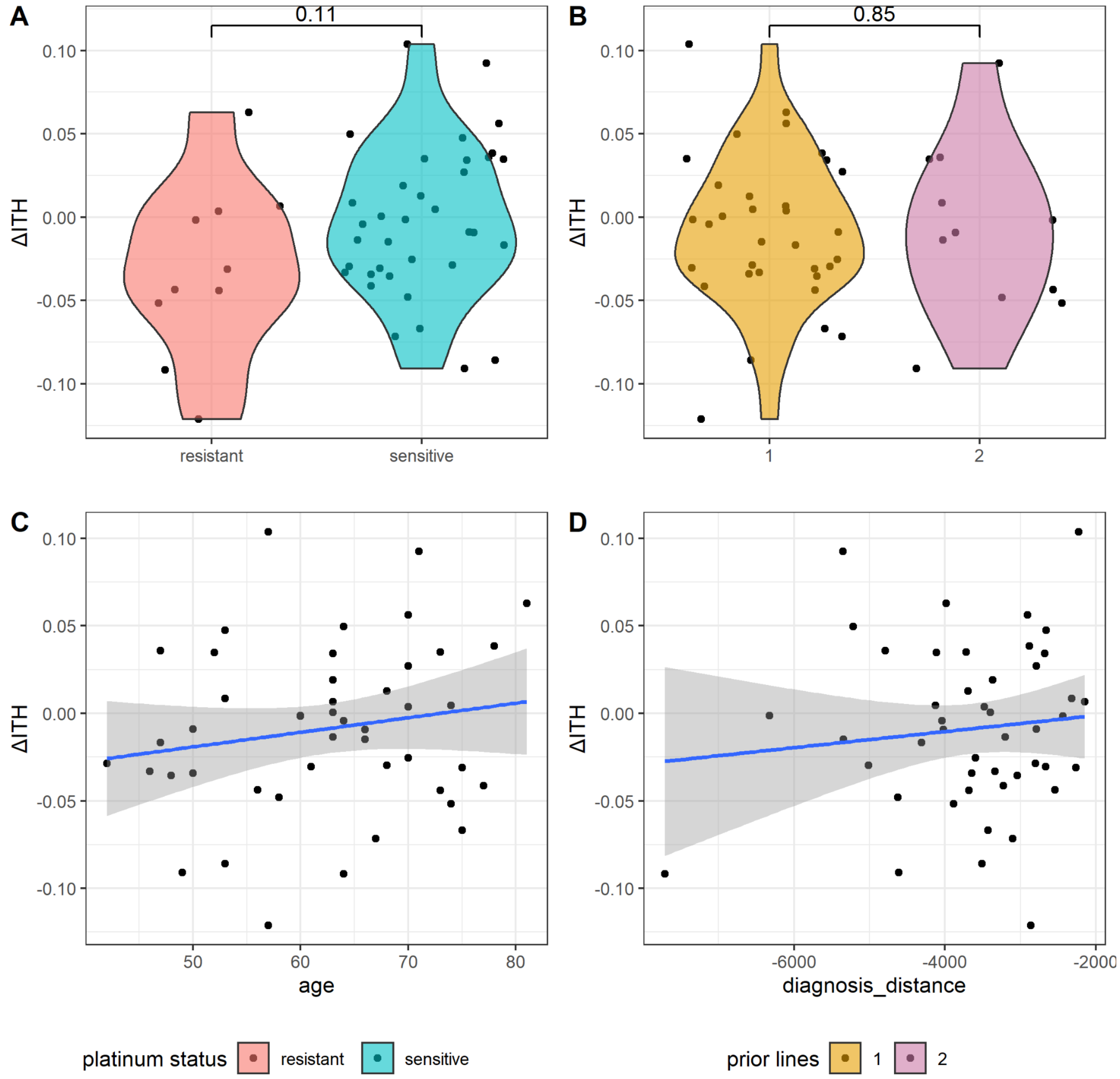

***Figure S20: Intra-tumoural heterogeneity change***

***A** - Violin plot comparing the patient-level comparison of estimated ΔITH as calculated from integer copy number segment deviations stratified by platinum-based therapeutic resistance or sensitivity (n = 9 & n = 35). Distributions were not shown to be non-statistically significant by Mann-Whitney U test.*

***B** - Violin plot comparing the patient-level comparison of estimated ΔITH as calculated from integer copy number segment deviations stratified by prior lines of therapy before study entry (n = 33 & n = 11), two patients had three and four prior lines of therapy, respectively, so were dropped from statistical testing.*

***C** - Scatter plot of estimated ΔITH to patient age demonstrating limited correlation of changing tumour heterogeneity with age at diagnosis*

***D** - Scatter plot of estimated ΔITH to distance from diagnosis, as a proxy for sample age. The linear regression demonstrates limited correlation of changing tumour heterogeneity with age at diagnosis.*

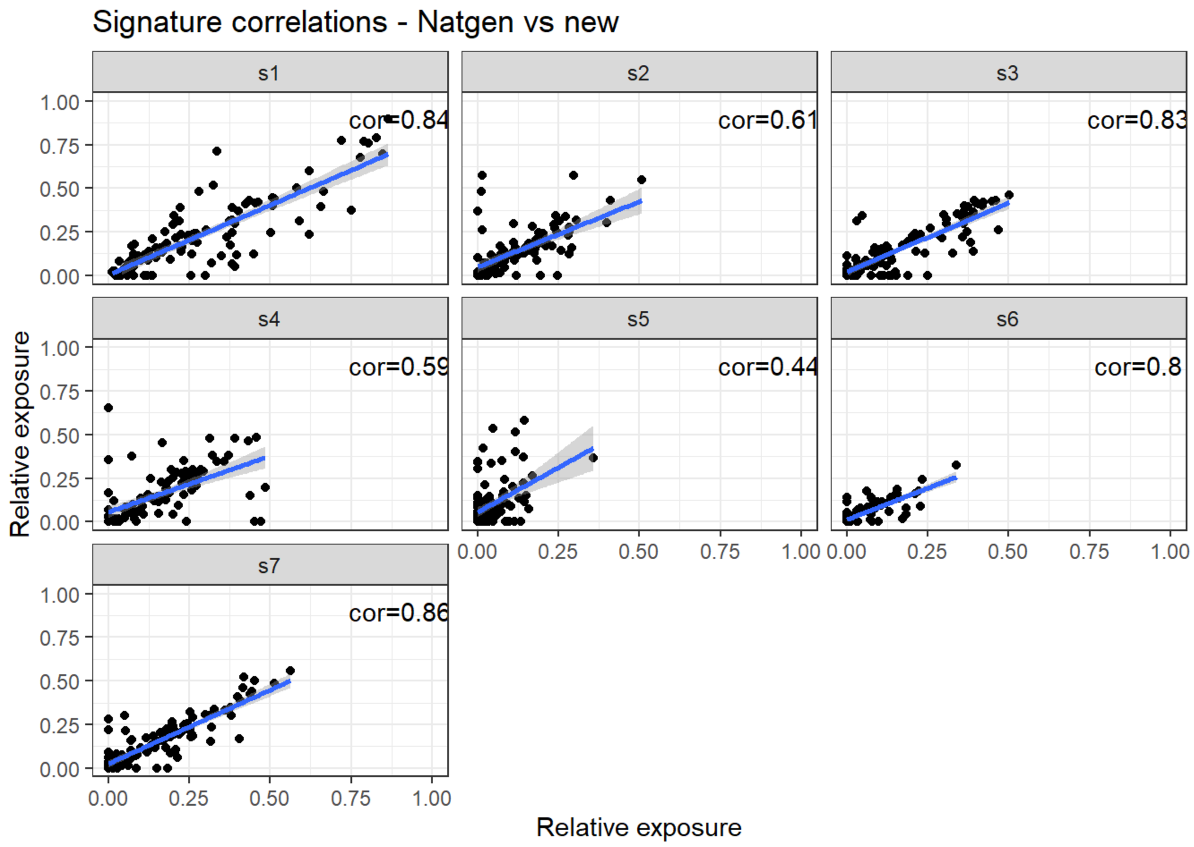

***Figure S21. Copy number signature correlations***

*Visualisation of the overall correlation between previously generated HGSC copy number signatures {Macintyre, 2018 #6710} and those generated here. Copy number signatures correlate though exposures for any given signature are variable due to different absolute copy number fitting methodologies between this study and the previous data.*

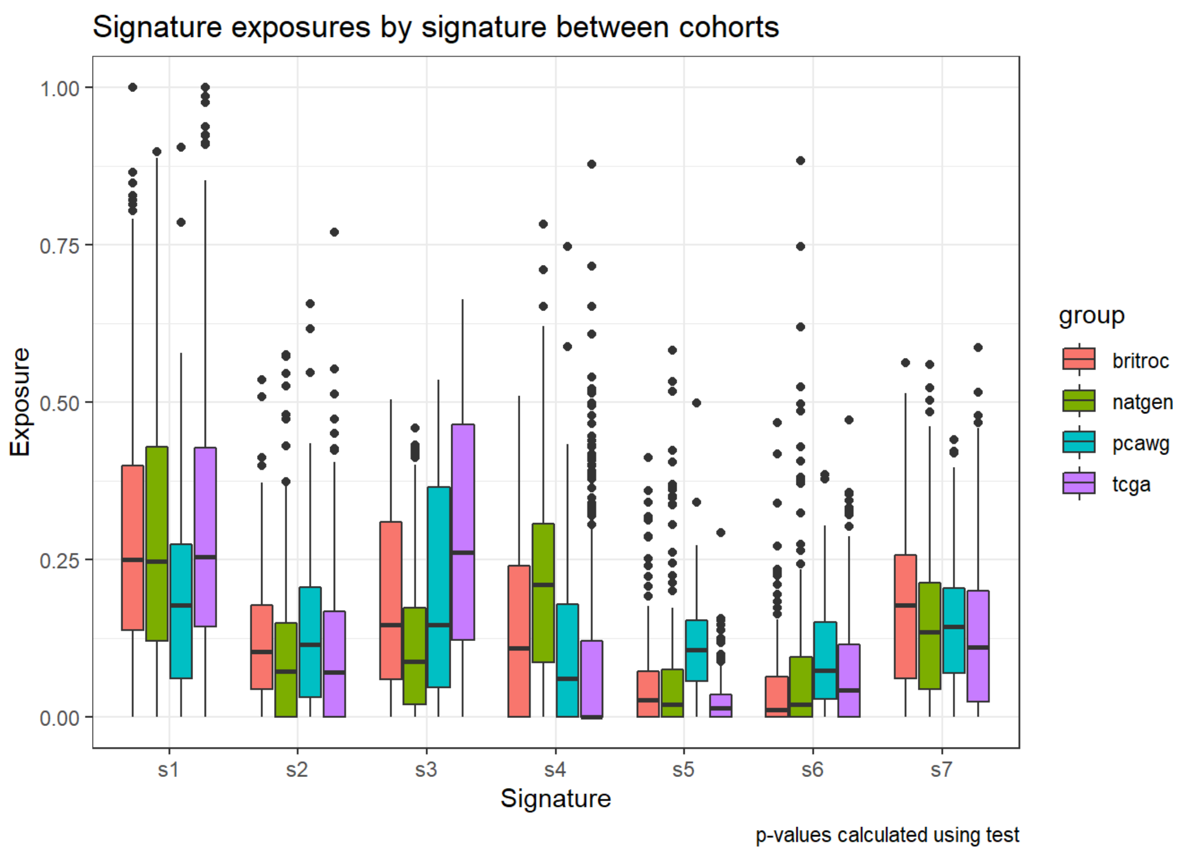

***Figure S22. Copy number signature exposure comparison to other HGSC cancer datasets***
*Comparison of copy number signature exposures for HGSC datasets across this study (Red - BriTROC), Macintyre et al. (Green-Natgen; subset of samples from this study with differing copy number methodology){Macintyre, 2018 #6710}, Pan-cancer analysis of Whole Genomes (Blue - PCAWG){ICGC/TCGA, 2020 #7462}, and The Cancer Genome Atlas (Purple - TCGA) {Weinstein, 2013 #5848}. Rates across differing HGSO cohorts are broadly stable despite differing sample distributions, features, and methodologies implemented prior to signature extraction*

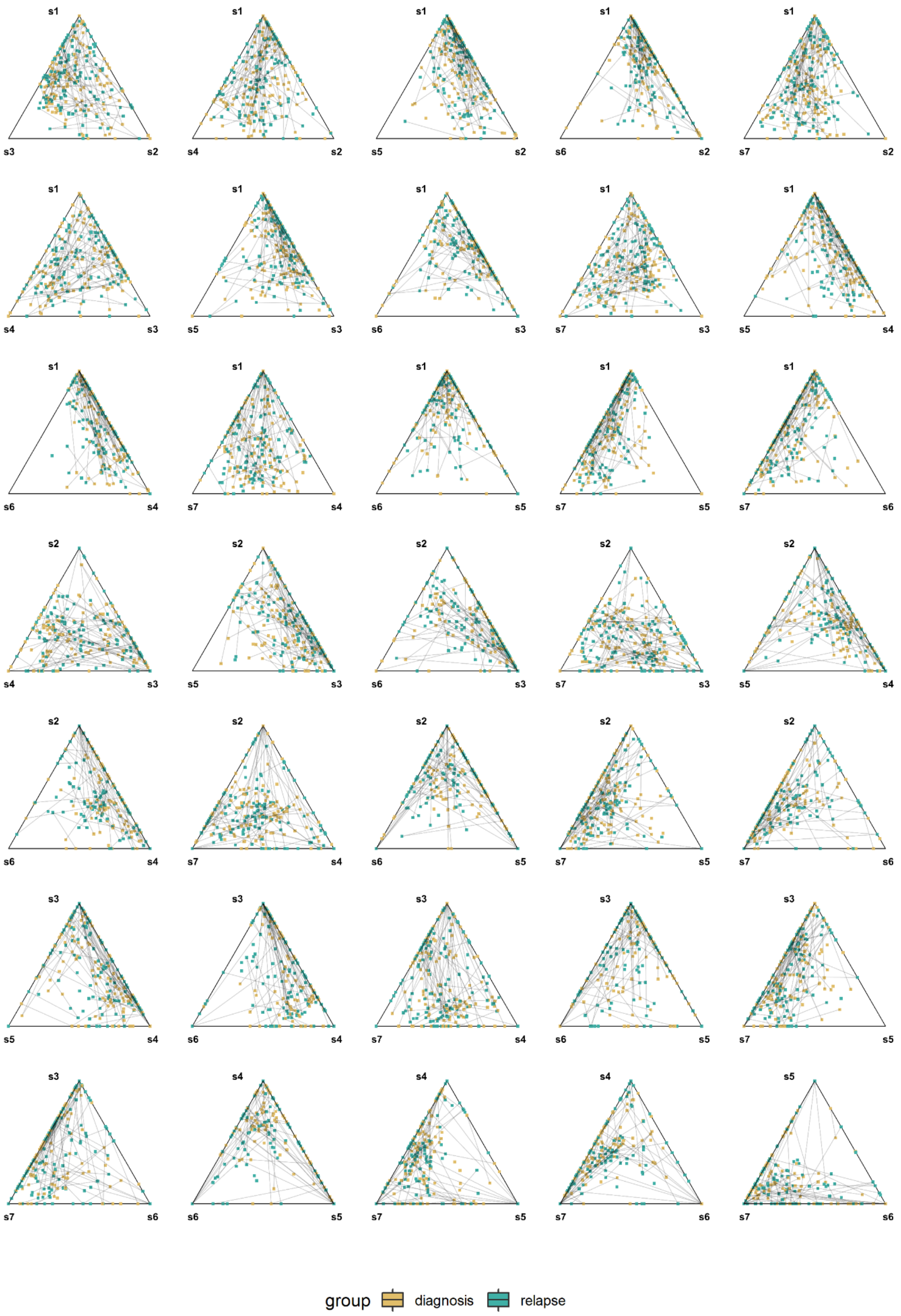

***Figure S23. Copy number signature simplex plots***

*Simplex plots describing the compositional copy number signature exposures between trios of copy number signatures. Colouration indicates the tumour grouping between diagnosis and relapse. Connecting lines indicate the intra-patient sample pairing, demonstrating the signature change between diagnosis and relapse.*

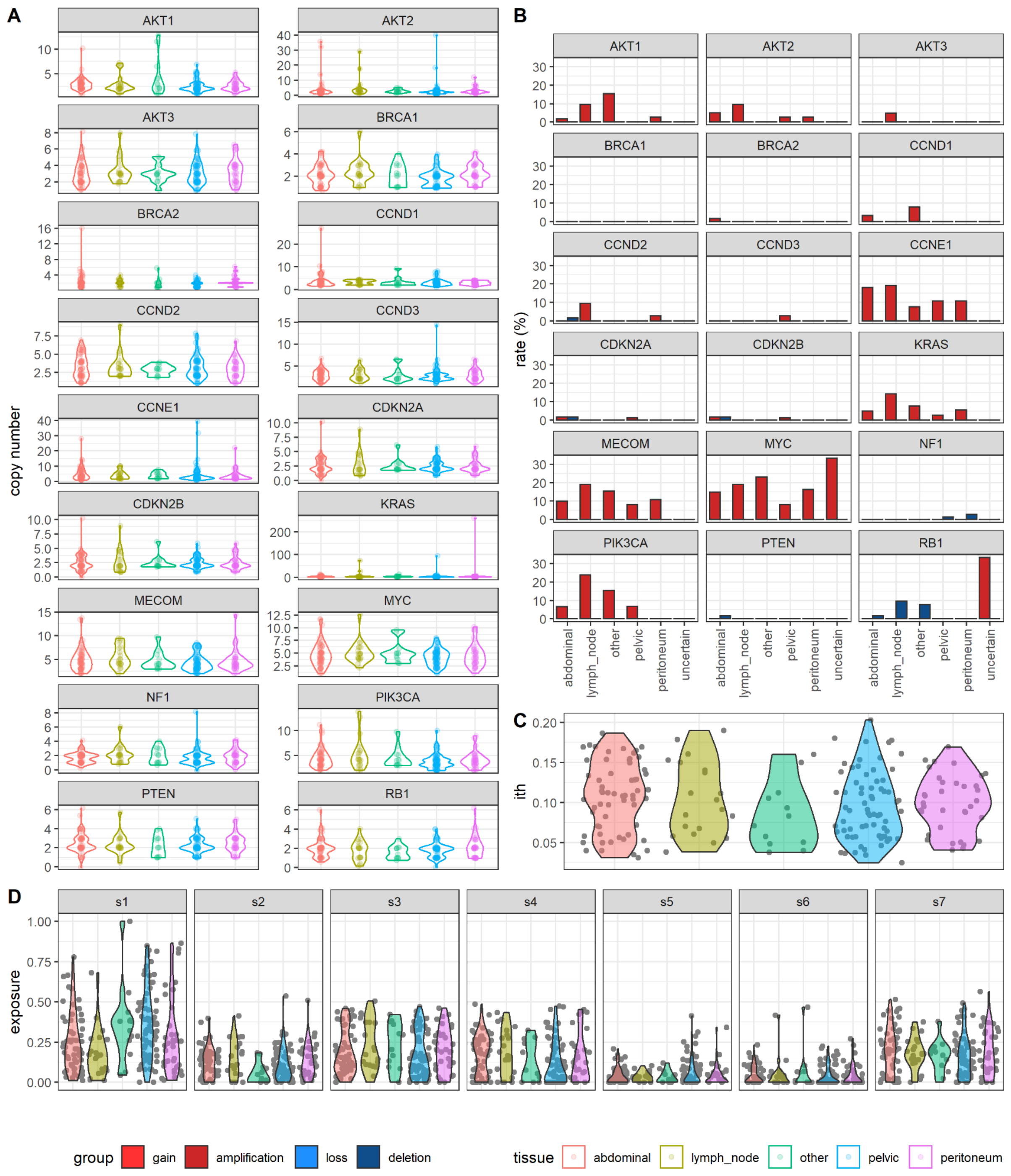

***Figure S24. Copy number features by tissue site of origin***

***A** - Copy number count summaries across 18 clinically relevant / frequently altered genes across tumour tissue site. Violin plots are colour by tissue and represent the distribution of copy number value for a given gene across all available samples.*

***B** - copy number alteration rates across 18 clinically relevant / frequently altered genes across tumour tissue site. Bar plots are colour by amplification and deletion rate for a given gene across all available samples.*

***C** - Distribution of ITH for each sample stratified by available tumour tissue type.*

***D** - Distributions for each copy number signature. Violin plots are colour by tissue and represent the distribution of copy number signature across all available samples by tumour tissue site. (sample n; intra-abdominal = 42, lymph = 9, other = 7, pelvic = 72, peritoneum = 19).*

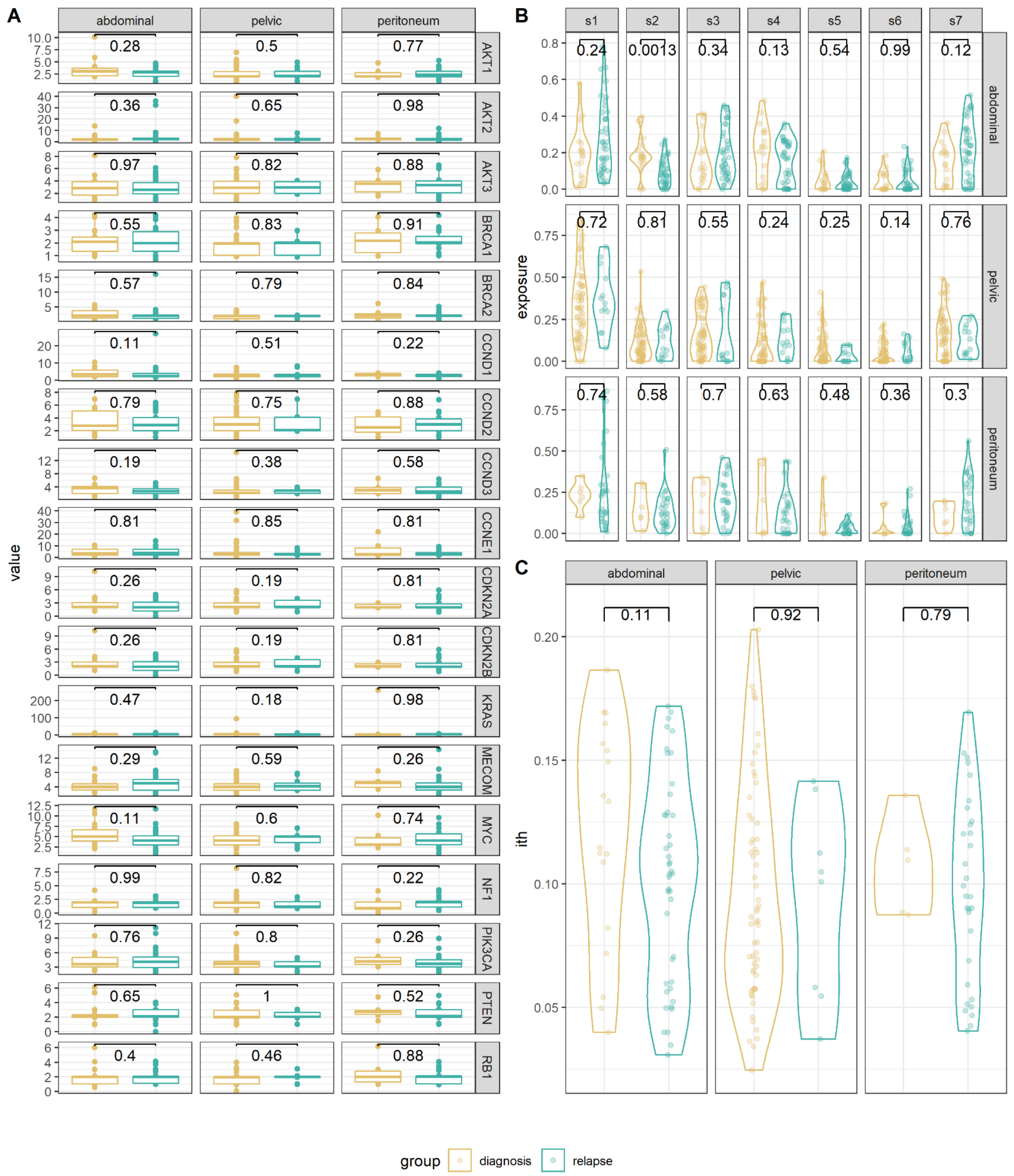

***Figure S25. Copy number features stratified by diagnosis and relapse across tissue site of origin***

***A** - Copy number count summaries across 18 clinically relevant / frequently altered genes across tumour tissue site. Box plots are colour by diagnosis and relapse sample status and represent the distribution of copy number value for a given gene (Mann-Whitney U test).*

***B** - Distributions for each copy number signature. Violin plots are colour by diagnosis and relapse sample status and represent the distribution of copy number signature across tumour tissue site.*

***C** - Distribution of ITH for each sample stratified by available tumour tissue type (sample n; intra-abdominal = 20 & 22, pelvic = 67 & 5, peritoneum = 4 & 15, diagnosis and relapse, respectively).*

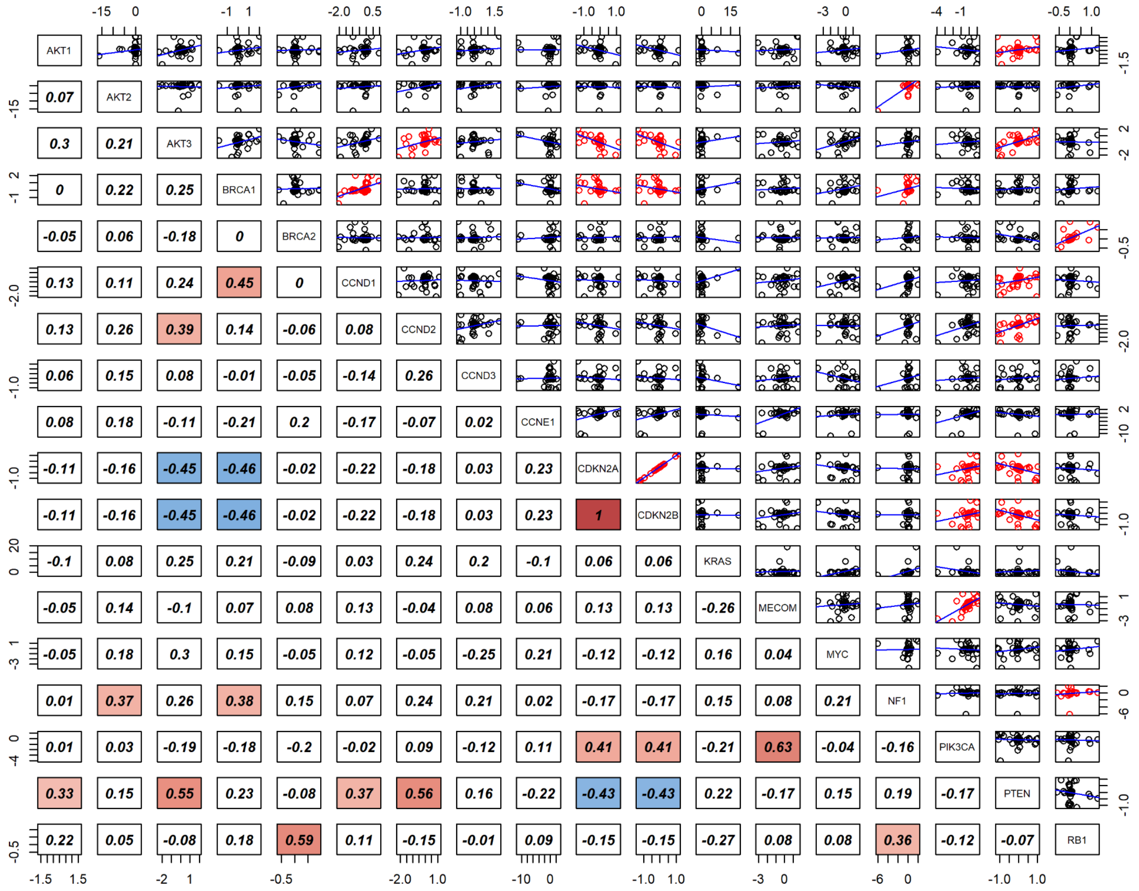

***Figure S26. Copy number change correlations in frequently altered genes***

*Correlation and scatter plot matrix of copy number change correlation between different clinically relevant / frequently altered genes. Scatter plots show the point distribution for any given pair of genes labelled in the diagonal. The blue line for each scatter plot in the linear fit for the given set of points; points in red are significantly correlation (spearman rank correlation). Numerical values in the lower portion state the correlation coefficient for a given gene pairing using the same colouration ascribed in Figure 6B.*

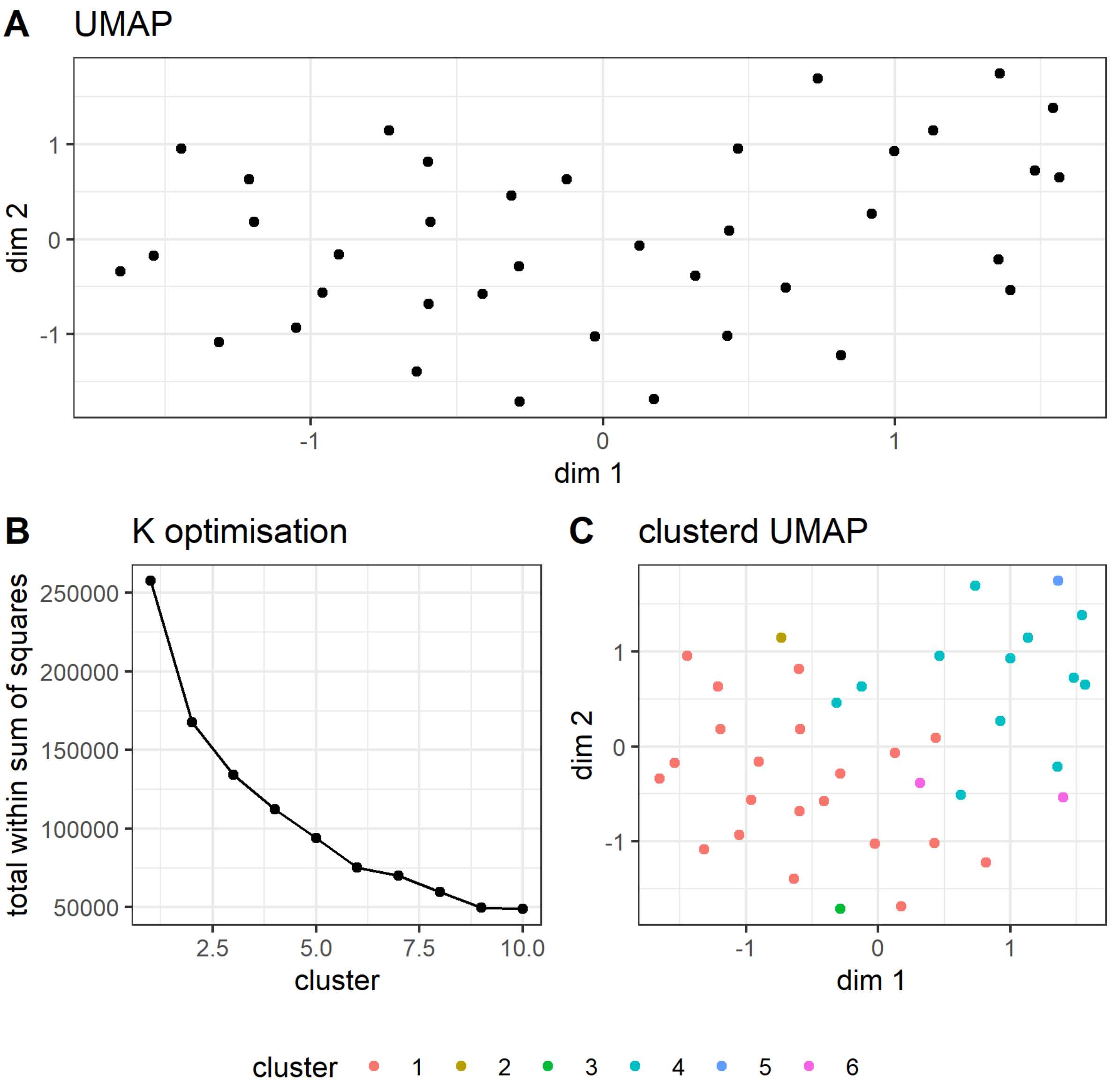

***Figure S27. Copy number change matrix clustering***

***A** - UMAP dimensional reduction of the copy number change matrix shown in figure 6A. No obvious patterns of patient clustering can be identified.*

***B** - Visualisation total within sum of squares calculation for cluster numbers 1 through 10 for k-means clustering. This process should typically identify an "elbow" to select as the optimal number of clusters.*

***C** - Re-visualisation of the UMAP dimensional reduction with the purported optimal k-means clusters which demonstrated little to no clustering of patients.*

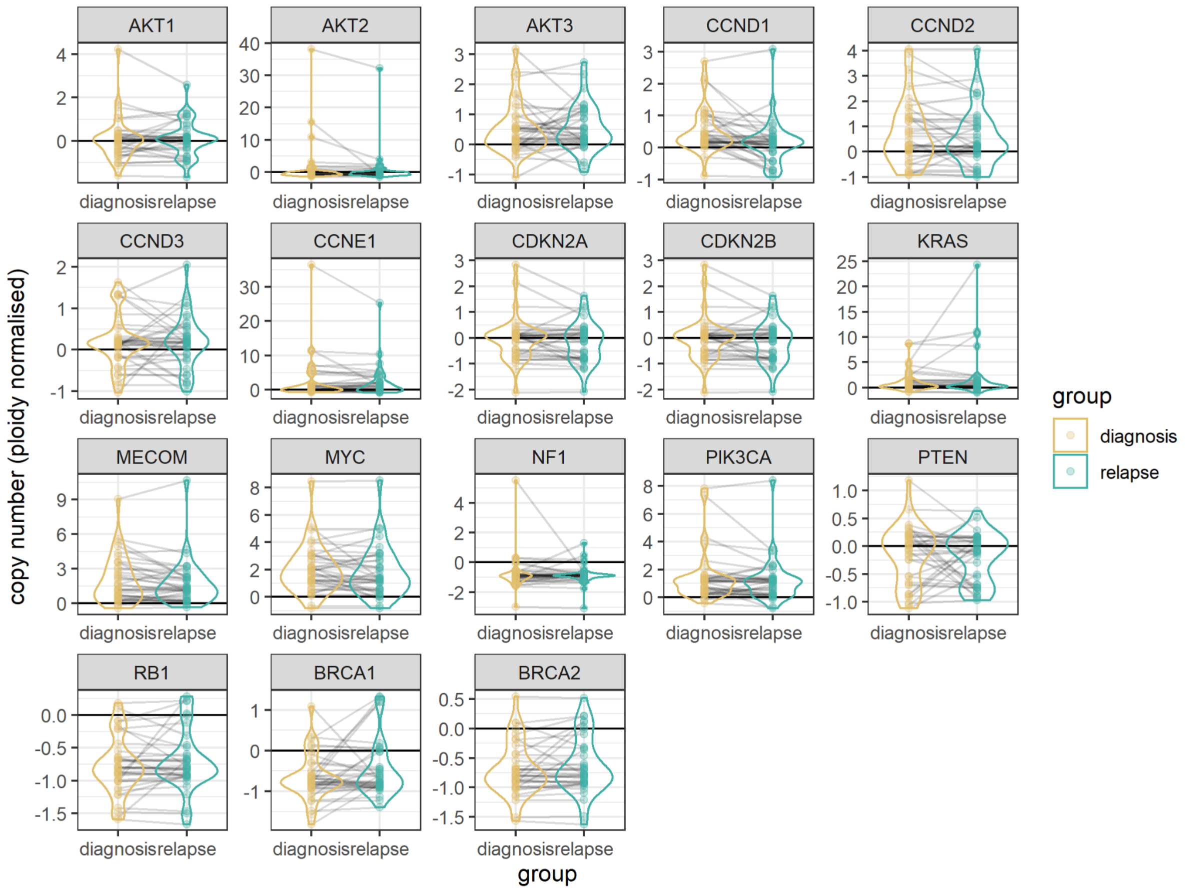

***Figure S28. Ploidy normalised copy number change across frequently altered genes***

*Copy number states of frequently altered / clinically relevant genes after ploidy normalisation (gene copy number - sample group ploidy), stratified by diagnosis and relapse groups. Violin plots visualise the distribution of copy number values between diagnosis and relapse groups, grey lines between points indicate the copy number change and direction of change between the diagnosis and relapse groups for a given patient. Black horizontal line indicates the zero change point where points at this line have gene copy numbers identical to the sample ploidy.*

***Figure S29. Individual patient data***

*A full compendium of patient vignettes is provided as an additional supplementary data file (supplemental data - Figure_S29_patient_vignettes.pdf). Each vignette presents clinical timeline, genome-wide copy number change, absolute copy number change for the eighteen recurrently altered genes, ITH, and copy number signature exposure for one patient. Red dots indicate data points for the individual patient compared to the whole cohort. PLD: peglyated liposomal doxorubicin.*

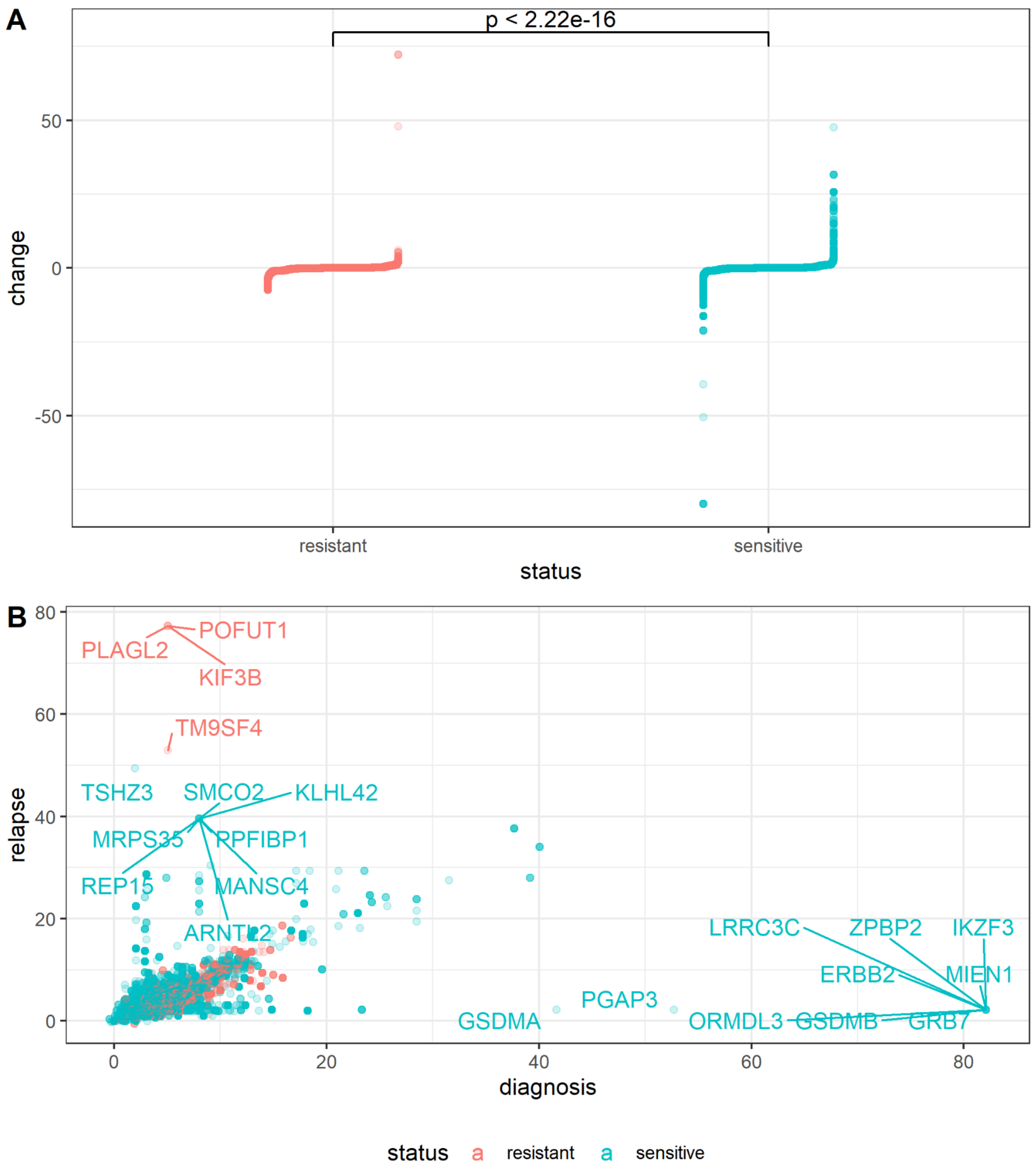

***Figure S30. Copy number change by response to platinum-based therapeutics spread***

*Plots demonstrating the increased number of 'extreme' copy number focal changes between resistant and sensitive HGSC patients (n = 9 & n = 29, resistant and sensitive patients, respectively).*

***A** - Jittered point plot of gene change between diagnosis and relapse patients stratified by response to platinum-based chemotherapeutics showing that sensitive patients have a wider distribution, corresponding to a greater number of extreme changes.*

***B** - Scatter plot of diagnosis versus relapse values for all gene loci coloured by response to platinum-based chemotherapeutics showing the wider spread of values above and below the diagonal.*

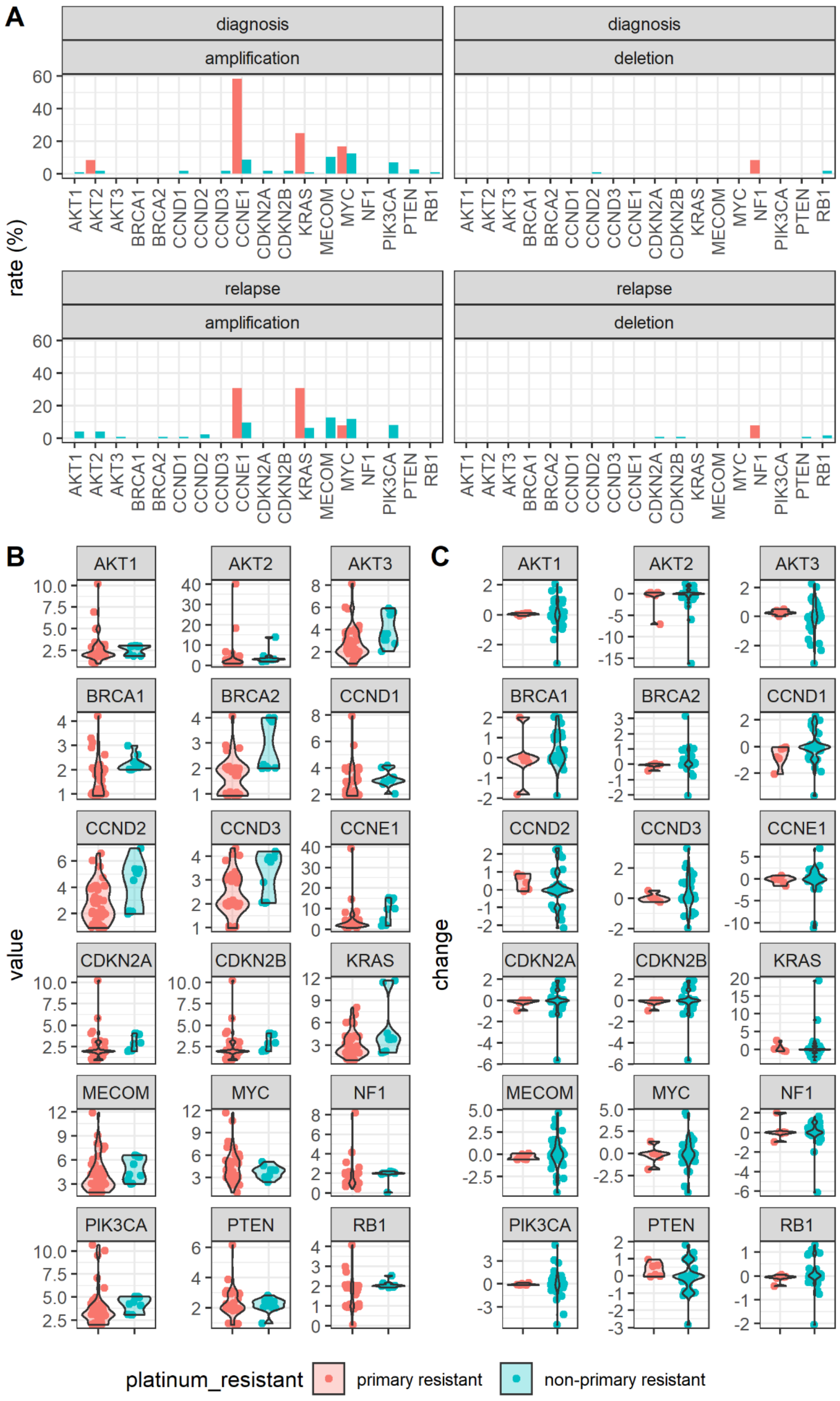

***Figure S31 Copy number alteration rates between primary platinum resistant samples***

***A** - Computed copy number alteration rate for clinically relevant / frequently altered genes. Faceted bar plot compares the copy number rate between primary platinum resistant samples against all others, stratified by tumour time point and copy number event type (n = 114, n = 12, n = 126 & n = 13; diagnosis-non-primary resistant, diagnosis-primary resistant, relapse-non-primary resistant, relapse-primary resistant, respectively).*

***B** - Copy number state violin distribution for clinically relevant / frequently altered genes comparing primary platinum resistant diagnosis samples versus other diagnosis samples (n = 6 & n = 41, respectively).*

***C** - Copy number violin change distribution for paired samples showing the changning copy number state for clinically relevant / frequently altered genes comparing primary platinum resistant patients versus other patients (n = 6 & n = 41, respectively).*

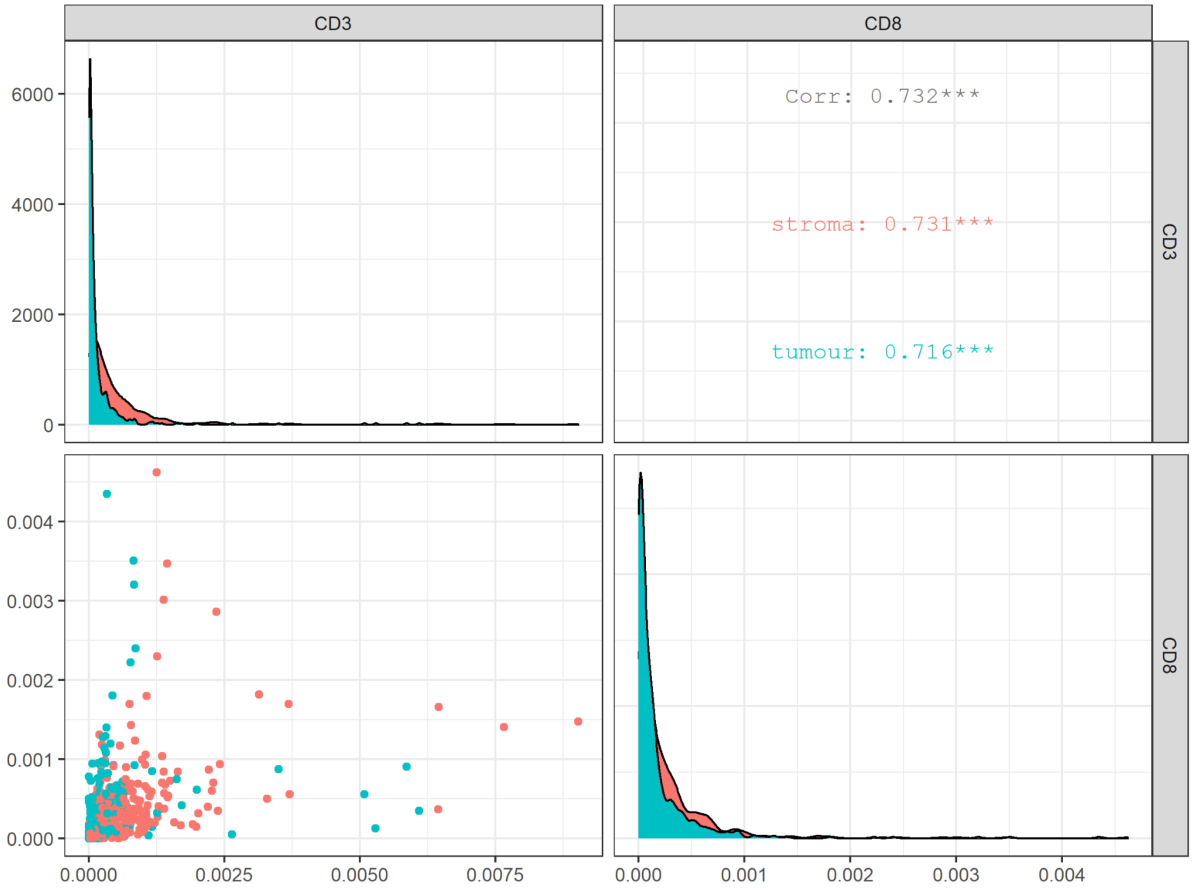

***Figure S32 IHC-derived Immune marker cell density***

*Paired correlation plot for IHC-derived immune markers CD3 and CD8 cell density across tissue groups tumour and stroma shown by green and red colouration, respectively. All markers were positively correlated with each other, including when stratified by tissue group, with a greater positive correlation being present in stromal regions compared with tumour regions (CD3 tumour n = 474, n = 474, n = 660, & n = 653; CD3-stroma, CD3-tumour, CD8-stroma, and CD8-tumour observations, respectively).*

### *Tables*

***Table S1 - Amplicon panel information***

***Table S2. FIGO stage at time of diagnosis.***

| ***Original FIGO stage**** | ***Number*** | ***%*** |
|---|---|---|
| *1a* | *3* | *1.1* |
| *1c* | *13* | *4.7* |
| ***Stage I*** | ***16*** | ***5.8*** |
| *2a* | *8* | *2.9* |
| *2b* | *8* | *2.9* |
| *2c* | *7* | *2.5* |
| ***Stage II*** | ***23*** | ***8.3*** |
| *3a* | *8* | *2.9* |
| *3b* | *16* | *5.8* |
| *3c* | *148* | *53.6* |
| *3NOS* | *1* | *0.4* |
| ***Stage III*** | ***173*** | ***62.7*** |
| *4* | *58* | *21.0* |
| *4b* | *2* | *0.7* |
| ***Stage IV*** | ***60*** | ***21.7*** |
| *NK* | *4* | *1.4* |
| ***TOTAL*** | ***276*** | ***100.0*** |

**As reported by recruiting site according classification in place at time of original diagnosis (2008 or 2014)*

***Table S3. Surgery undertaken during first-line treatment and extent of residual disease following first line surgery.***

| ***SURG_TYP_1*** | ***Number*** | | ***Residual disease at first surgery*** | ***Number*** |
|---|---|---|---|---|
| *Interval* | *96* | | *No residual - R0* | *42* |
| *No surgery* | *24* | | *Optimal - R1* | *156* |
| *Primary surgery* | *152* | | *Suboptimal - R2* | *38* |
| *Salvage surgery* | *3* | | *Not known* | *15* |
| *Missing* | *1* | | *No surgery* | *24* |
| ***Total*** | ***276*** | | *Missing* | *1* |
| | | | ***Total*** | ***276*** |

*Table S4A - BriTROC-1 biopsy locations by sample*

| ***Biopsy site summary*** | ***Tissue samples*** | ***DNA Samples*** |
|---|---|---|
| *Peritoneum* | *56* | *62* |
| *Lymph node** | *72* | *85* |
| *Omentum* | *28* | *33* |
| *Colon, mesentery, small bowel, pericolic fat* | *20* | *24* |
| *Liver* | *17* | *20* |
| *Subcutaneous* | *9* | *10* |
| *Gynae organs (uterus, ovary, fallopian tube, vaginal vault)* | *16* | *22* |
| *Peri-splenic* | *3* | *3* |
| *Brain* | *4* | *5* |
| *Pelvis* | *2* | *3* |
| *Diaphragm* | *2* | *2* |
| *Chest (lung, mediastinum, trachea)* | *3* | *3* |
| *Breast* | *3* | *4* |
| *Bladder* | *3* | *4* |
| *Perinephric fat* | *1* | *1* |
| *Multiple* | *8* | *11* |
| *Total* | | |

***Table S4B - BriTROC-1 lymph node locations by sample***

| **Lymph node summary* | *Number* |
|---|---|
| *PELVIC* | *14* |
| *RETROPERITONEAL* | *23* |
| *AXILLARY* | *14* |
| *CERVICAL* | *1* |
| *INGUINAL* | *12* |
| *MEDIASTINAL* | *1* |
| *RIGHT ILIAC FOSSA* | *1* |
| *SUPRA-CLAVICULAR* | *7* |
| *Total* | *71* |

***Table S5 - Systemic anti-cancer therapy prior to study entry***

***Table S6 - Response to first treatment following study entry as reported by recruiting site***

| ***Response (N)*** | ***Overall*** | ***Sensitive*** | ***Resistant*** |
|---|---|---|---|
| *Complete Response* | *36* | *35* | *1* |
| *Partial Response* | *78* | *67* | *11* |
| *Stable Disease* | *50* | *36* | *14* |
| *Progressive Disease* | *63* | *35* | *28* |
| *Not Evaluable* | *7* | *5* | *2* |
| *Not Known* | *32* | *27* | *5* |
| *No treatment* | *10* | *4* | *6* |
| *Total* | *276* | *209* | *67* |
| *Response rate (N)* | *114* | *102* | *12* |
| *Response Rate (CR+PR) (%)* | *50.2* | *59.0* | *22.2* |

***Table S7 - List of clinically relevant and/or frequently altered genes in HGSC***

| *Gene name* | *Chromosome* | *Gene start (bp)* | *Gene end (bp)* | *Cyto* | *expected* | *ensembl* |
|---|---|---|---|---|---|---|
| *AKT1* | *14* | *105235686* | *105262088* | *14q32.33* | *AMP* | *ENSG00000142208* |
| *AKT2* | *19* | *40736224* | *40791443* | *19q13.2* | *AMP* | *ENSG00000105221* |
| *AKT3* | *1* | *243651535* | *244014381* | *1q44* | *AMP* | *ENSG00000117020* |
| *CCND1* | *11* | *69455855* | *69469242* | *11q13.3* | *AMP* | *ENSG00000110092* |
| *CCND2* | *12* | *4382938* | *4414516* | *12p13.32* | *AMP* | *ENSG00000118971* |
| *CCND3* | *6* | *41902671* | *42018095* | *6p21.1* | *AMP* | *ENSG00000112576* |
| *CCNE1* | *19* | *30302805* | *30315215* | *19q12* | *AMP* | *ENSG00000105173* |
| *CDKN2A* | *9* | *21967751* | *21995300* | *9p21.3* | *DEL* | *ENSG00000147889* |
| *CDKN2B* | *9* | *22002902* | *22009362* | *9p21.3* | *DEL* | *ENSG00000147883* |
| *KRAS* | *12* | *25357723* | *25403870* | *12p12.1* | *AMP* | *ENSG00000133703* |
| *MECOM* | *3* | *168801287* | *169381406* | *3q26.2* | *AMP* | *ENSG00000085276* |
| *MYC* | *8* | *128747680* | *128753674* | *8q24.21* | *AMP* | *ENSG00000136997* |
| *NF1* | *17* | *29421945* | *29709134* | *17q11.2* | *DEL* | *ENSG00000196712* |
| *PIK3CA* | *3* | *178865902* | *178957881* | *3q26.32* | *AMP* | *ENSG00000121879* |
| *PTEN* | *10* | *89622870* | *89731687* | *10q23.31* | *DEL* | *ENSG00000171862* |
| *RB1* | *13* | *48877887* | *49056122* | *13q14.2* | *DEL* | *ENSG00000139687* |

*Table S8 - Significantly altered copy number signature by tissue site*

| signature | anova pval | posthoc group | posthoc pval | q.val |
|---|---|---|---|---|
| s1 | 0.003523 | pelvic-lymph_node | 0.009558 | 0.019117 |
| s1 | 0.003523 | pelvic-abdominal | 0.039269 | 0.078538 |

***Table S9 - Ploidy change scoring***

| ***patient*** | ***samples*** | ***category 1*** | ***category 2*** | ***category 3*** | ***star rating*** |
|---|---|---|---|---|---|
| *BRITROC-209* | *IM_295, JBLAB-4960* | *TRUE* | *FALSE* | *TRUE* | ** |
| *BRITROC-216* | *IM_336, IM_337, IM_338, IM_339, IM_340, IM_341, IM_342, JBLAB-4965* | *TRUE* | *FALSE* | *FALSE* | * |
| *BRITROC-23* | *IM_56, JBLAB-4128, JBLAB-4967* | *TRUE* | *FALSE* | *TRUE* | ** |
| *BRITROC-241* | *IM_423,JBLAB-4996* | *TRUE* | *TRUE* | *FALSE* | ** |
| *BRITROC-248* | *IM_403, JBLAB-19302, JBLAB-19303* | *TRUE* | *FALSE* | *TRUE* | ** |
| *BRITROC-267* | *IM_383,JBLAB-19330* | *TRUE* | *TRUE* | *FALSE* | ** |
| *BRITROC-274* | *IM_395, IM_396, IM_397, JBLAB-19337, JBLAB-19338* | *TRUE* | *FALSE* | *FALSE* | * |
| *BRITROC-67* | *IM_115, JBLAB-4179* | *TRUE* | *TRUE* | *FALSE* | ** |
| *BRITROC-74* | *IM_124, JBLAB-4186, JBLAB-4187, JBLAB-4188, JBLAB-4189* | *TRUE* | *TRUE* | *TRUE* | *** |

*NB - Details of the ploidy change rating and designation can be found in the supplemental methods*

***Table S10 - Short Variant Table***

A table of all short variants detected in the analysis
