## Supplementary material for "The genomic landscape of recurrent ovarian high grade serous carcinoma: the BriTROC-1 study": Figure S29

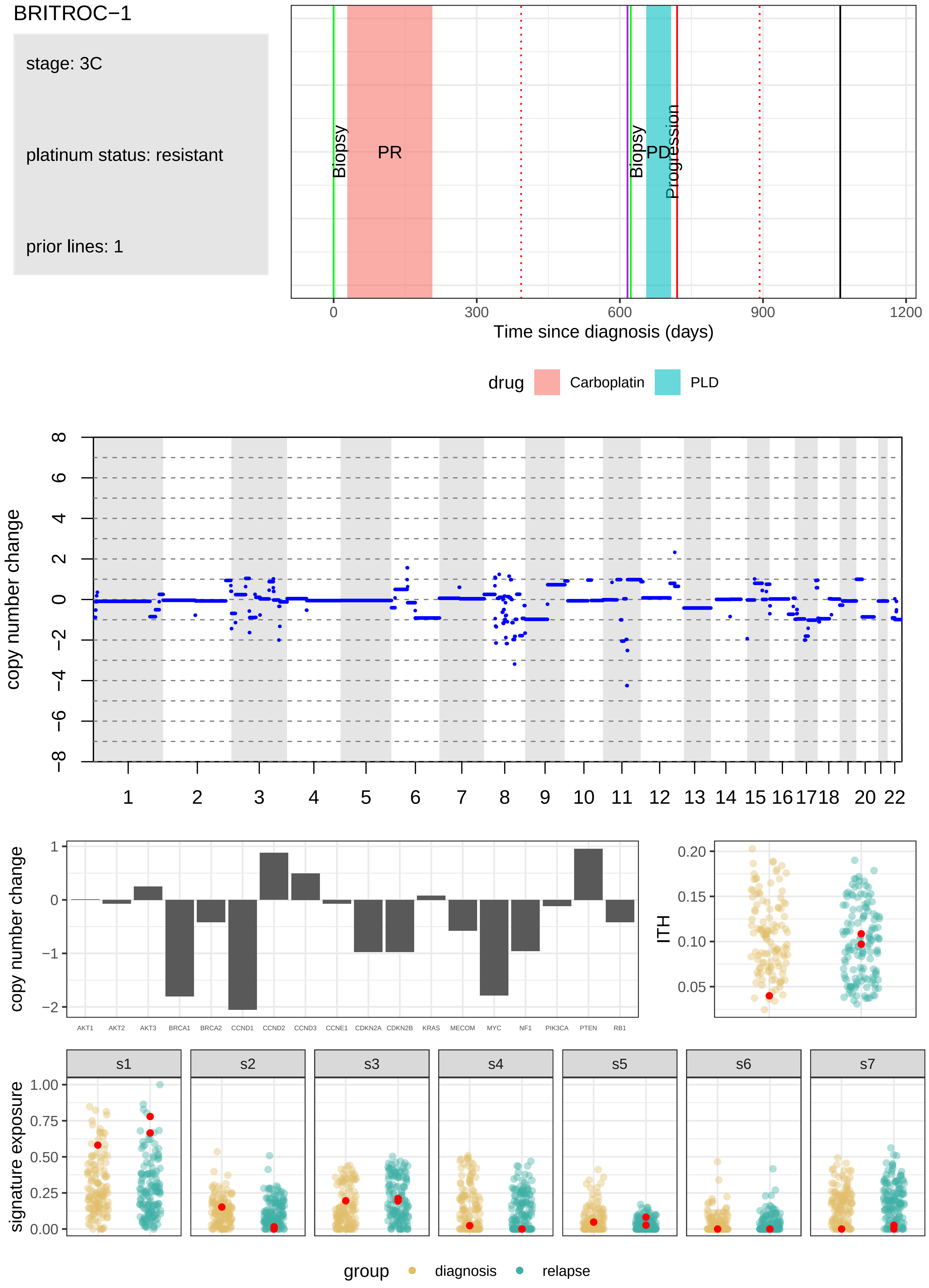
BRITROC−1
stage: 3C
platinum status: resistant
prior lines: 1
Biopsy
PR
Biopsy
PD
Progression
0
300
600
900
1200
Time since diagnosis (days)
drug
Carboplatin
PLD
copy number change
8
6
4
2
0
−2
−4
−6
−8
20
22
copy number change
1
0
−1
−2
AKT1
AKT2
AKT3
BRCA1
BRCA2
CCND1
CCND2
CCND3
CCNE1
CDKN2A
CDKN2B
KRAS
MECOM
MYC
NF1
PIK3CA
PTEN
RB1
ITH
0.20
0.15
0.10
0.05
s1
s2
s3
s4
s5
s6
s7
signature exposure
1.00
0.75
0.50
0.25
0.00
group
diagnosis
relapse

BRITROC−106

stage: 3C

platinum status: sensitive

prior lines: 2

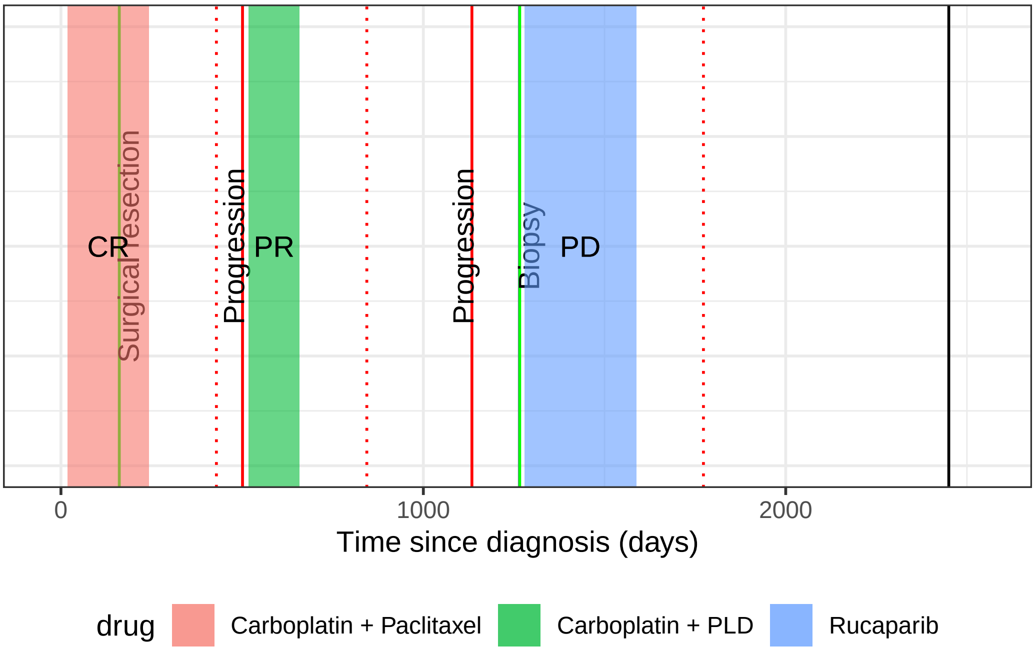

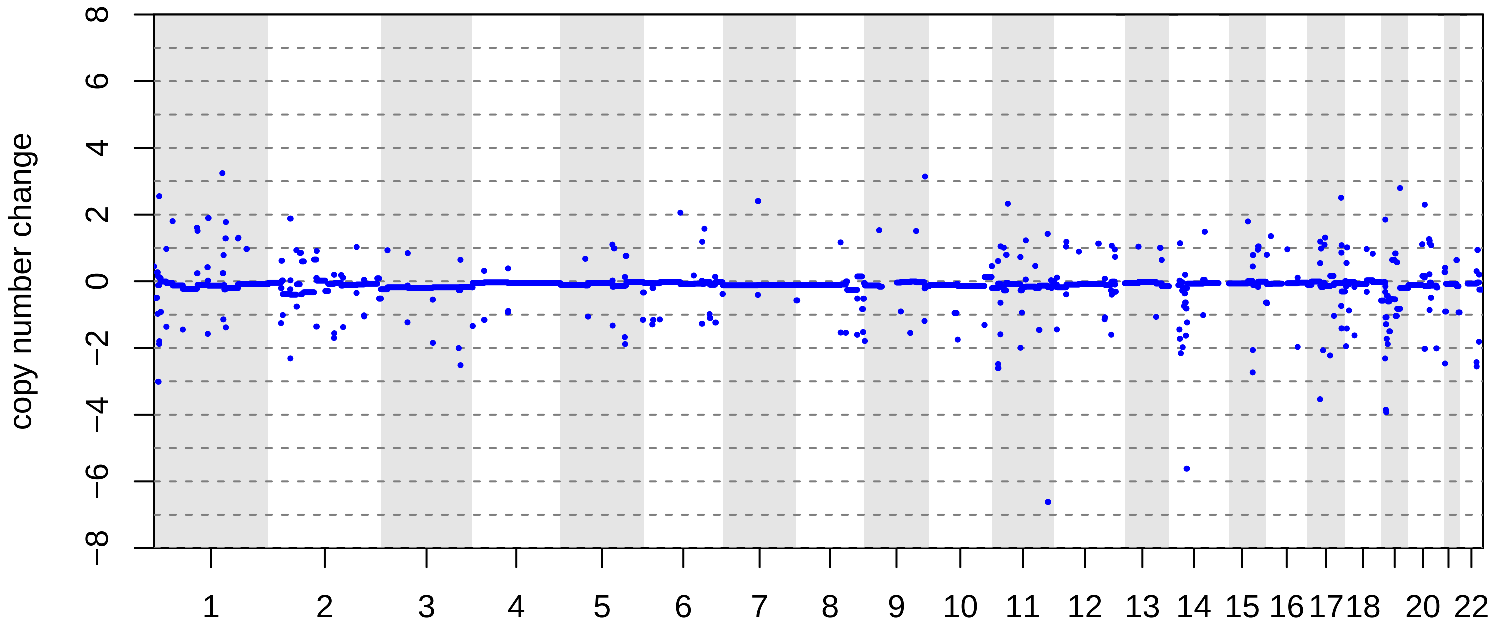

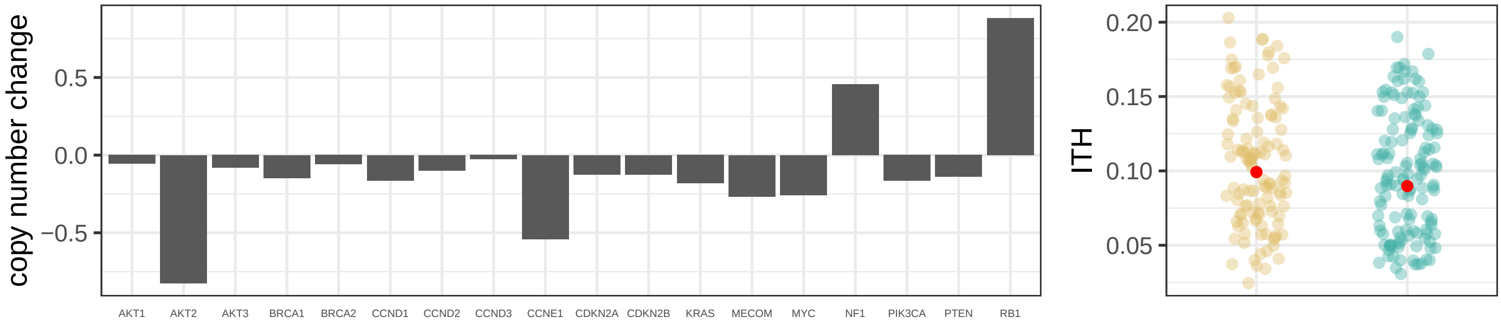

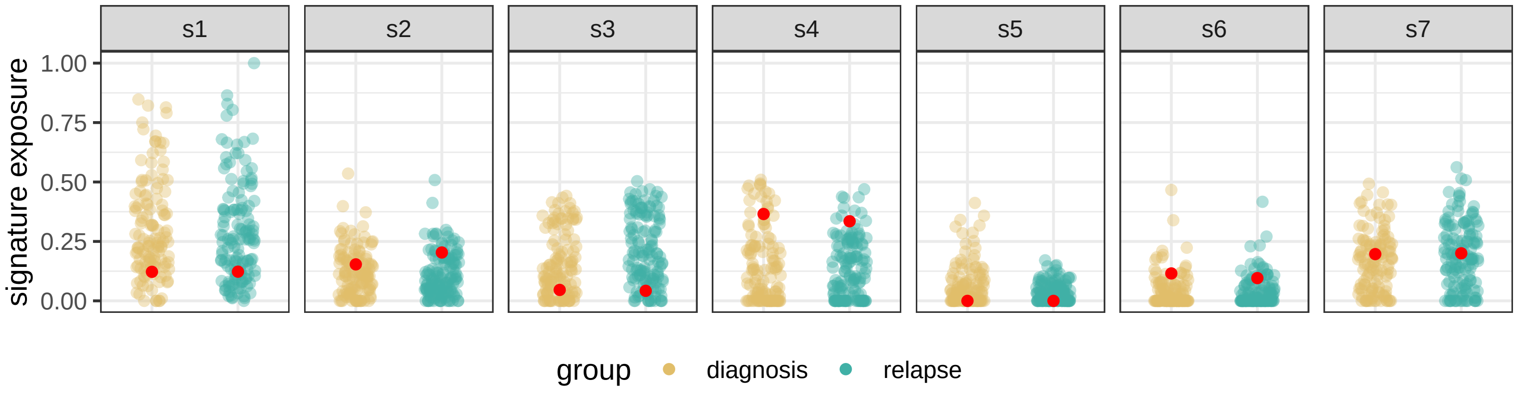

BRITROC−11

stage: 3C

platinum status: sensitive

prior lines: 1

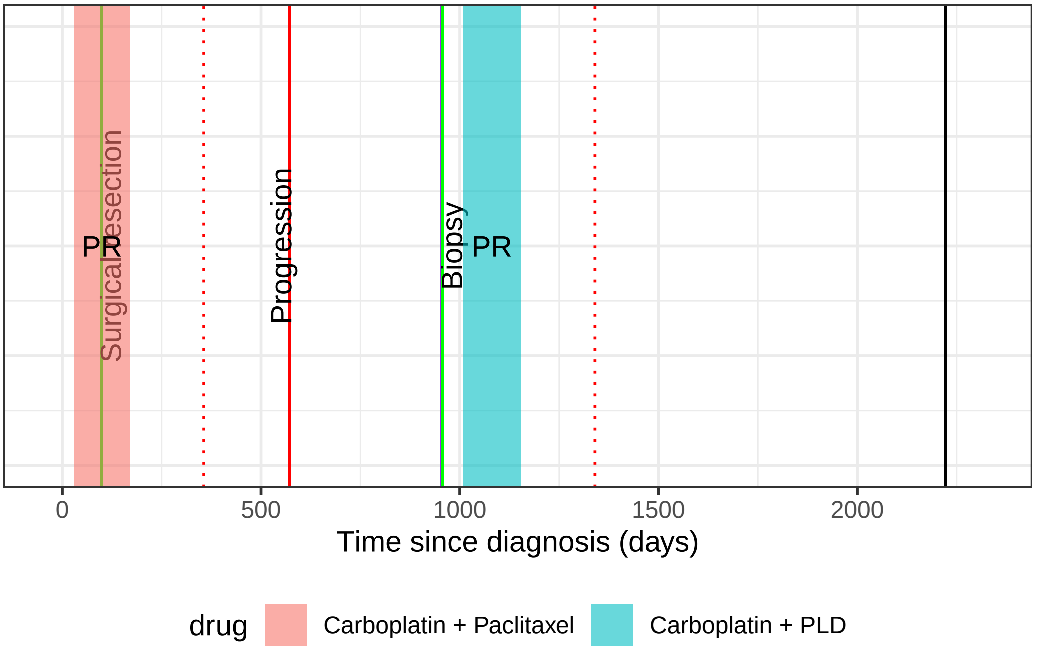

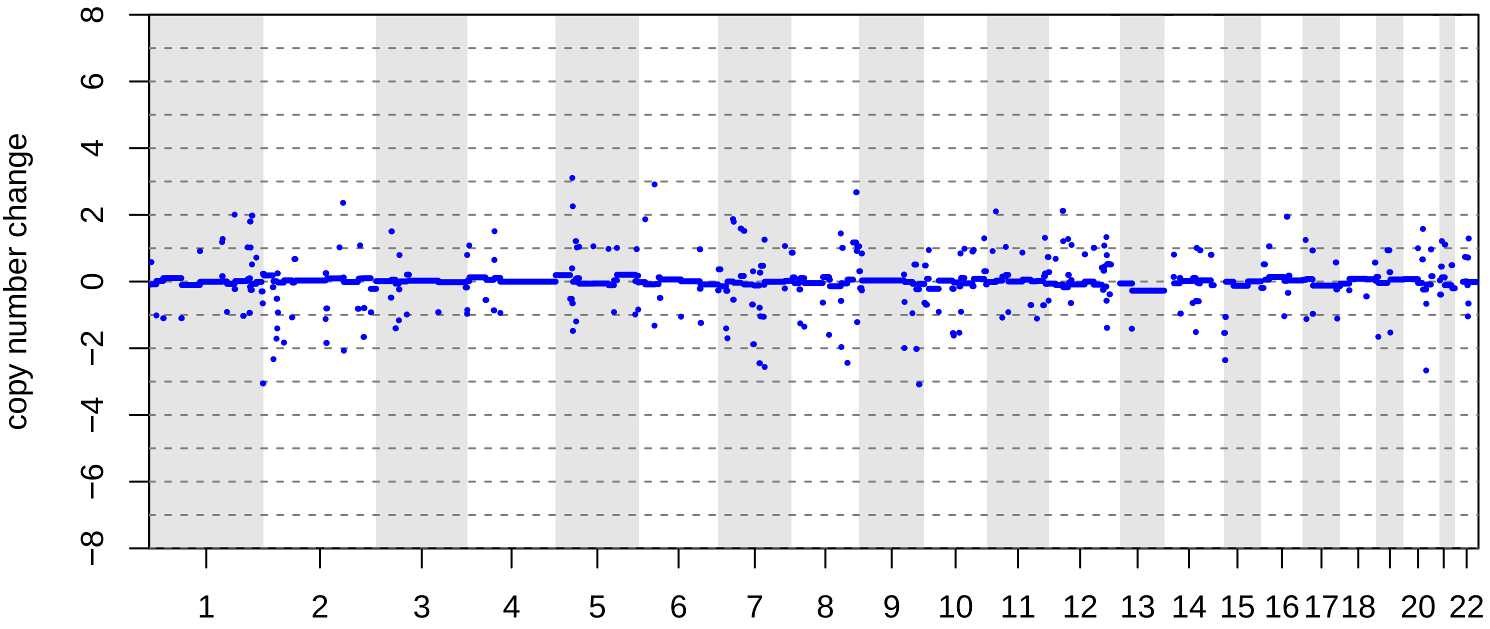

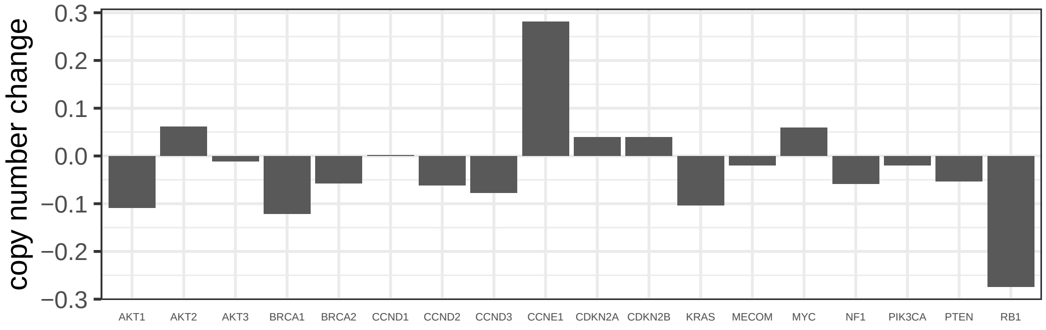

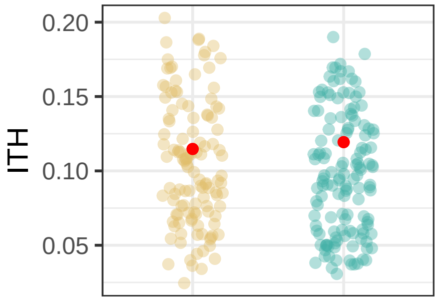

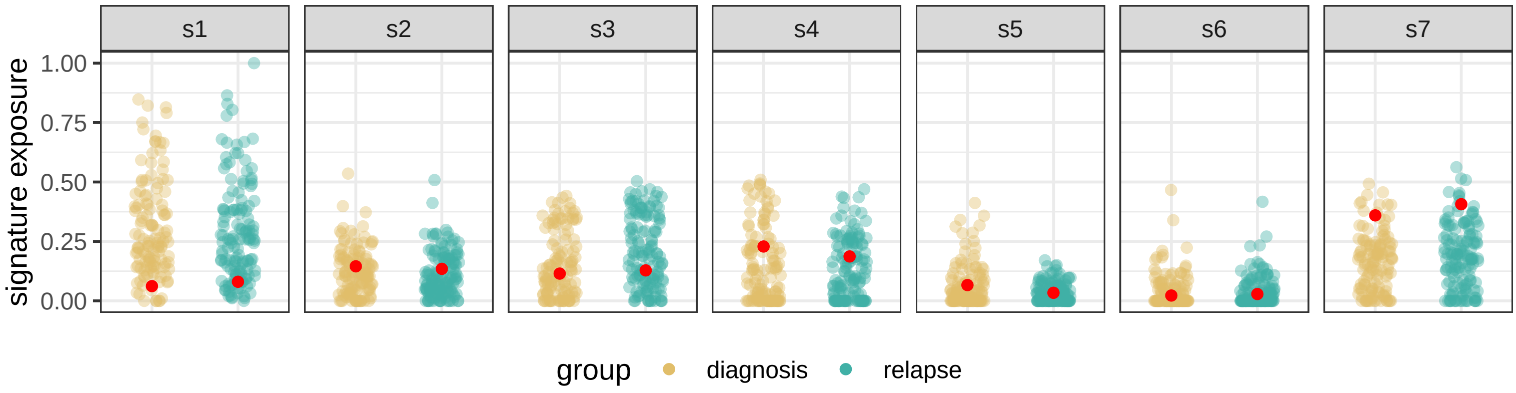

BRITROC−124

stage: 3C

platinum status: sensitive

prior lines: 1

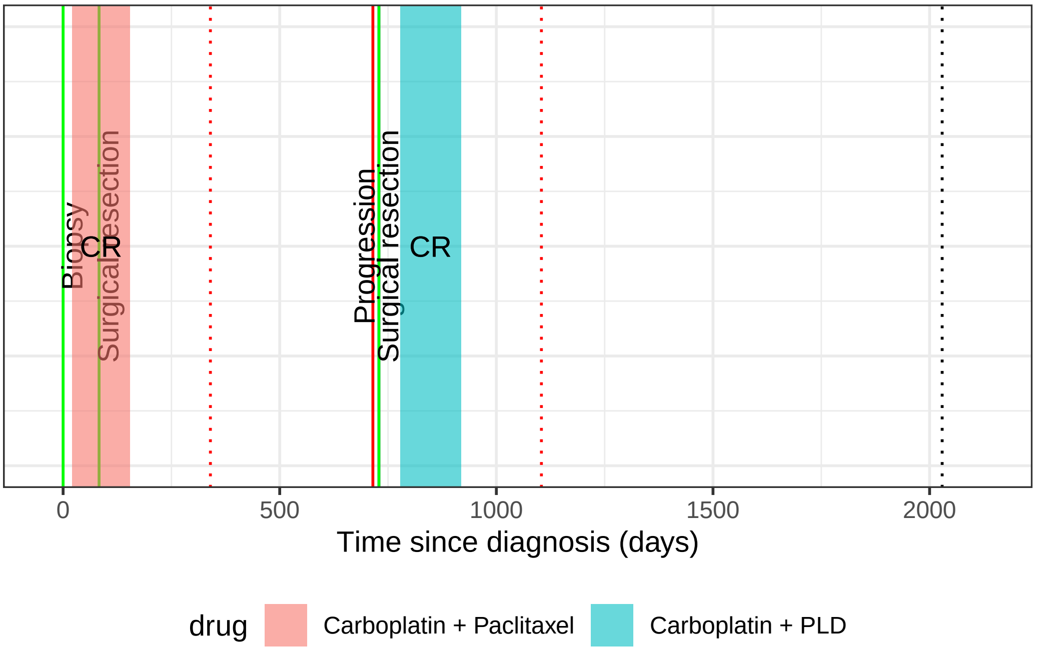

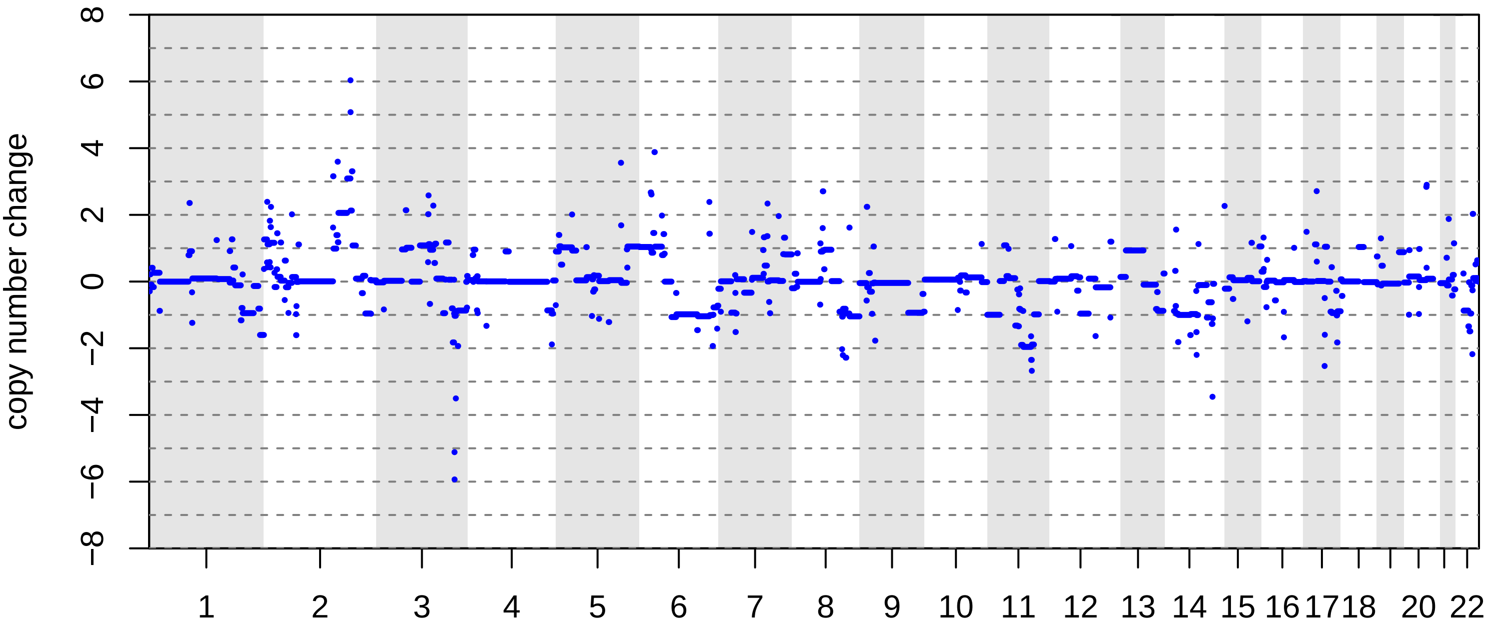

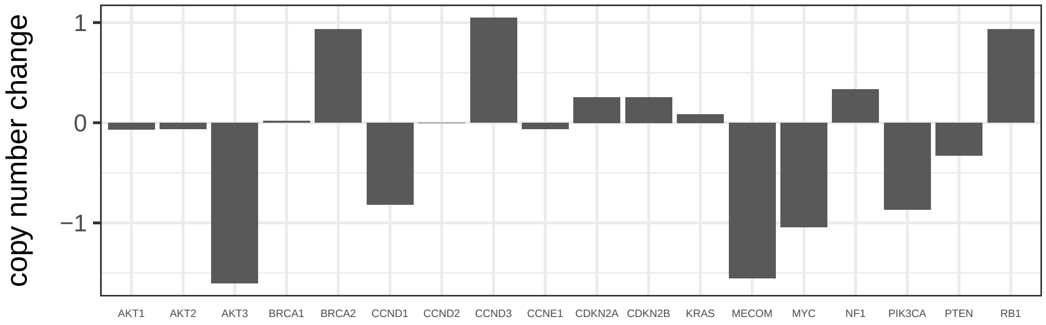

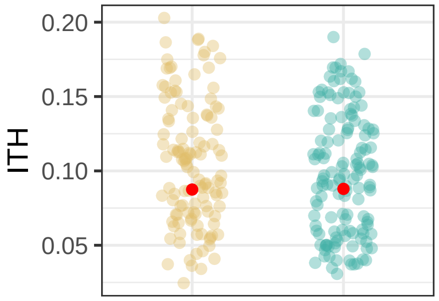

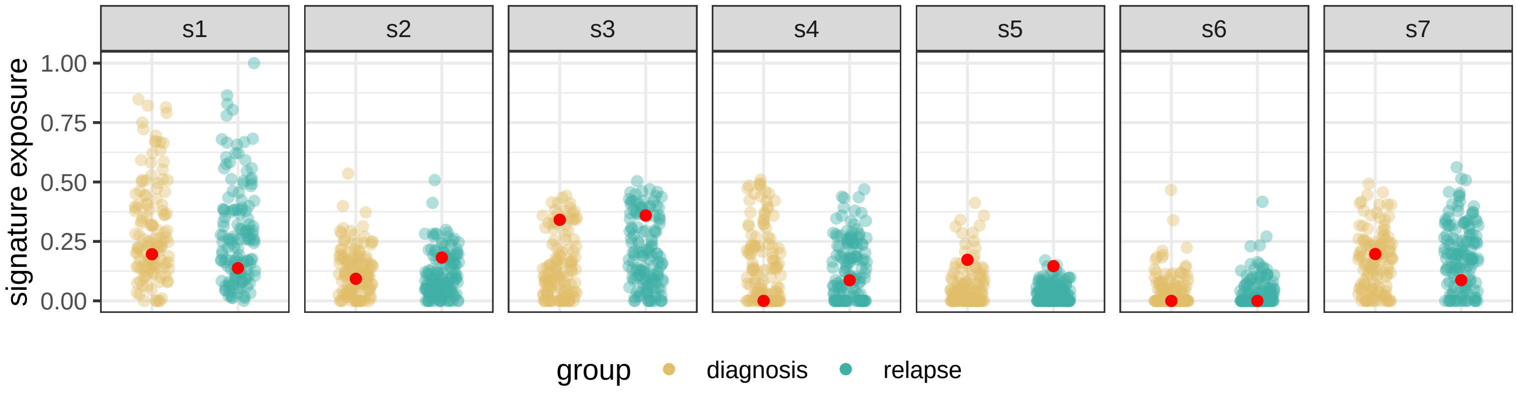

BRITROC−132

stage: 3B

platinum status: sensitive

prior lines: 1

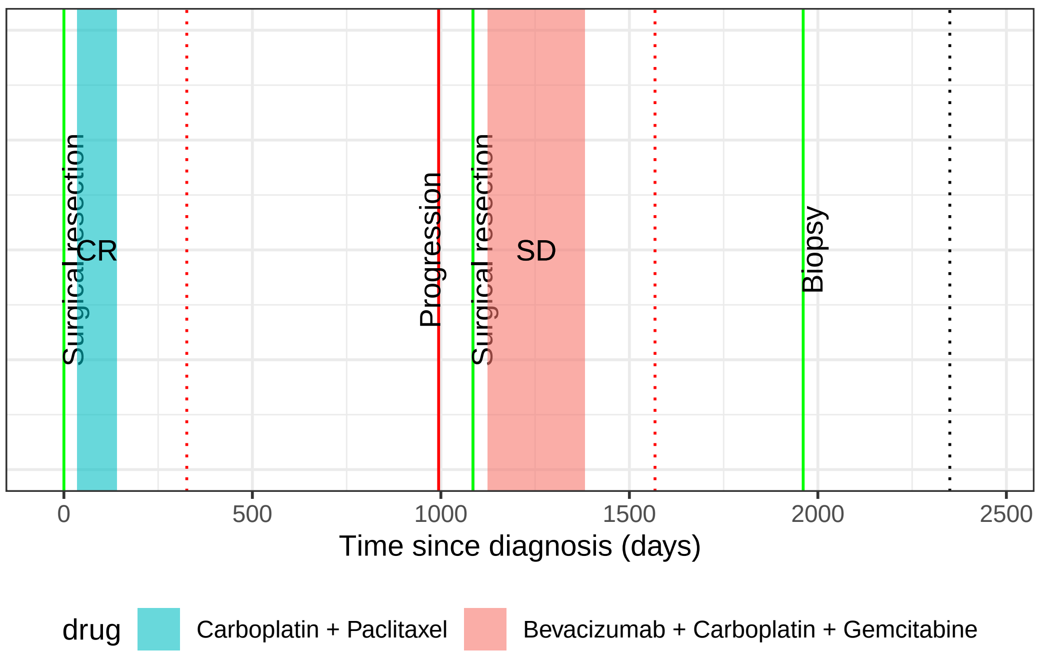

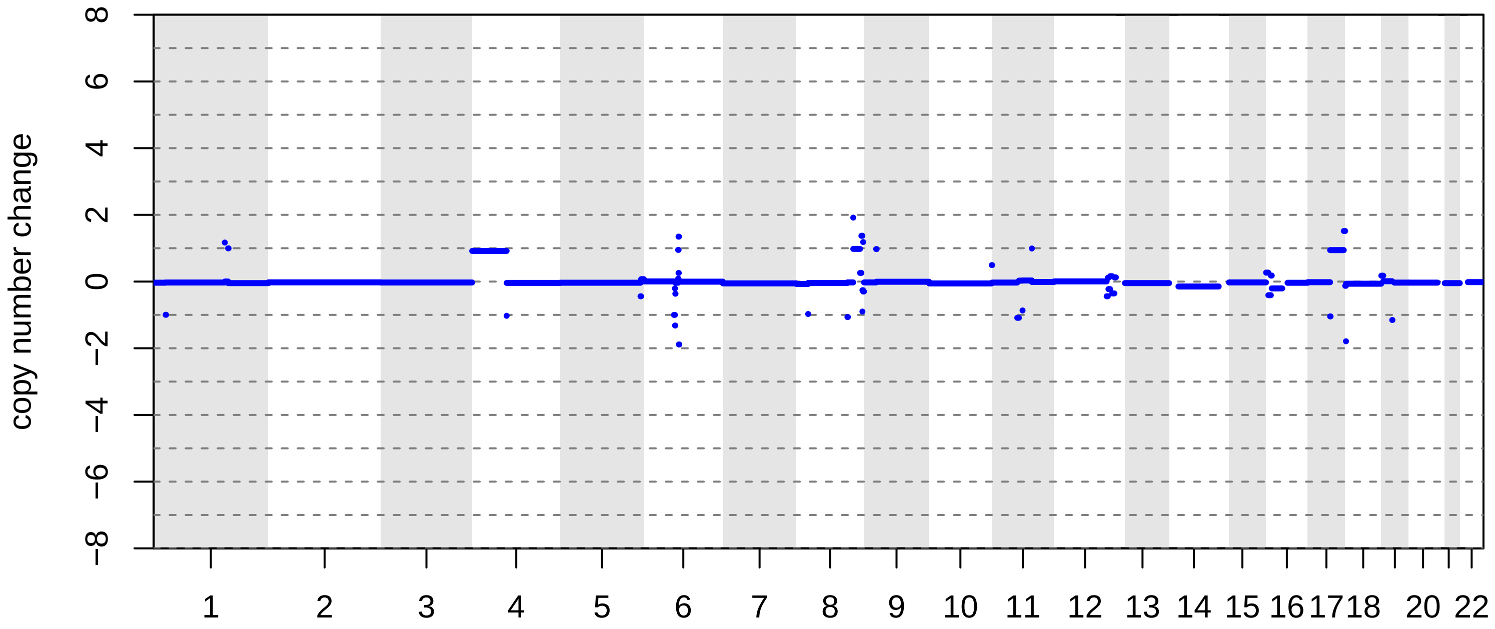

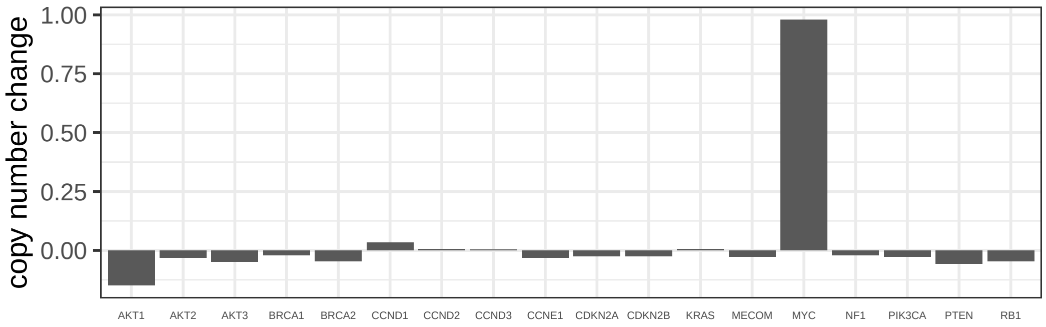

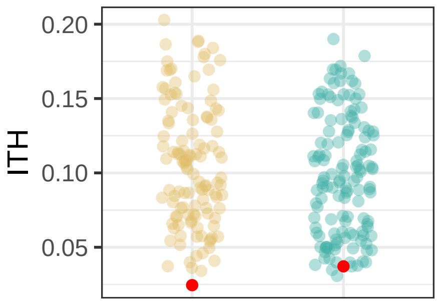

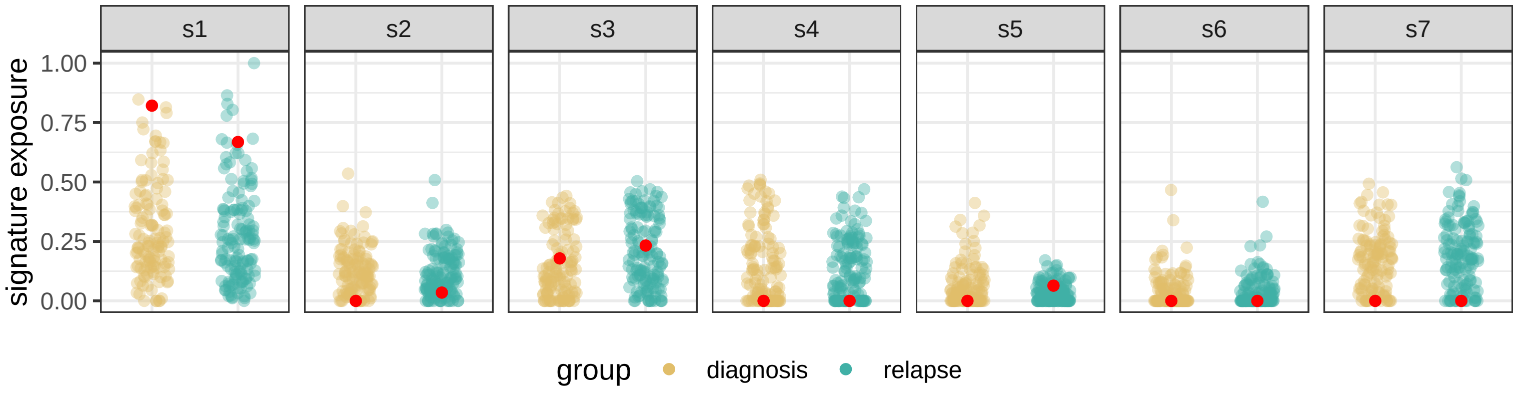

BRITROC−147

stage: 4NA

platinum status: resistant

prior lines: 1

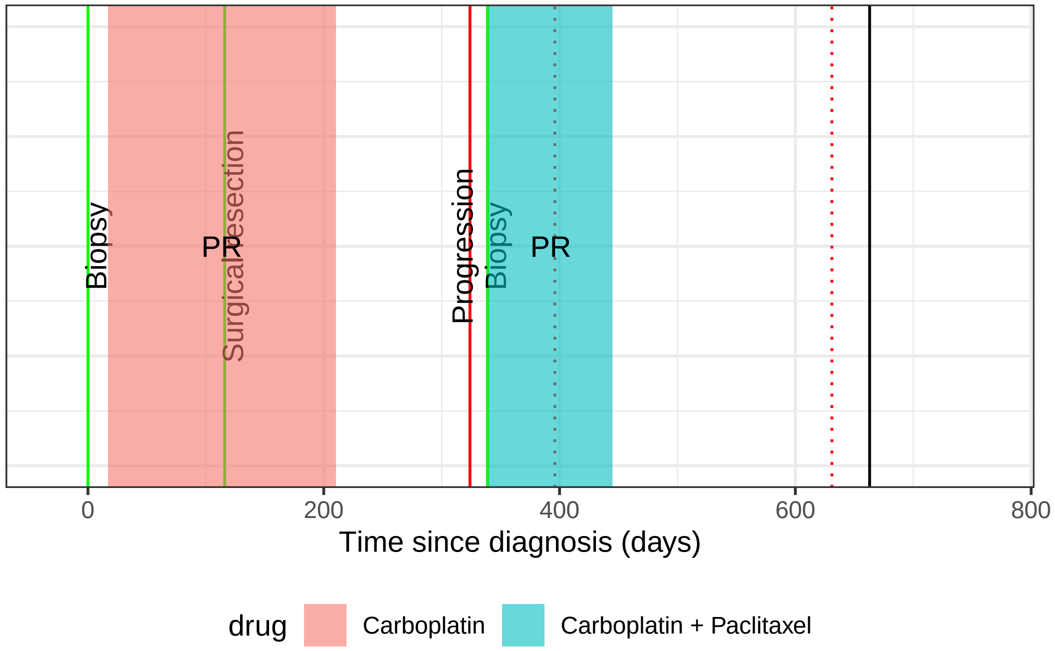

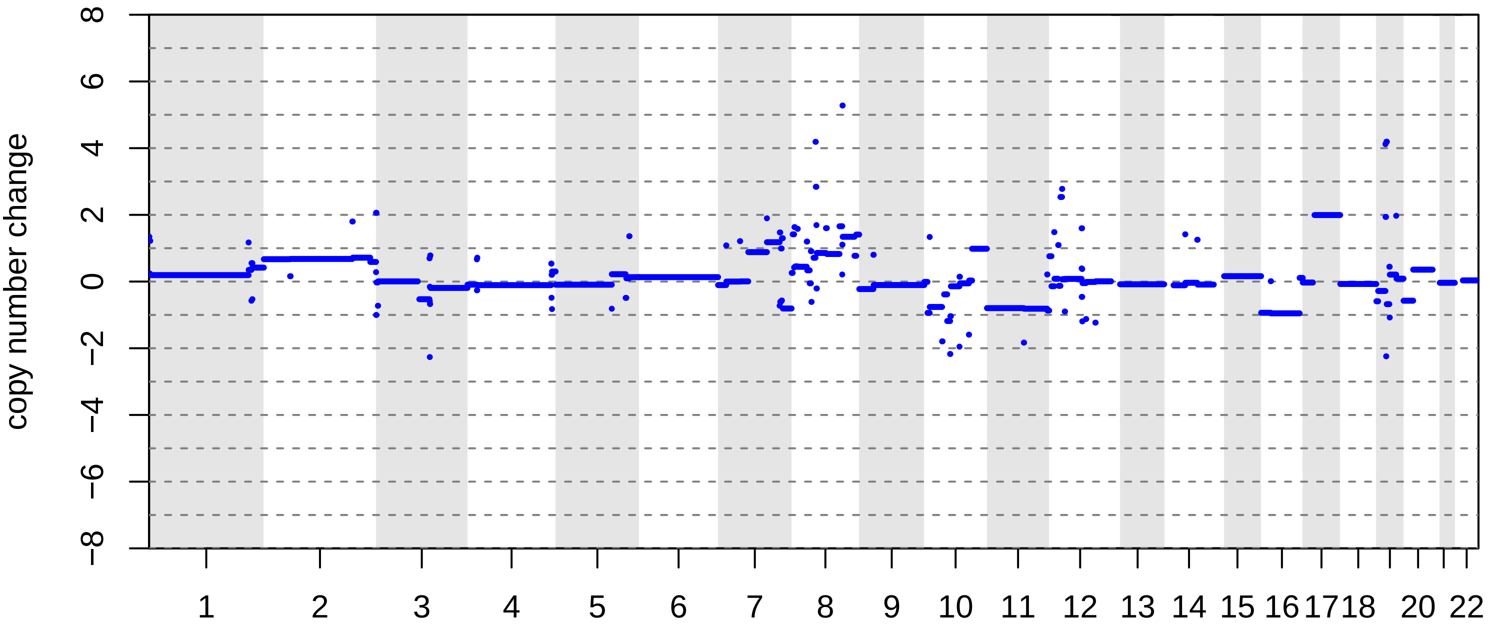

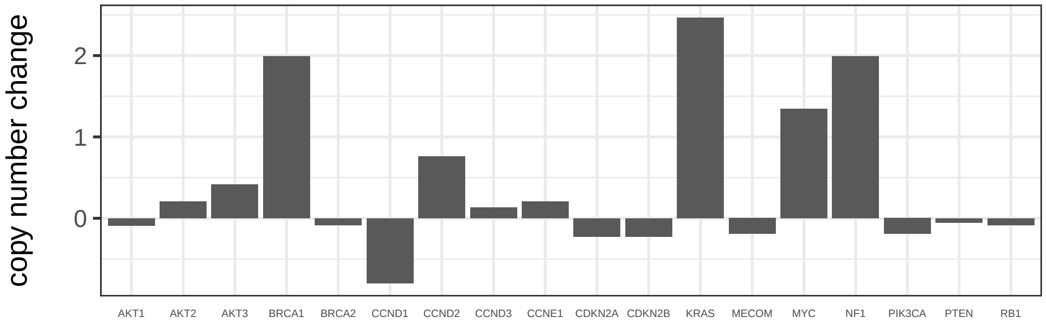

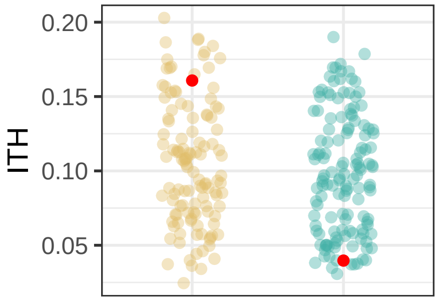

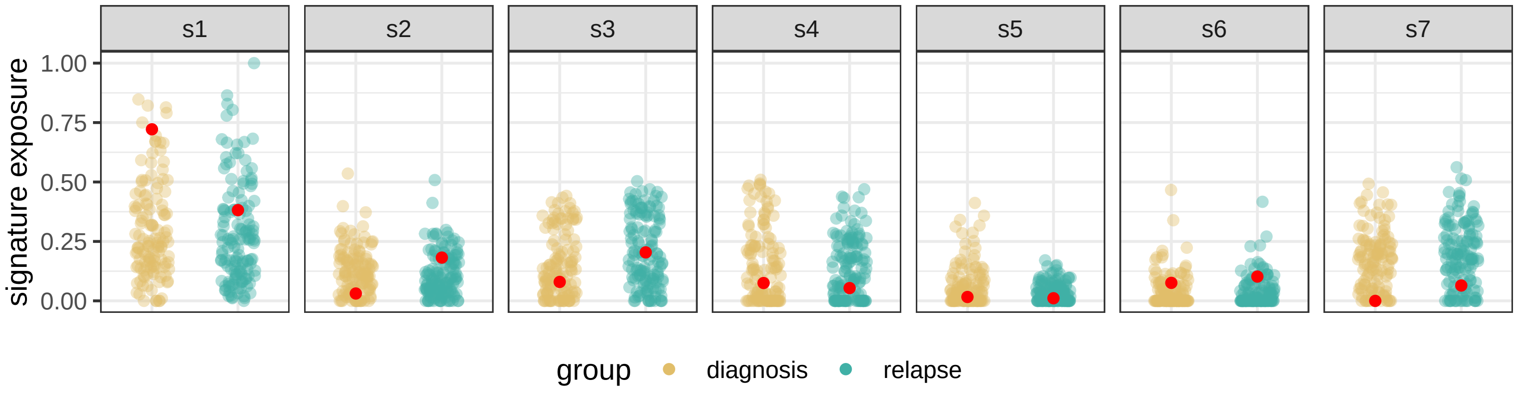

BRITROC−148

stage: 4NA

platinum status: resistant

prior lines: 2

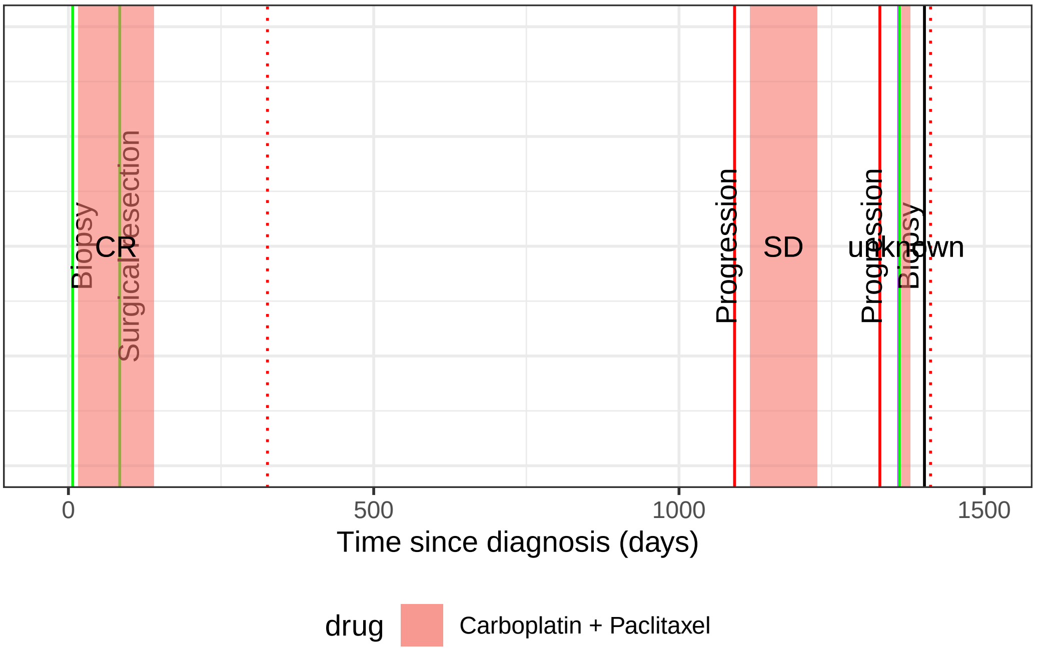

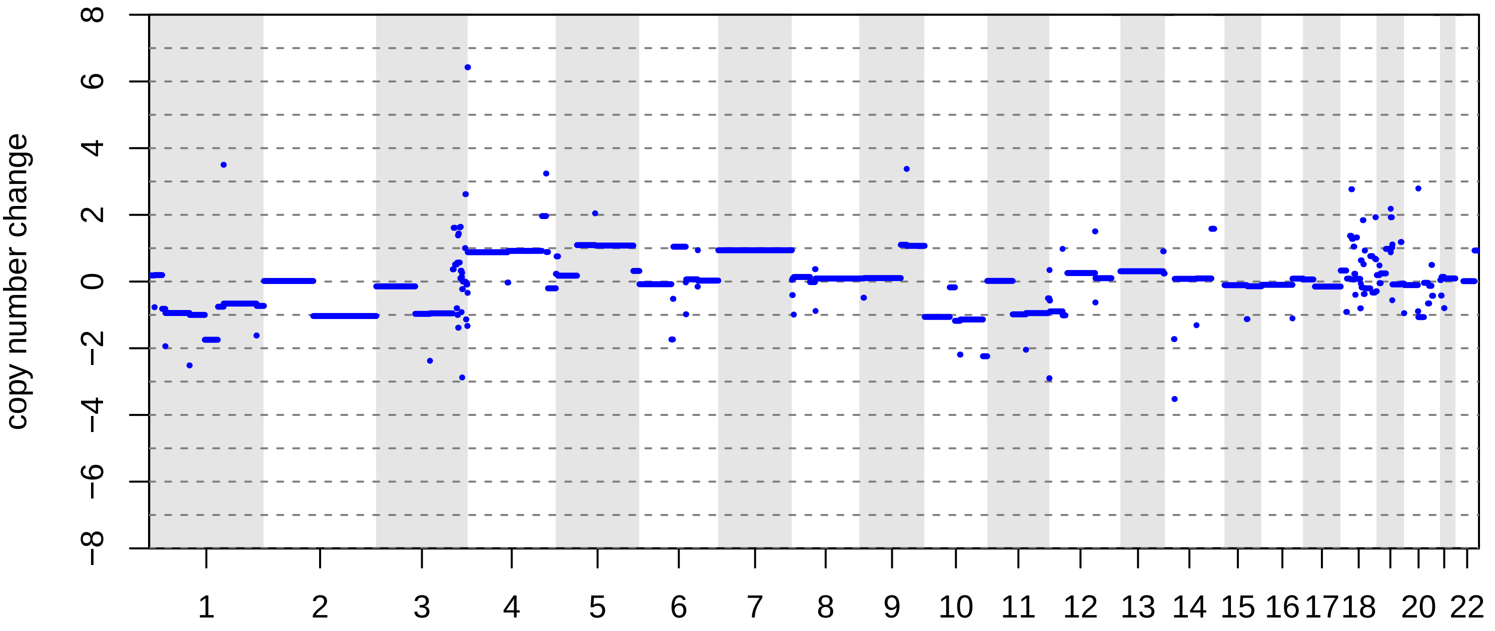

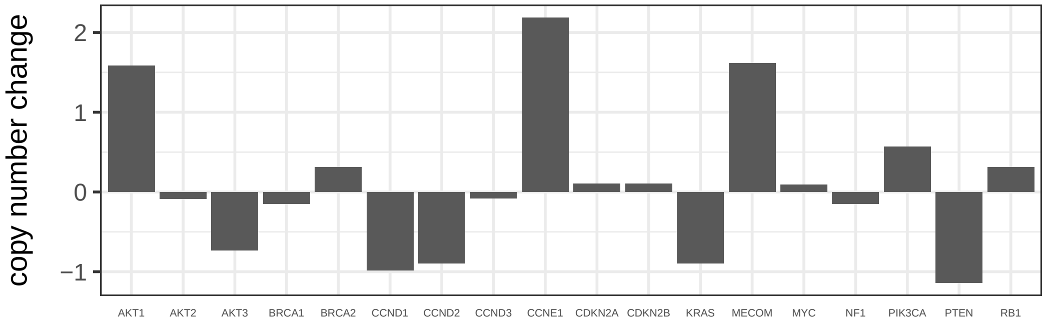

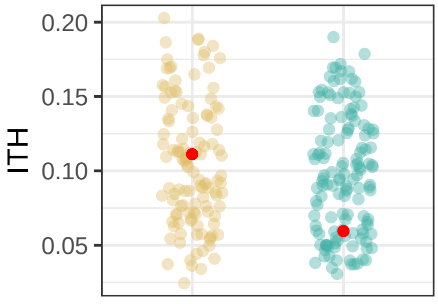

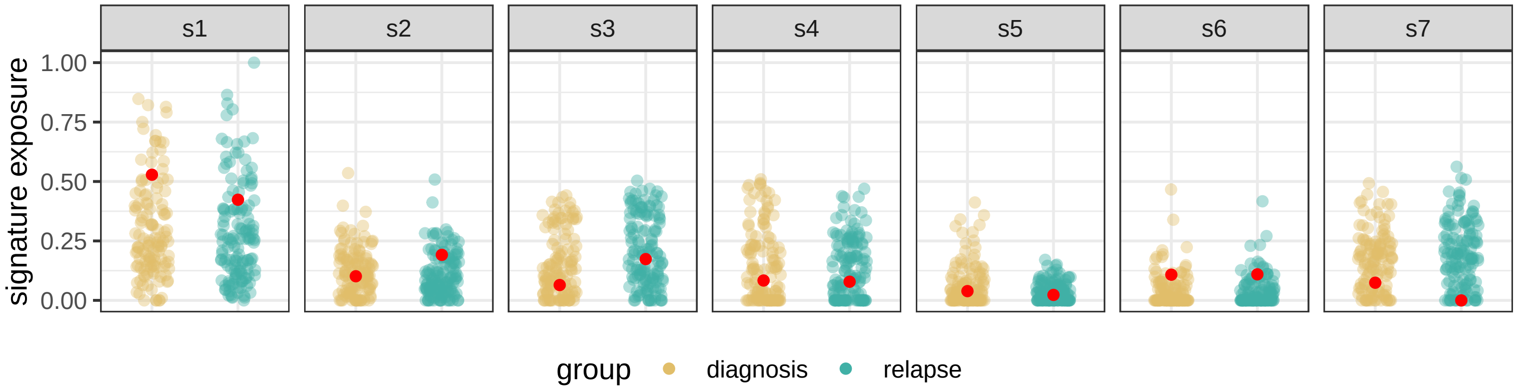

BRITROC−157

stage: 4NA

platinum status: sensitive

prior lines: 1

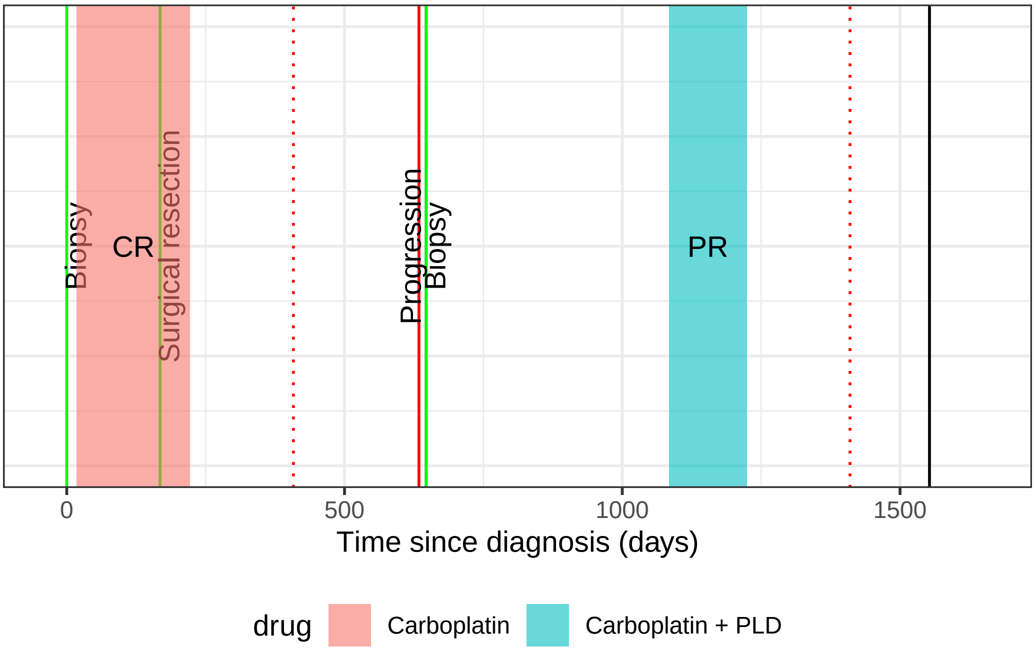

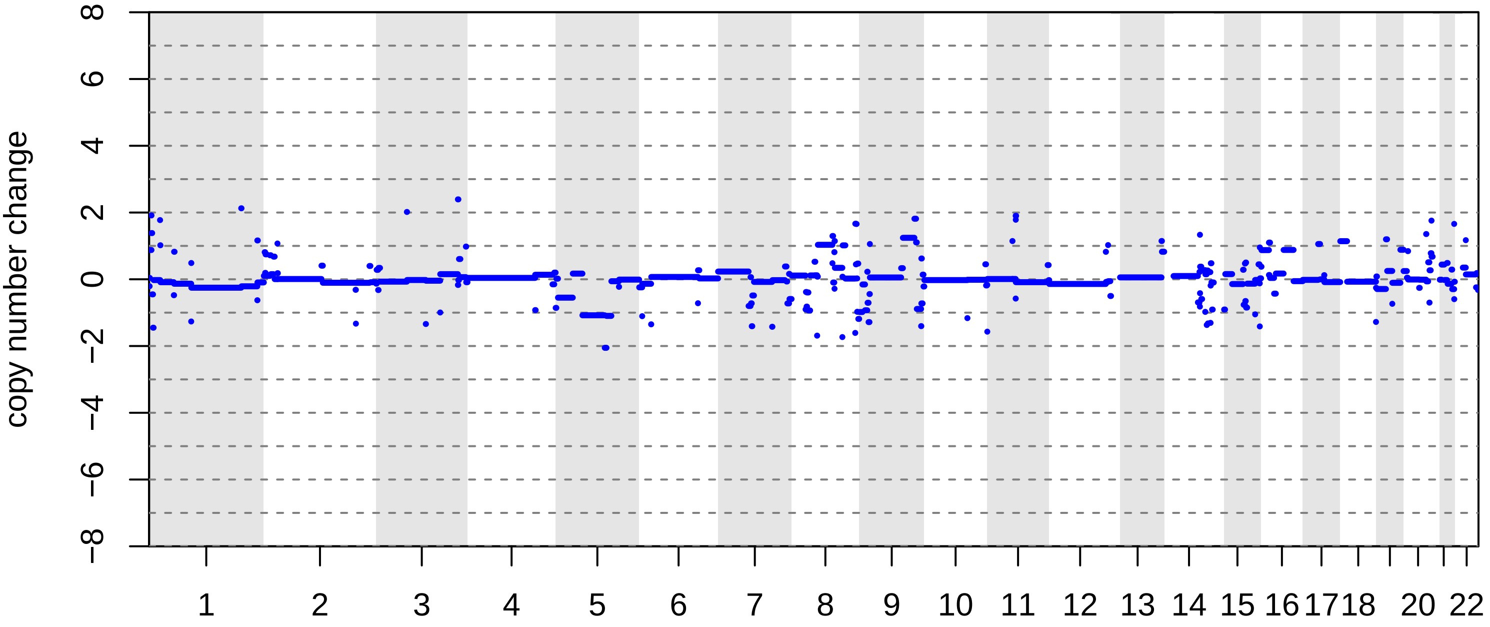

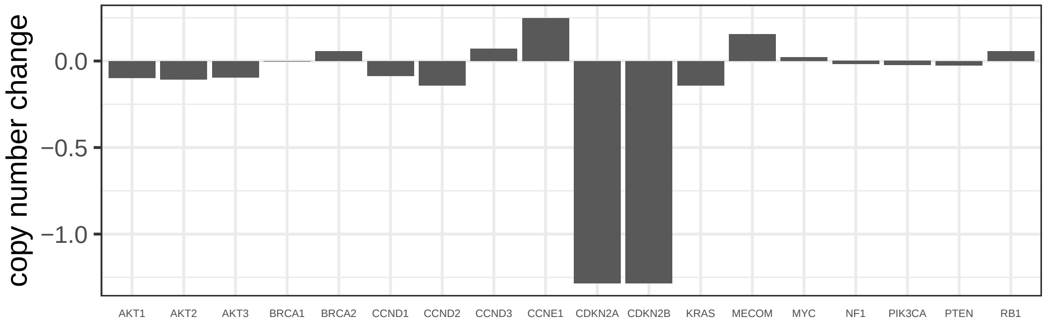

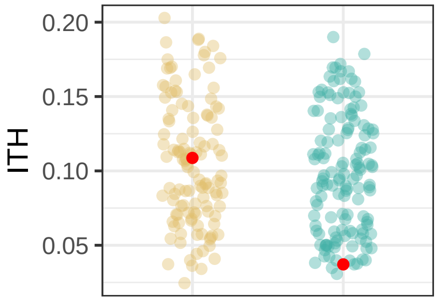

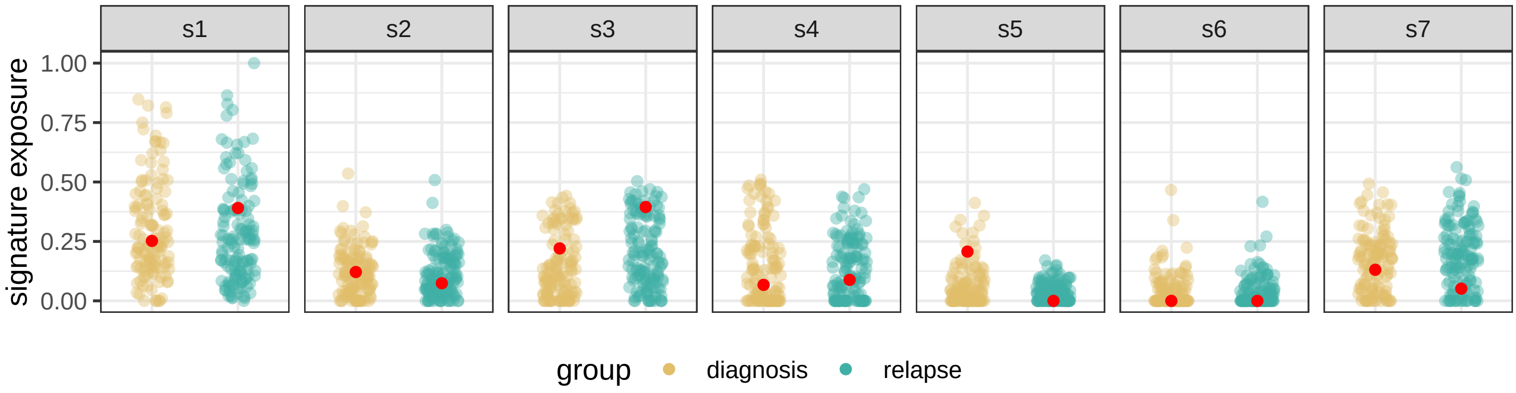

### BRITROC−187

stage: 1A

platinum status: sensitive

prior lines: 1

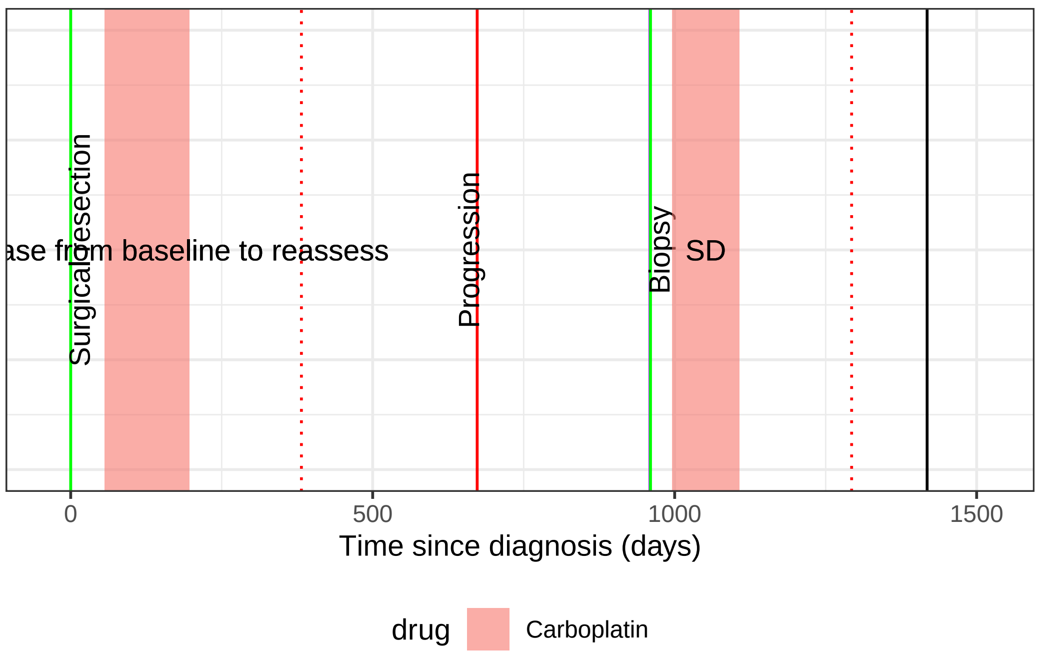

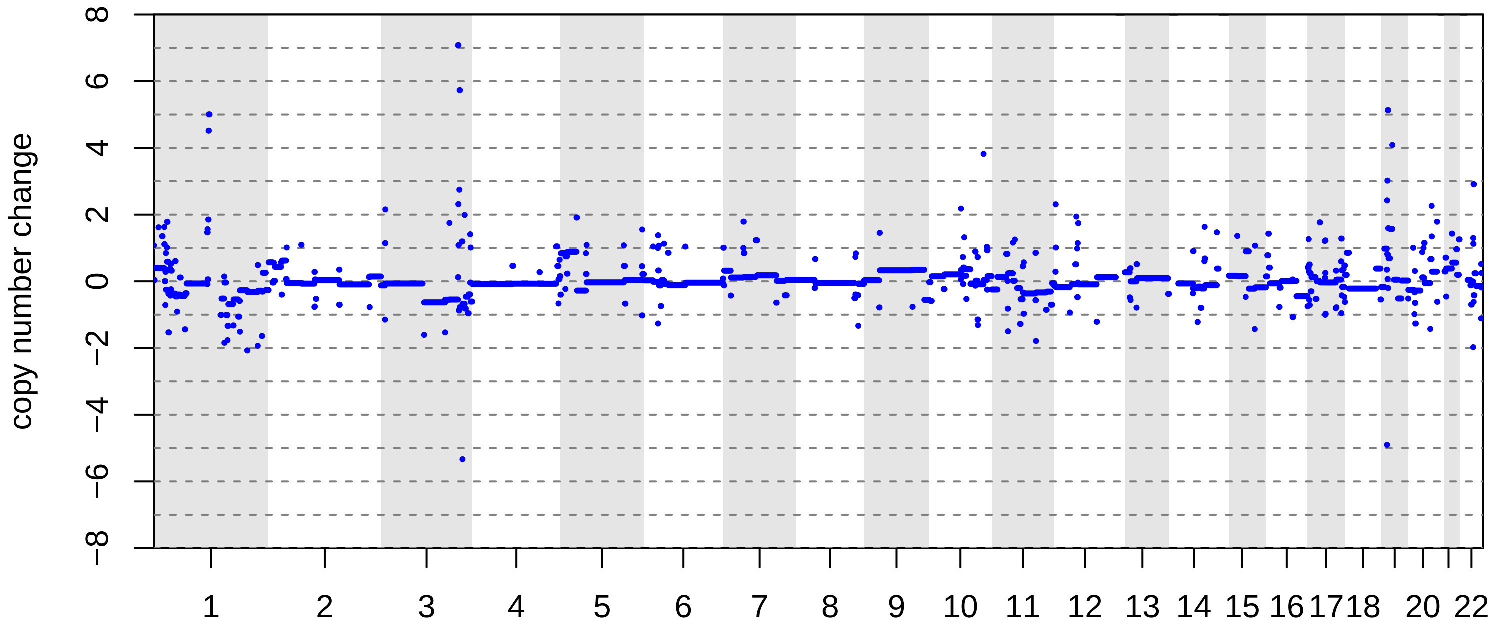

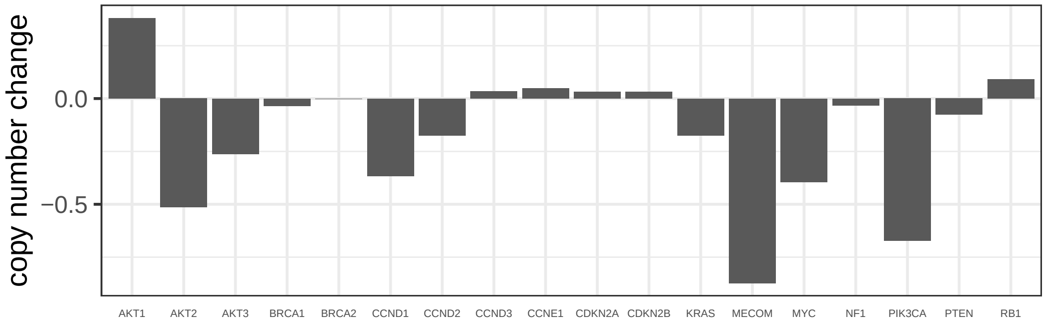

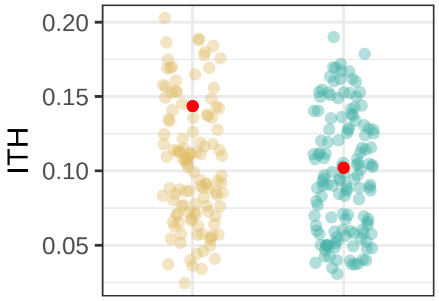

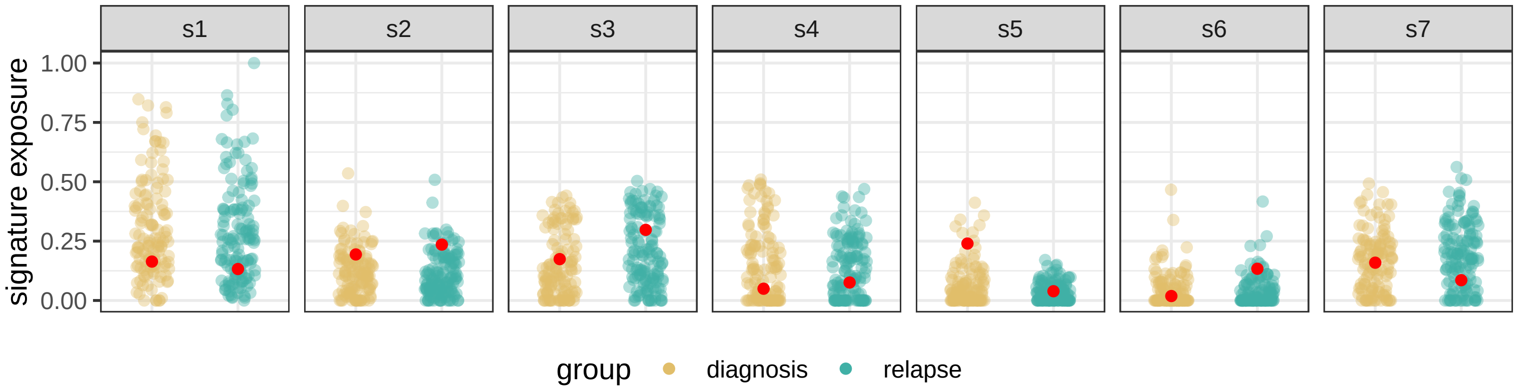

BRITROC−192

stage: 3C

platinum status: sensitive

prior lines: 1

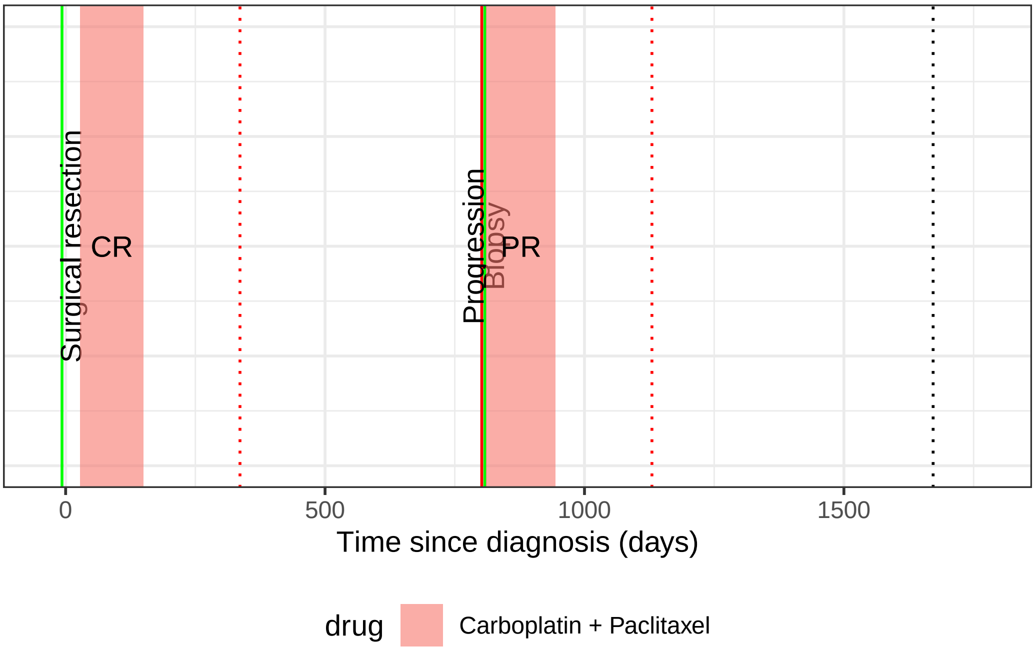

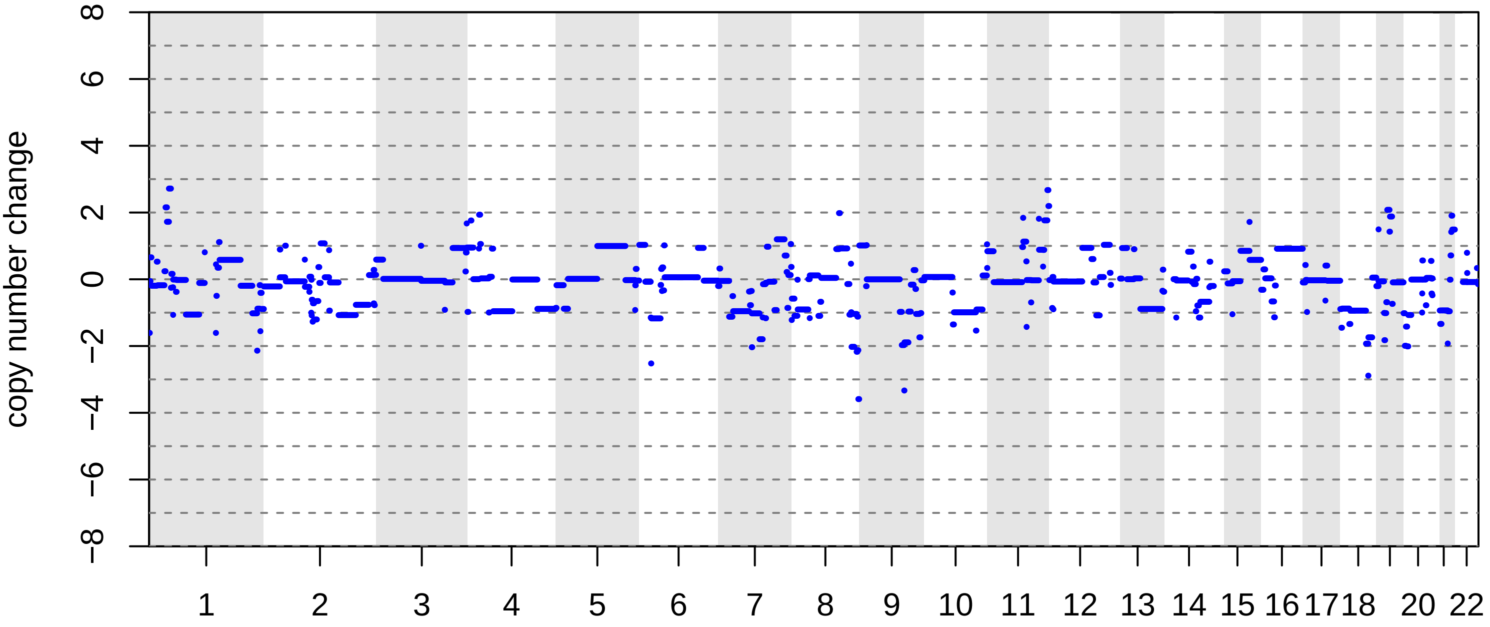

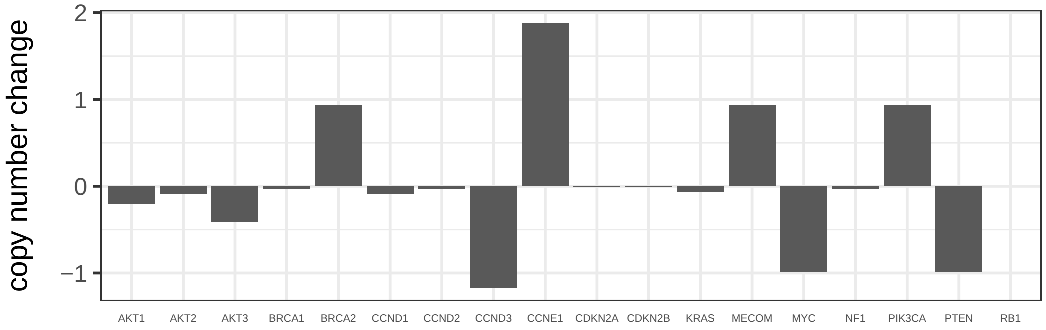

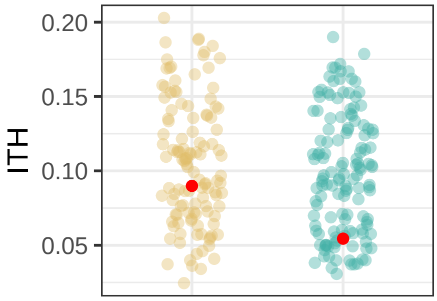

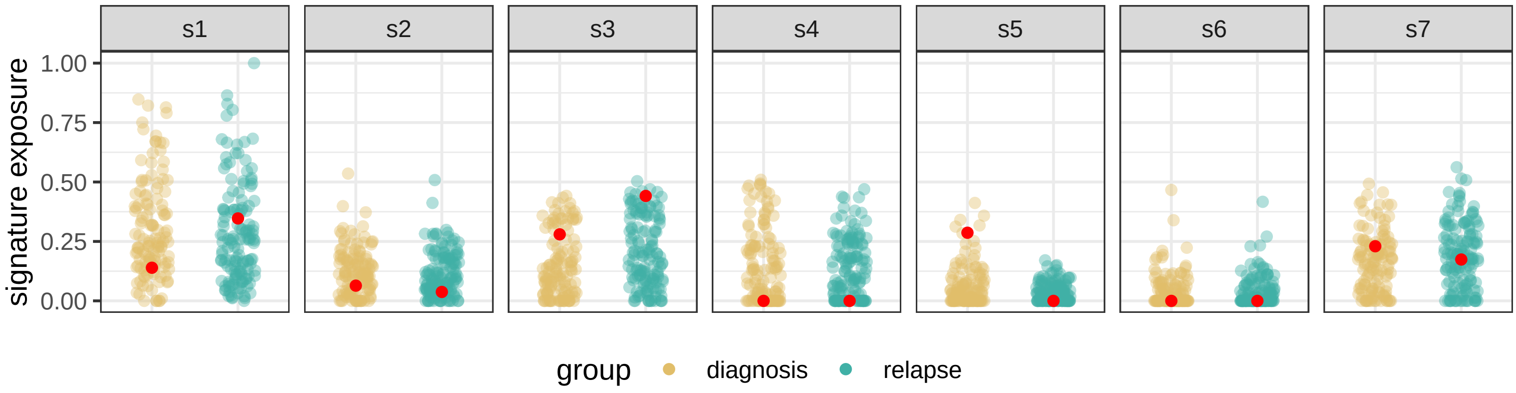

BRITROC−2

stage: 3C

platinum status: resistant

prior lines: 1

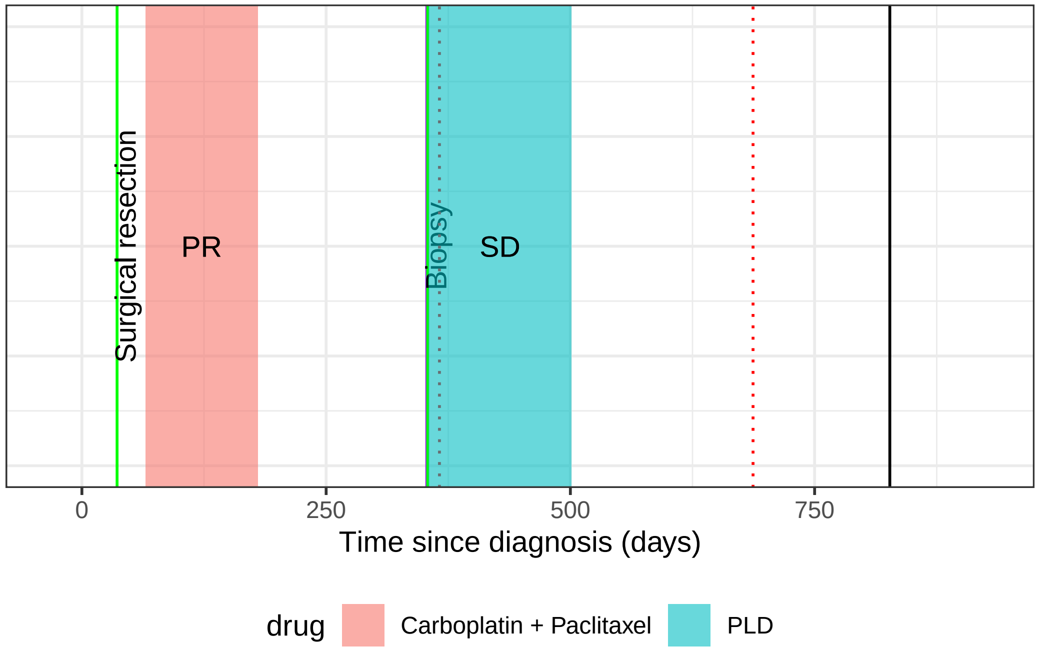

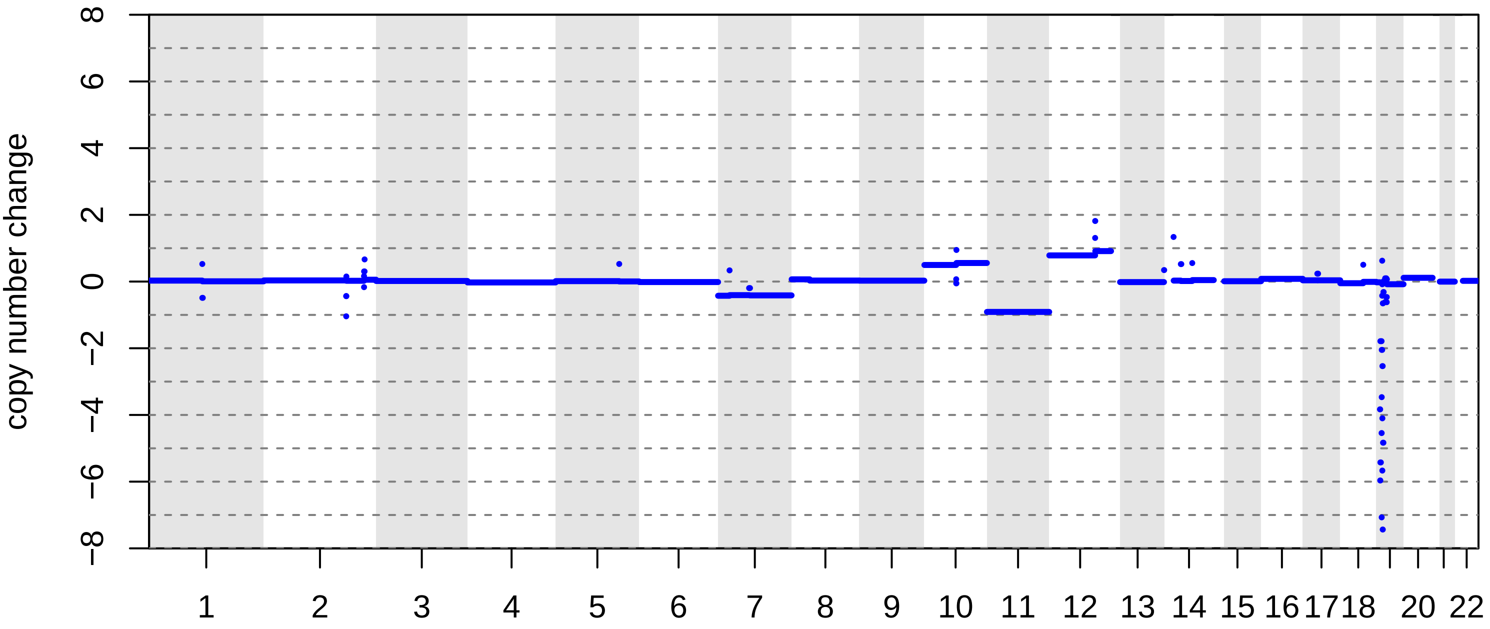

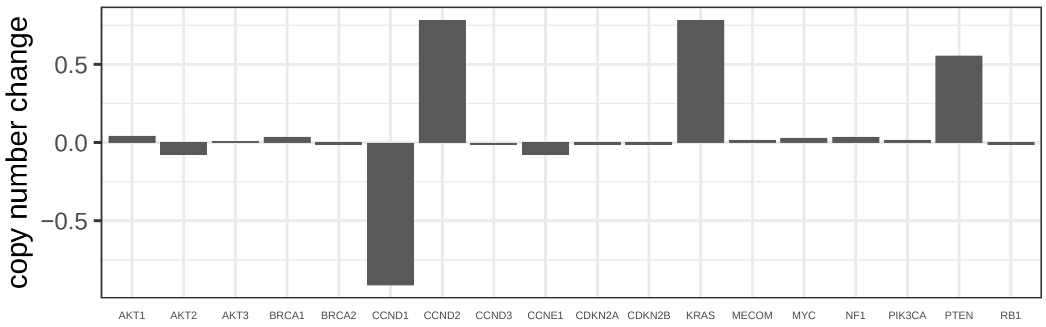

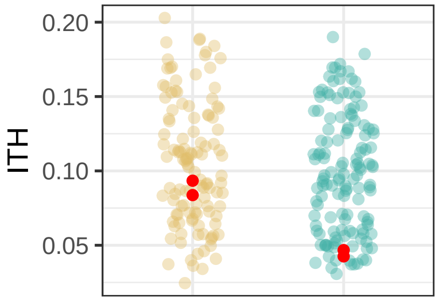

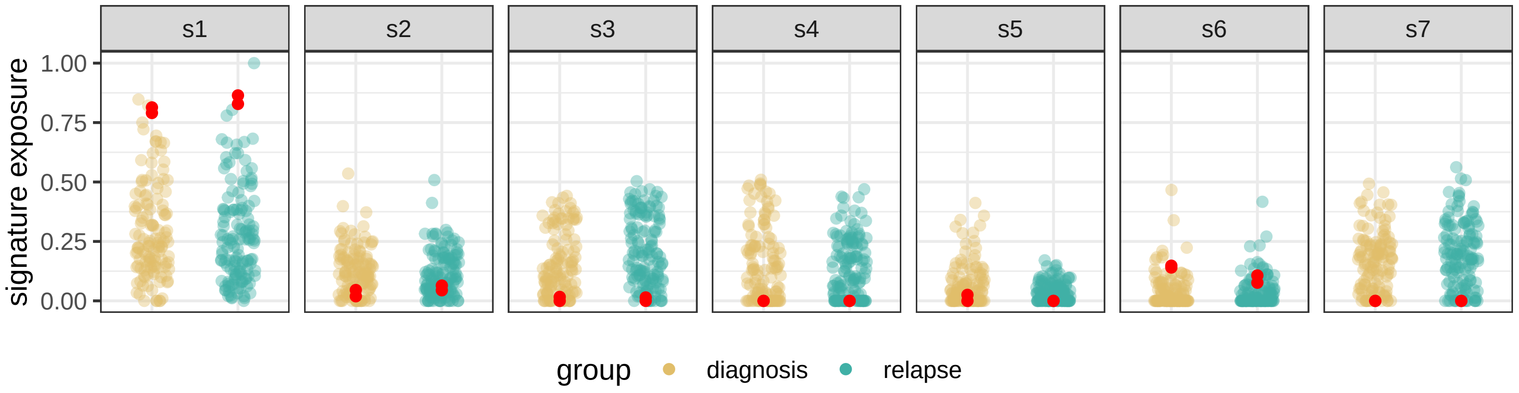

### BRITROC−204

stage: 2B

platinum status: sensitive

prior lines: 1

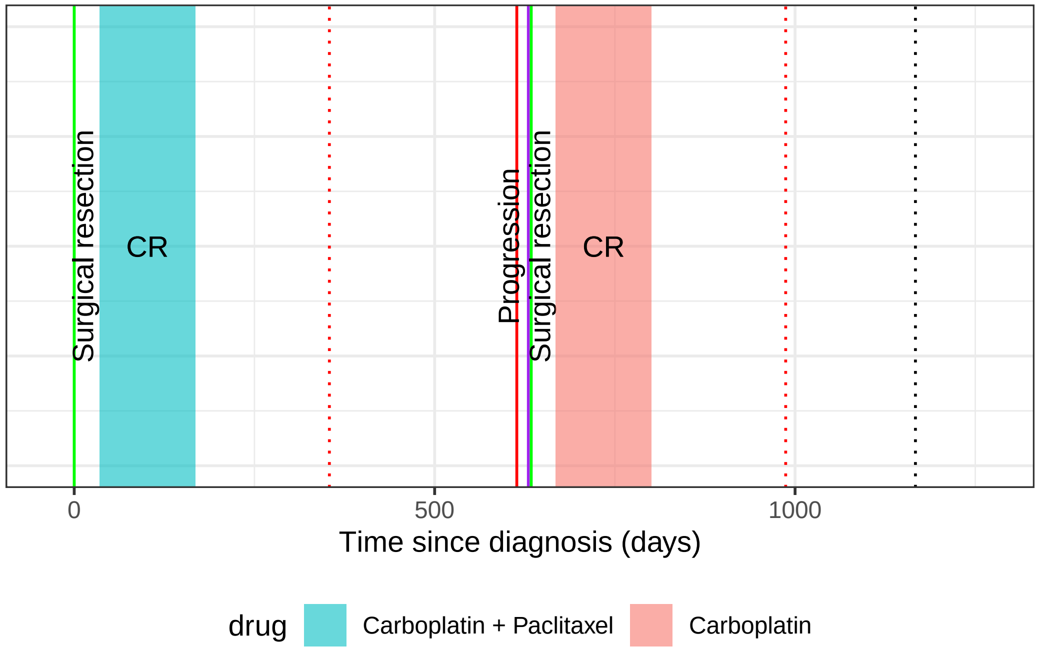

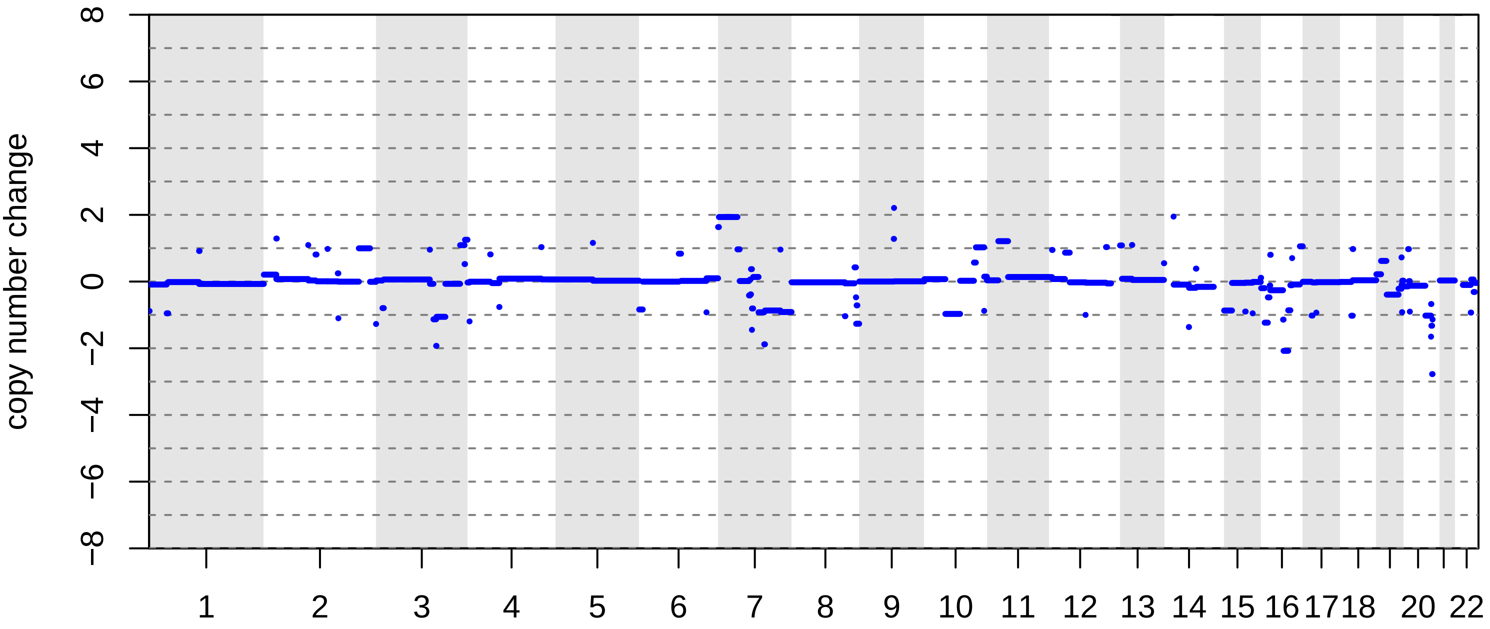

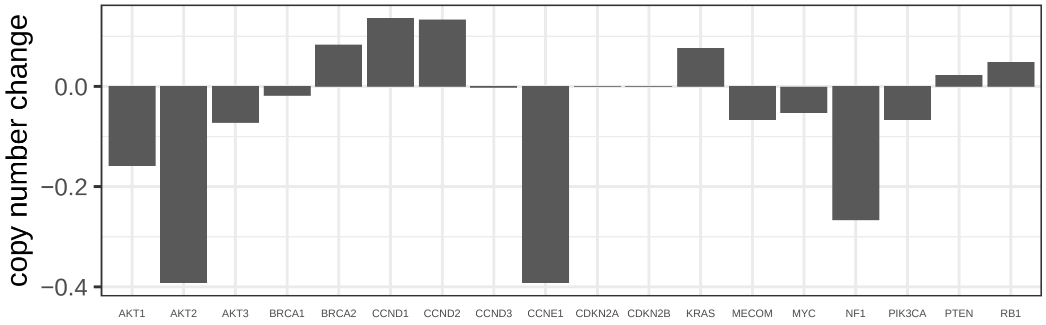

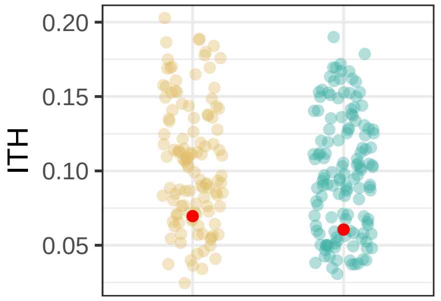

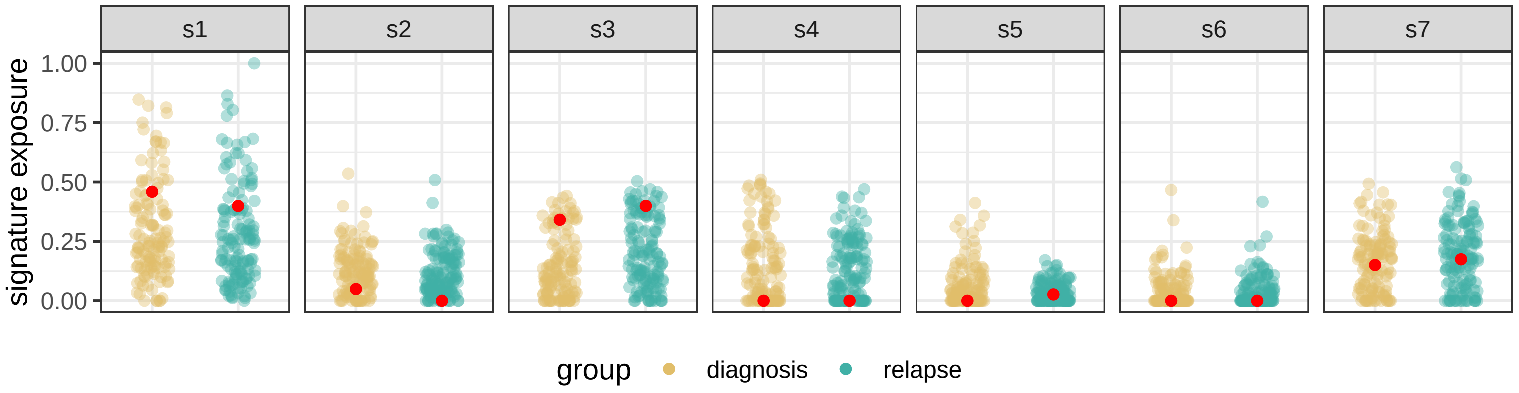

### BRITROC−205

stage: 3C

platinum status: sensitive

prior lines: 1

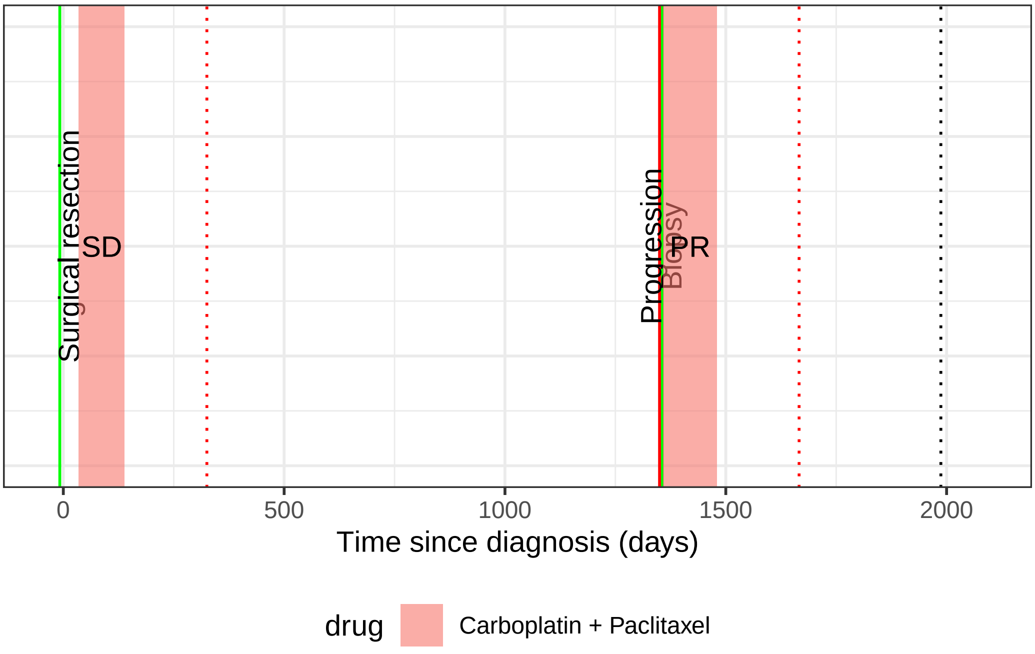

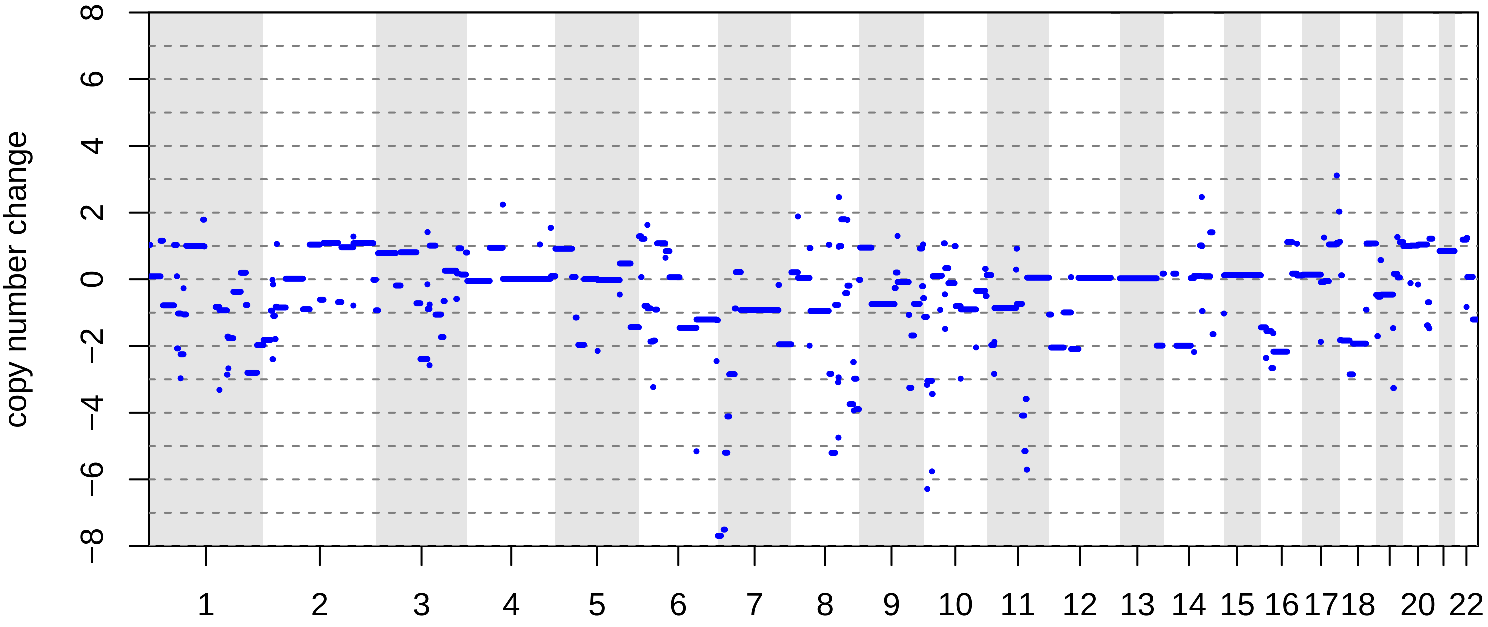

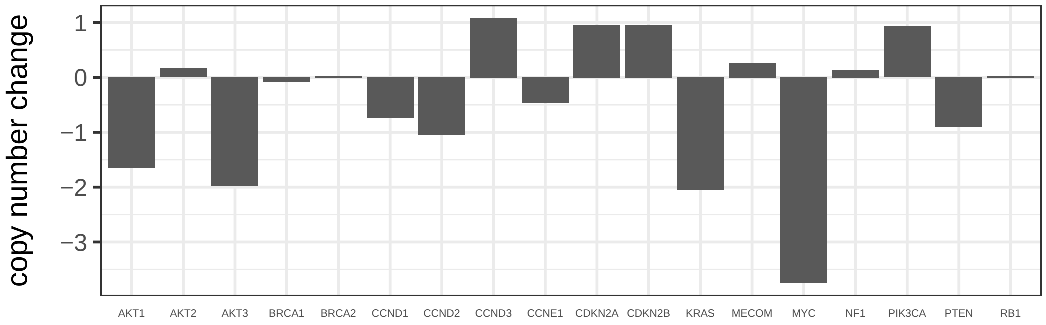

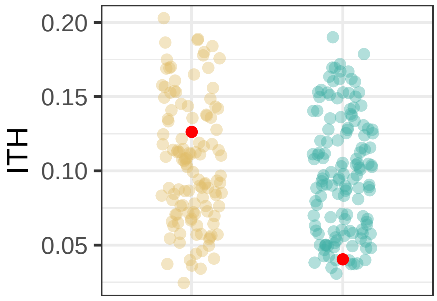

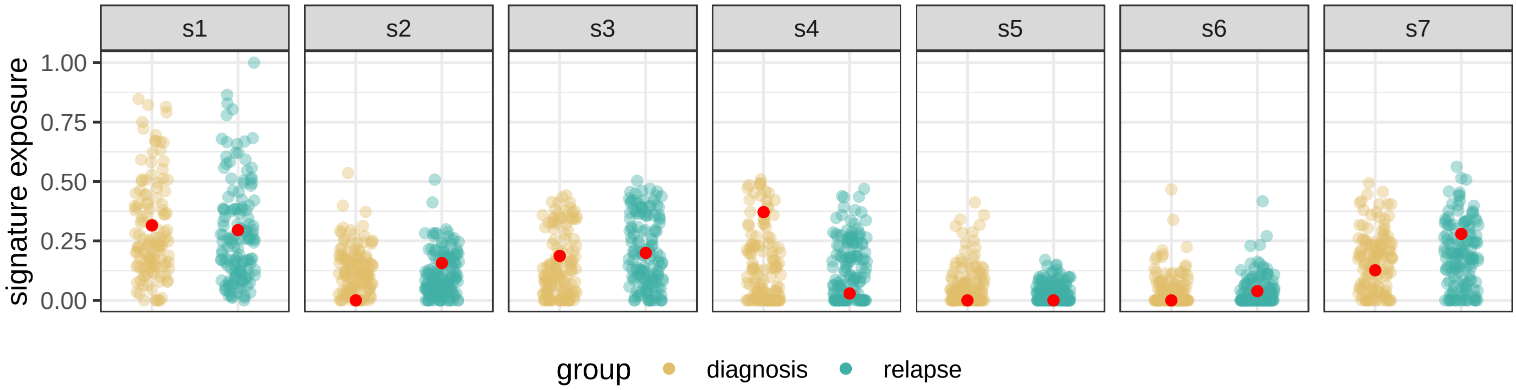

#### BRITROC−209

stage: 3C

platinum status: sensitive

prior lines: 1

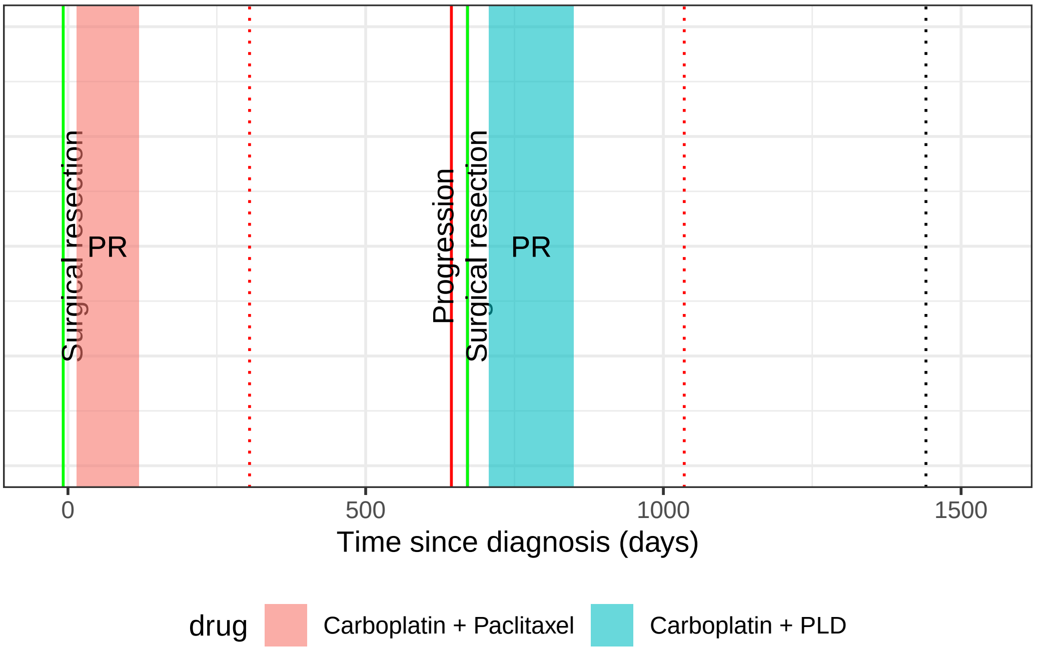

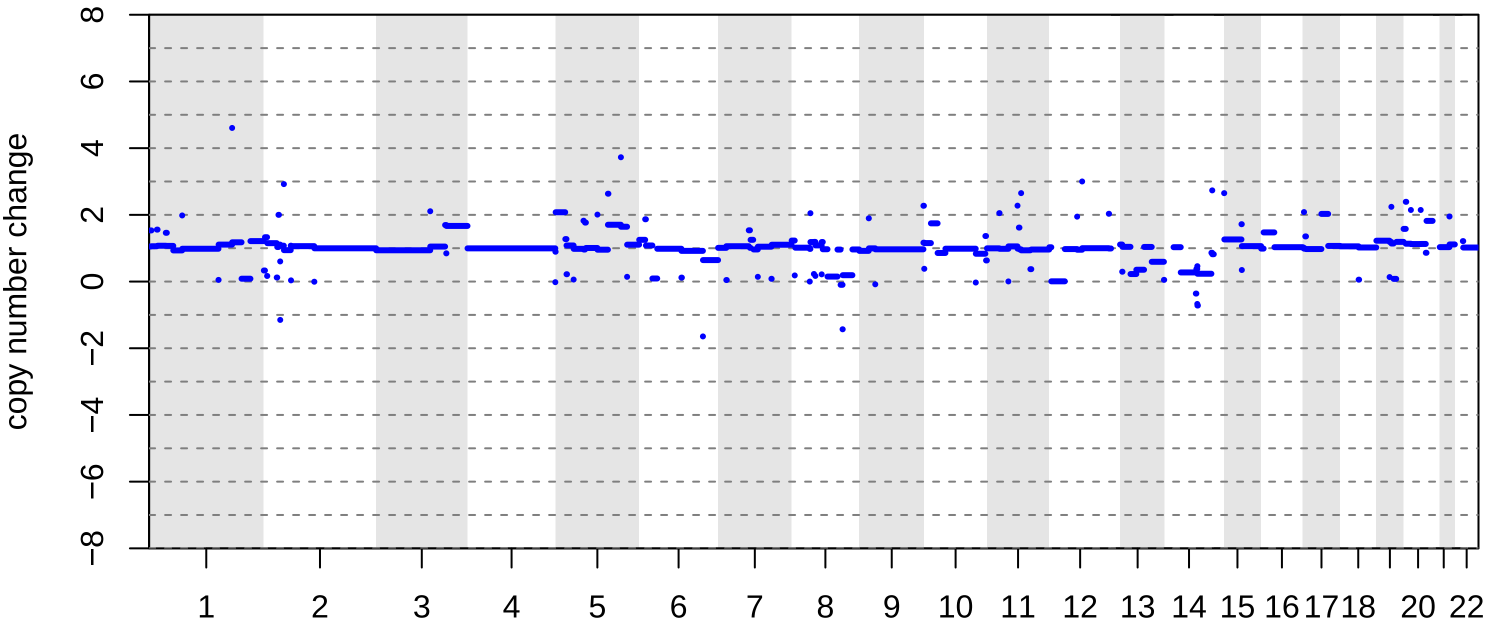

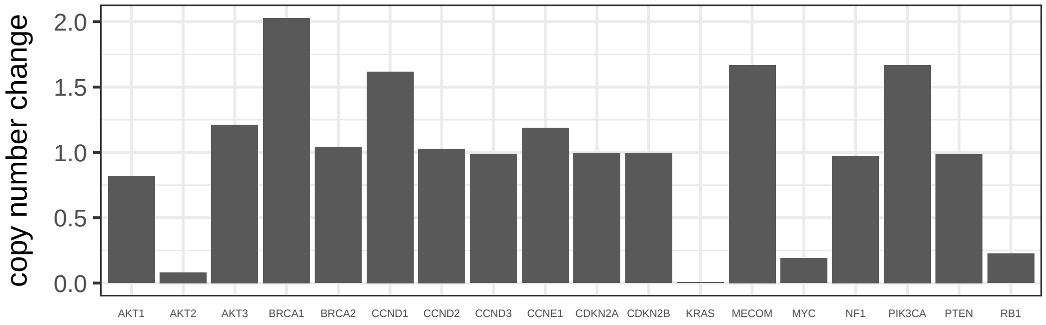

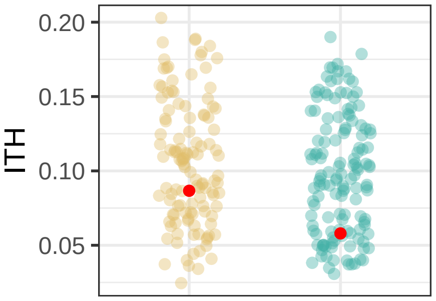

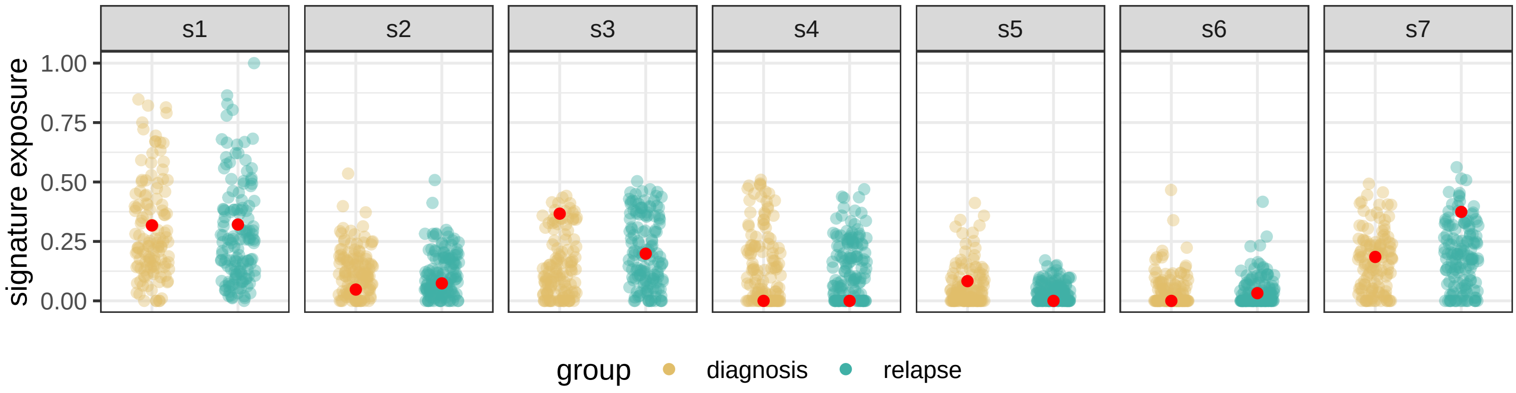

BRITROC−216

stage: 3B

platinum status: sensitive

prior lines: 1

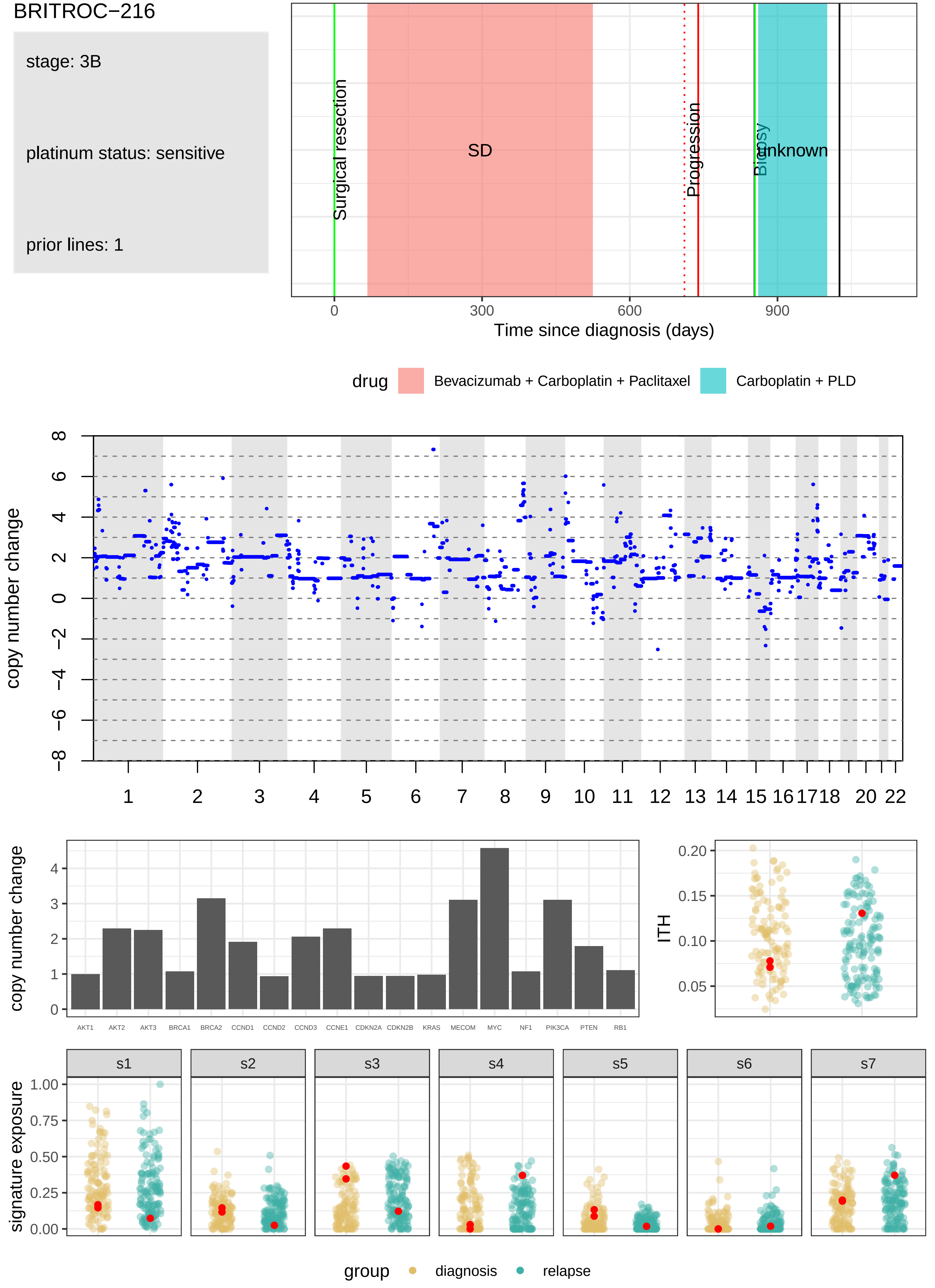

BRITROC−225

stage: 3C

platinum status: sensitive

prior lines: 1

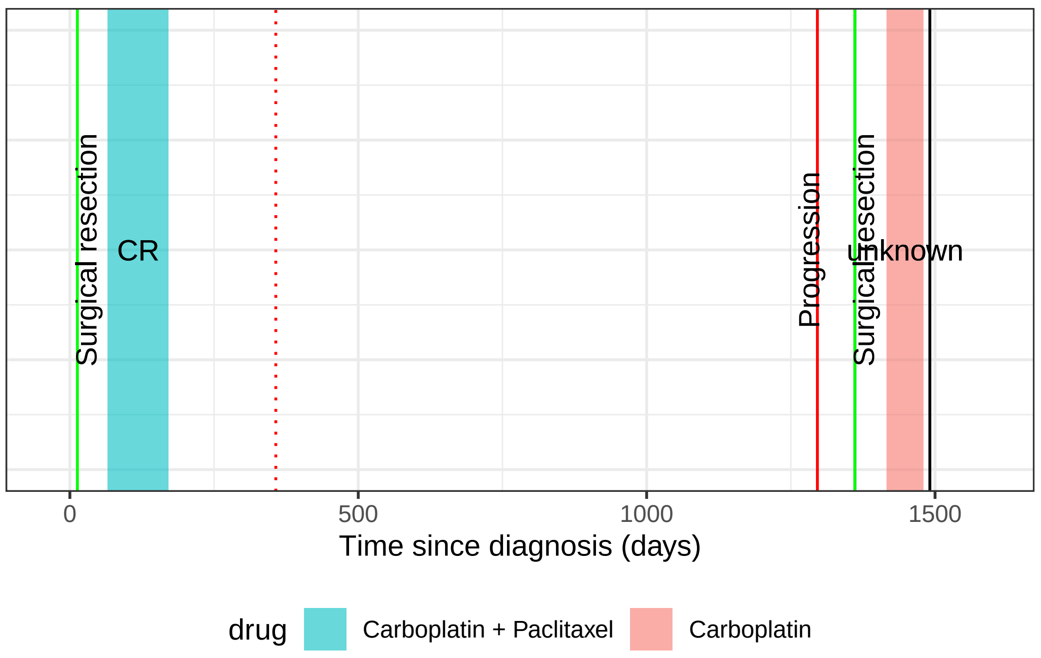

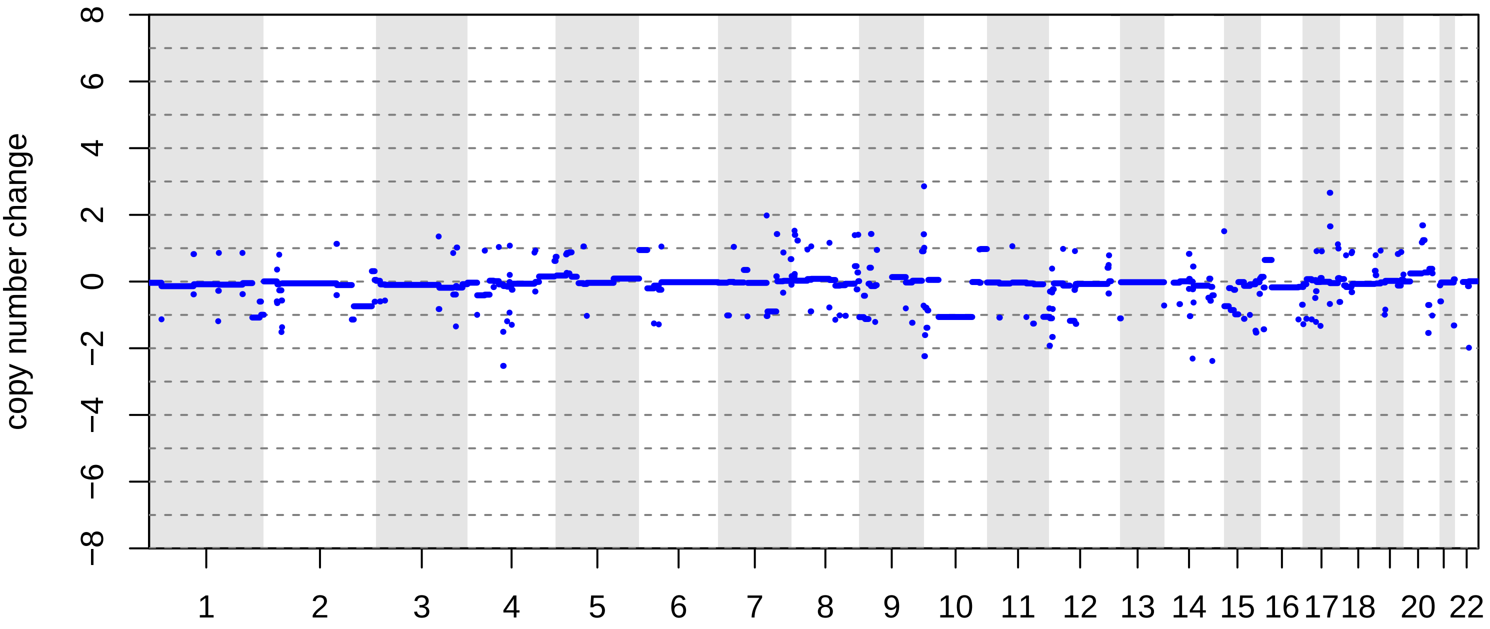

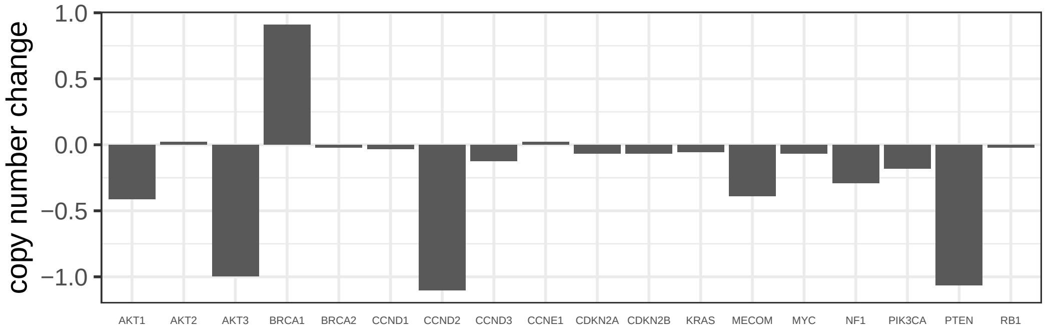

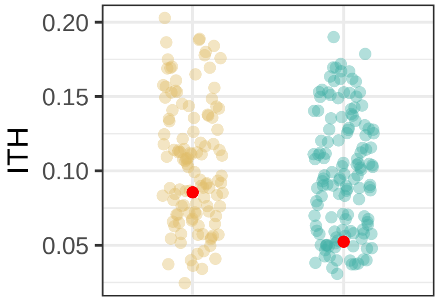

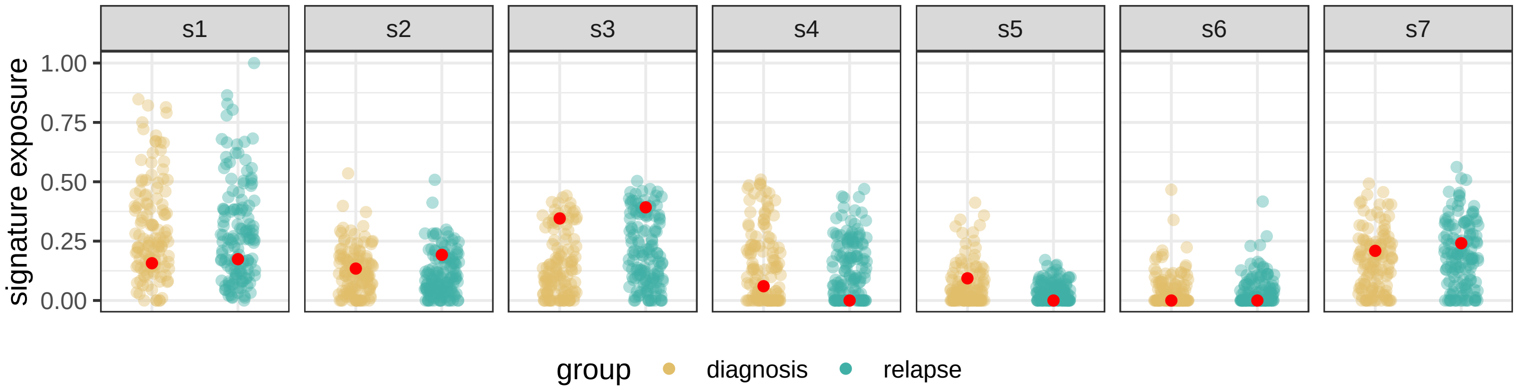

BRITROC−226

stage: 3B

platinum status: sensitive

prior lines: 1

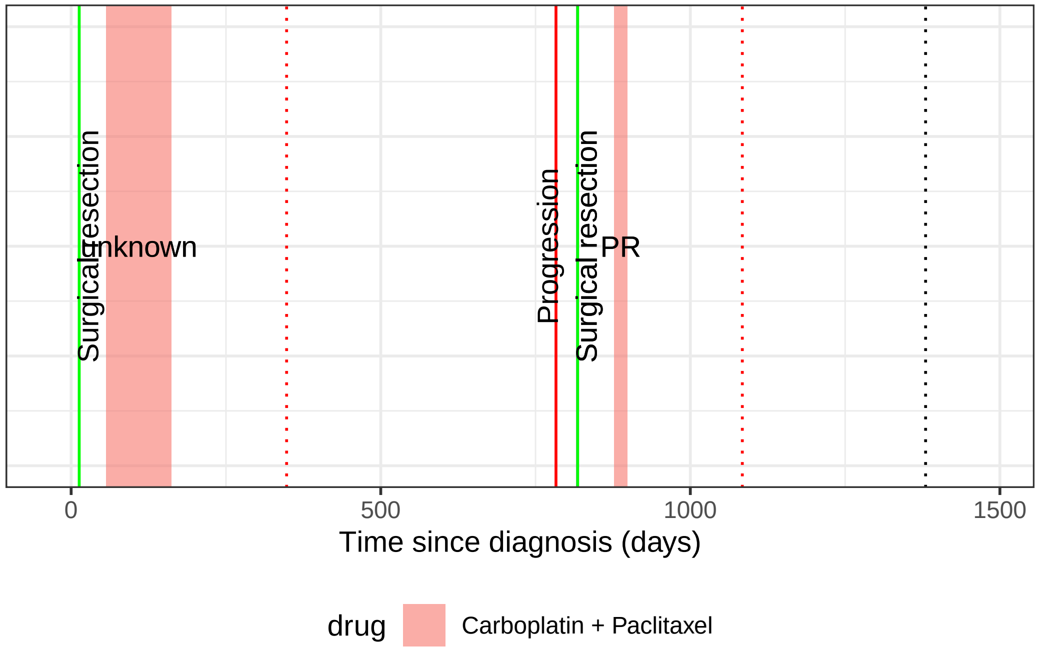

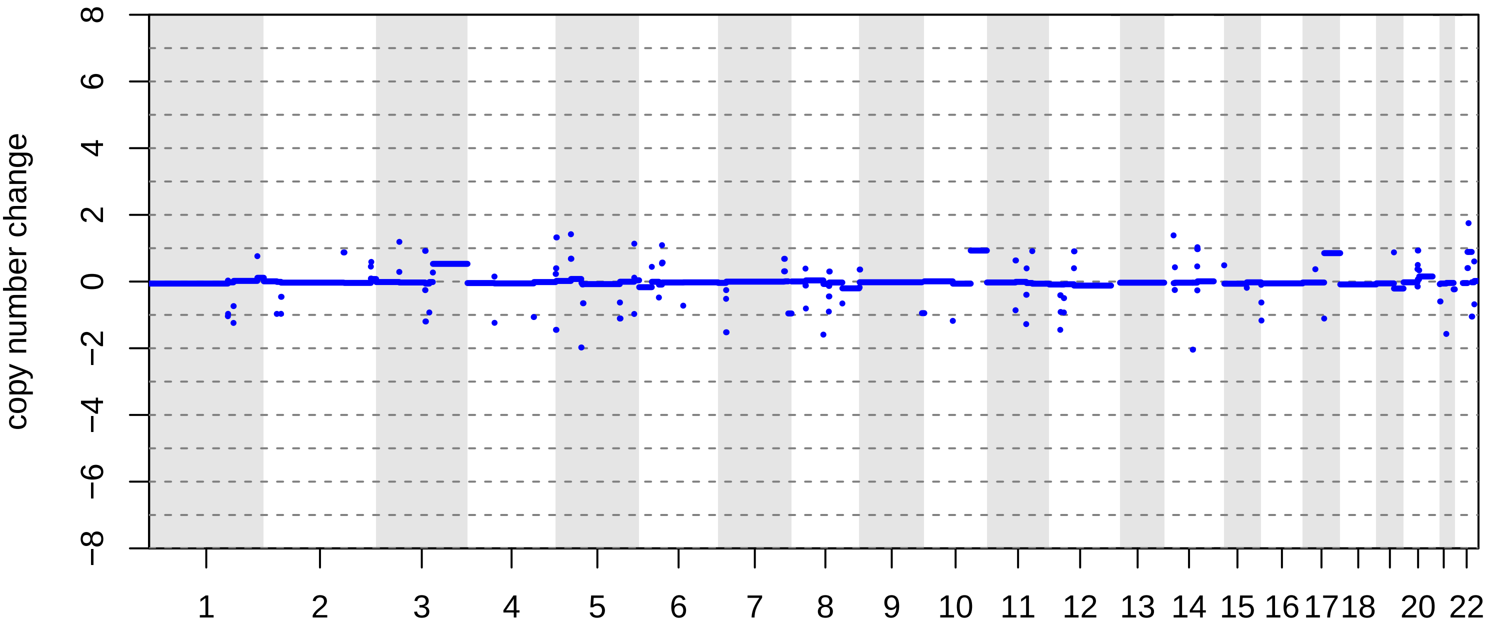

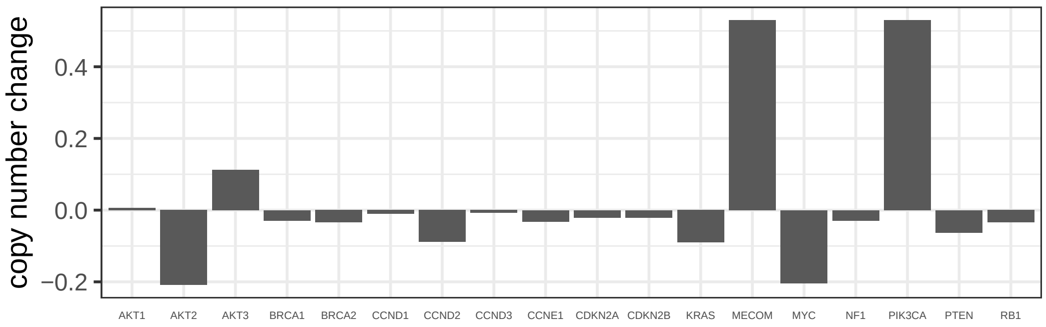

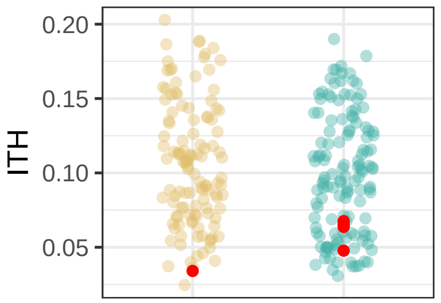

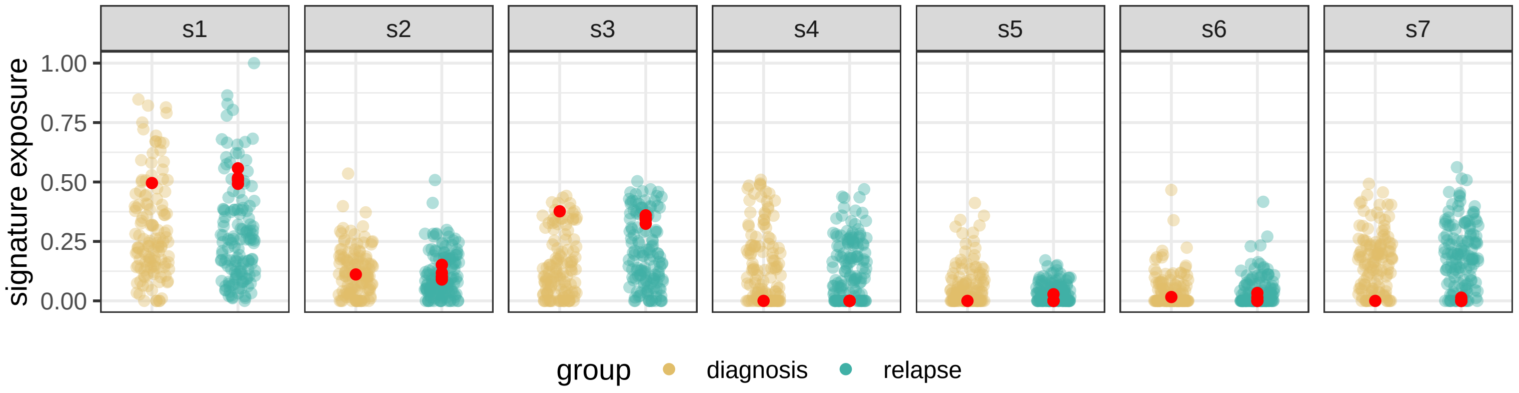

BRITROC−229

stage: 3C

platinum status: resistant

prior lines: 1

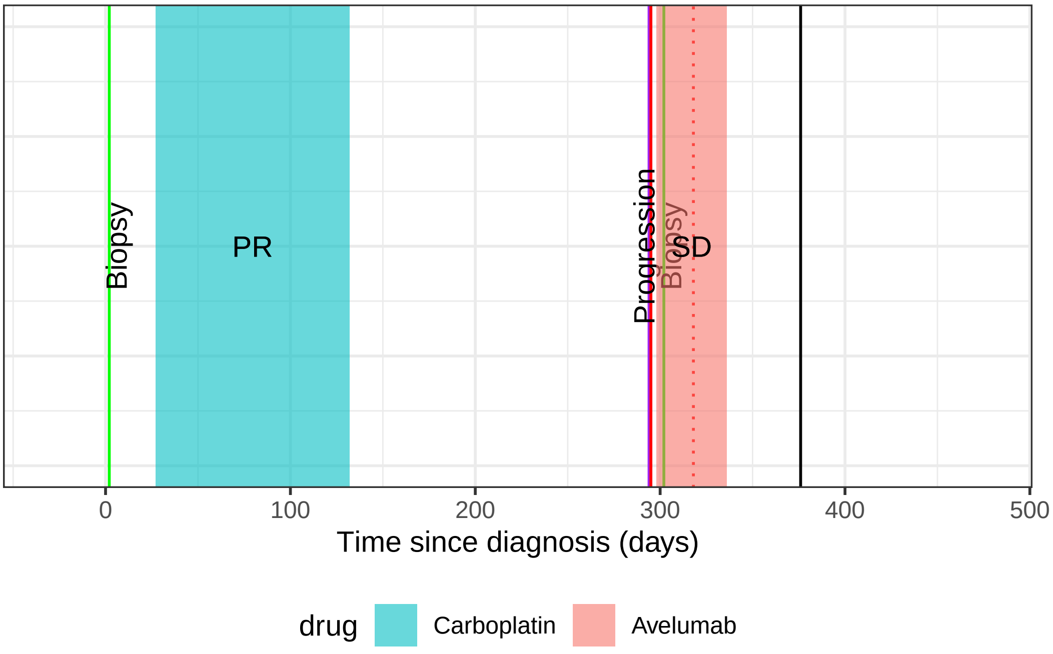

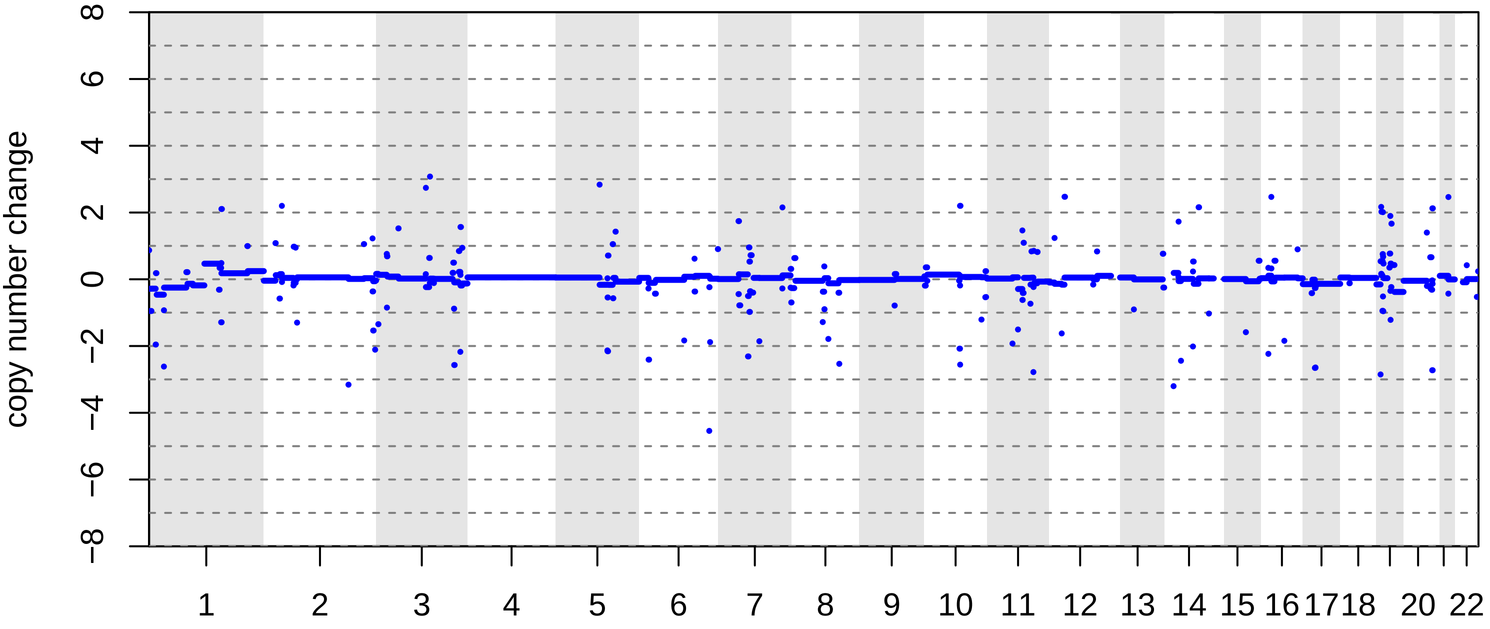

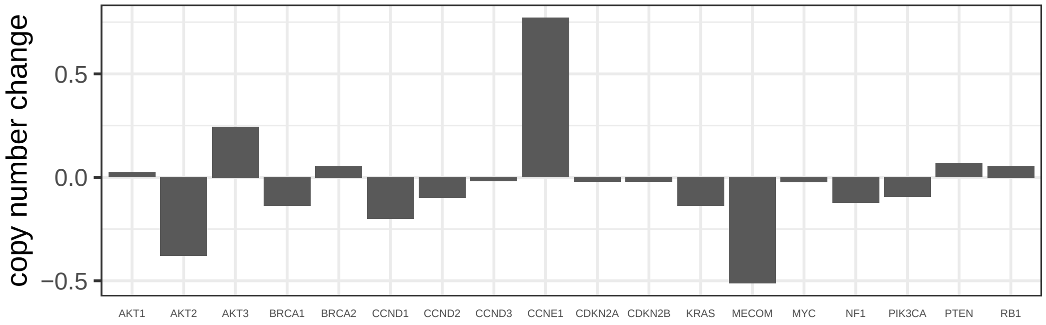

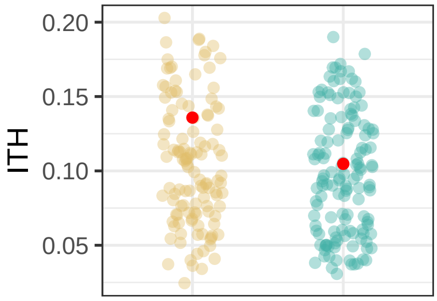

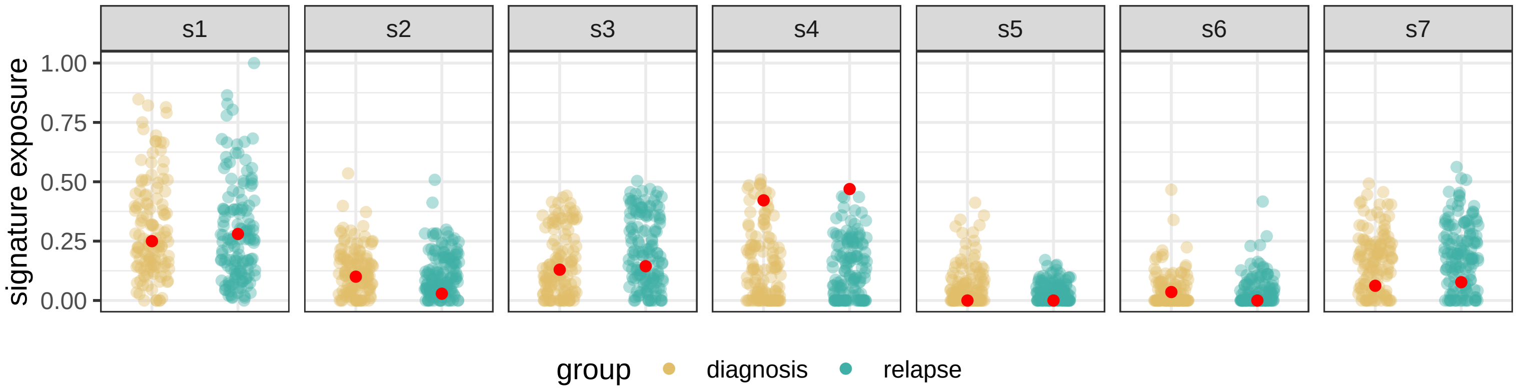

### BRITROC−23

stage: 3C

platinum status: sensitive

prior lines: 2

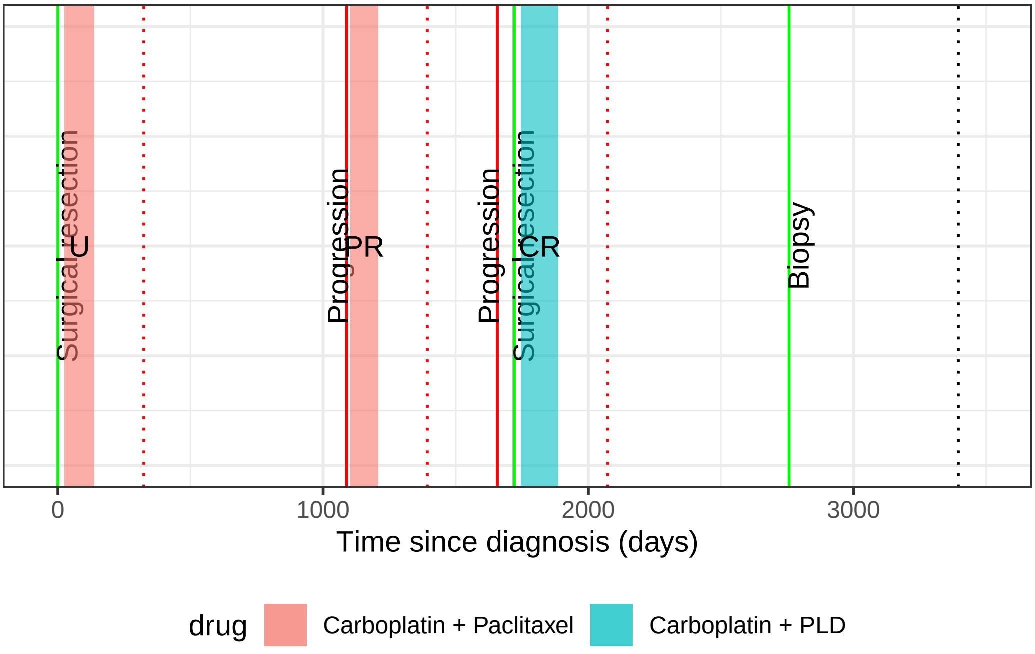

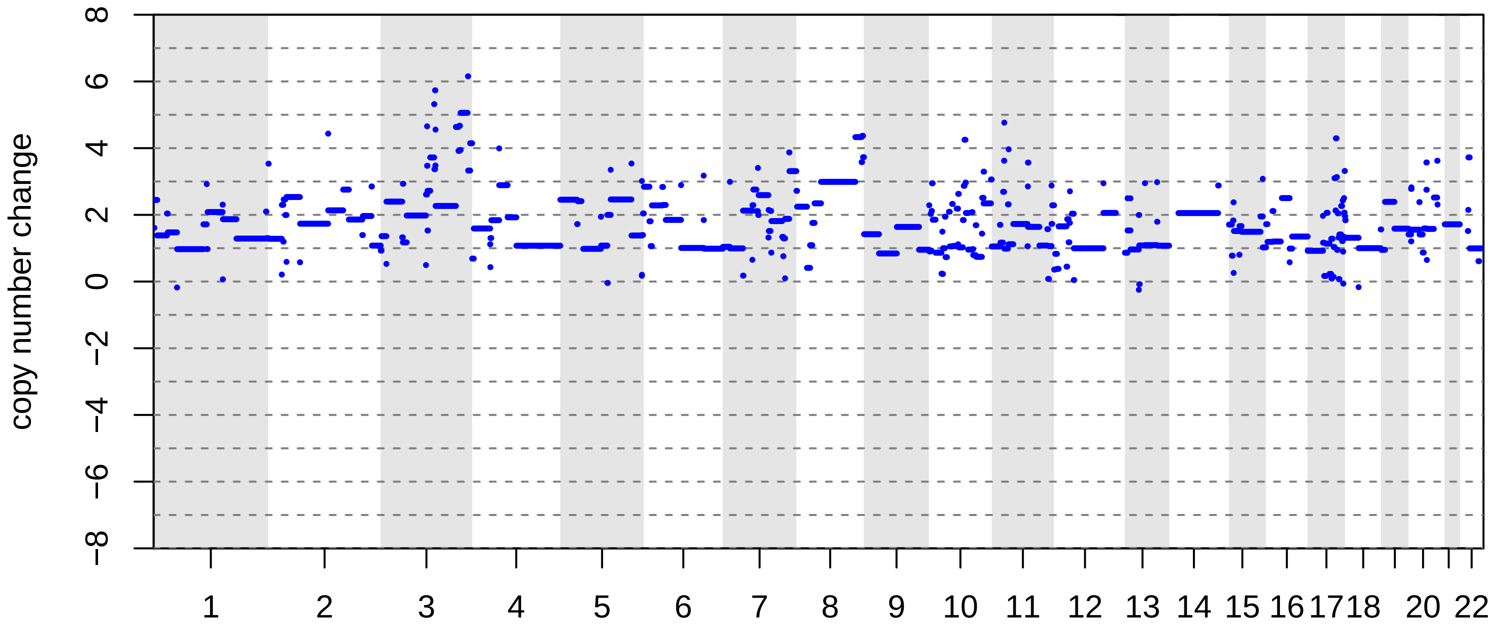

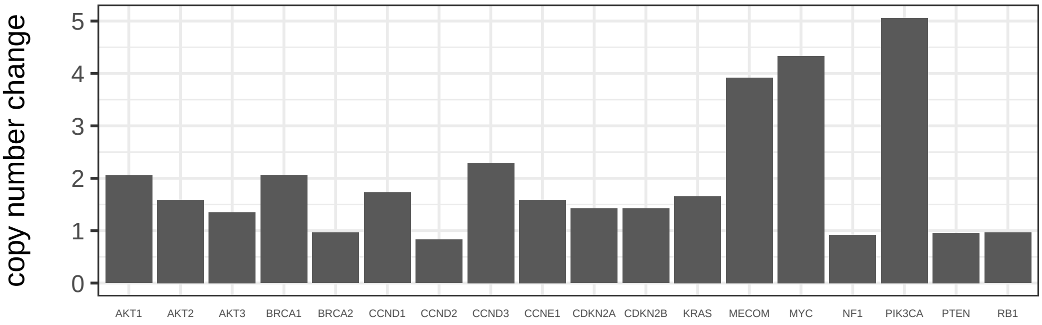

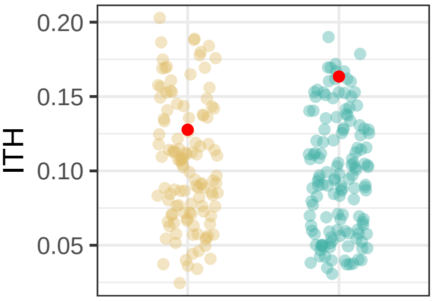

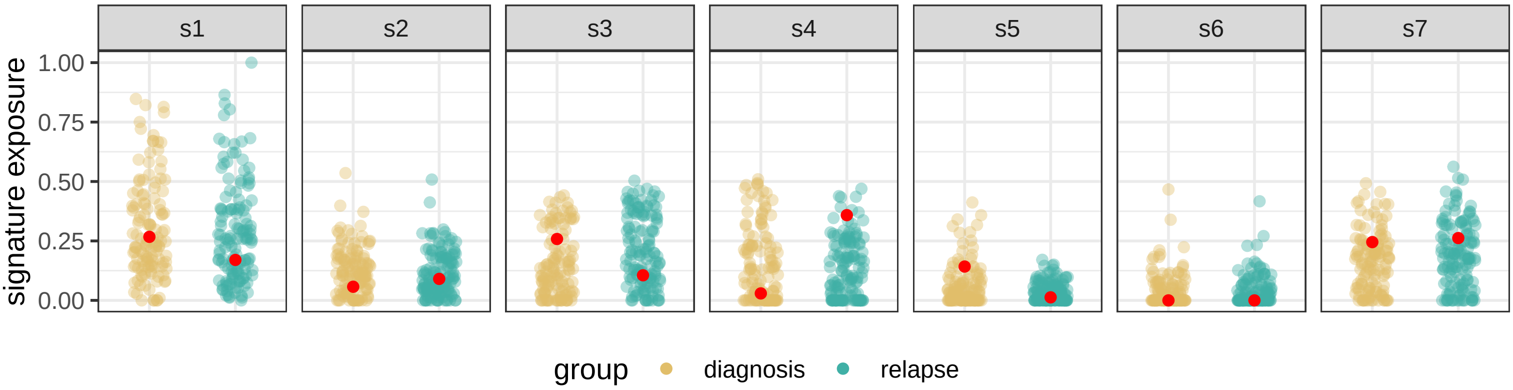

BRITROC−232

stage: 2B

platinum status: sensitive

prior lines: 1

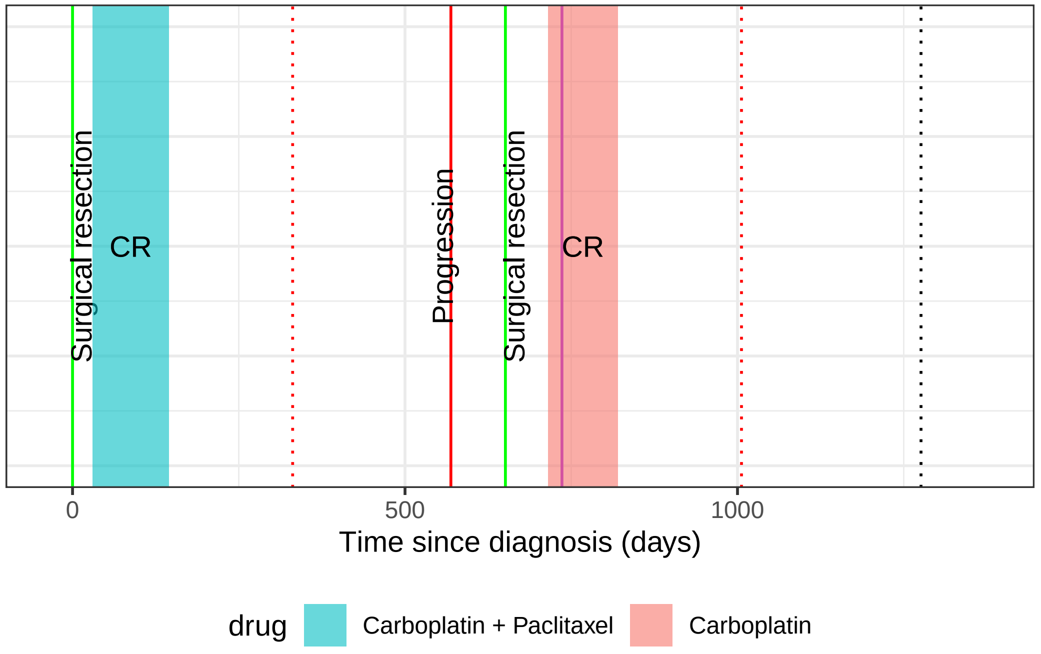

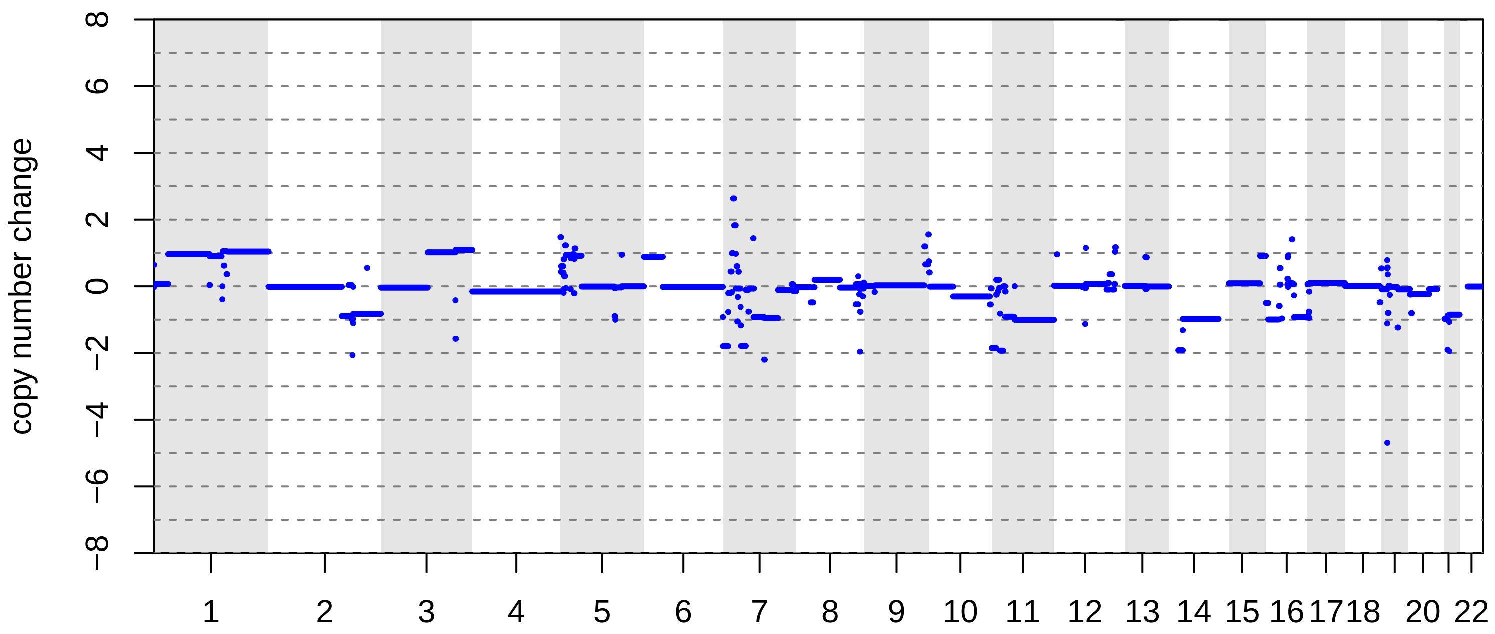

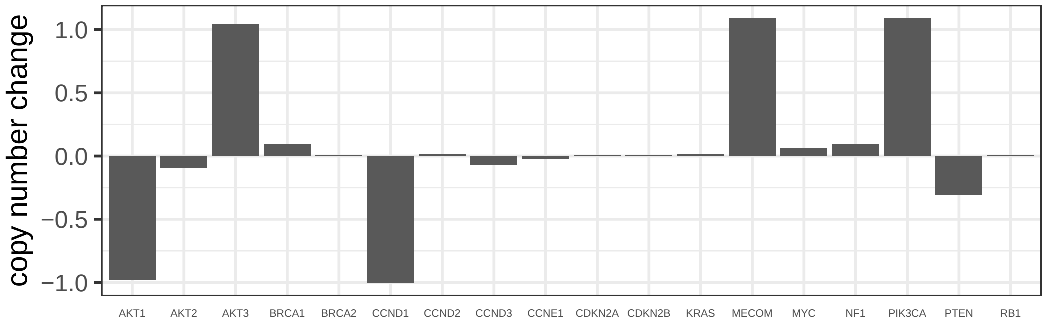

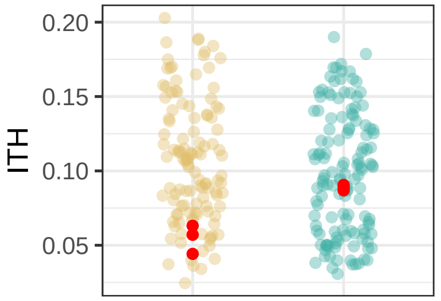

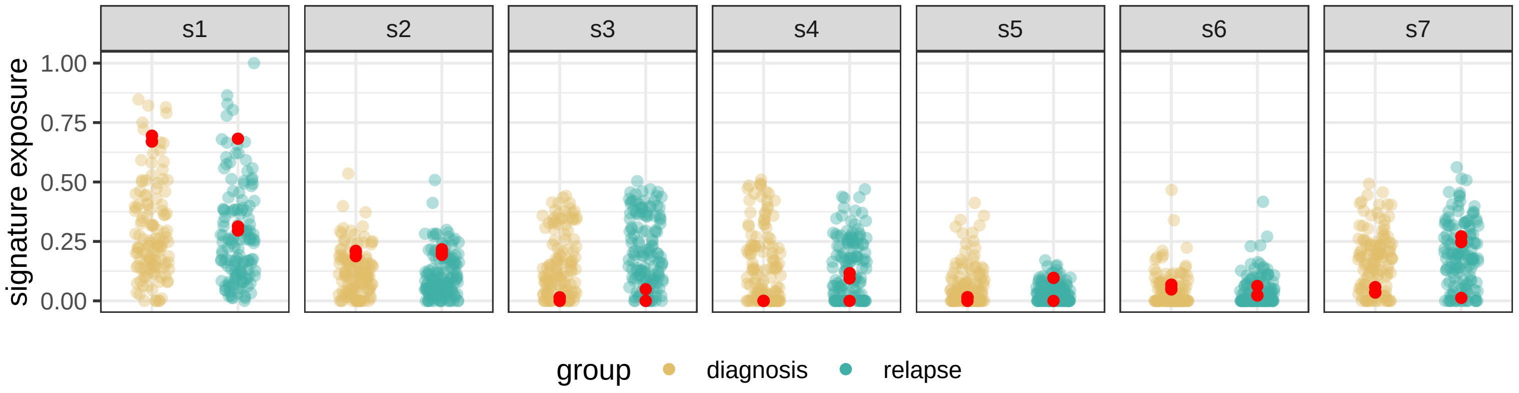

BRITROC−234

stage: 4NA

platinum status: resistant

prior lines: 1

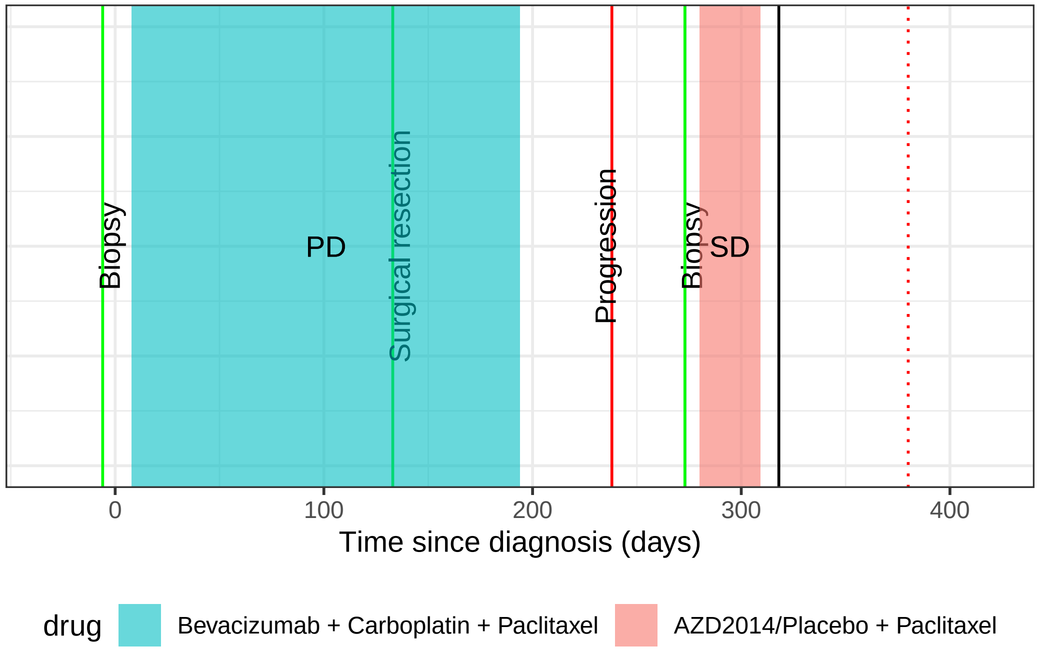

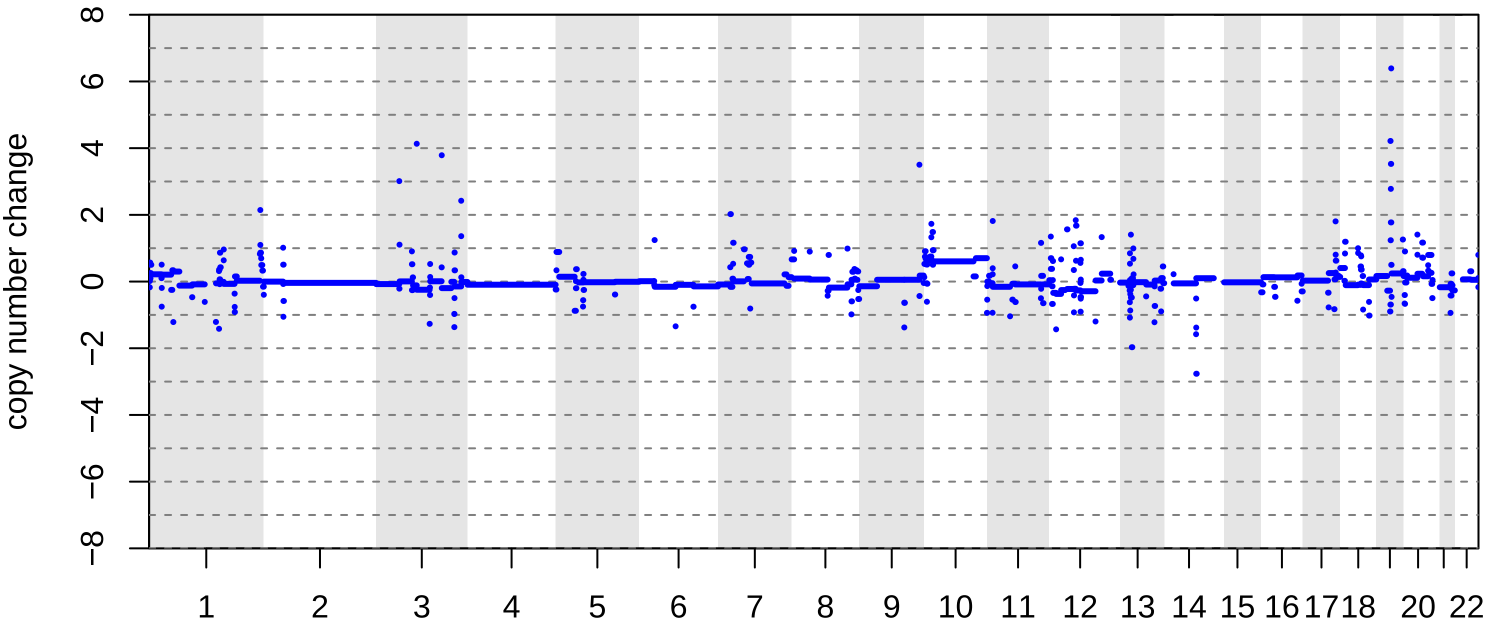

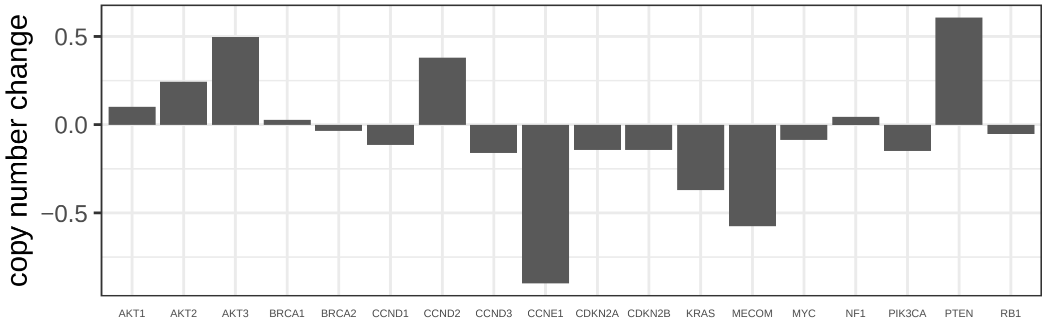

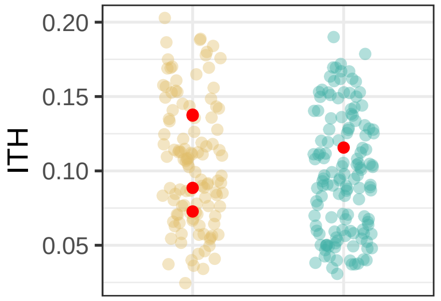

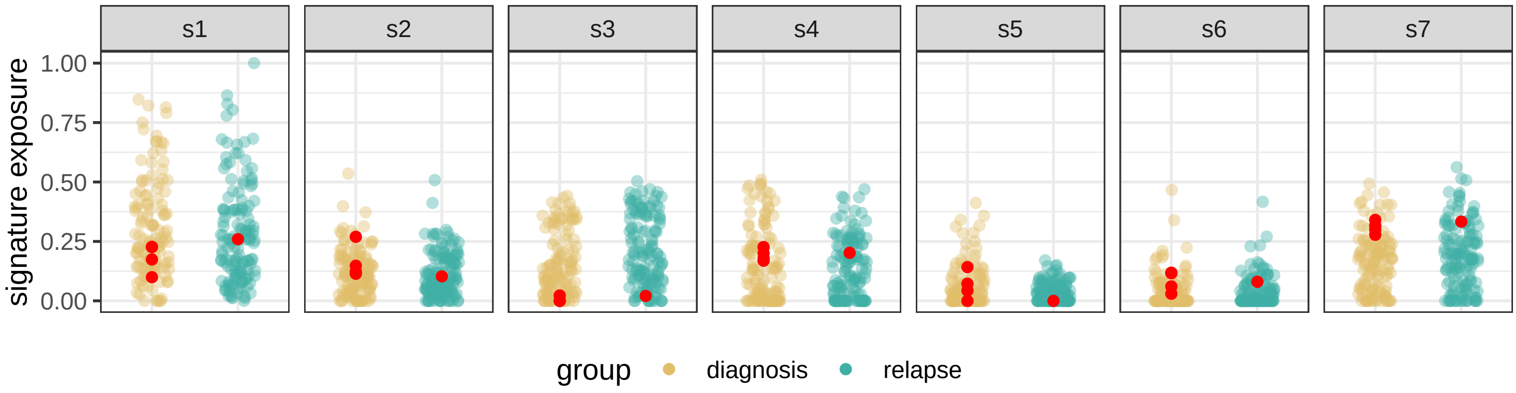

BRITROC−241

stage: 4NA

platinum status: sensitive

prior lines: 3

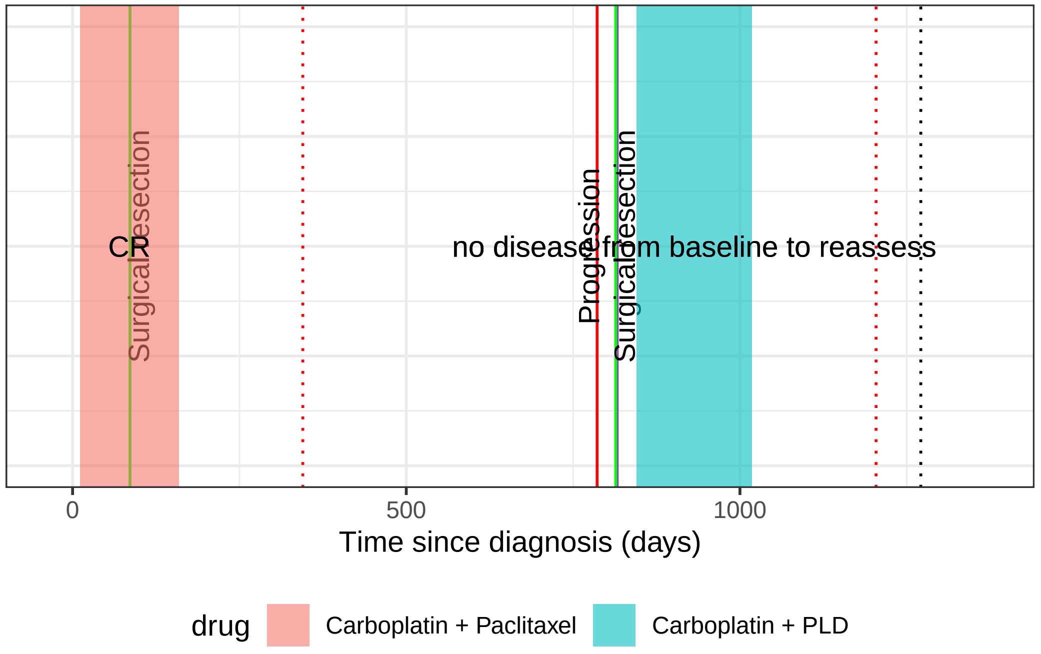

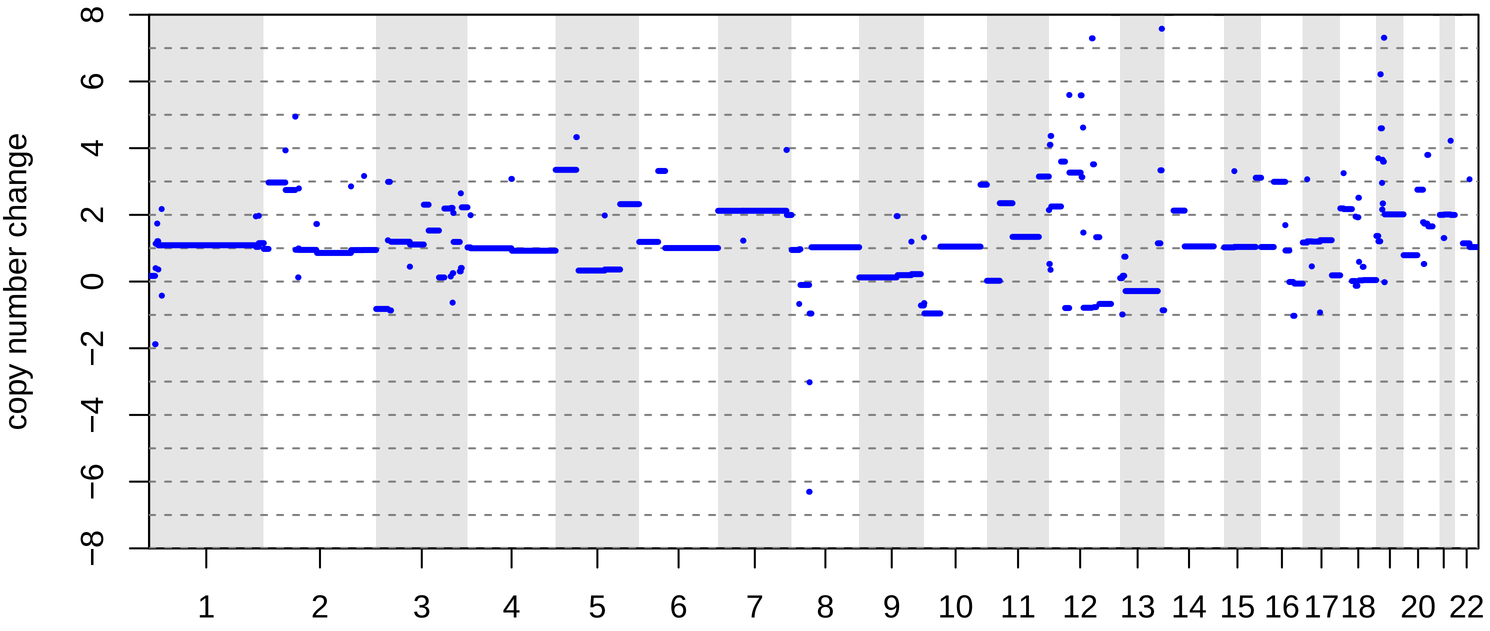

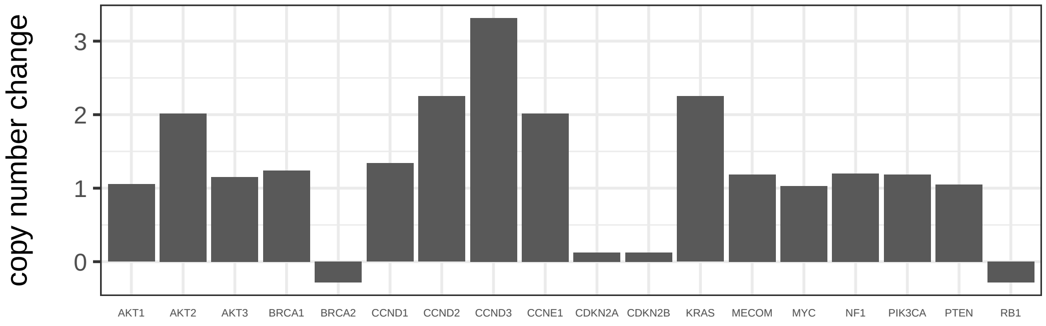

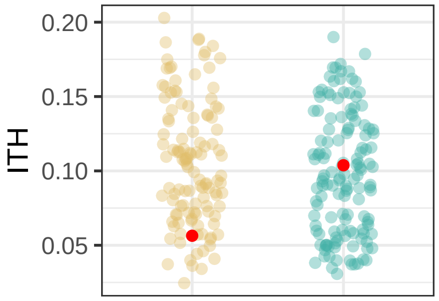

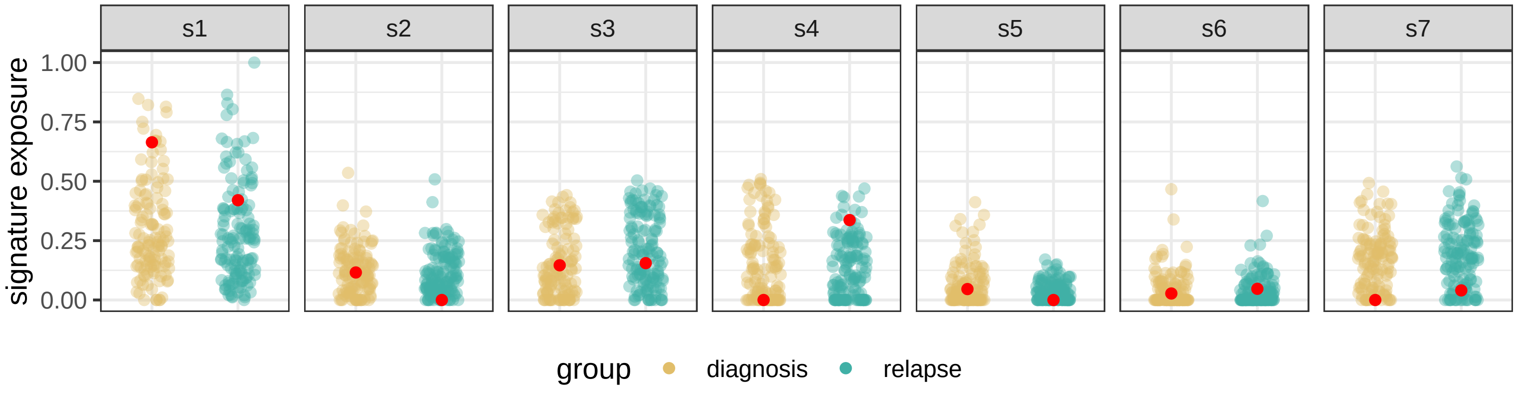

BRITROC−242

stage: 3C

platinum status: sensitive

prior lines: 1

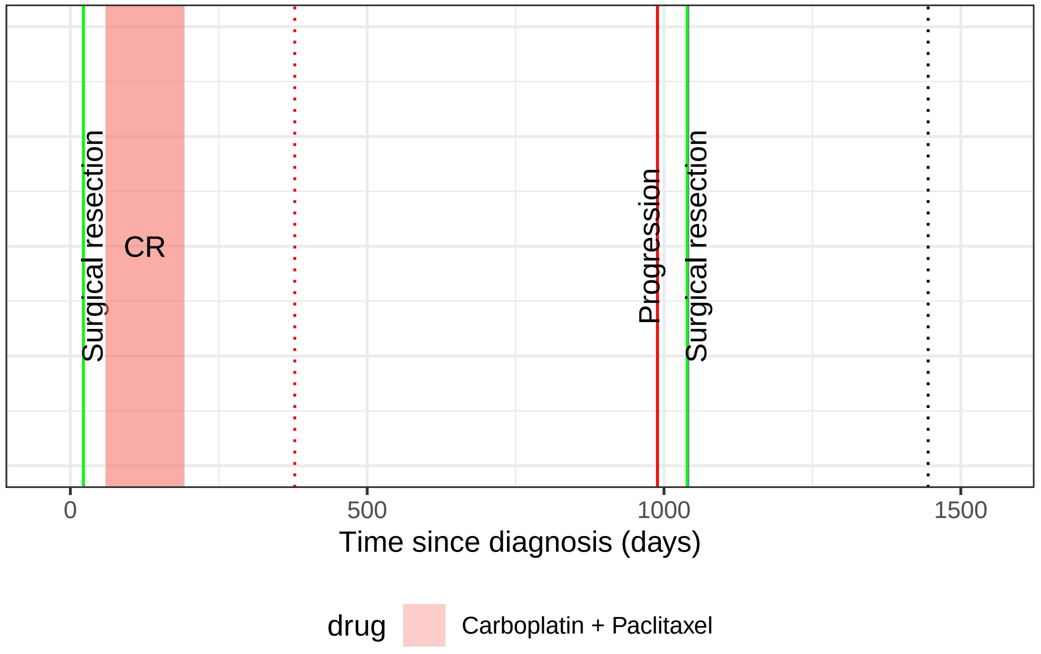

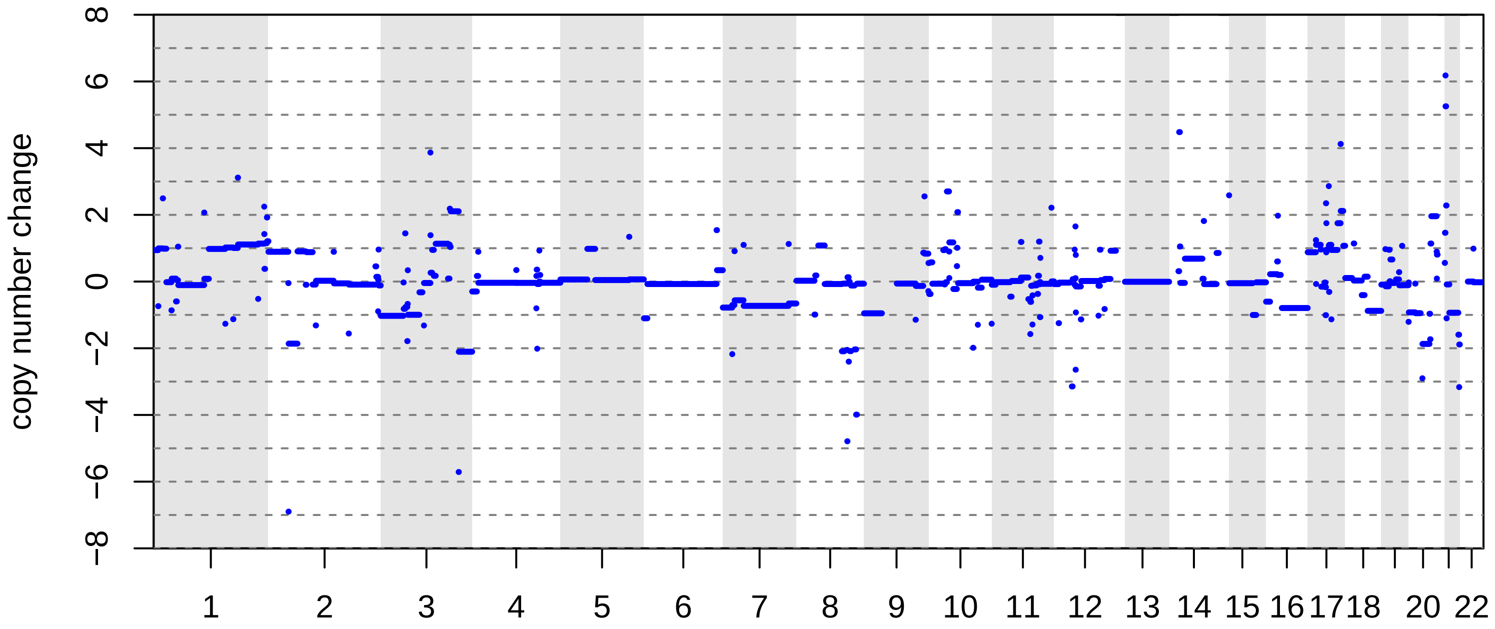

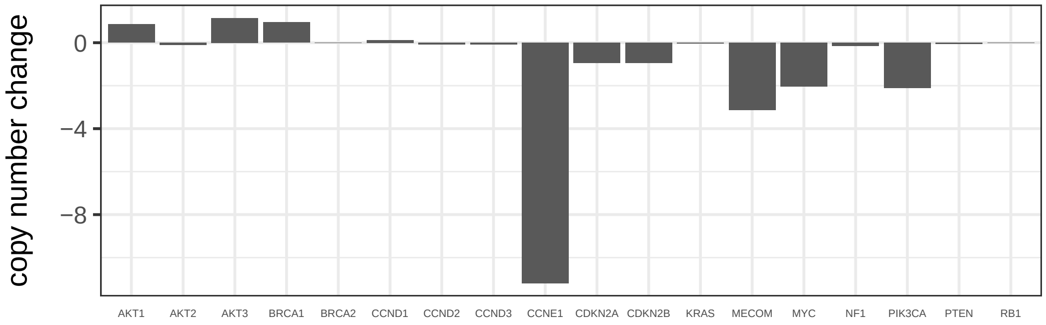

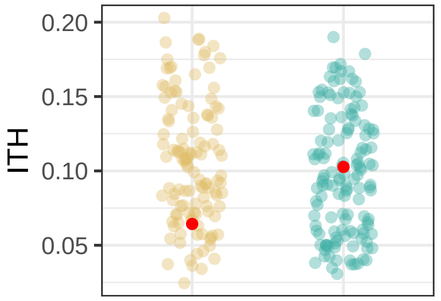

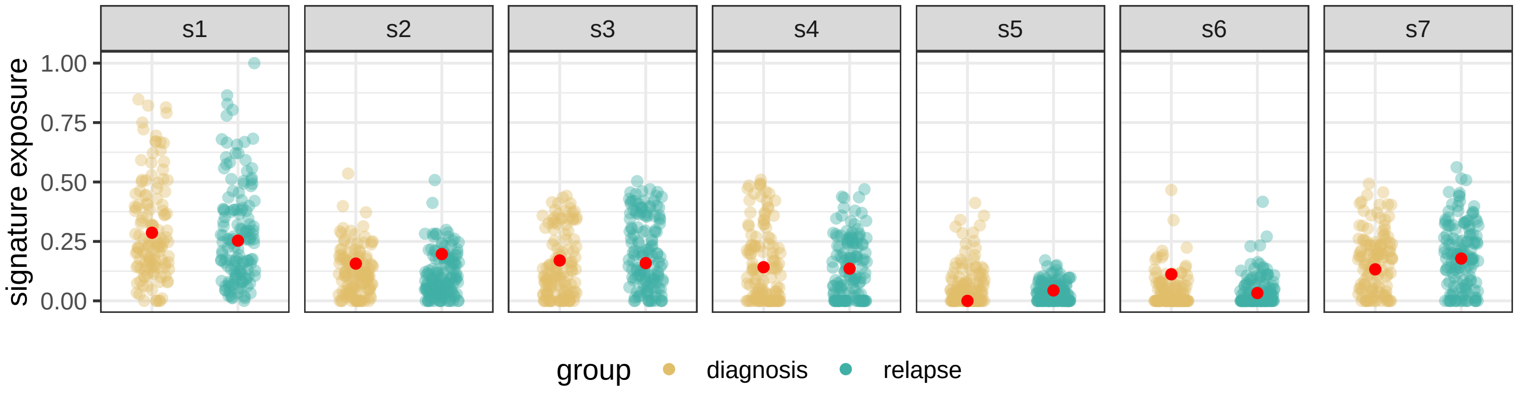

BRITROC−246

stage: 3C

platinum status: sensitive

prior lines: 1

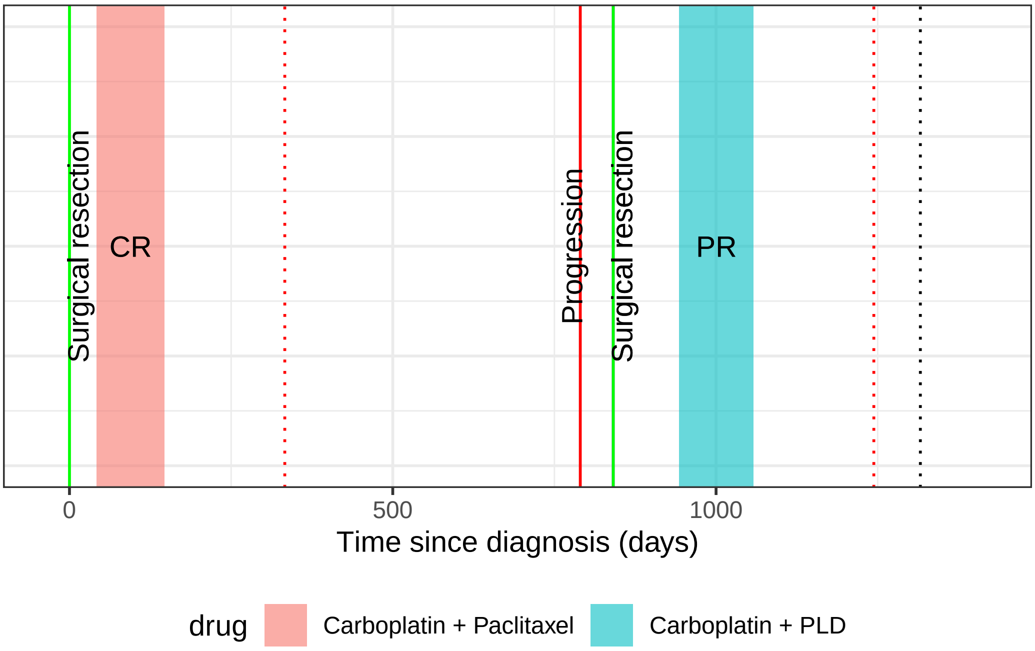

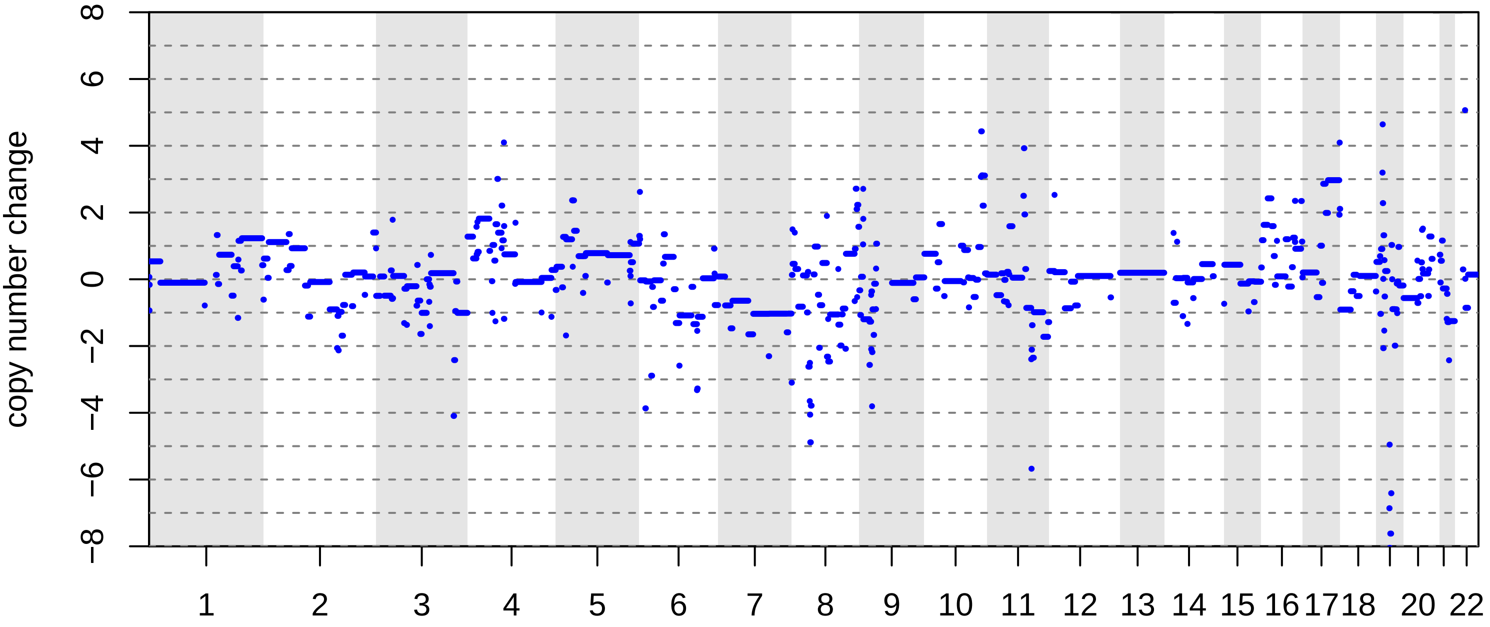

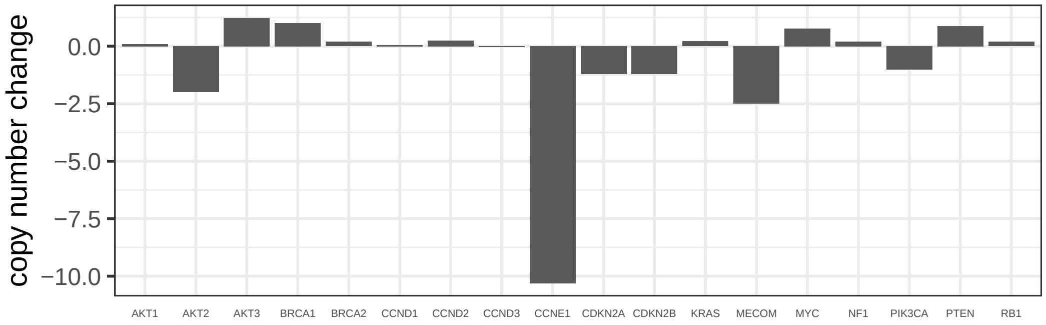

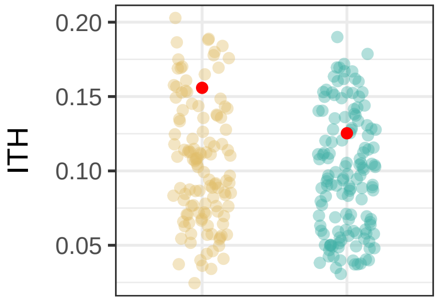

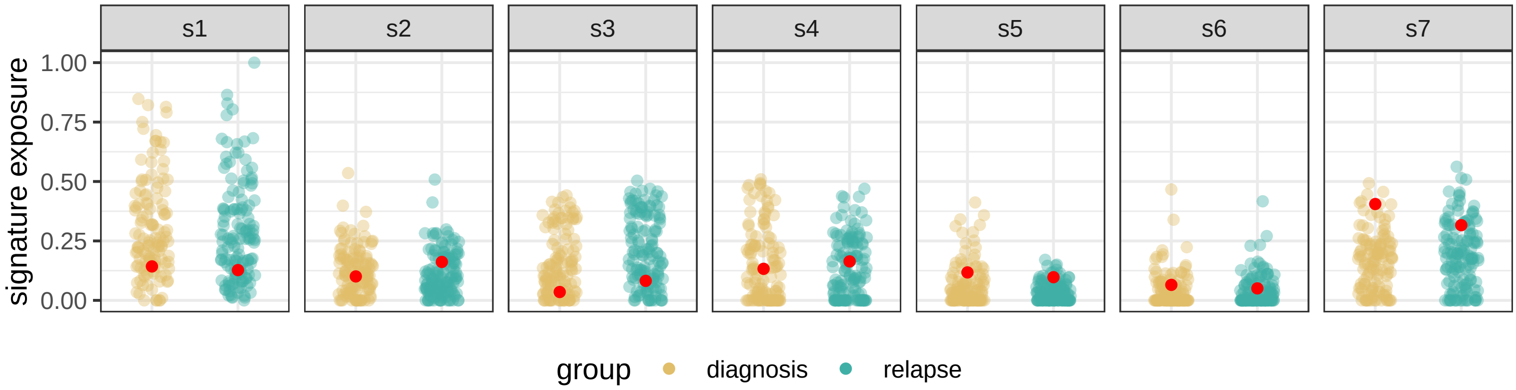

BRITROC−248

stage: 4NA

platinum status: resistant

prior lines: 2

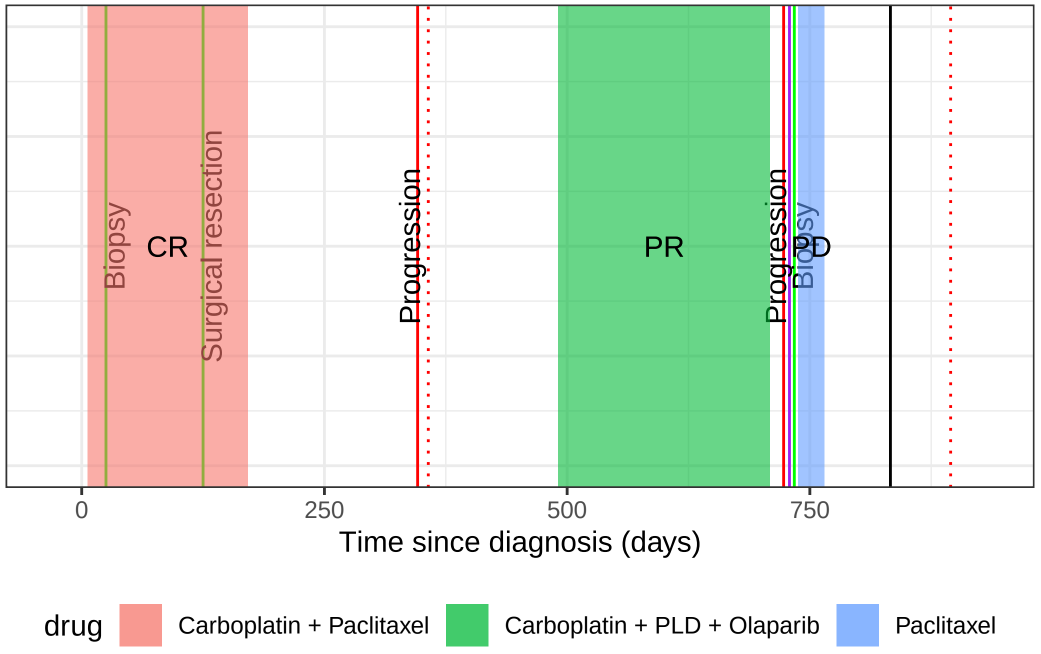

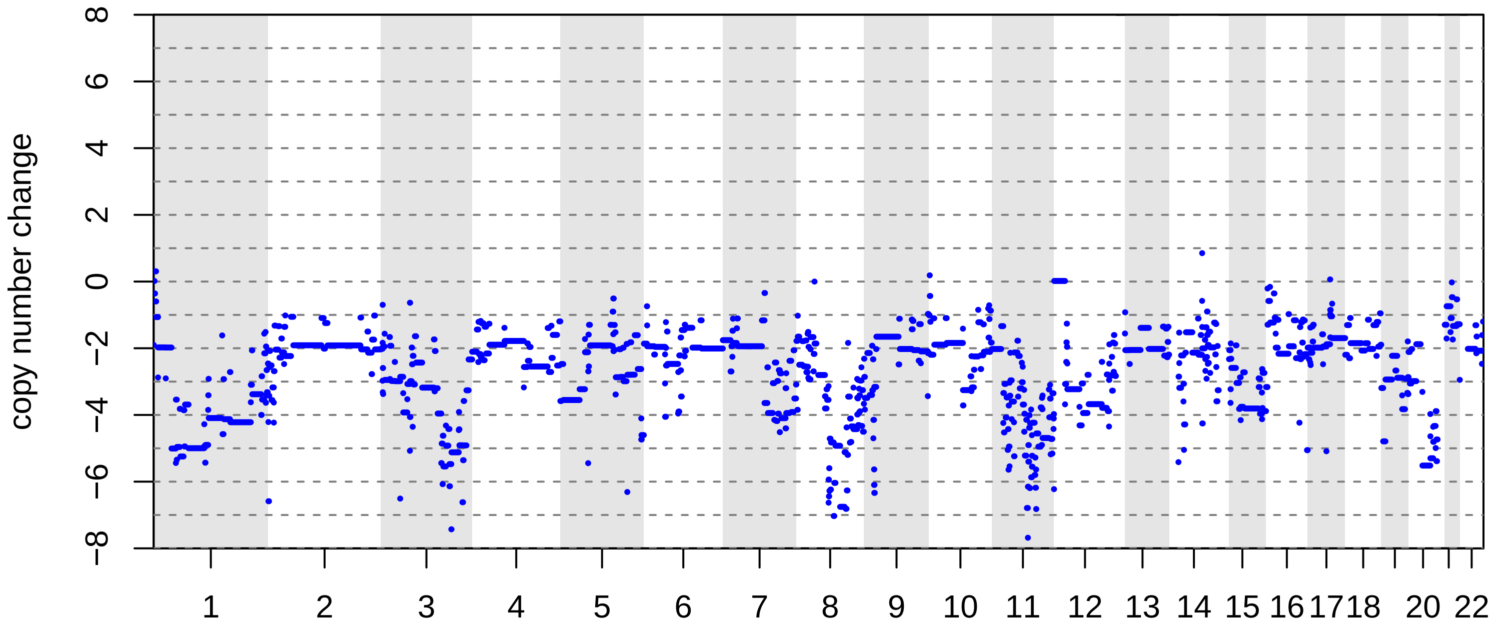

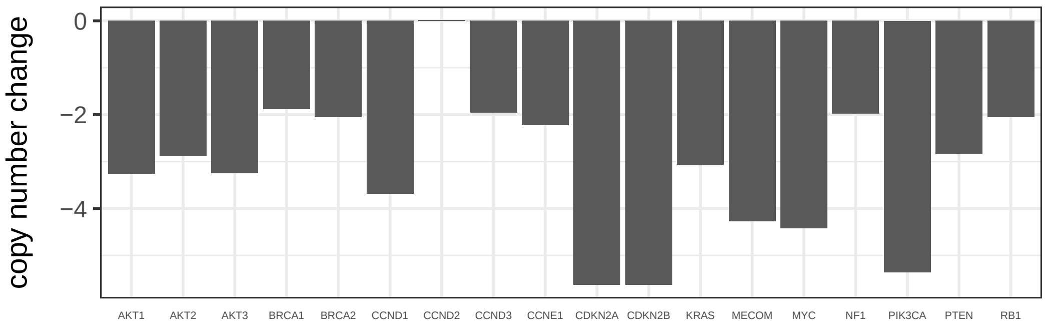

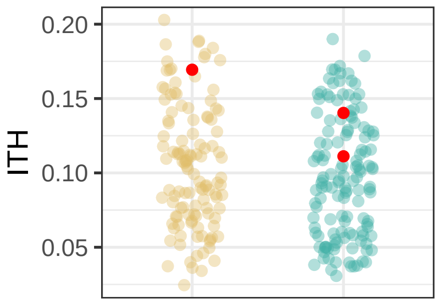

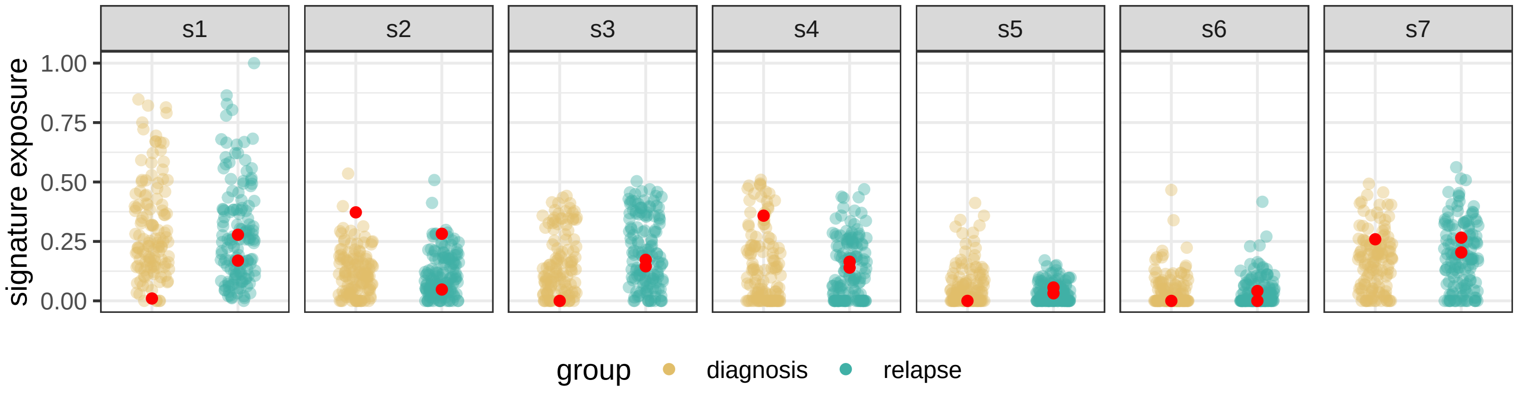

#### BRITROC−256

stage: 3C

platinum status: sensitive

prior lines: 2

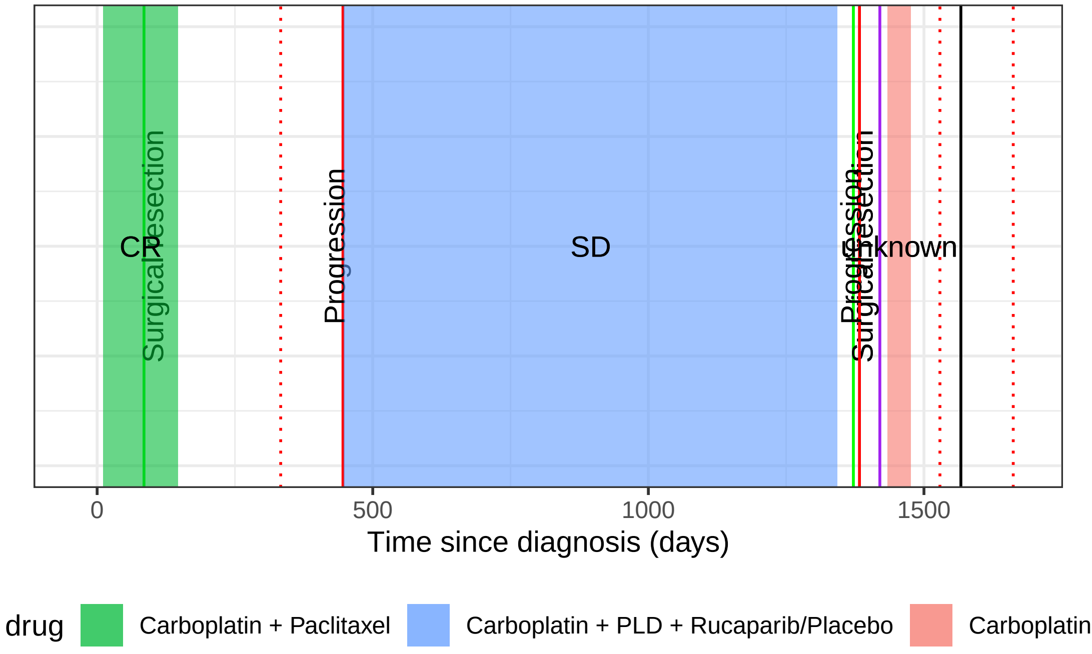

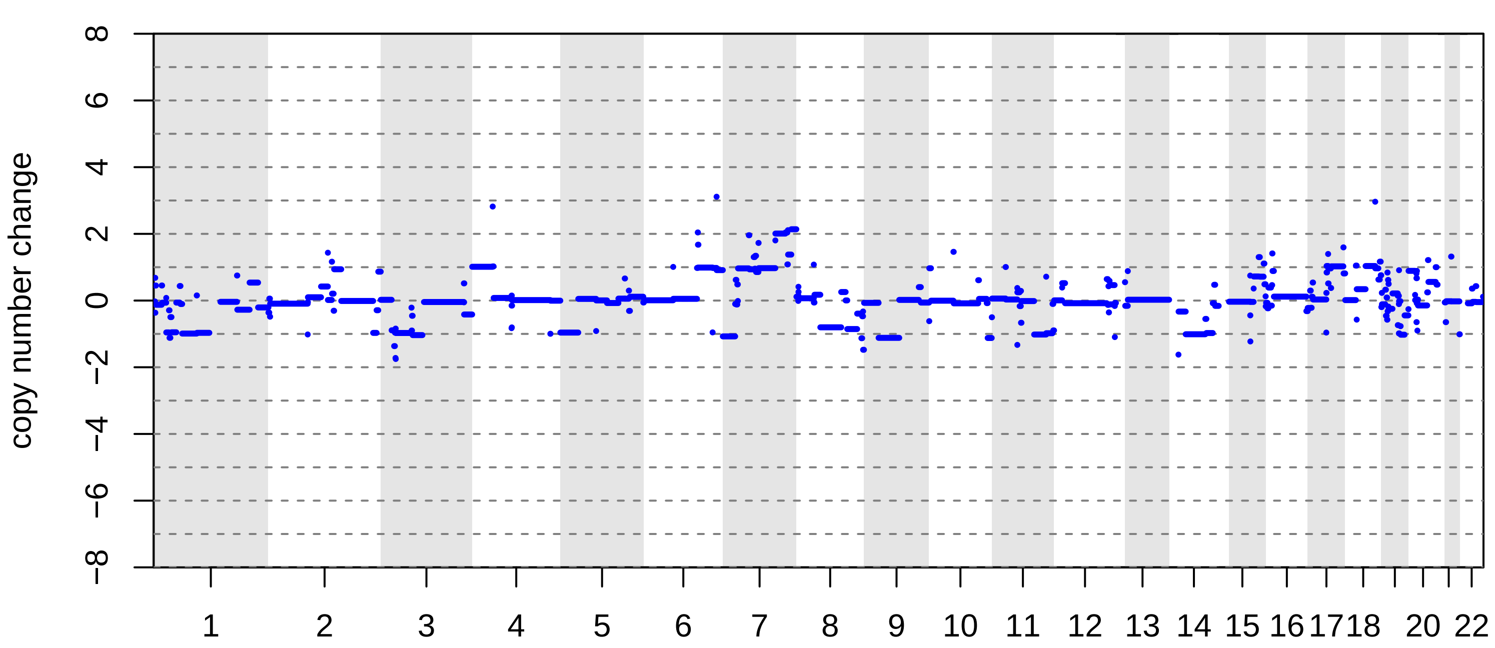

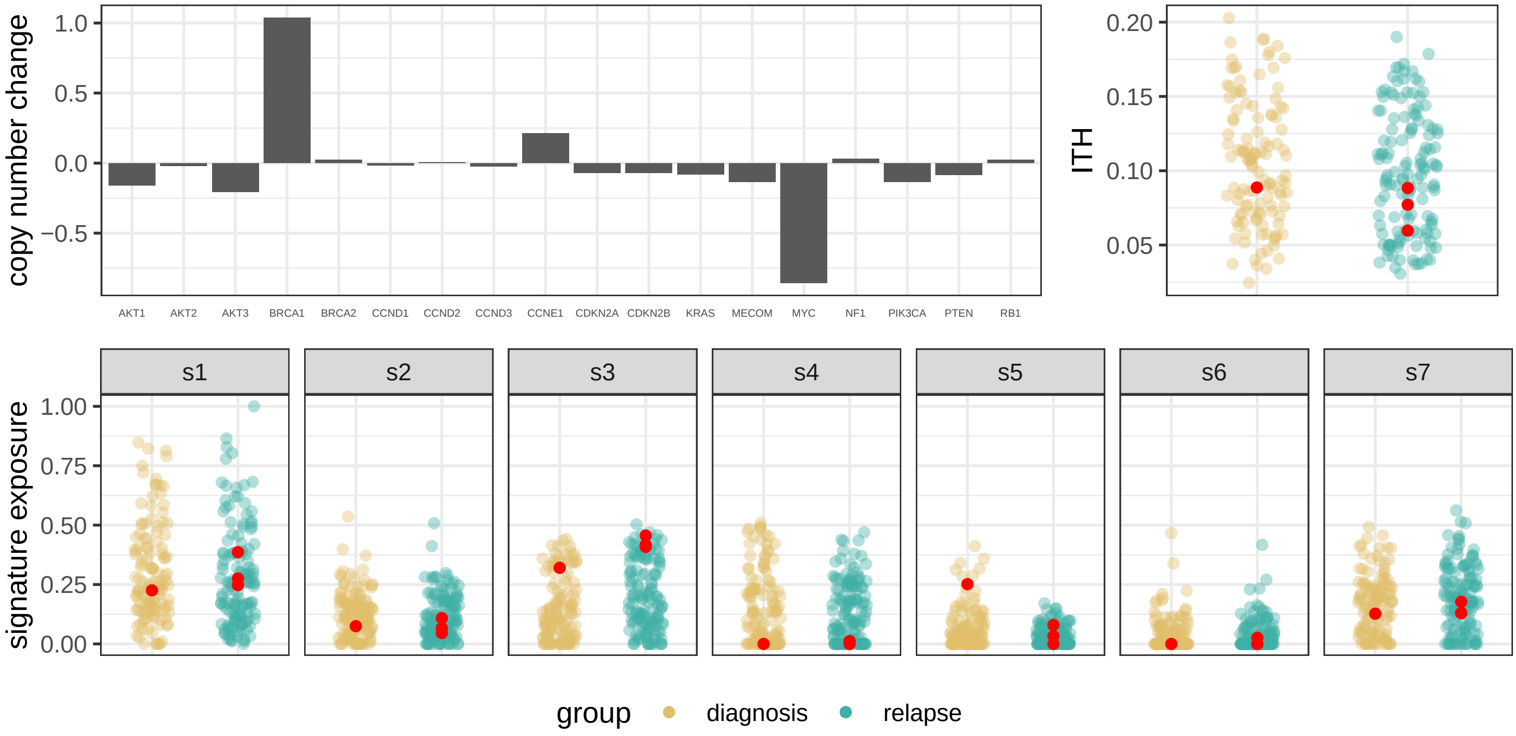

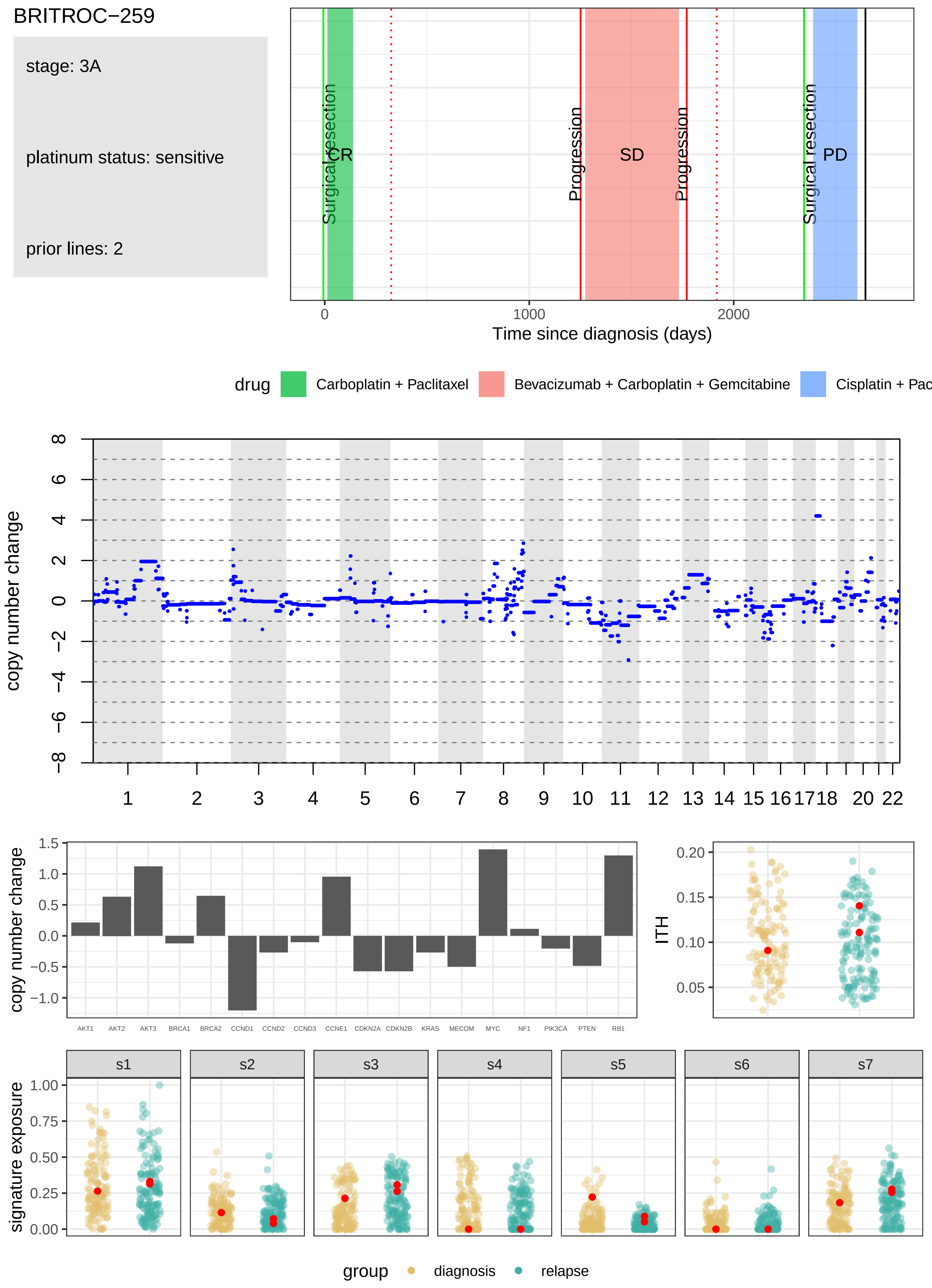
BRITROC−259
stage: 3A
platinum status: sensitive
prior lines: 2
Surgical resection
CR
Progression
SD
Progression
Surgical resection
PD
0
1000
2000
Time since diagnosis (days)
drug
Carboplatin + Paclitaxel
Bevacizumab + Carboplatin + Gemcitabine
Cisplatin + Pac
copy number change
1 2 3 4 5 6 7 8 9 10 11 12 13 14 15 16 17 18 20 22
AKT1 AKT2 AKT3 BRCA1 BRCA2 CCND1 CCND2 CCND3 CCNE1 CDKN2A CDKN2B KRAS MECOM MYC NF1 PIK3CA PTEN RB1
ITH
s1 s2 s3 s4 s5 s6 s7
signature exposure
group
diagnosis
relapse

### BRITROC−260

stage: 2A

platinum status: sensitive

prior lines: 1

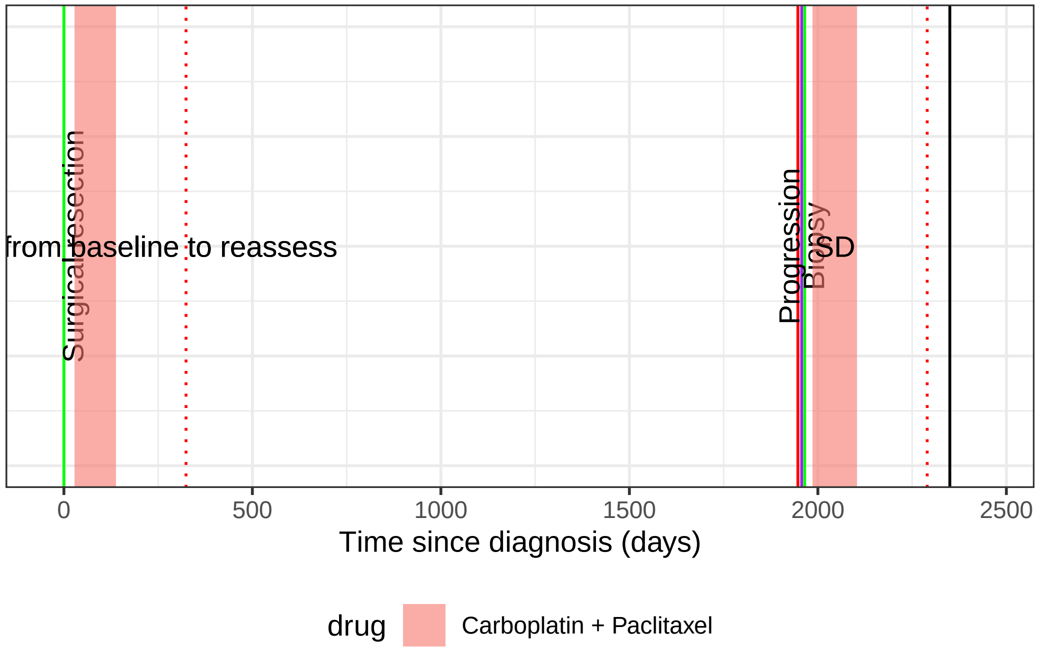

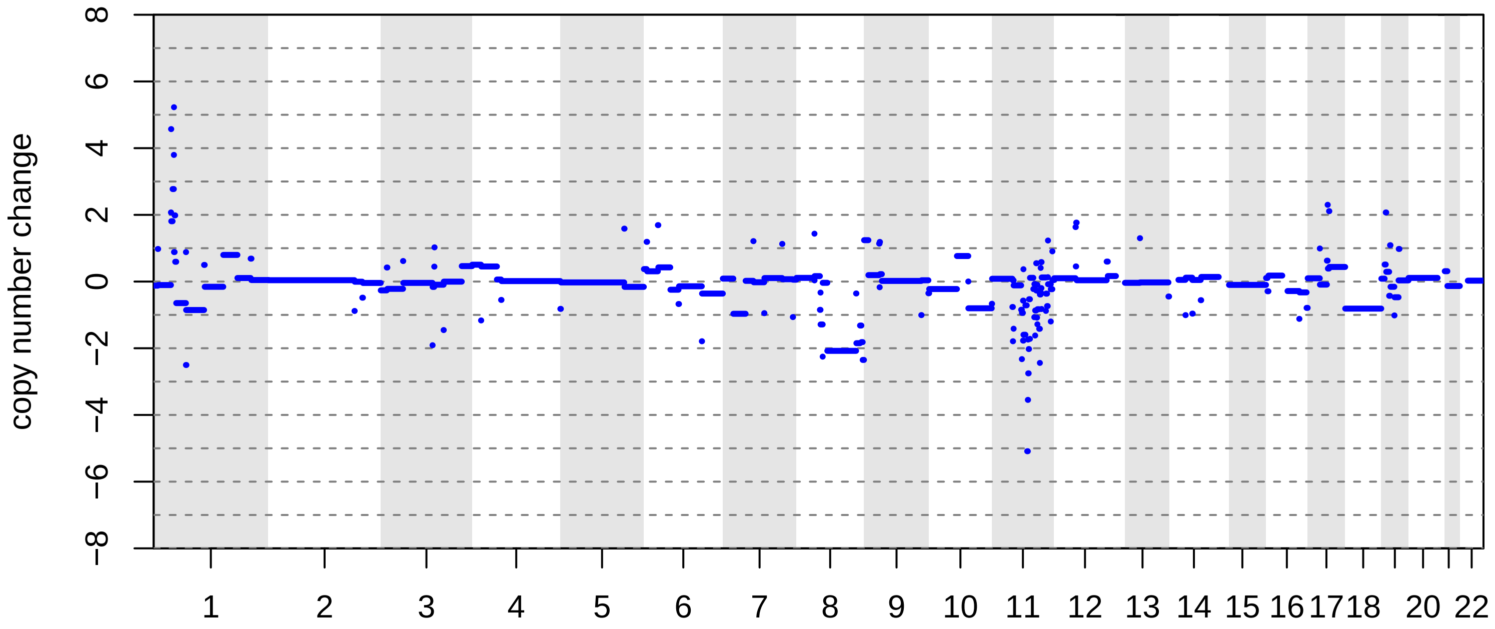

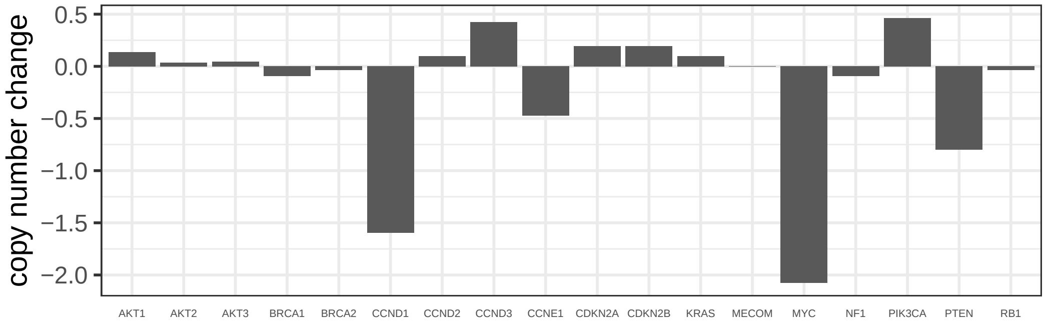

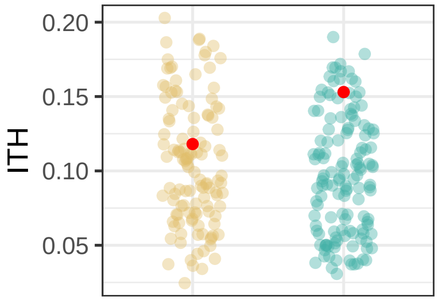

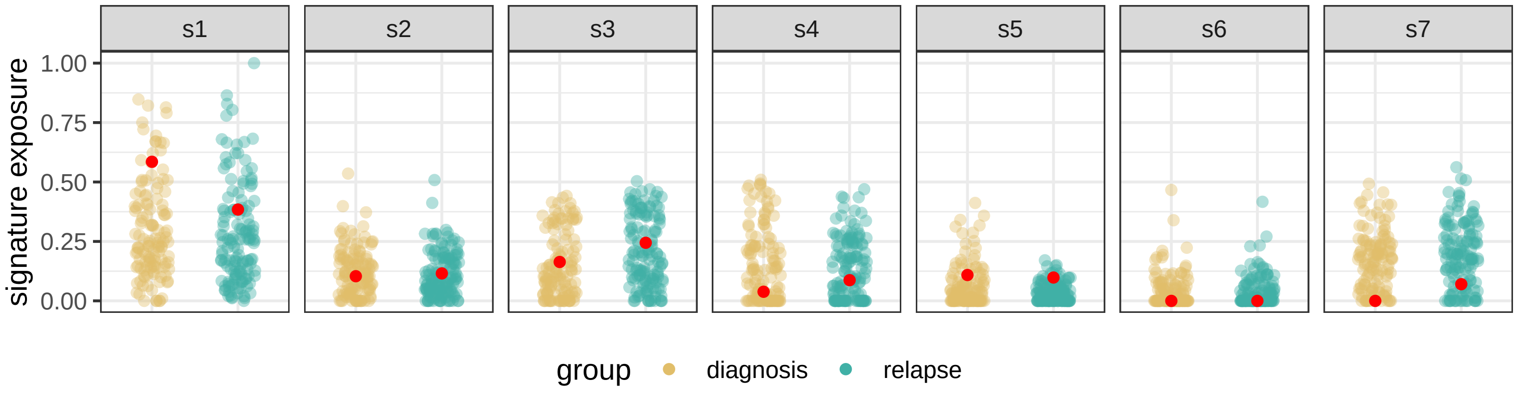

BRITROC−267

stage: 3B

platinum status: sensitive

prior lines: 1

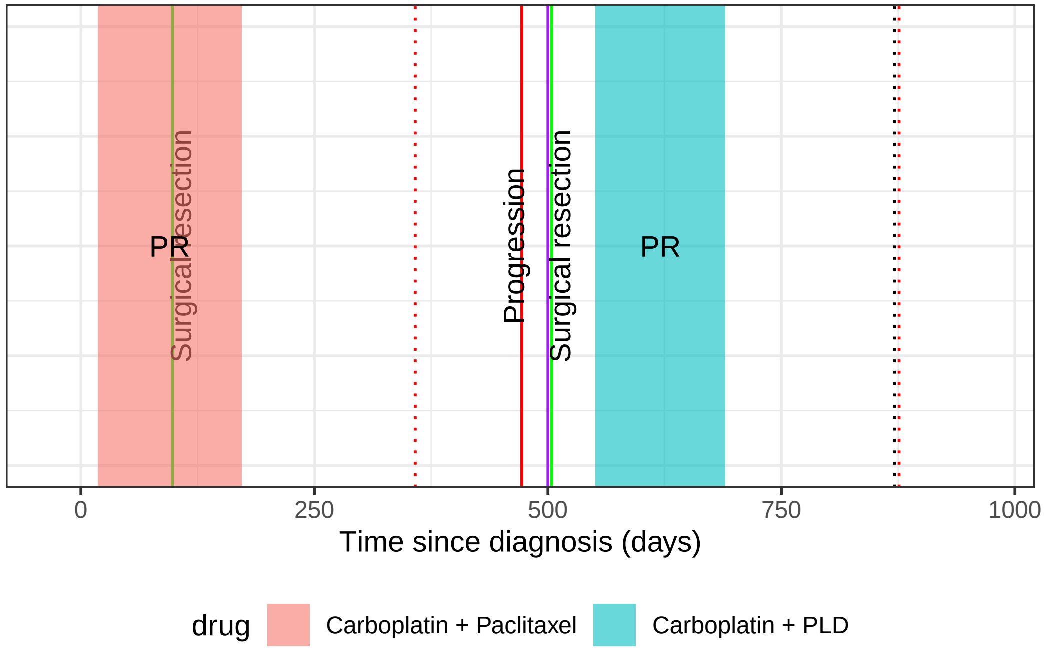

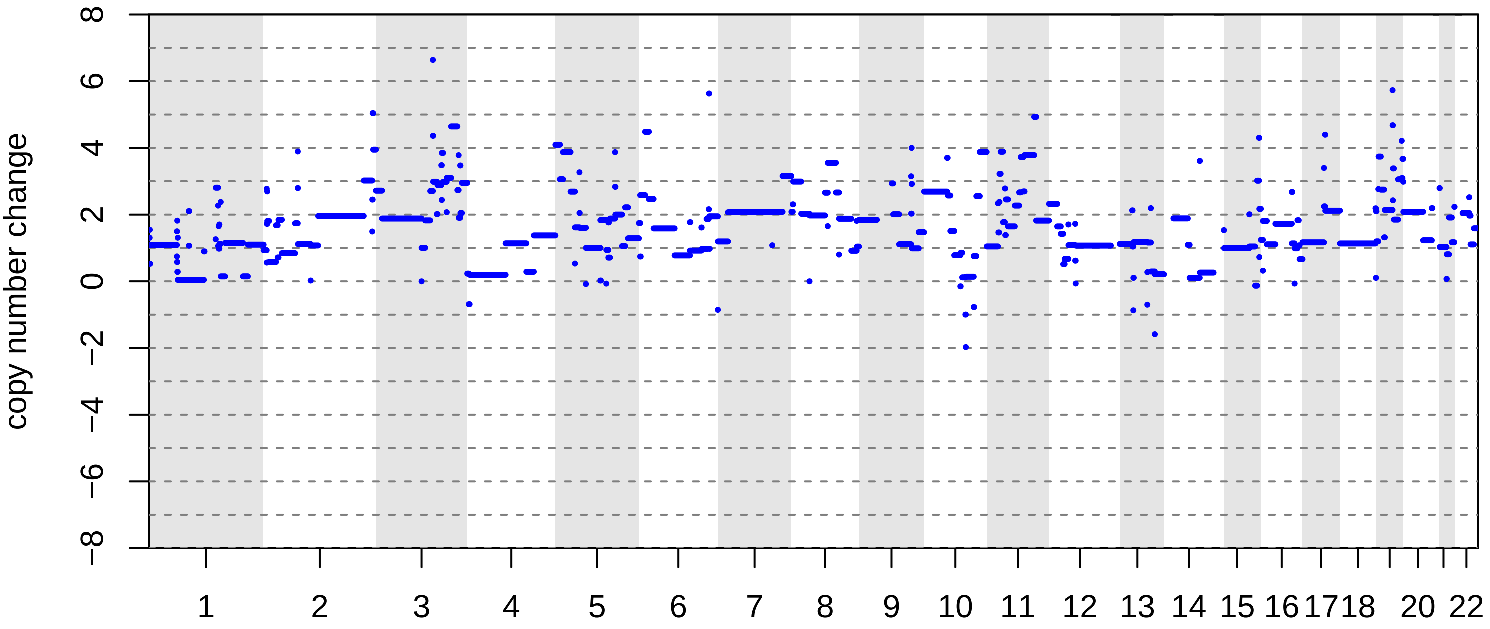

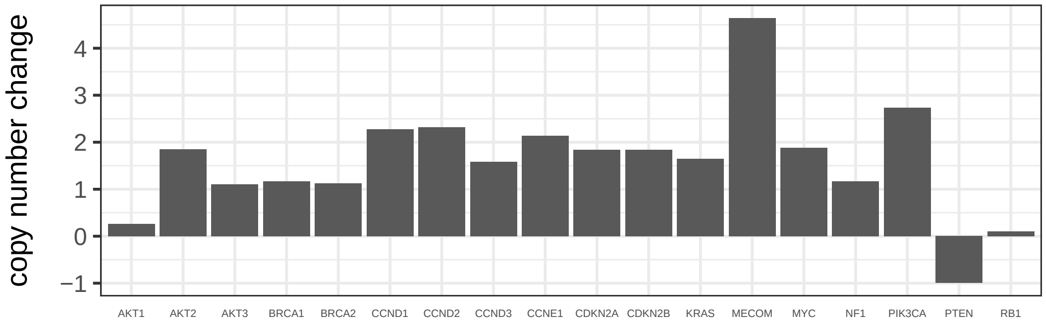

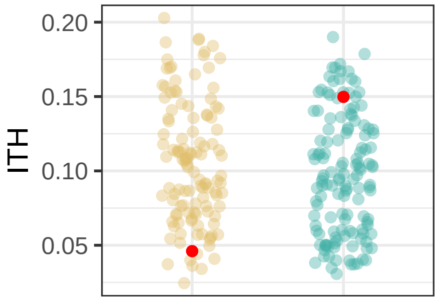

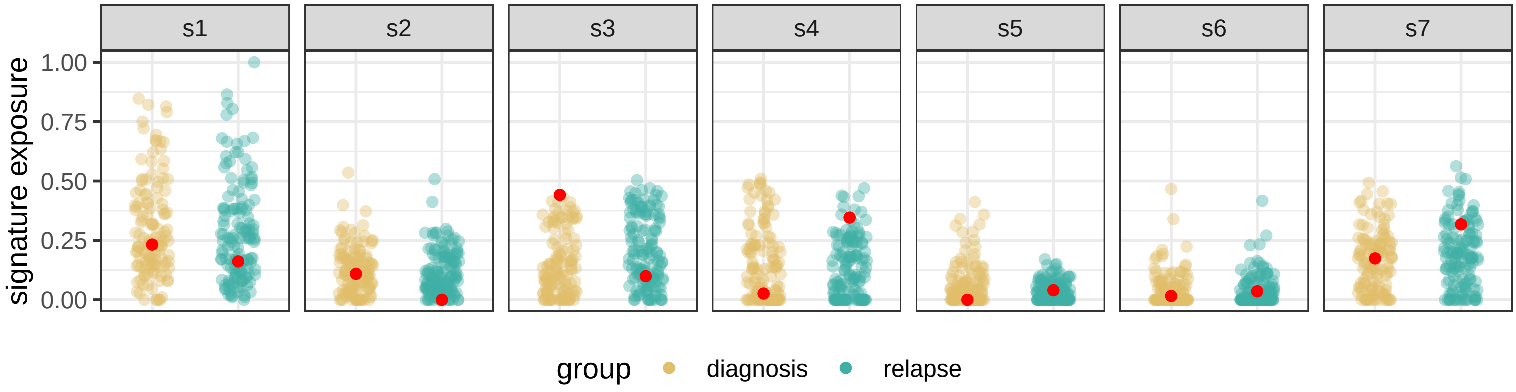

BRITROC−268

stage: 3C

platinum status: resistant

prior lines: 2

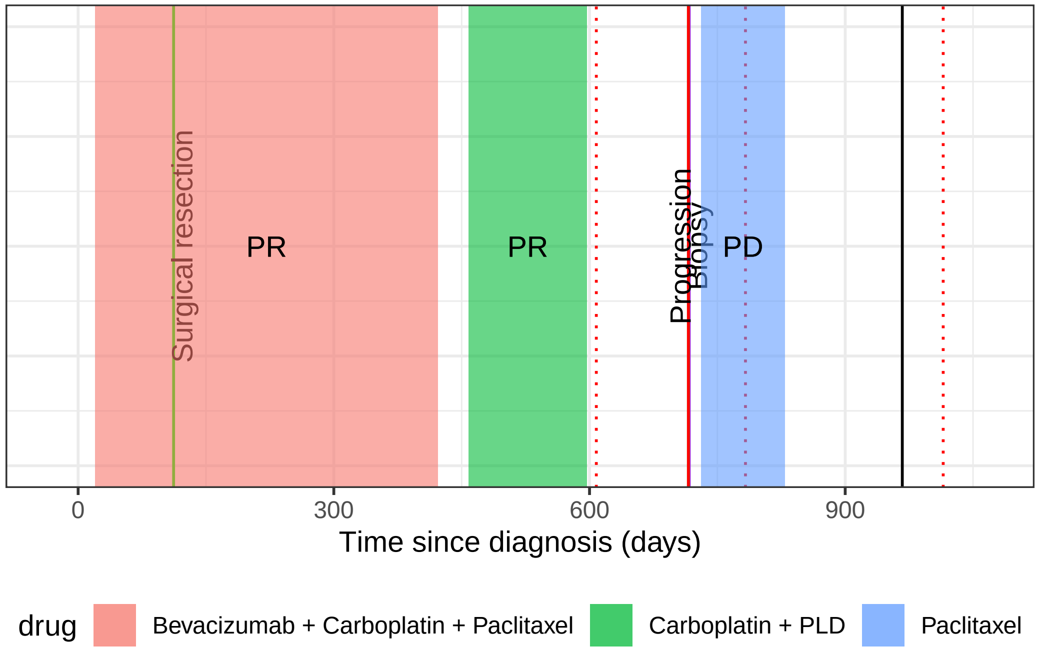

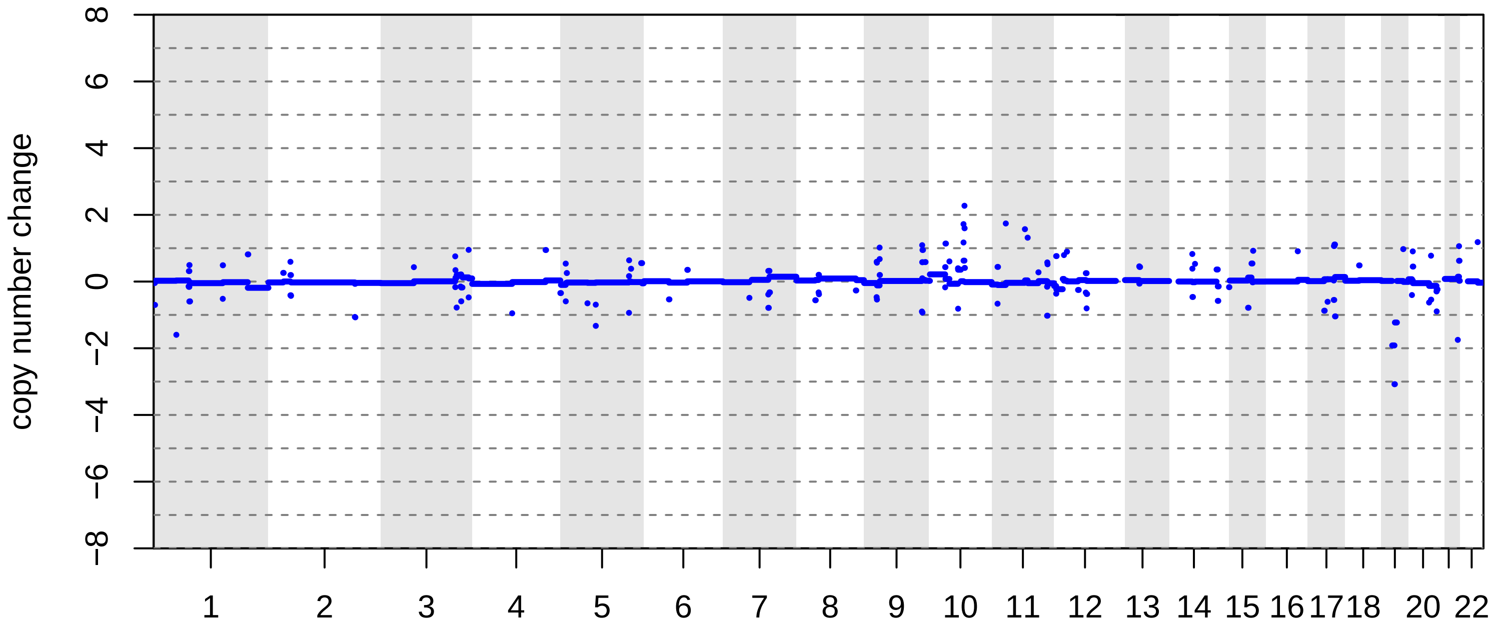

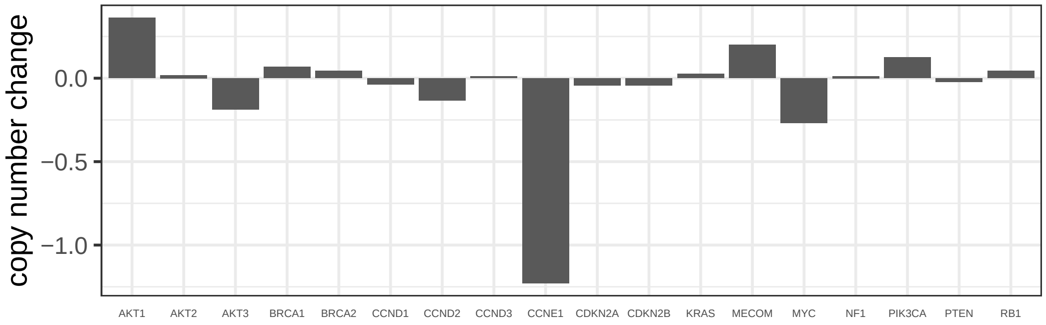

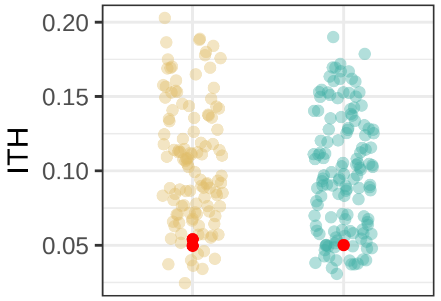

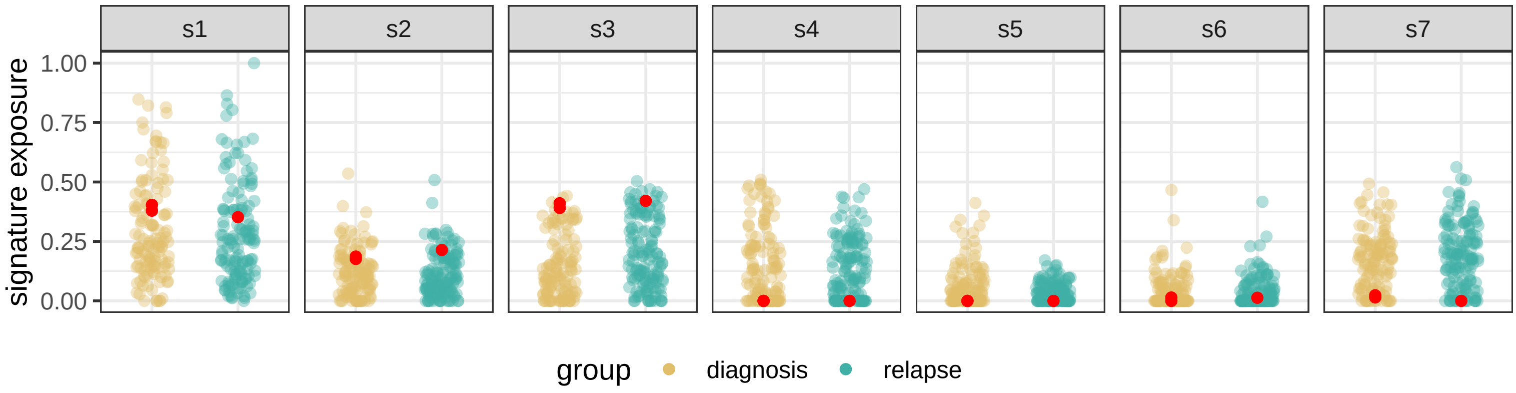

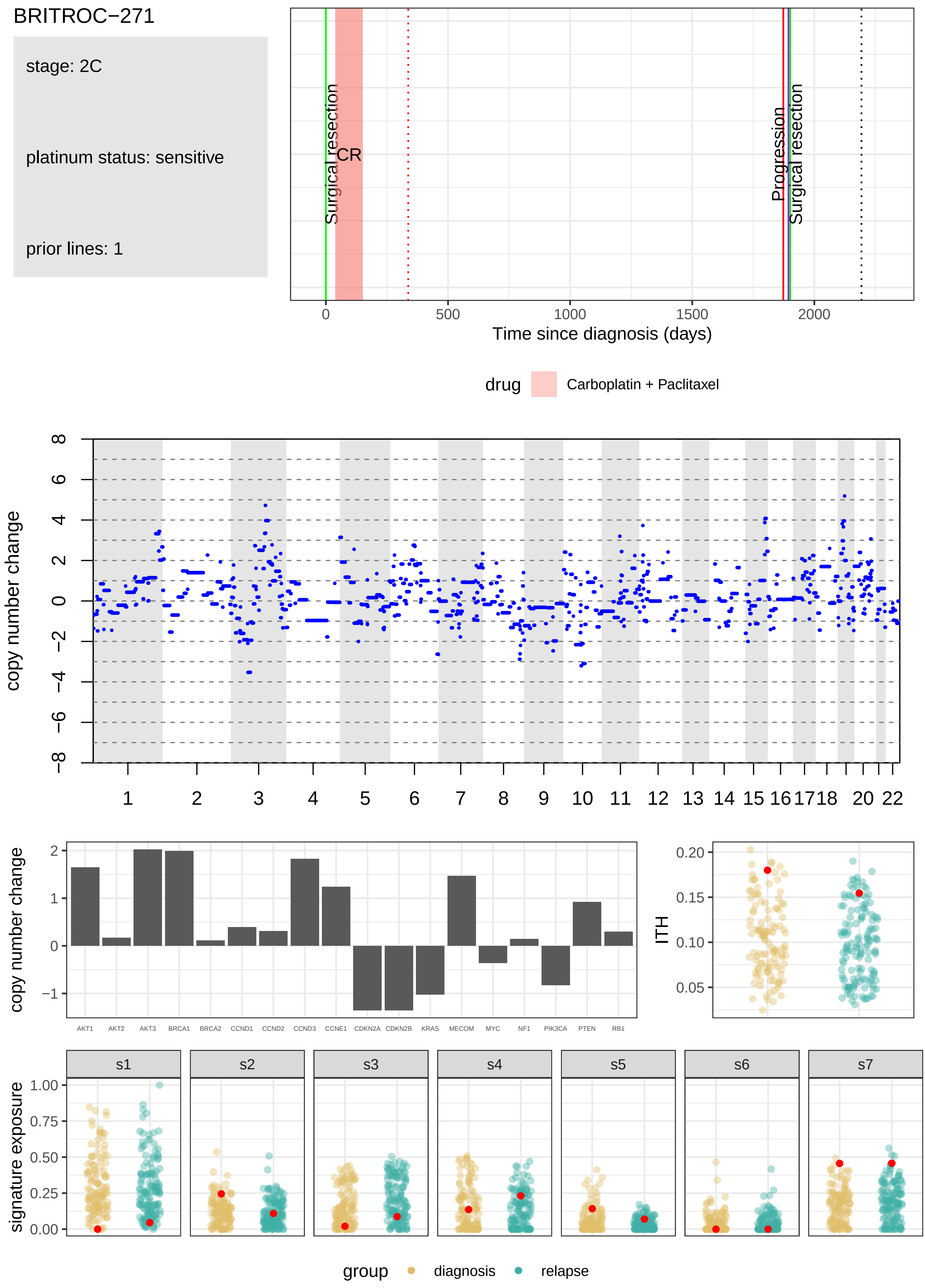

BRITROC−271
stage: 2C
platinum status: sensitive
prior lines: 1
Surgical resection
CR
Progression
Surgical resection
Time since diagnosis (days)
drug
Carboplatin + Paclitaxel
copy number change
copy number change
AKT1
AKT2
AKT3
BRCA1
BRCA2
CCND1
CCND2
CCND3
CCNE1
CDKN2A
CDKN2B
KRAS
MECOM
MYC
NF1
PIK3CA
PTEN
RB1
ITH
signature exposure
s1
s2
s3
s4
s5
s6
s7
group
diagnosis
relapse

BRITROC−274

stage: 2B

platinum status: sensitive

prior lines: 2

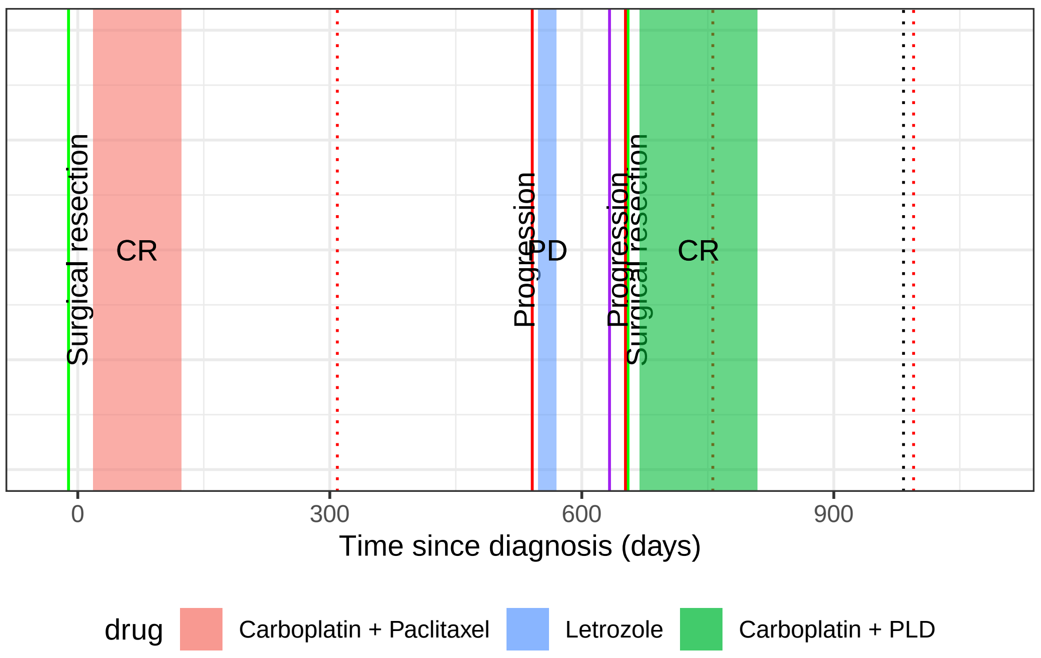

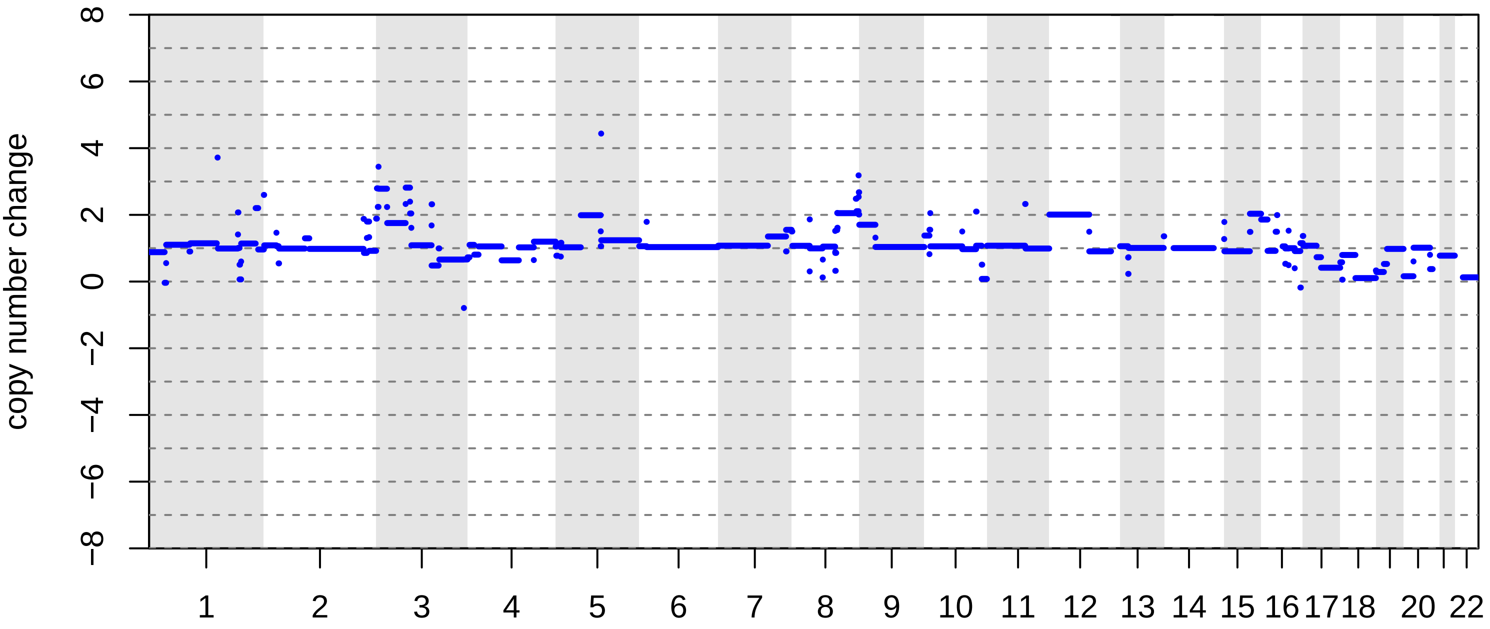

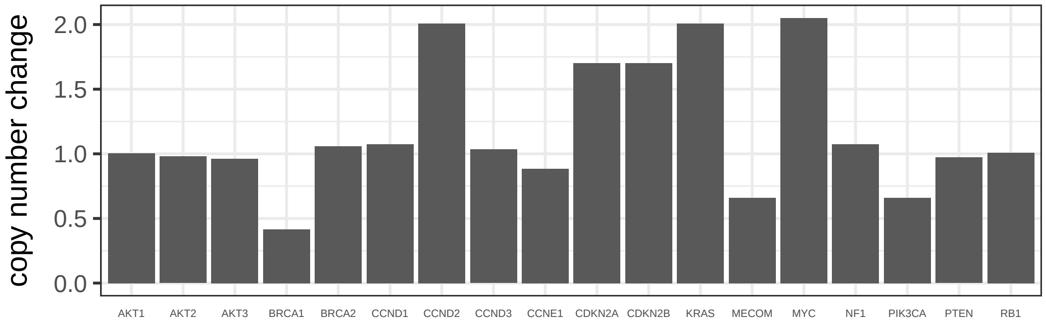

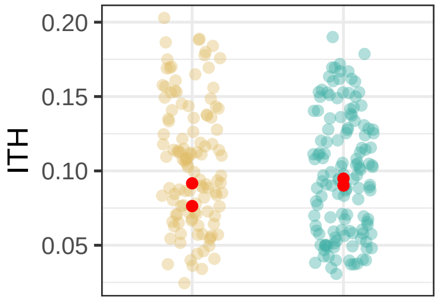

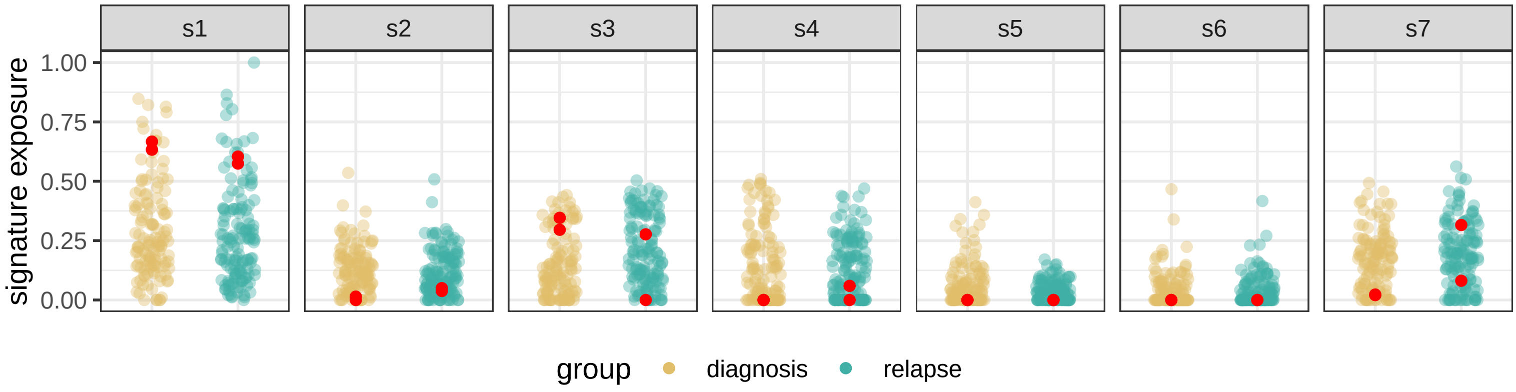

### BRITROC−32

stage: 3C

platinum status: resistant

prior lines: 1

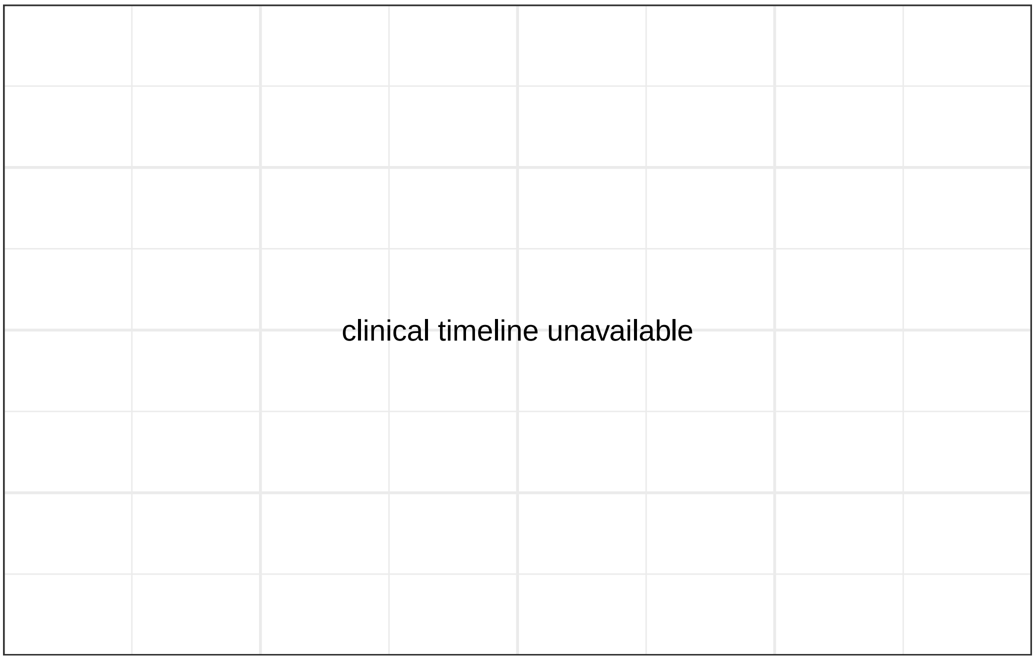

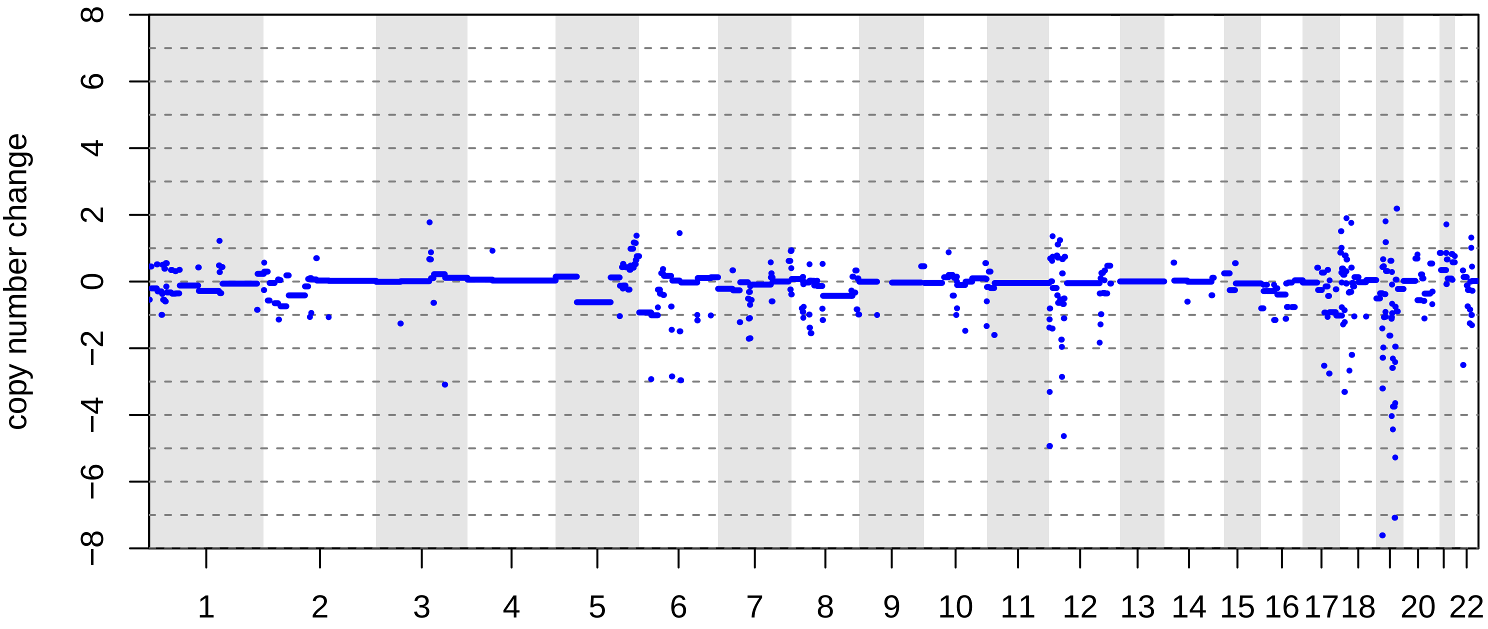

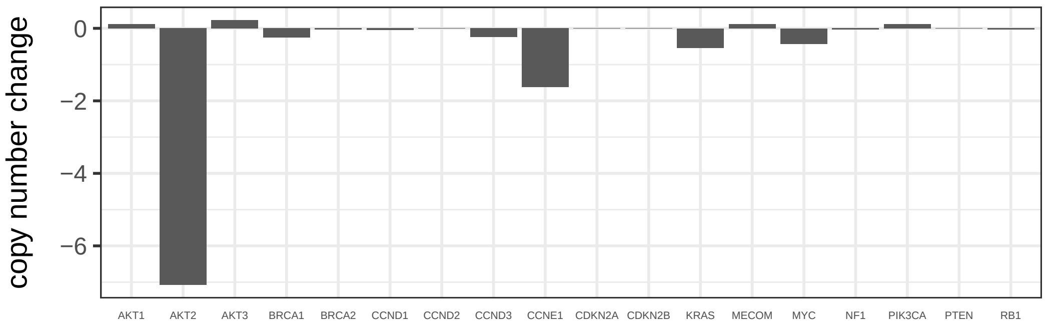

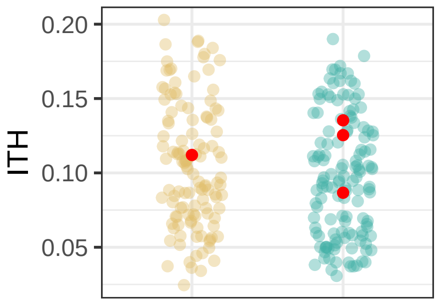

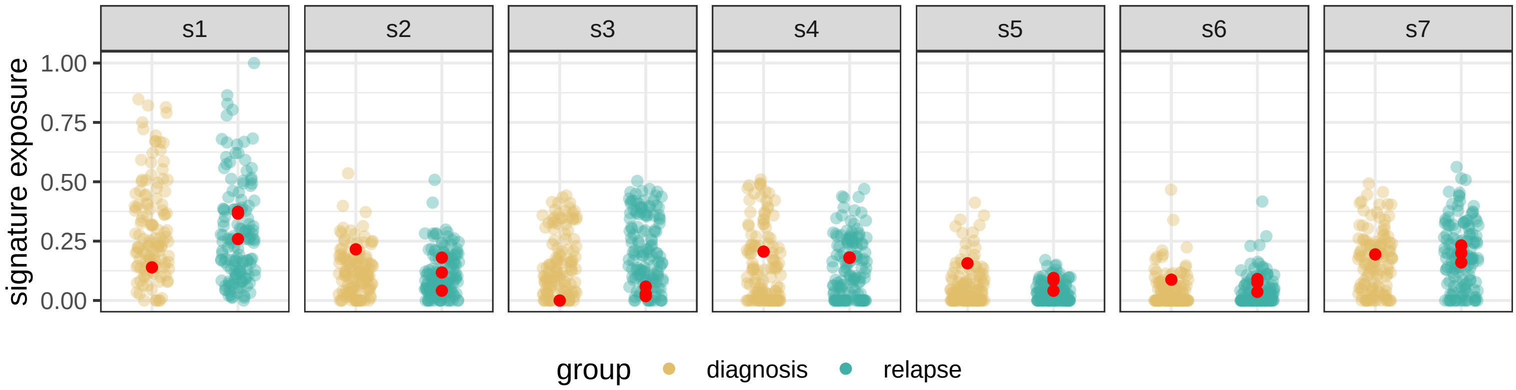

BRITROC−34

stage: 3C

platinum status: sensitive

prior lines: 1

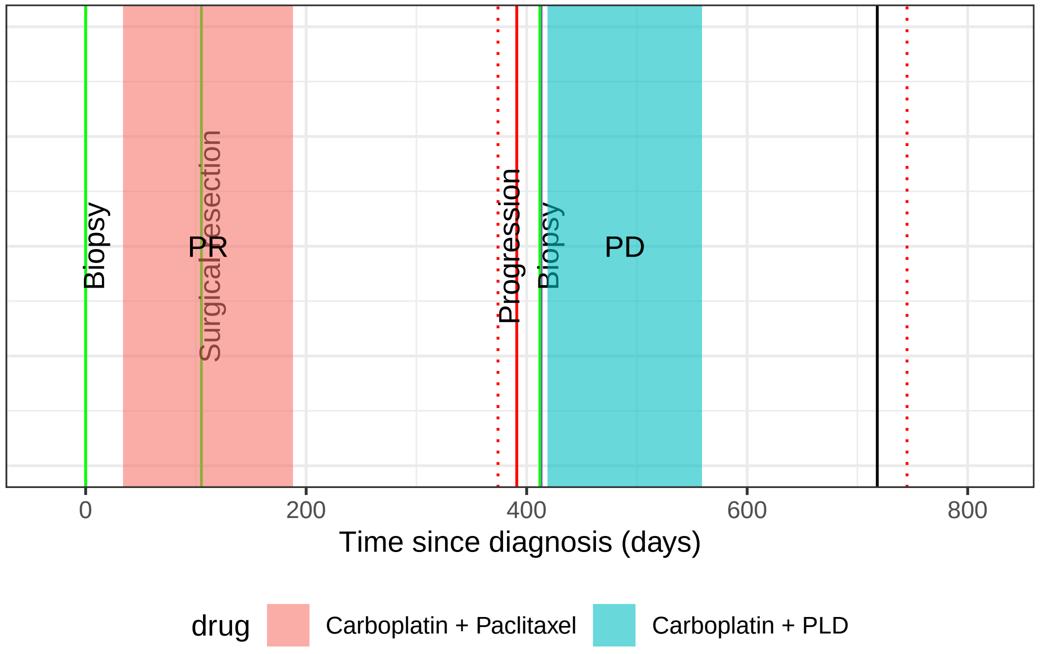

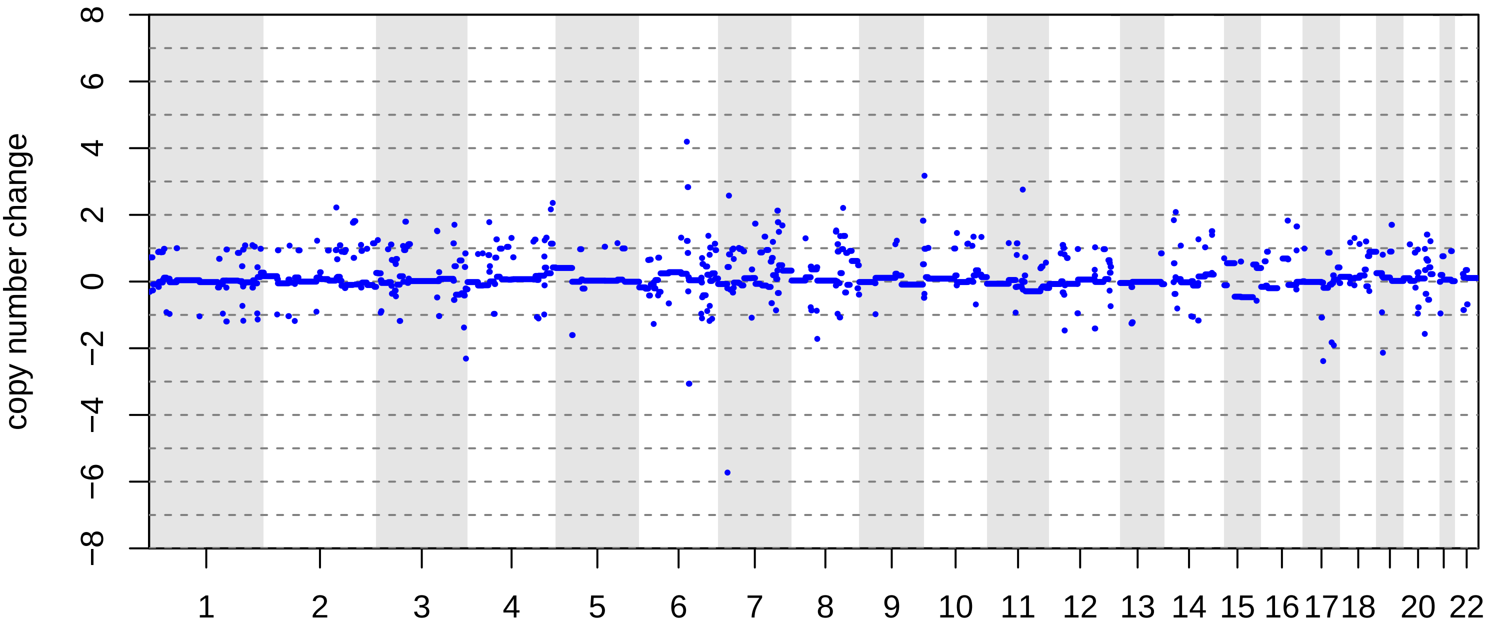

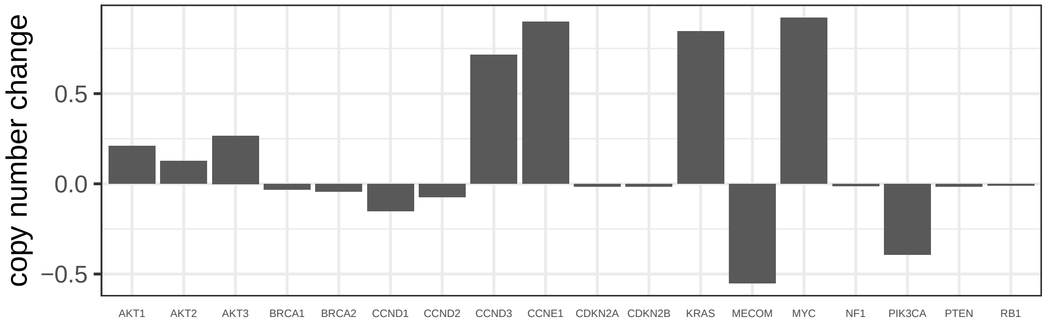

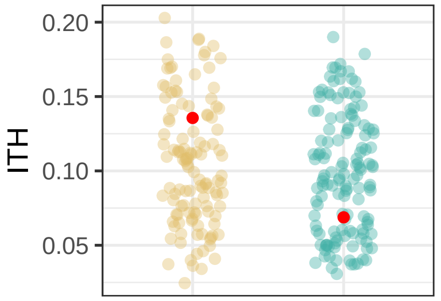

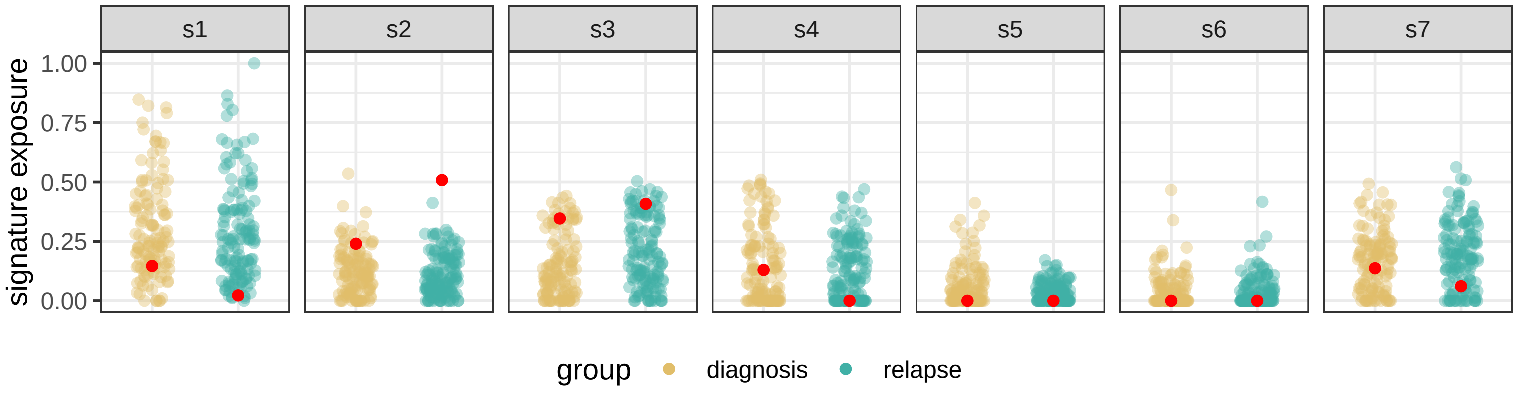

### BRITROC−36

stage: 3B

platinum status: sensitive

prior lines: 1

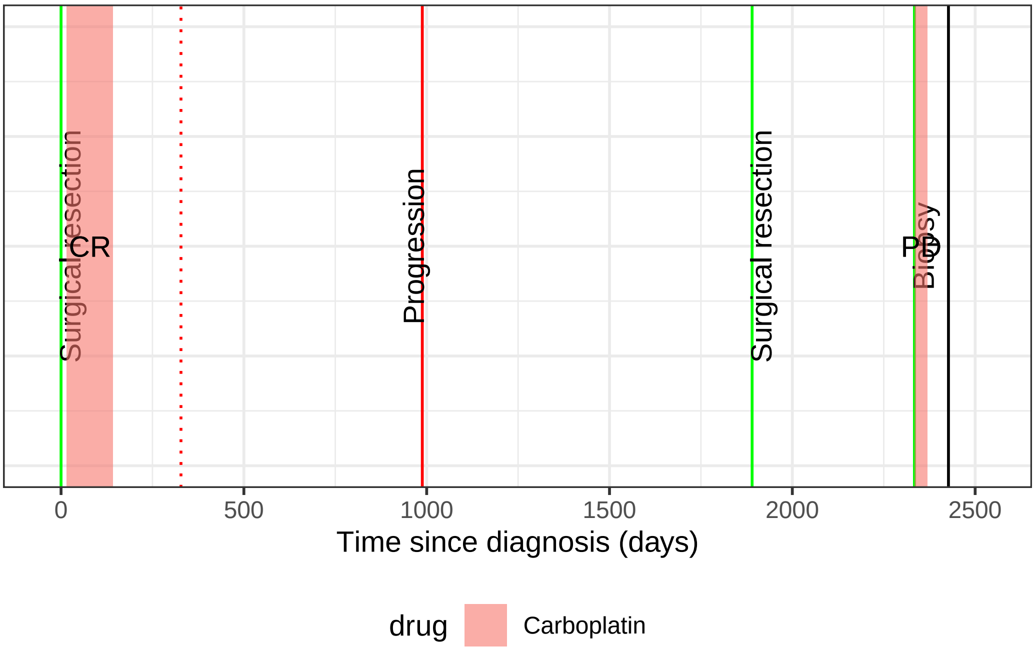

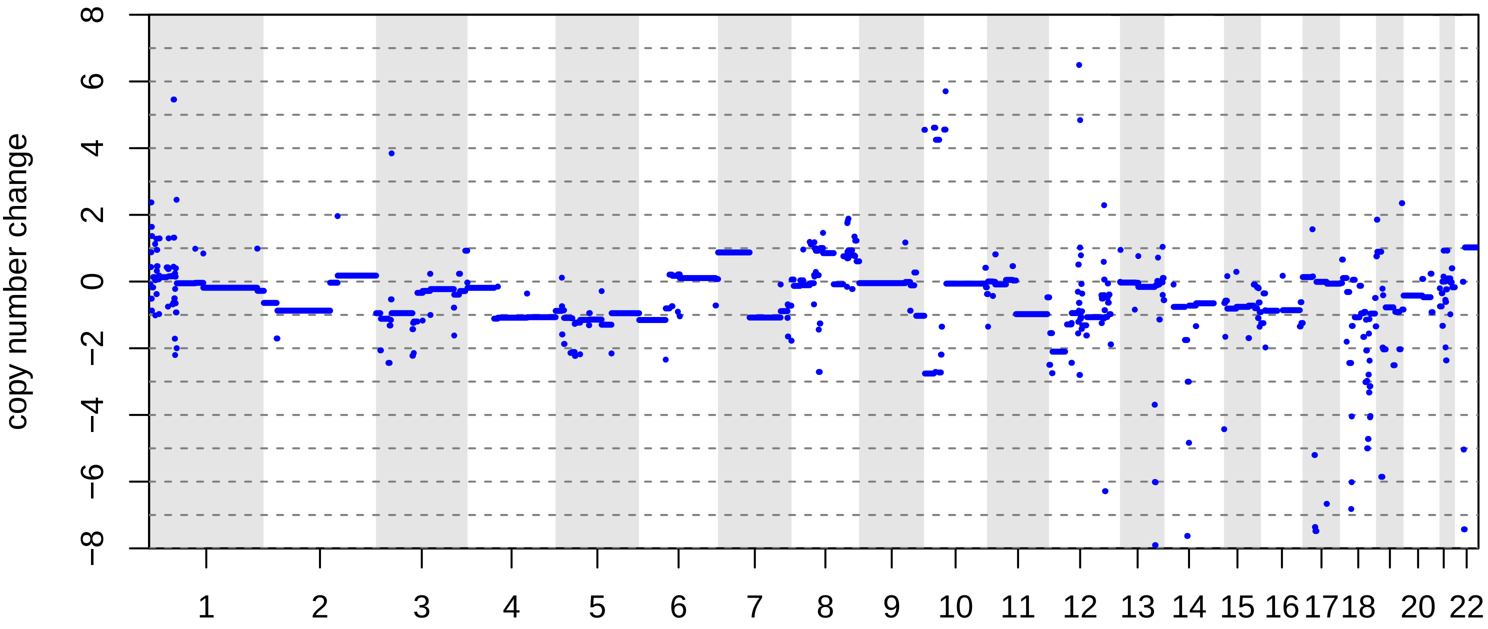

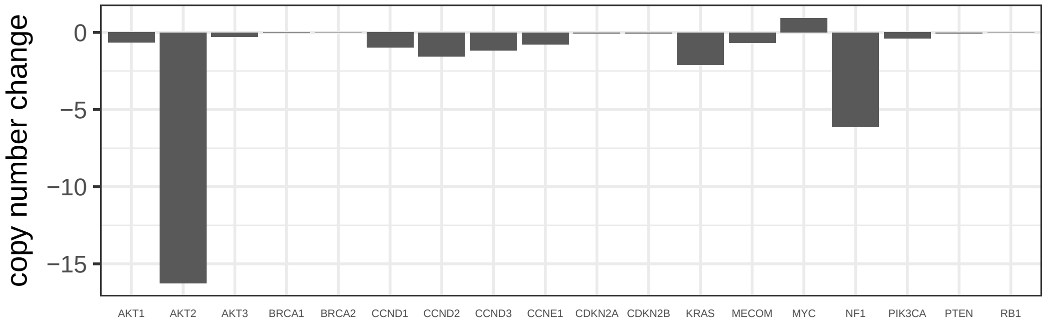

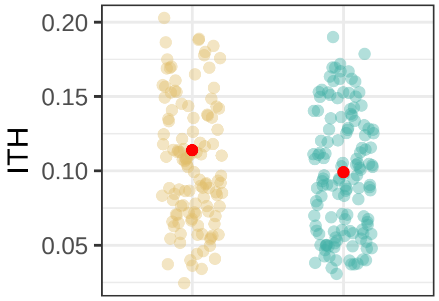

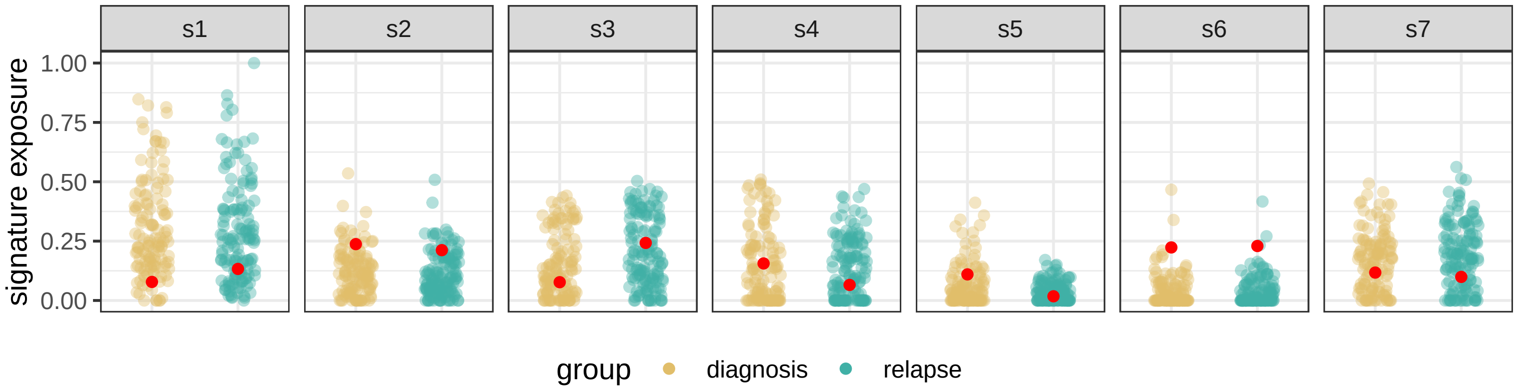

BRITROC−37

stage: 3C

platinum status: sensitive

prior lines: 1

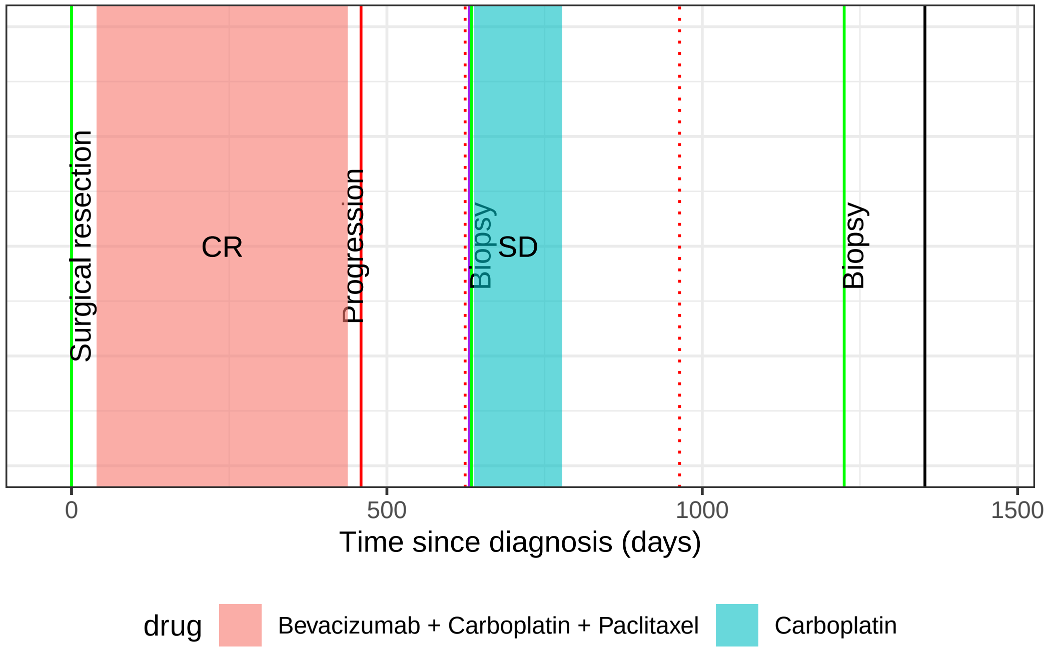

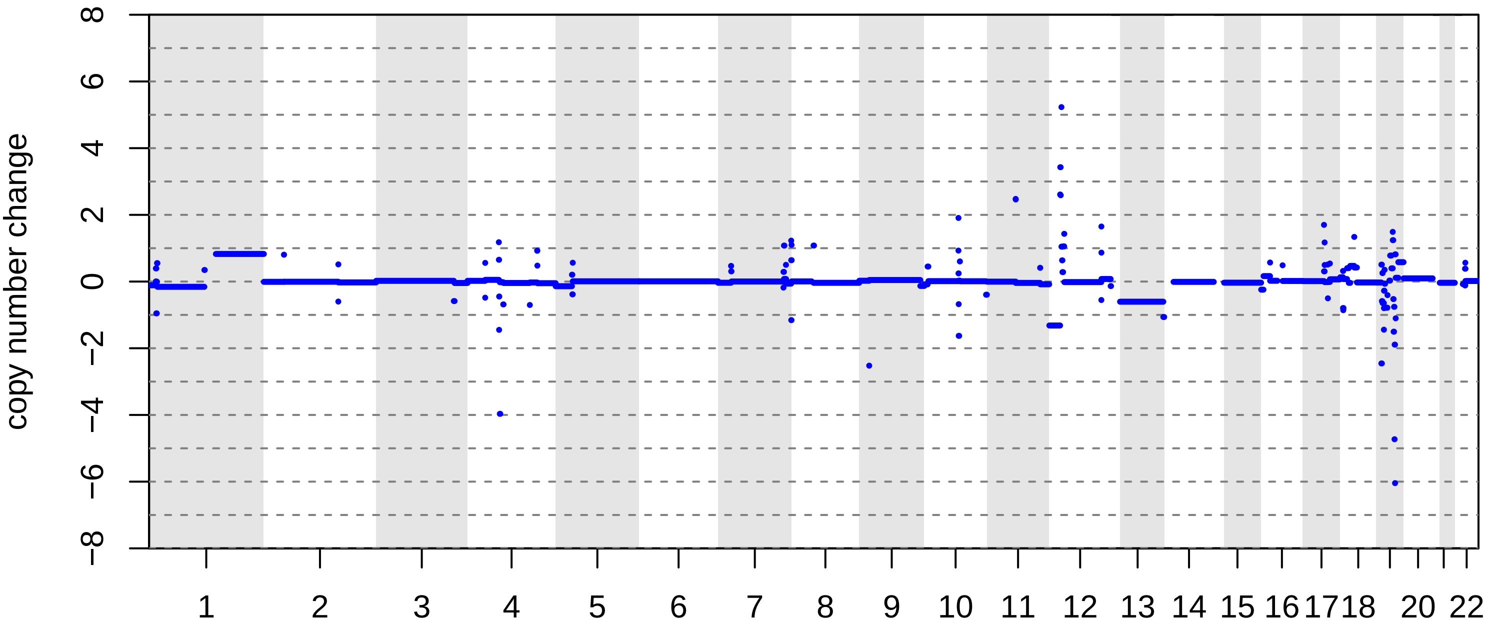

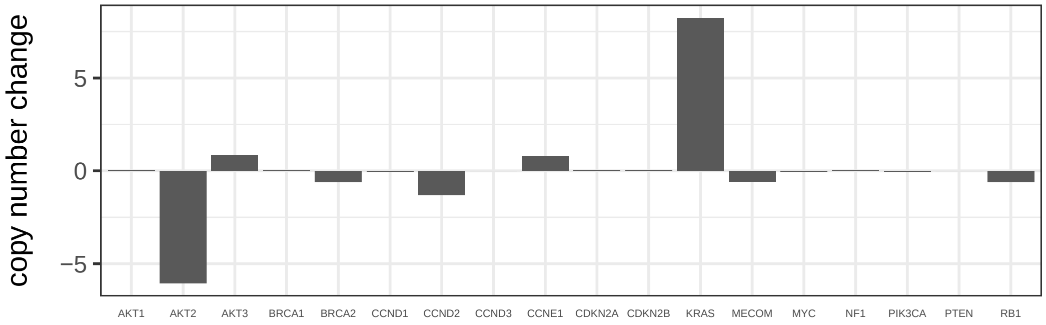

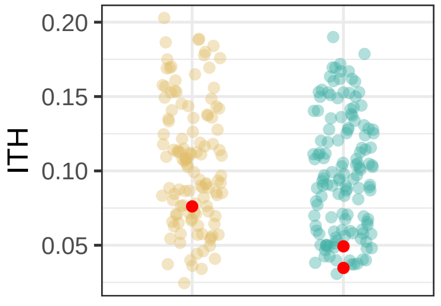

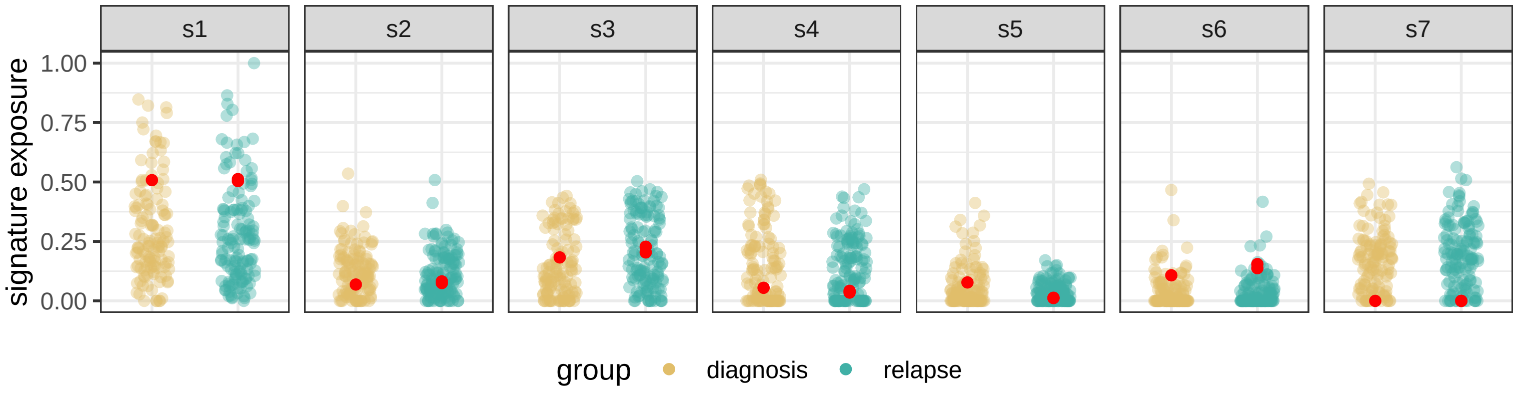

BRITROC−39

stage: 3A

platinum status: sensitive

prior lines: 1

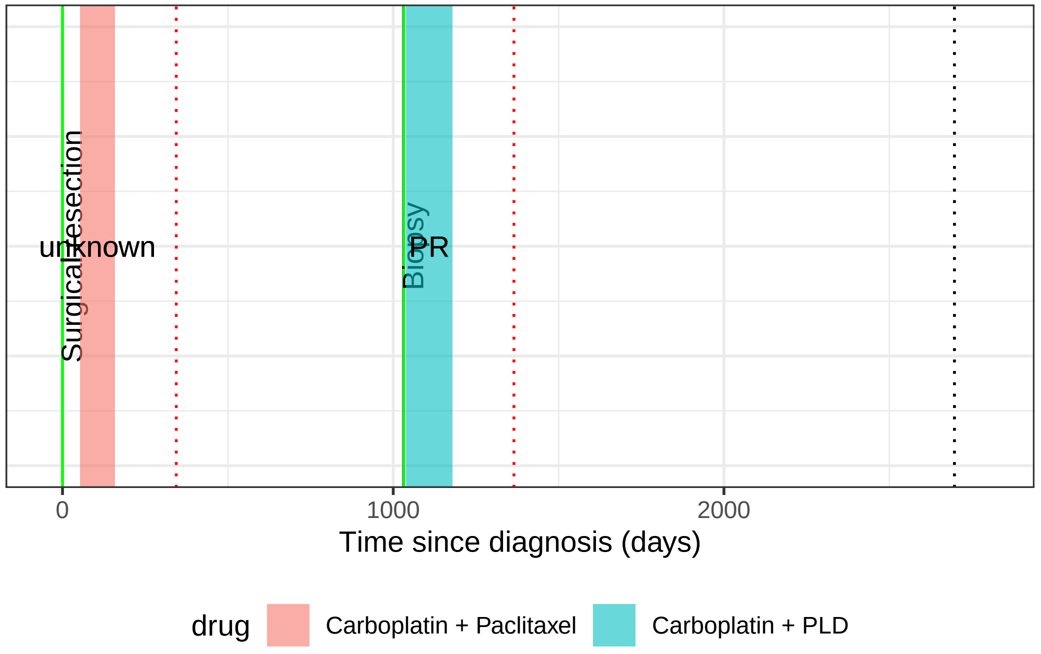

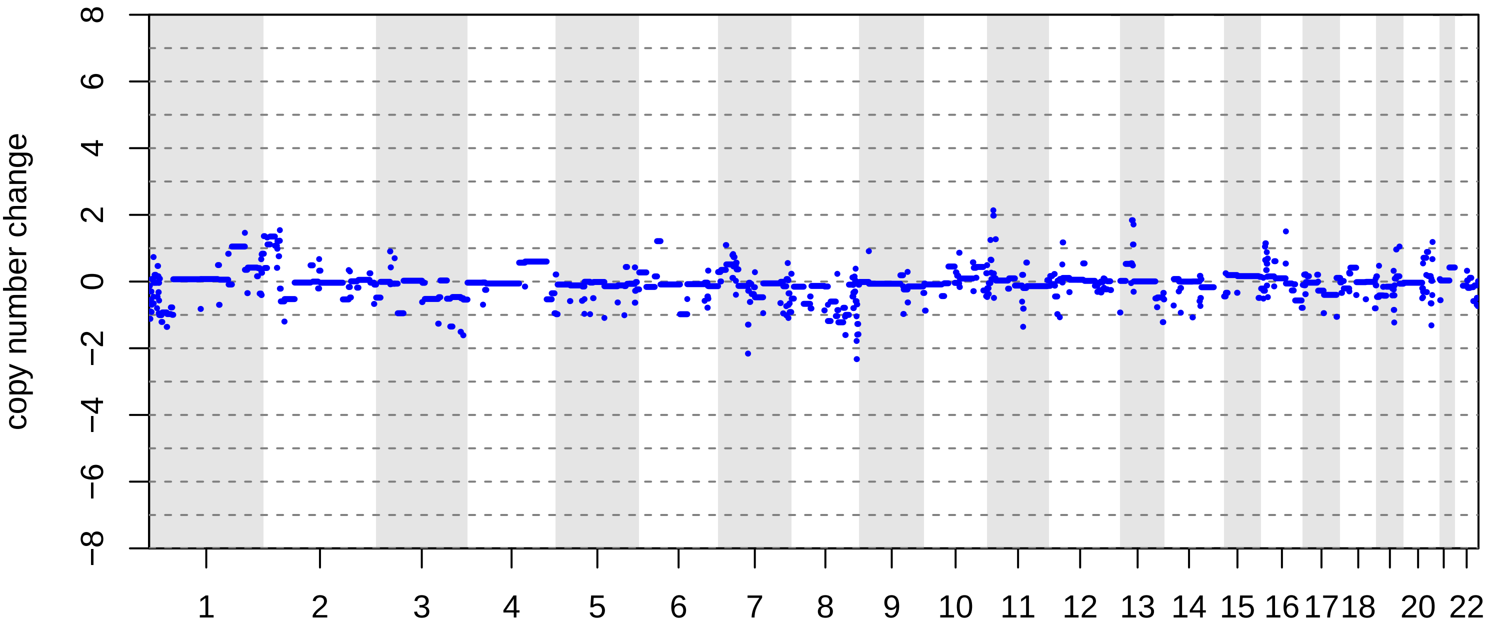

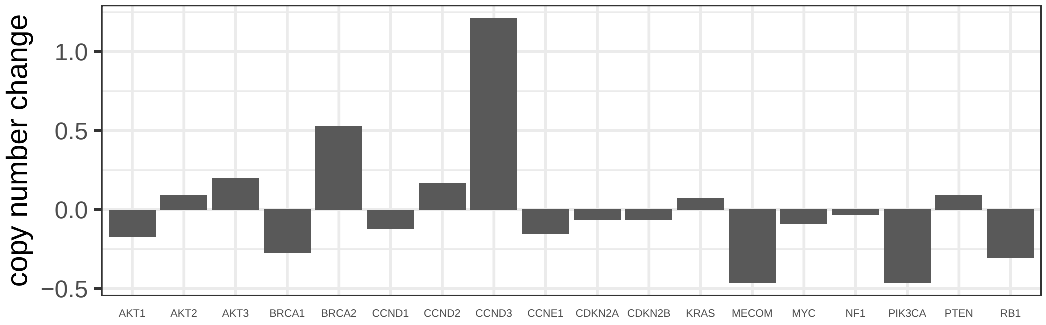

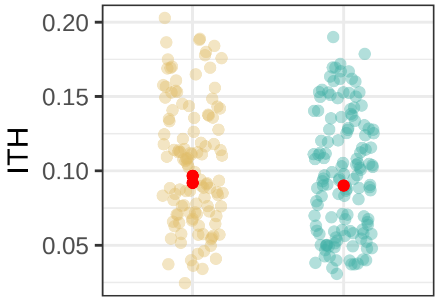

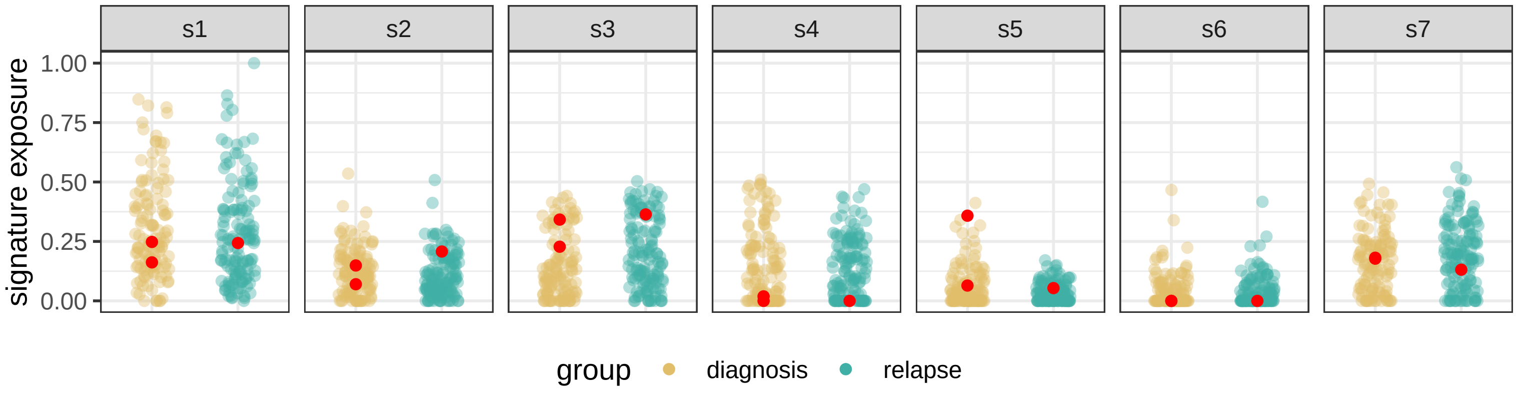

BRITROC−5

stage: 3C

platinum status: sensitive

prior lines: 2

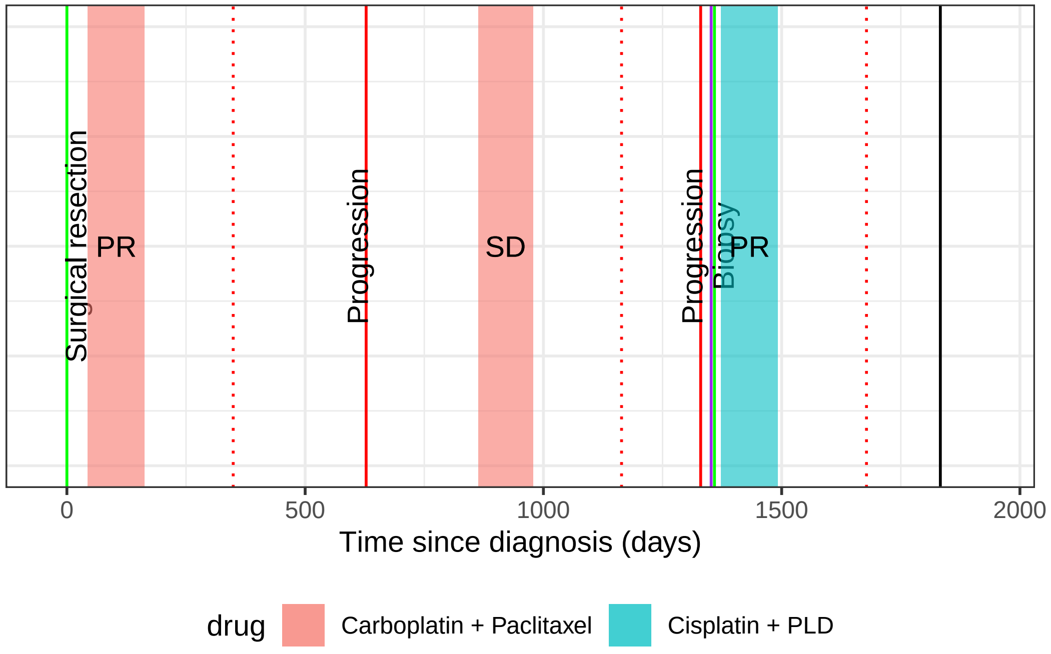

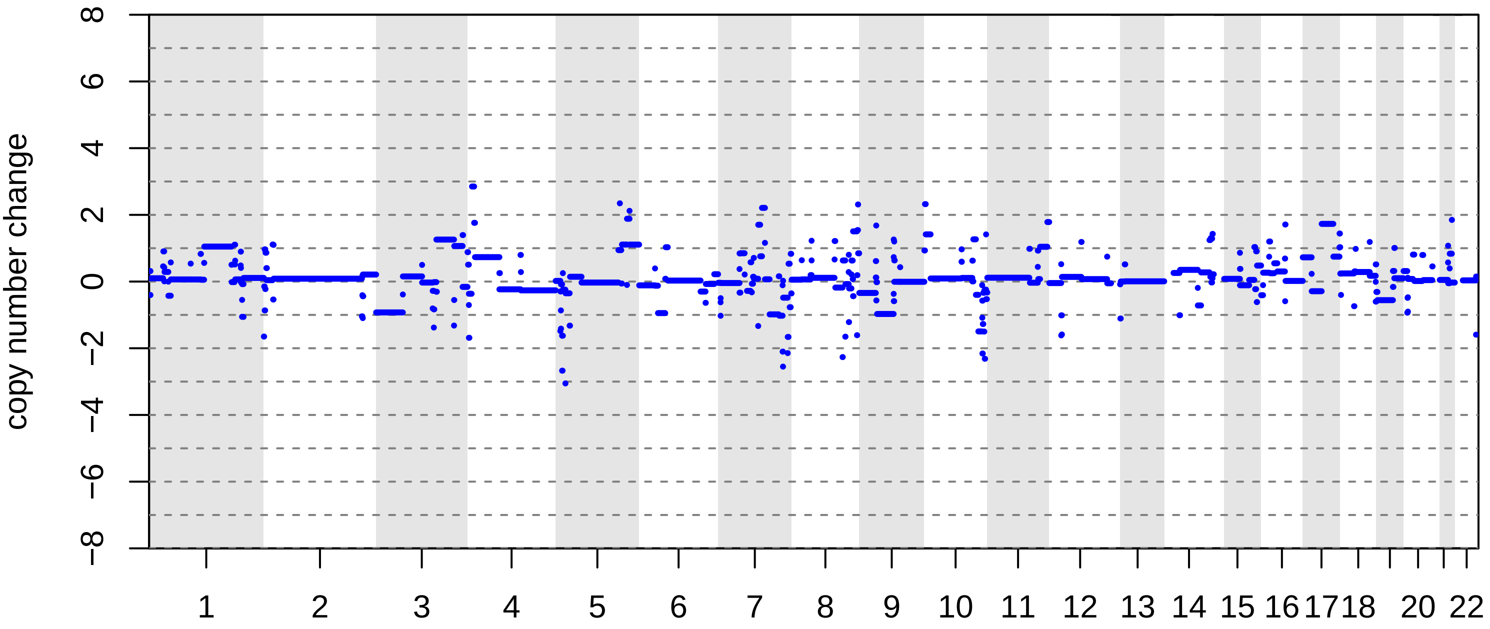

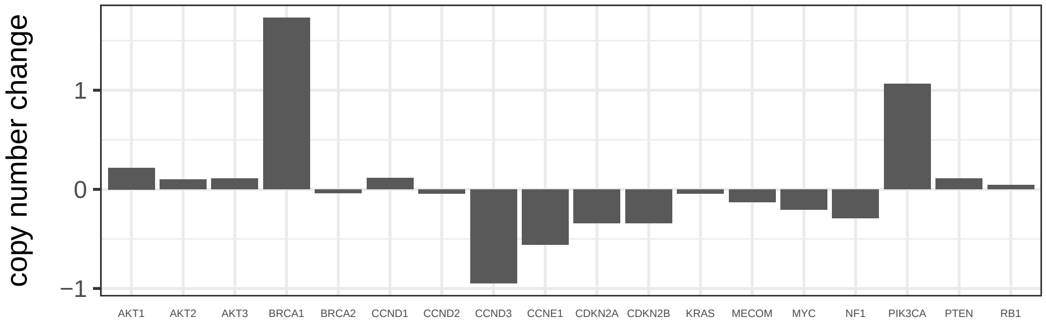

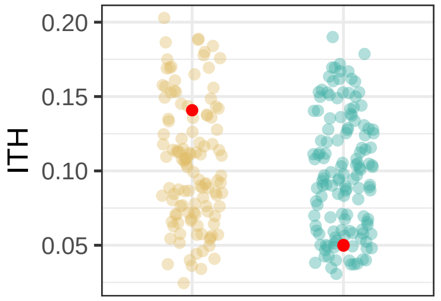

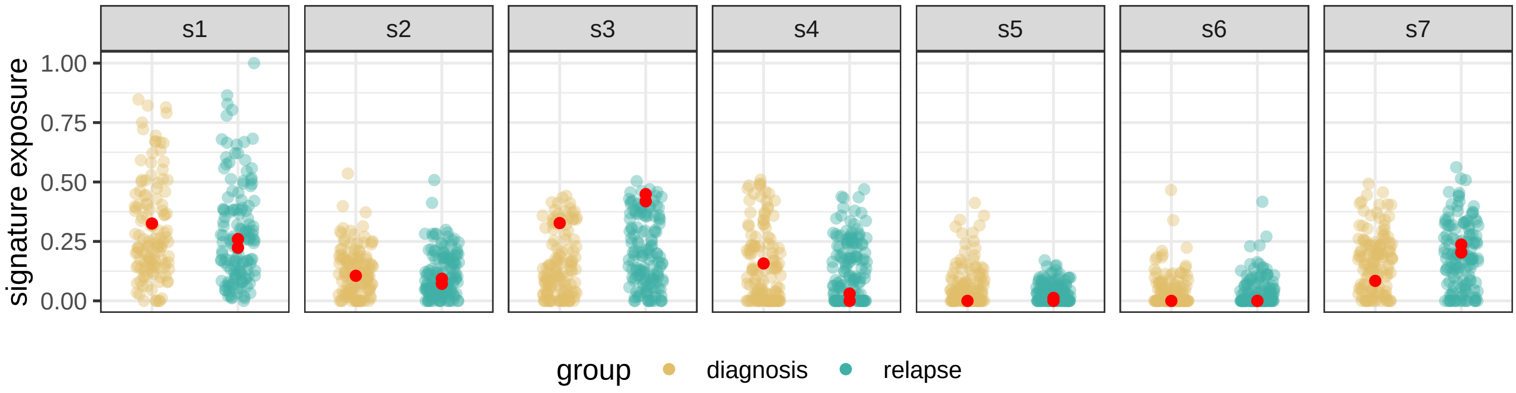

BRITROC−55

stage: 1C

platinum status: sensitive

prior lines: 2

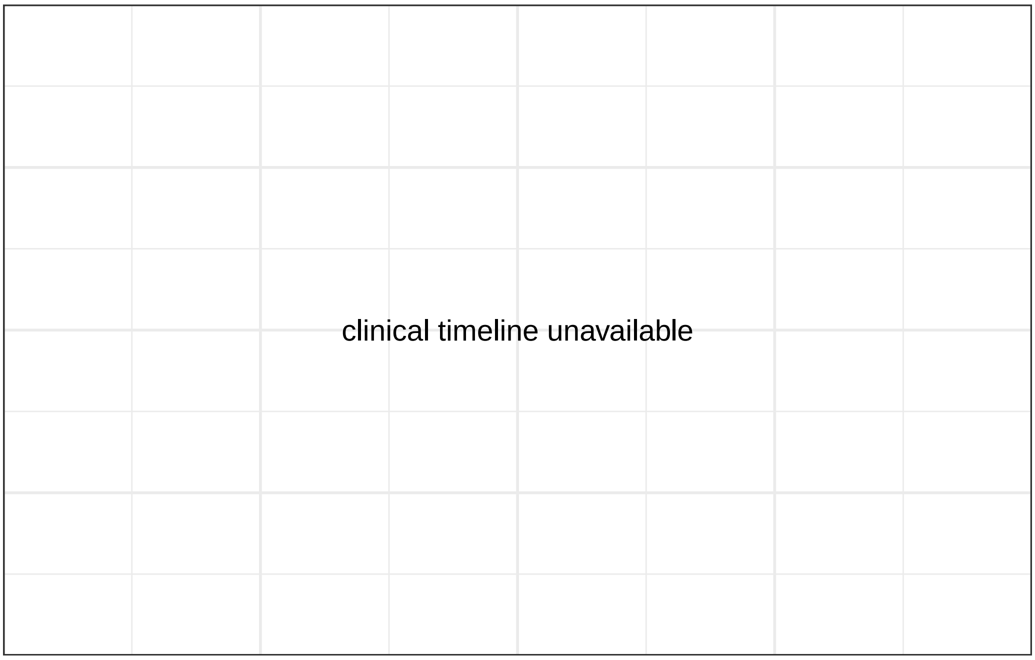

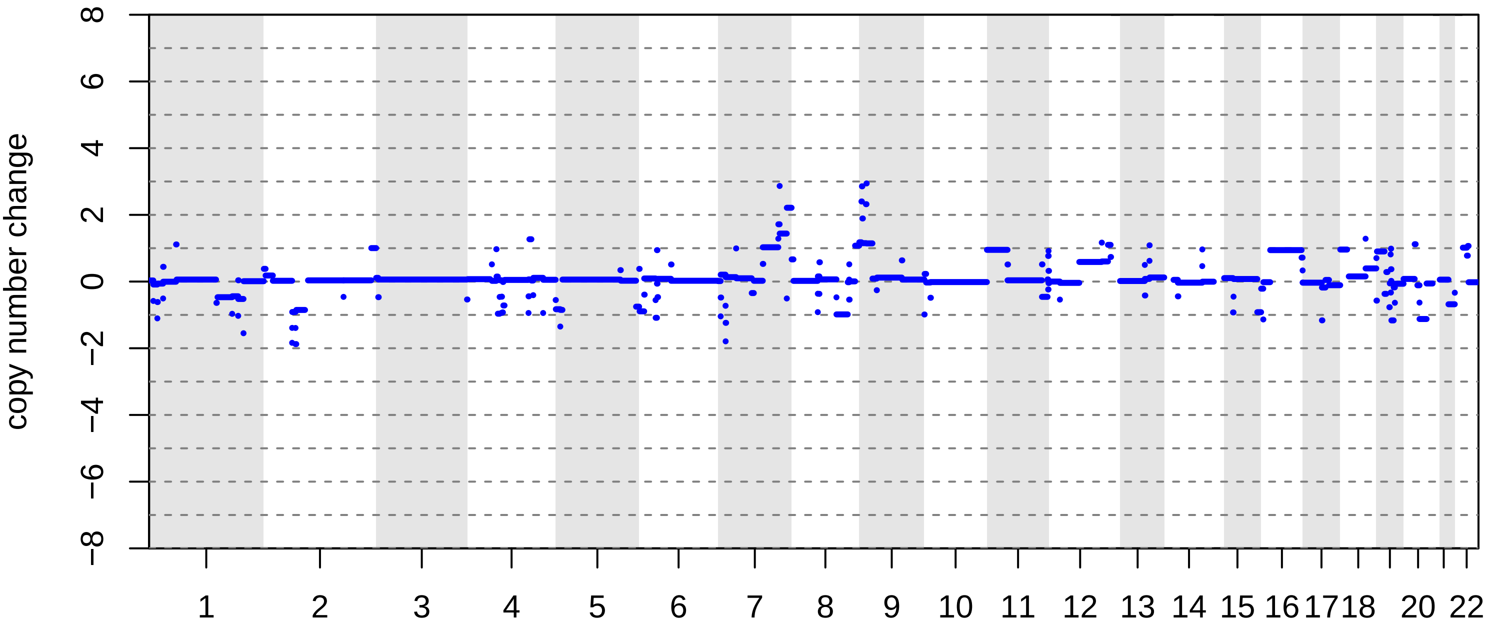

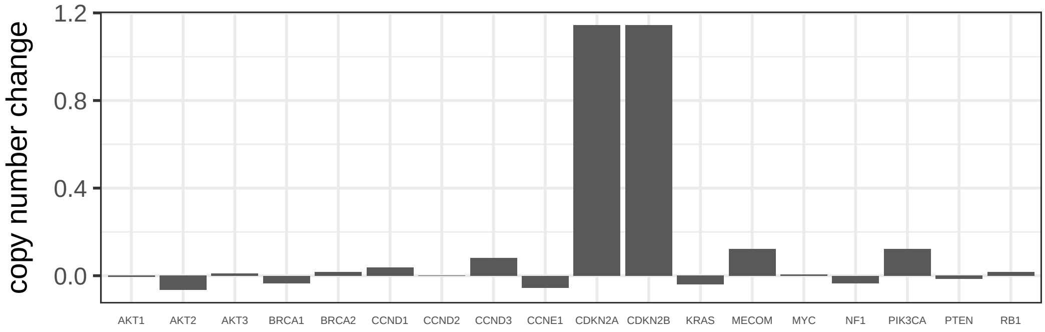

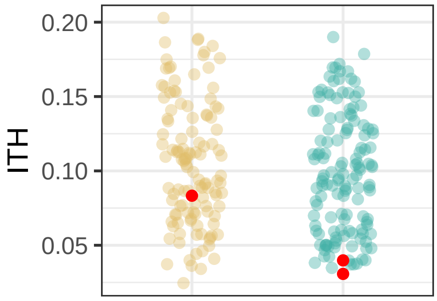

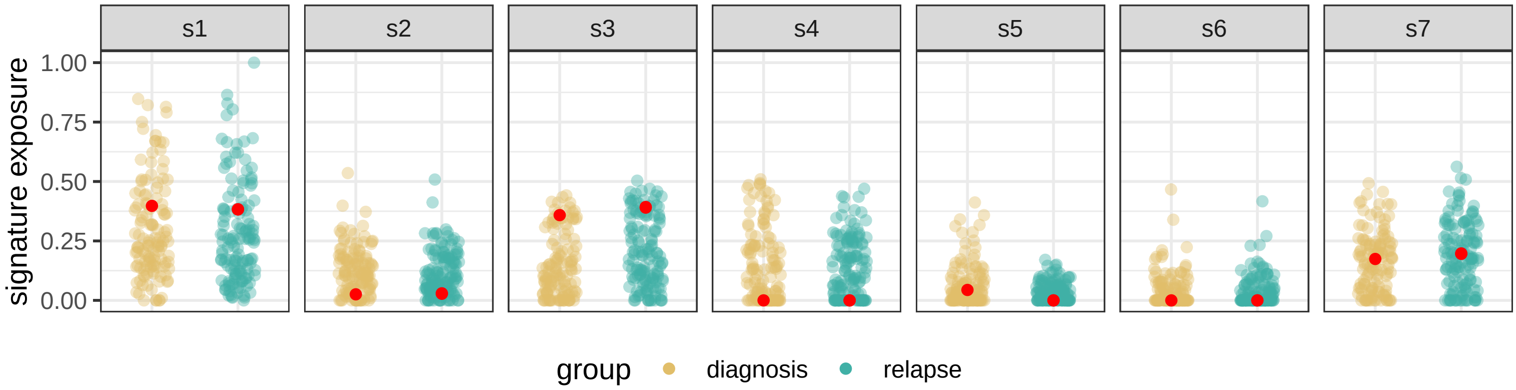

BRITROC−65

stage: 3C

platinum status: sensitive

prior lines: 1

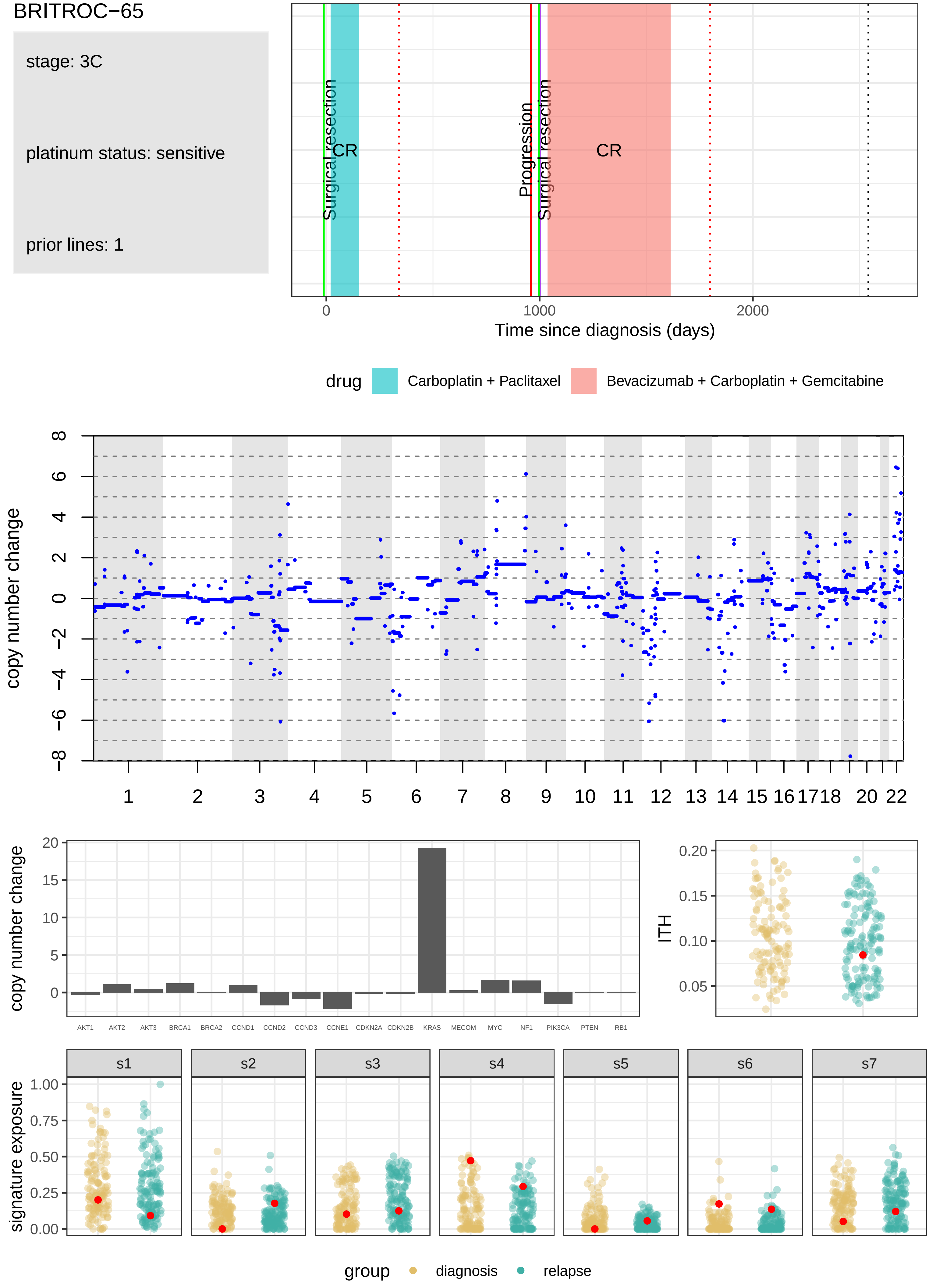

BRITROC−67

stage: 3C

platinum status: sensitive

prior lines: 1

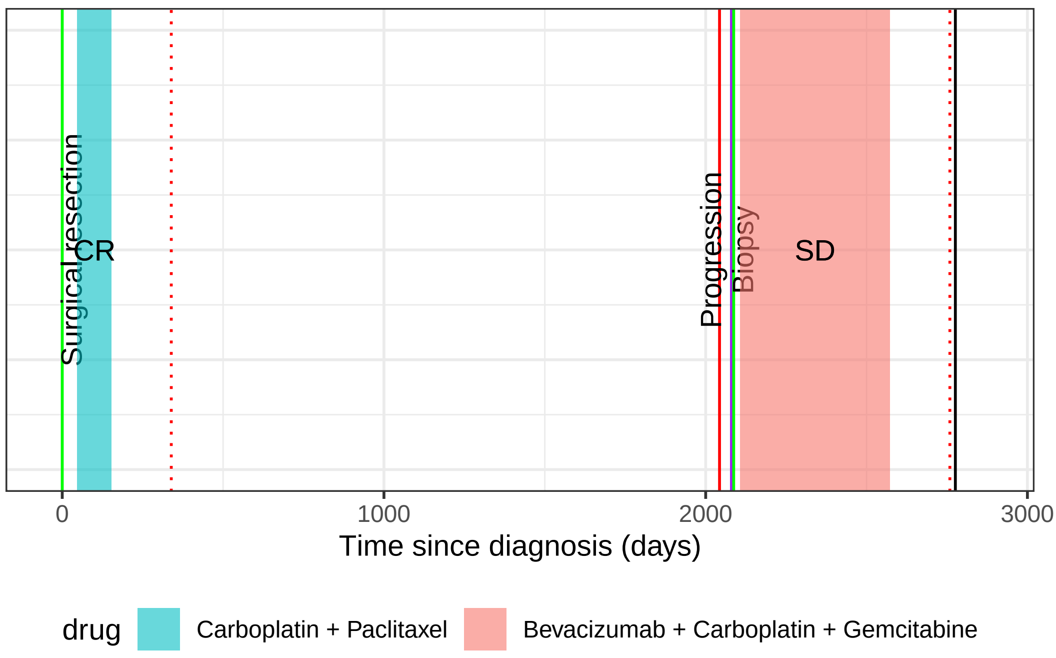

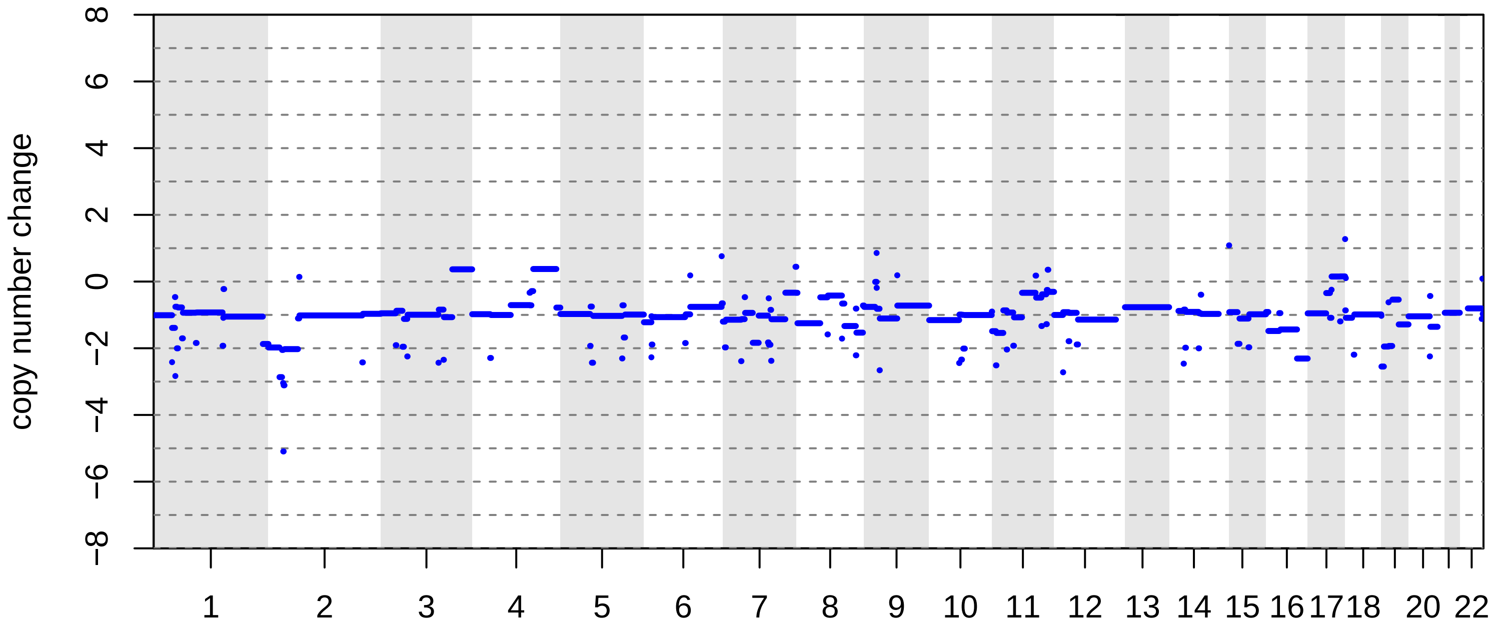

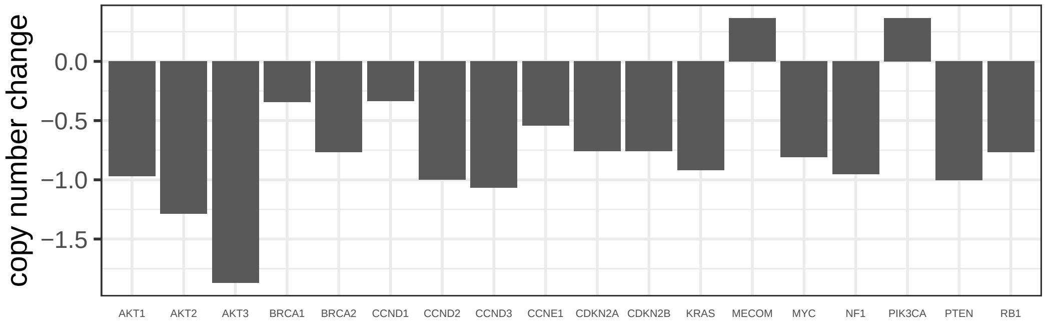

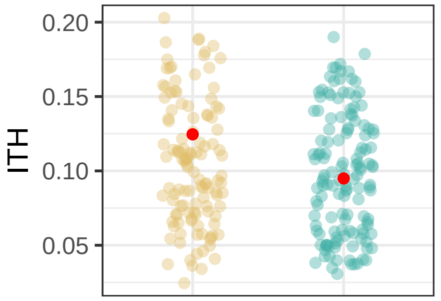

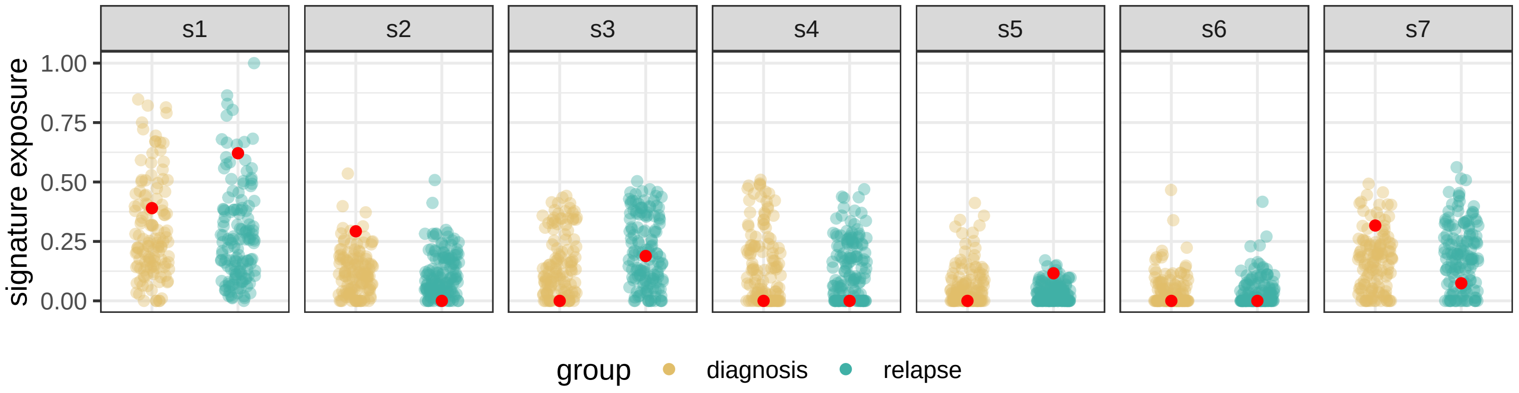

BRITROC−74

stage: 3C

platinum status: sensitive

prior lines: 1

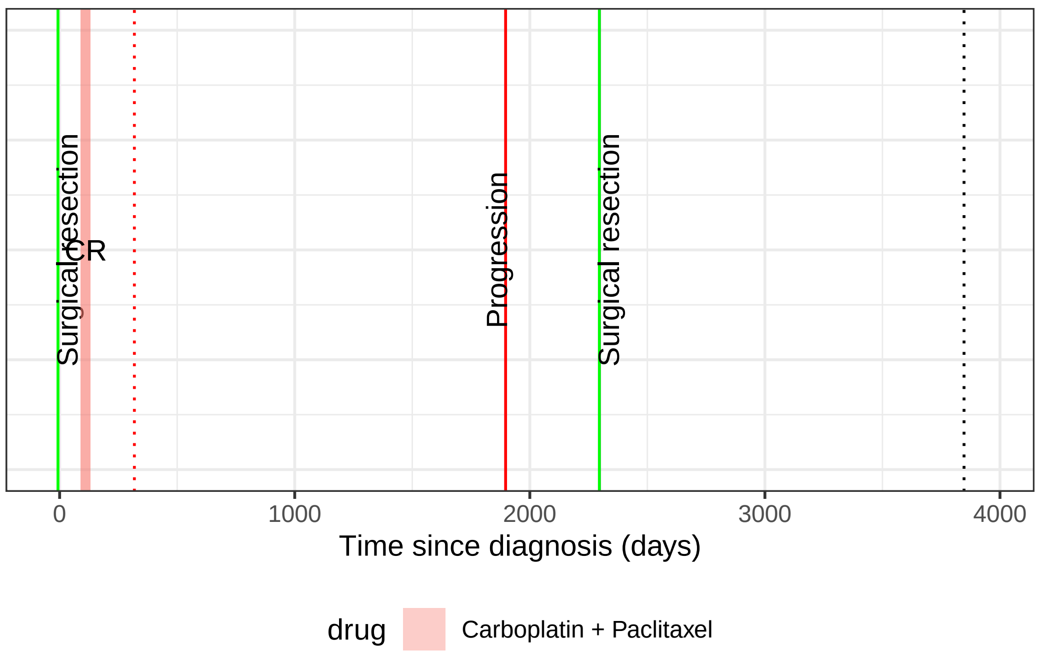

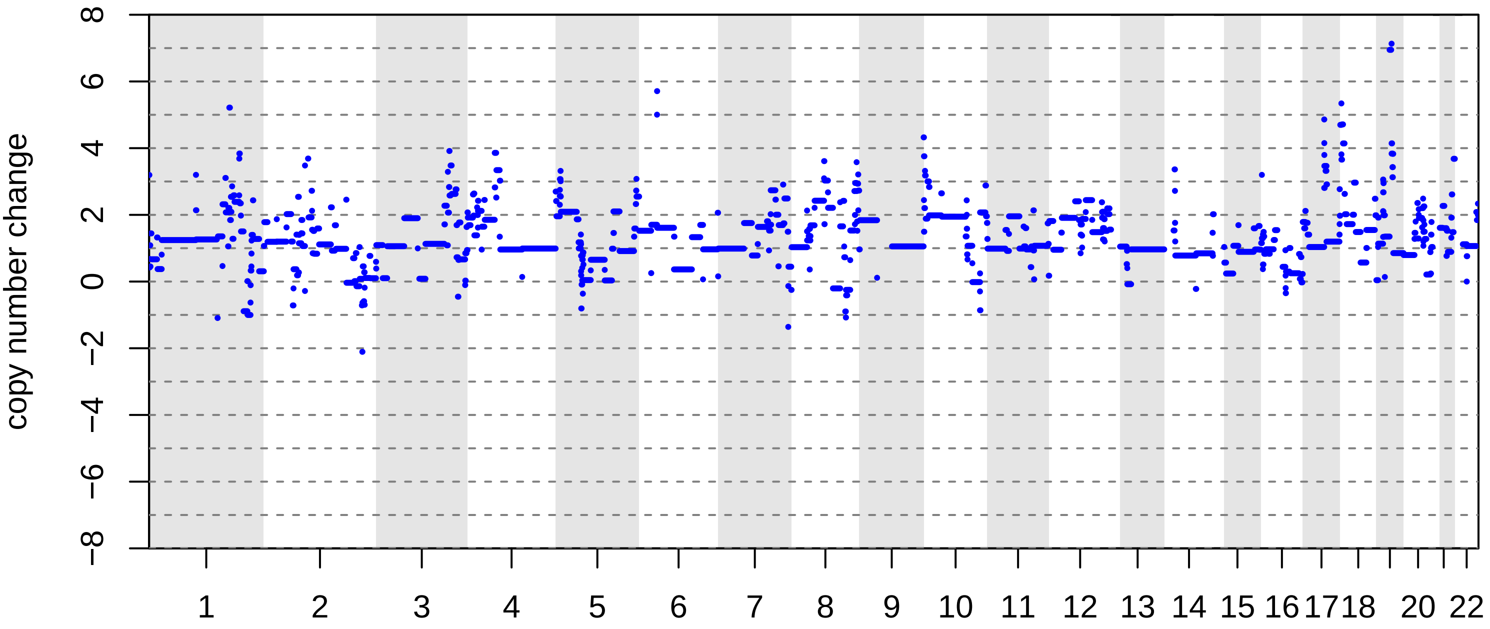

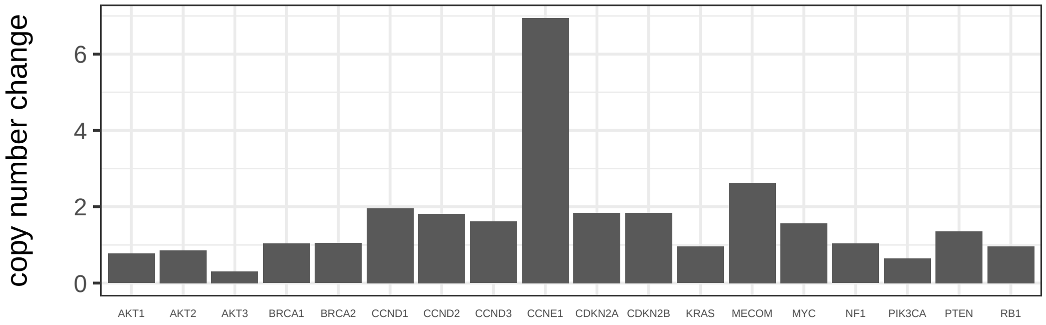

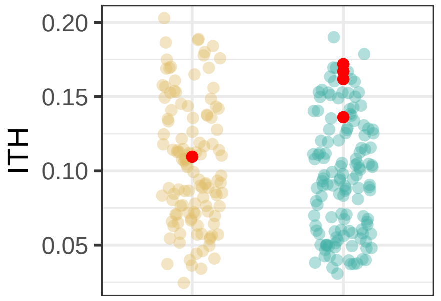

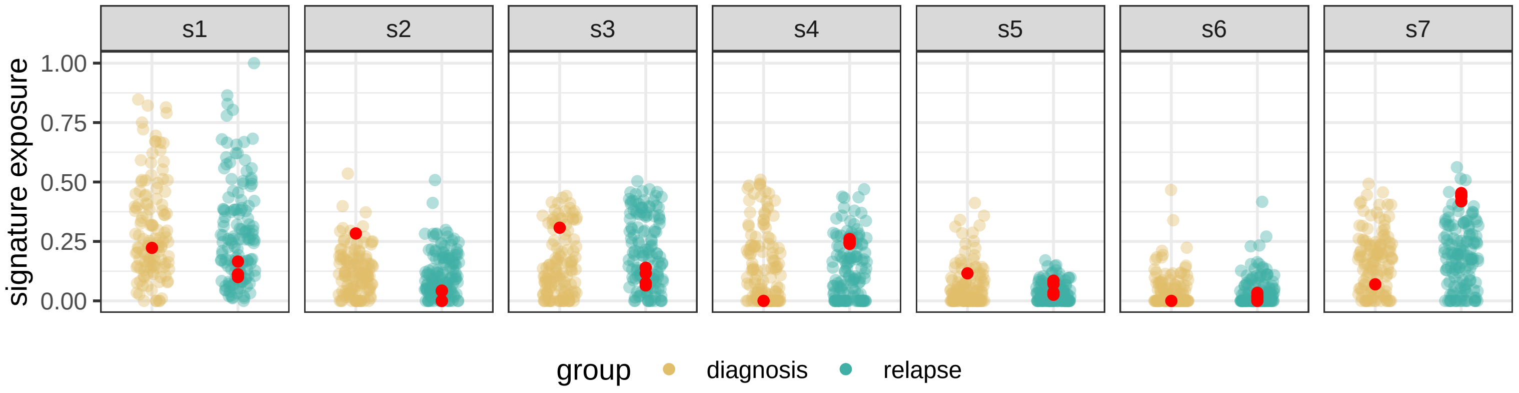

BRITROC−8

stage: 2B

platinum status: sensitive

prior lines: 1

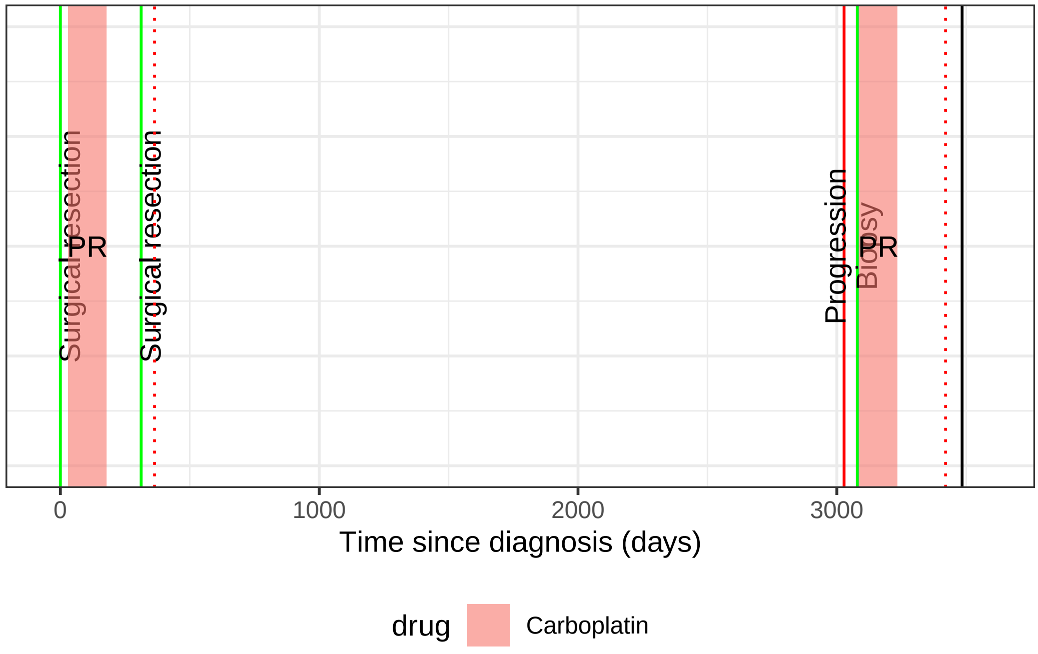

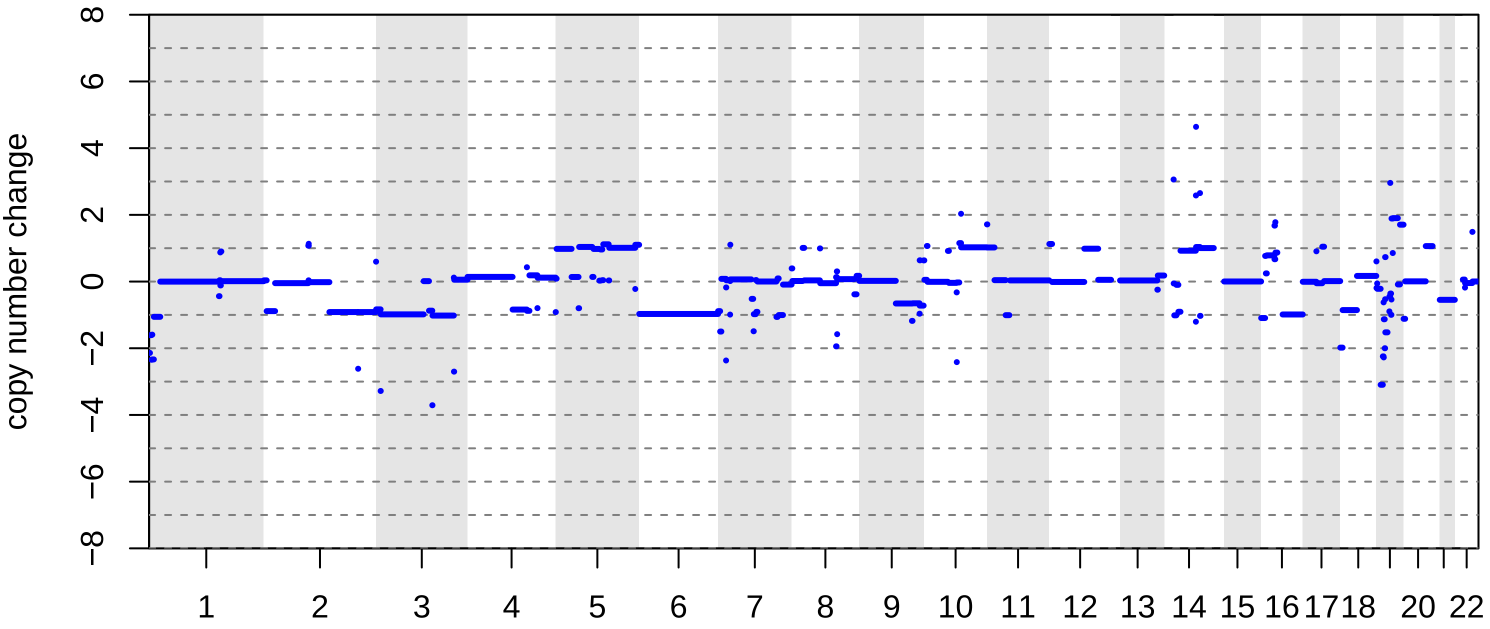

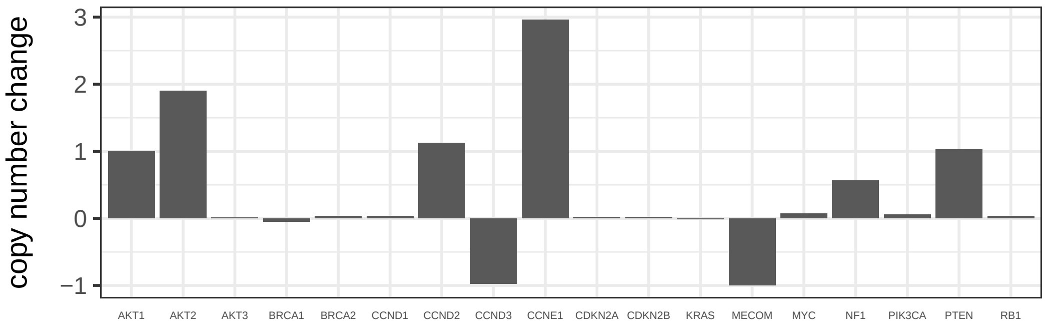

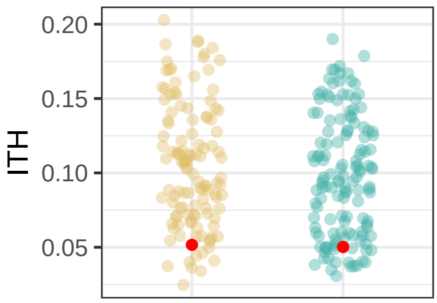

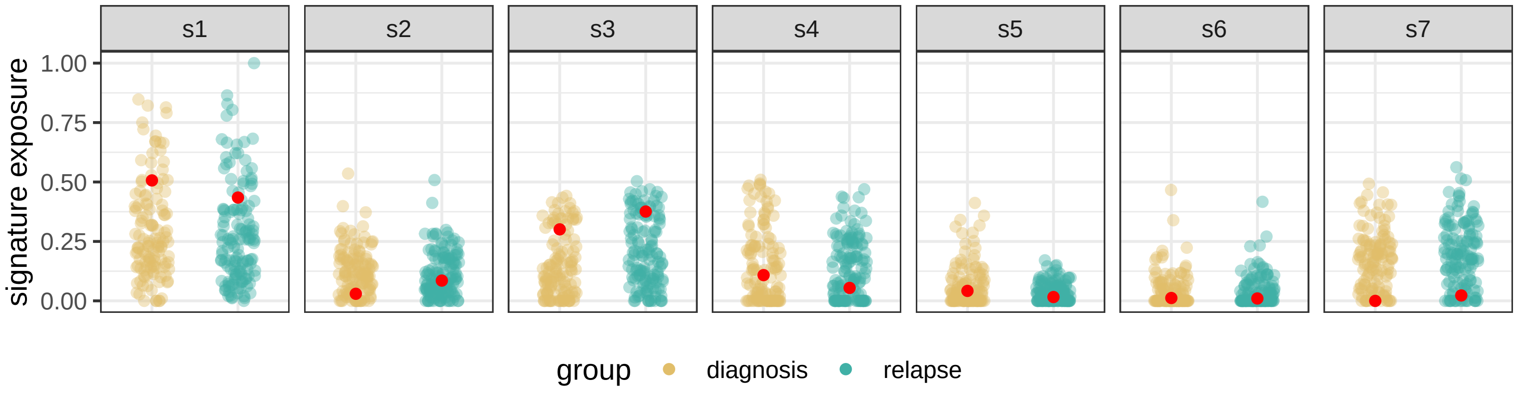

BRITROC−9

stage: 3C

platinum status: sensitive

prior lines: 1

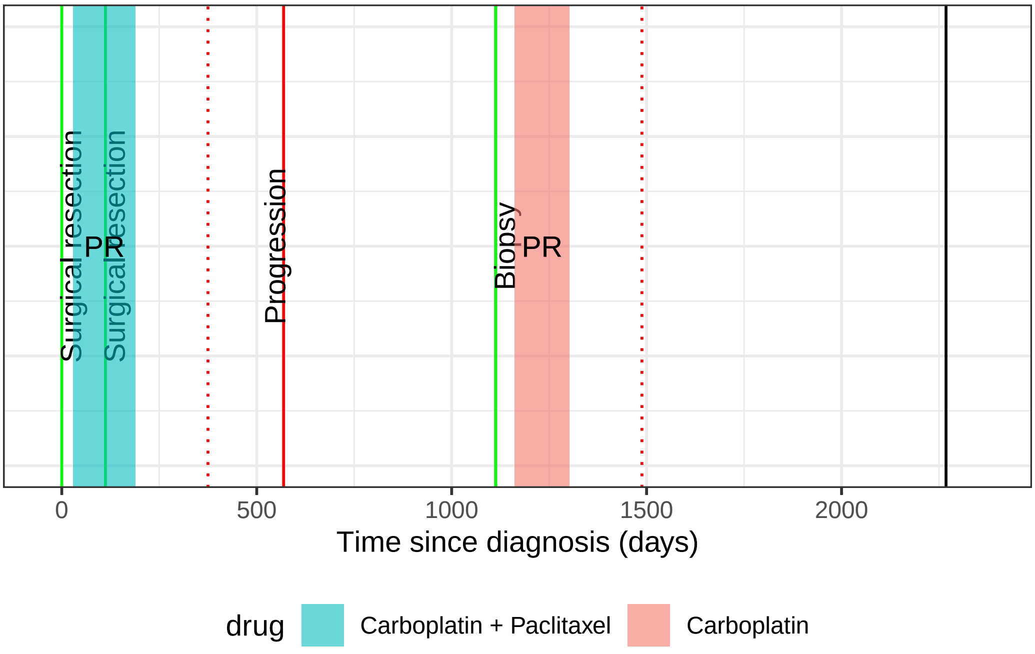

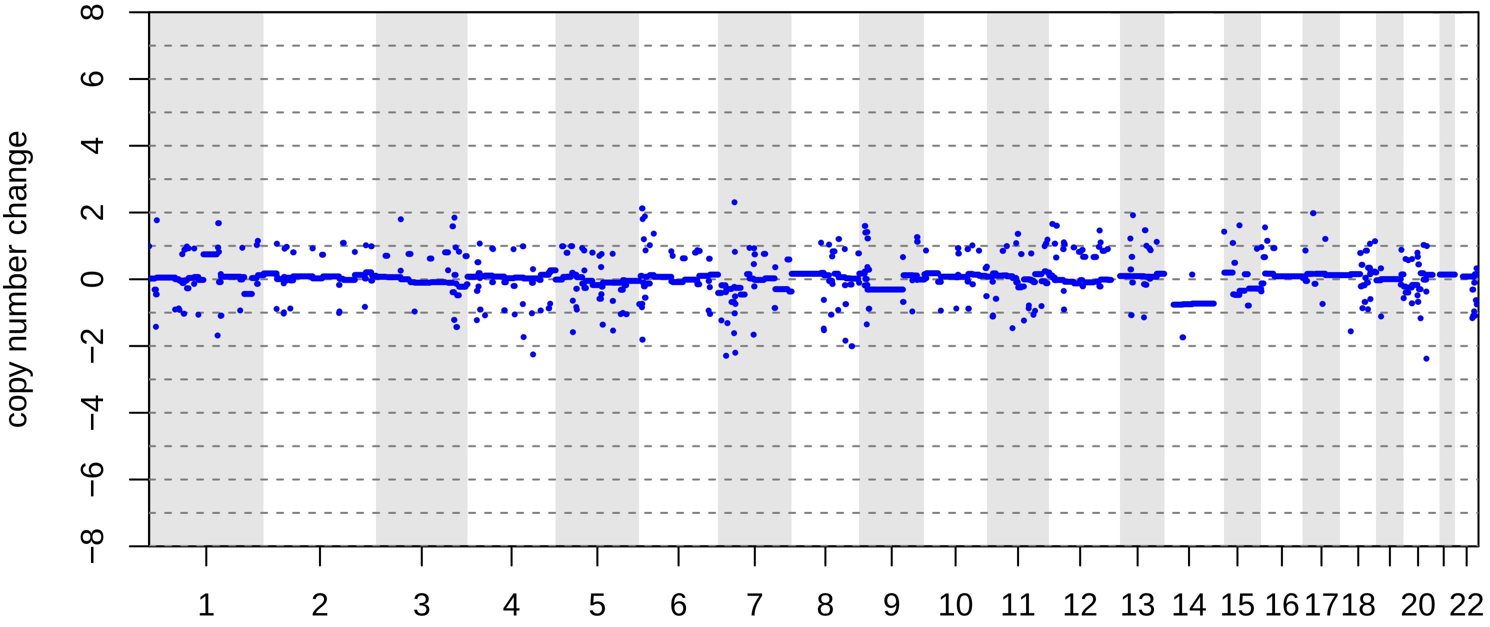

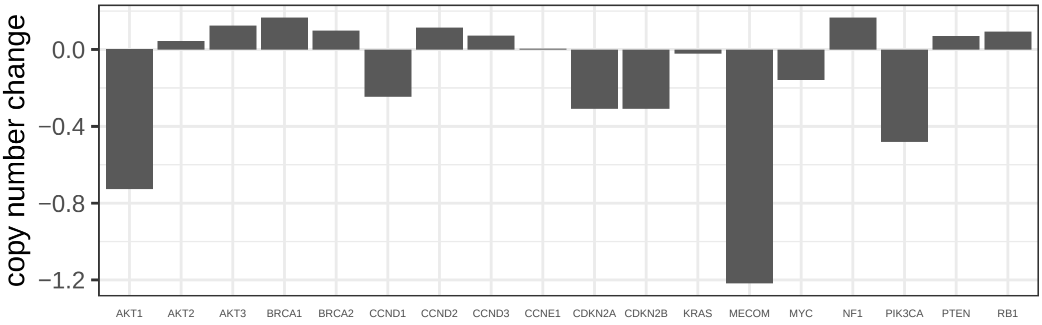

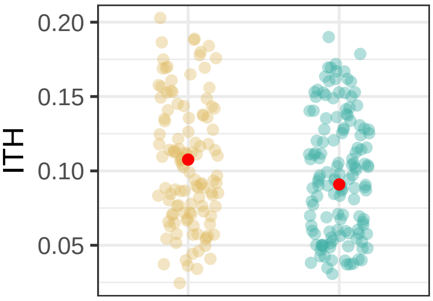

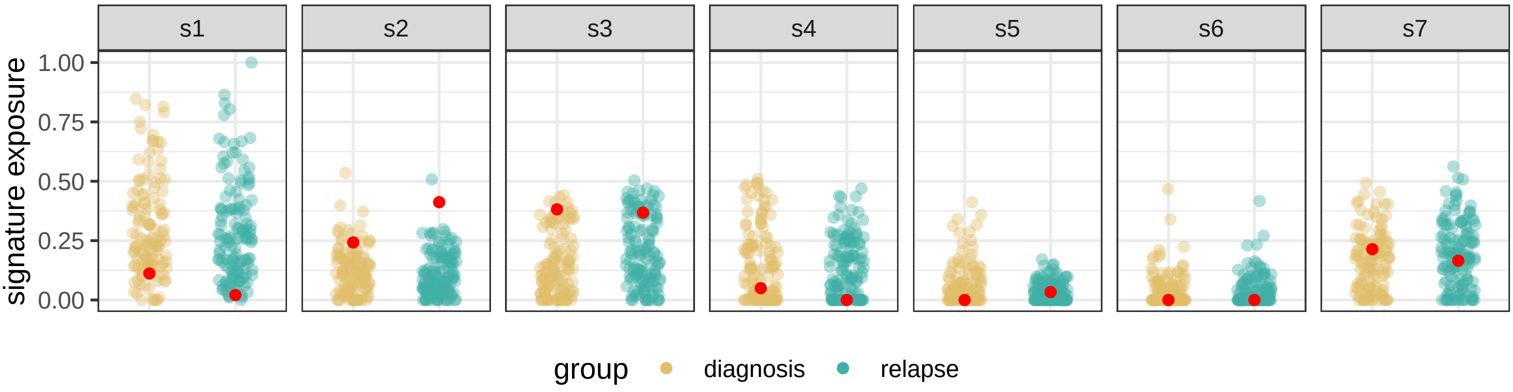

BRITROC−94

stage: 2B

platinum status: sensitive

prior lines: 2

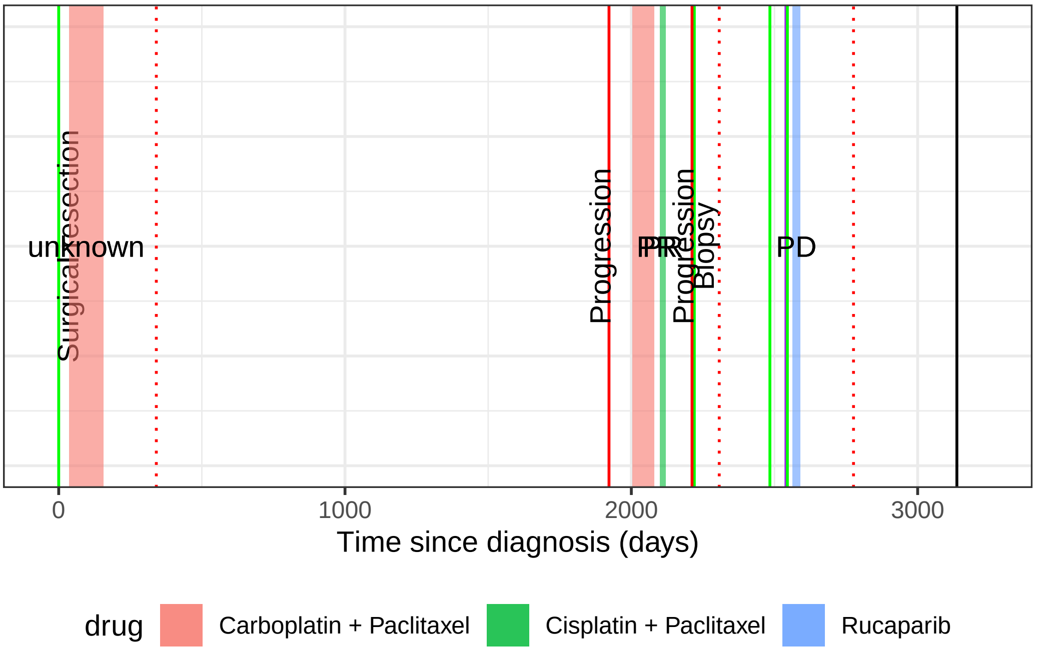

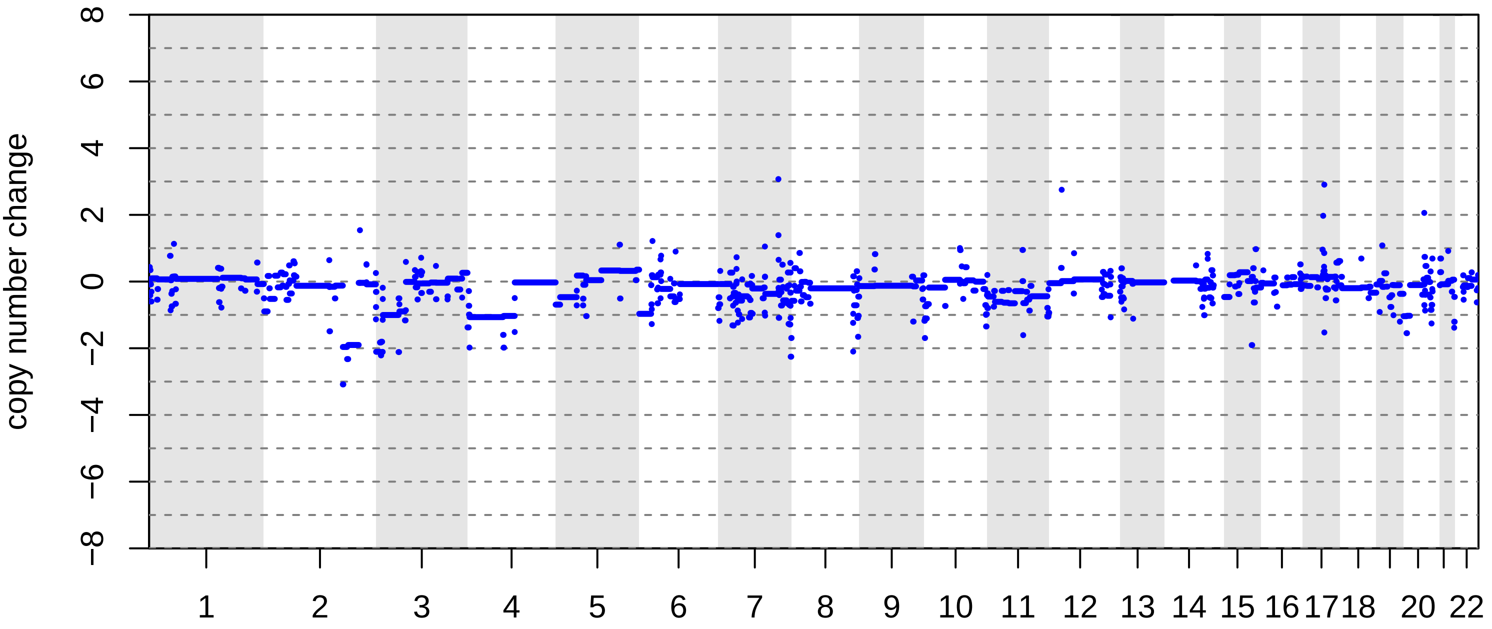

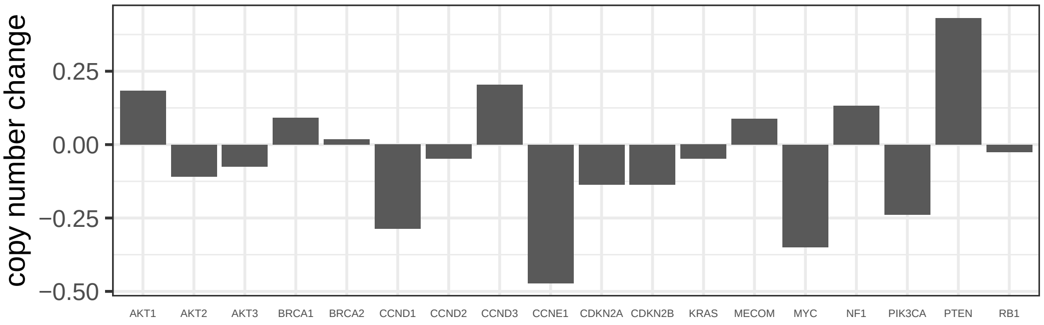

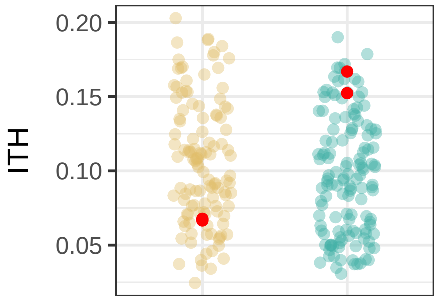

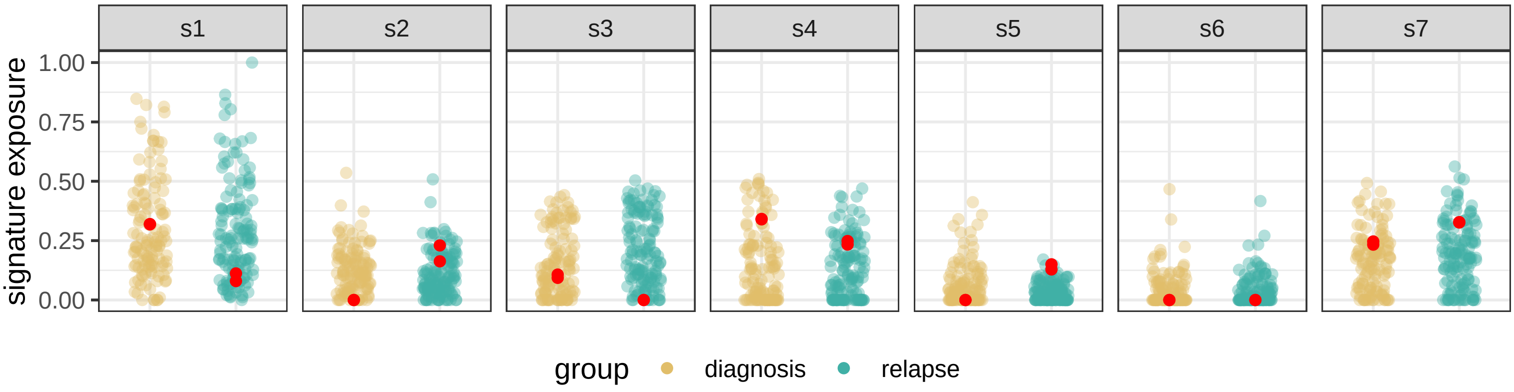

BRITROC−96

stage: 3B

platinum status: resistant

prior lines: 4

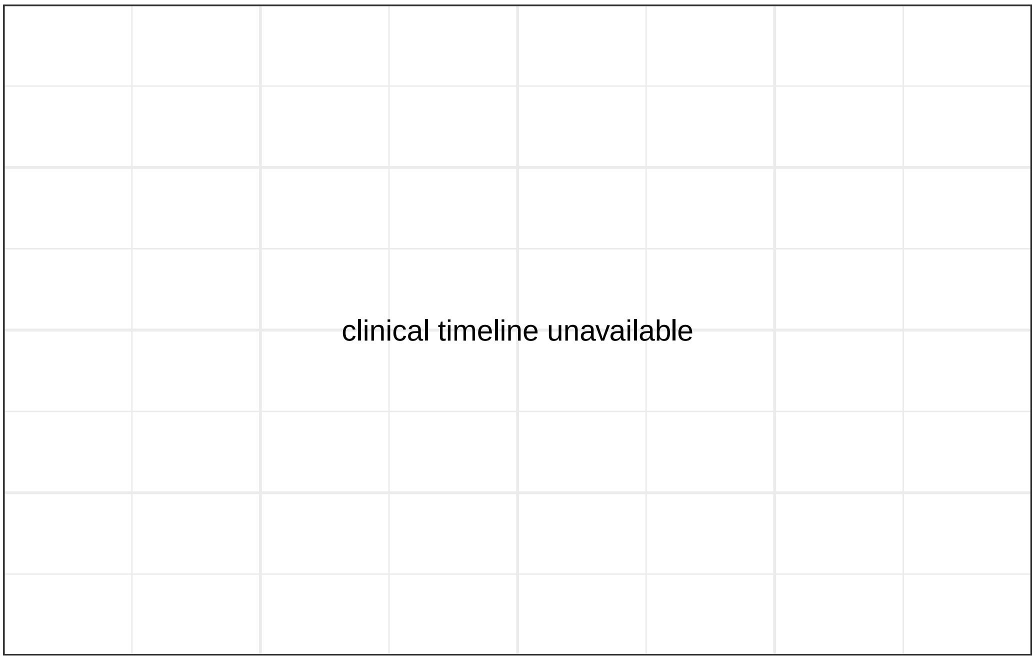

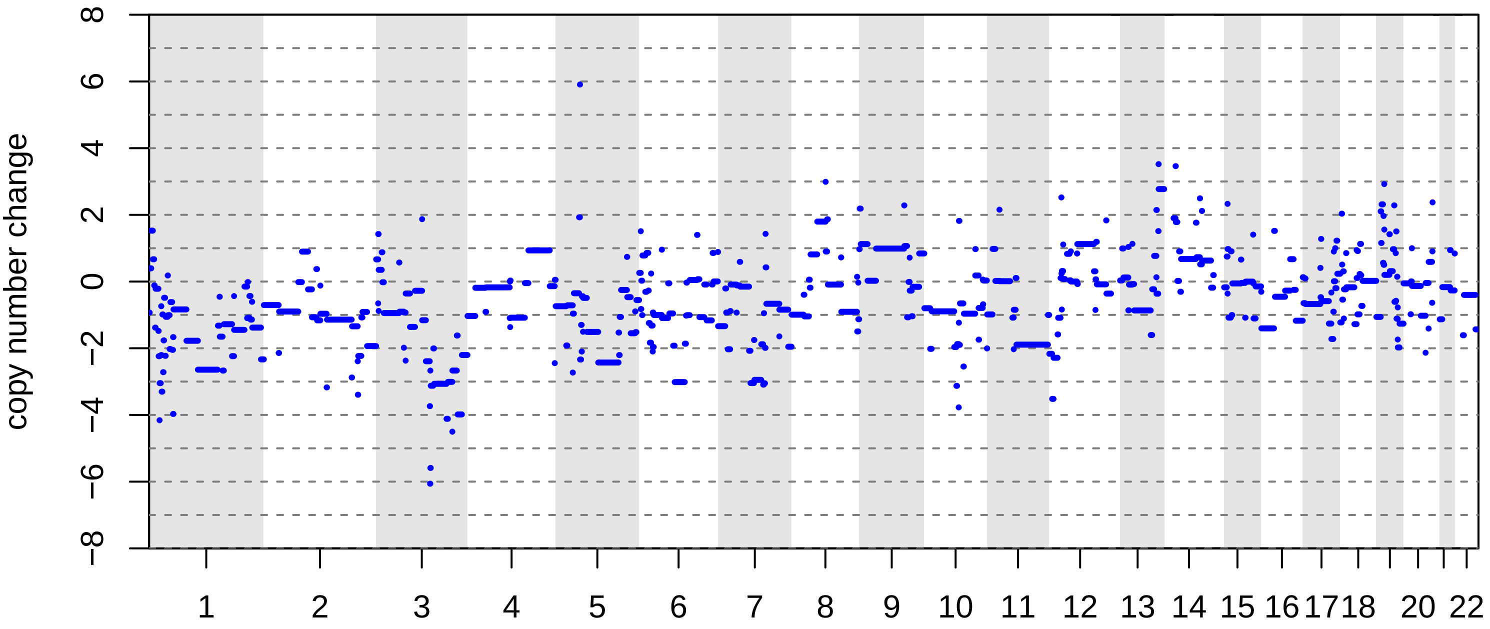

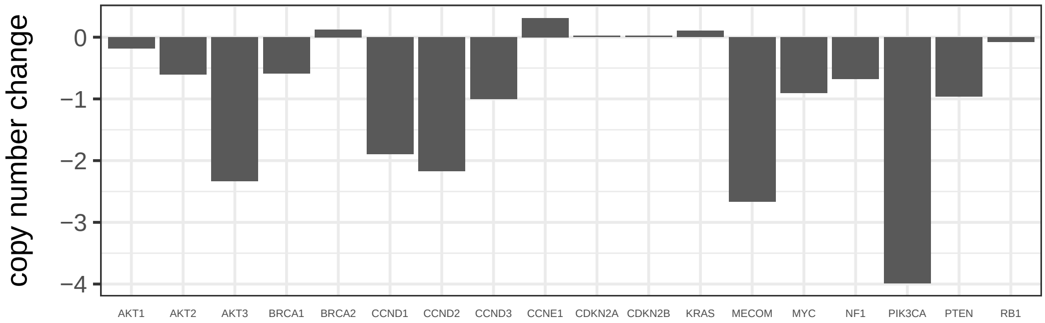

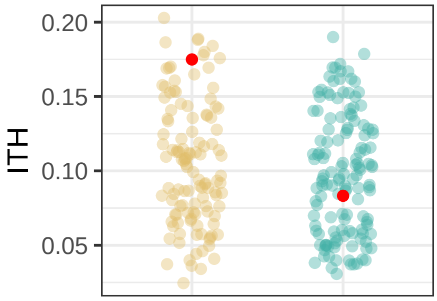

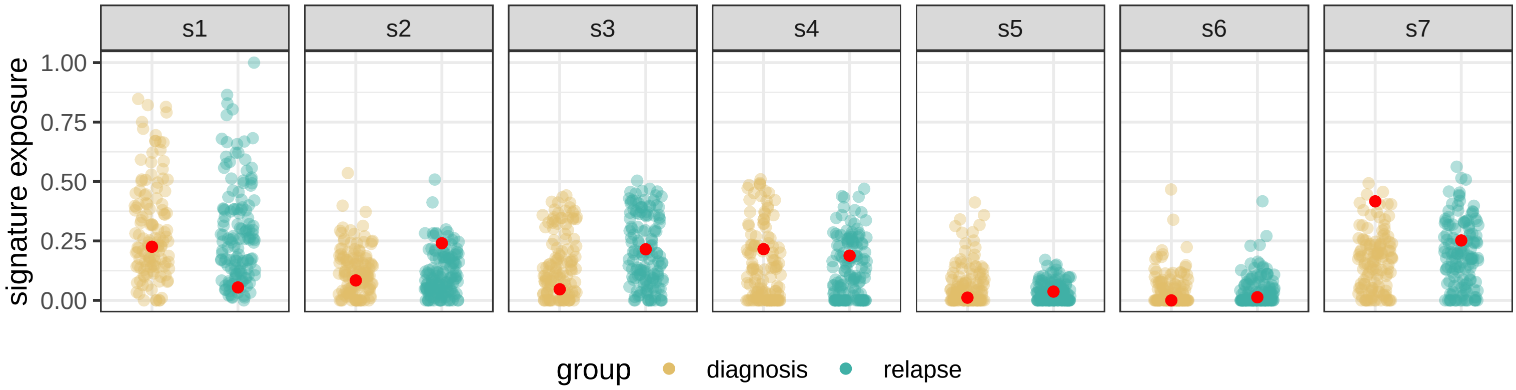

#### BRITROC−99

stage: 4NA

platinum status: sensitive

prior lines: 1

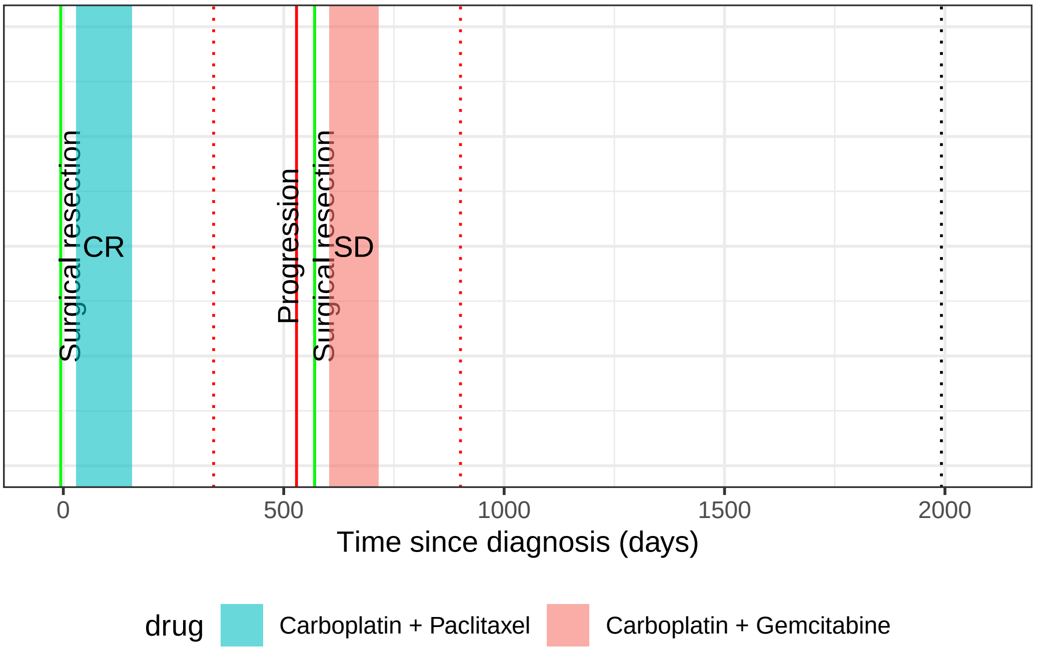

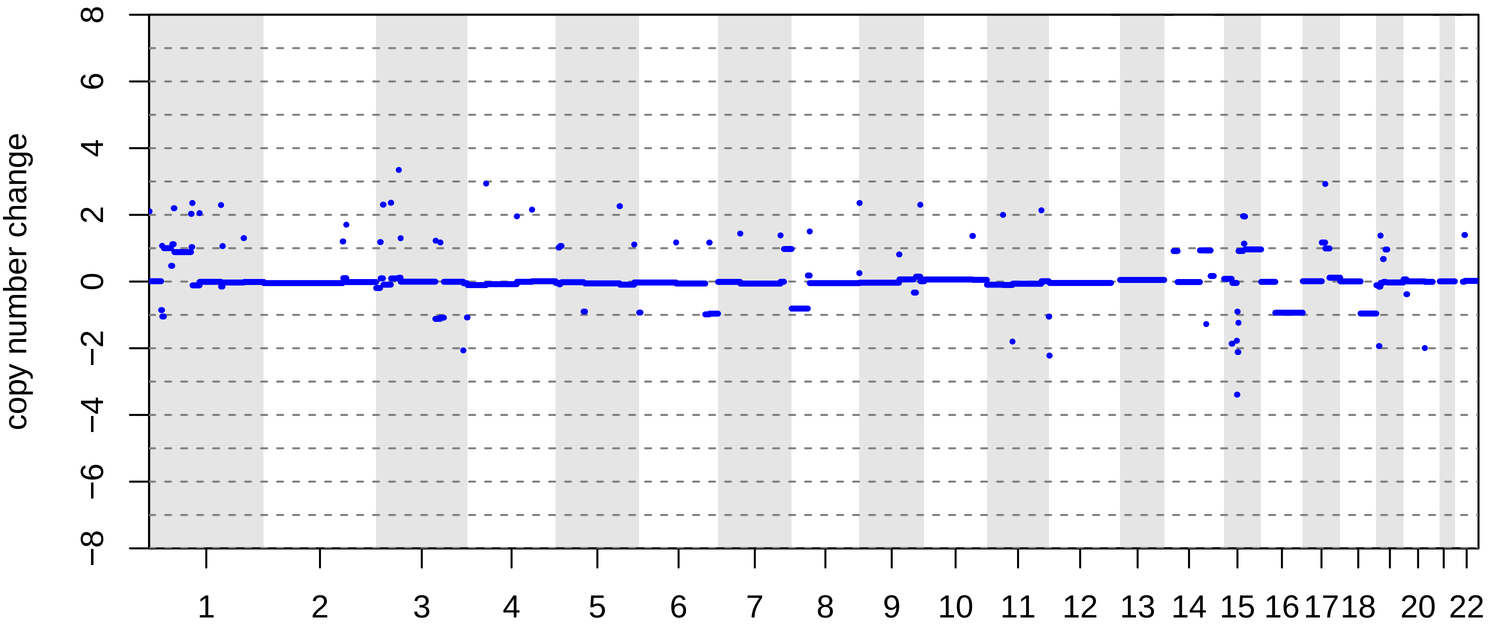

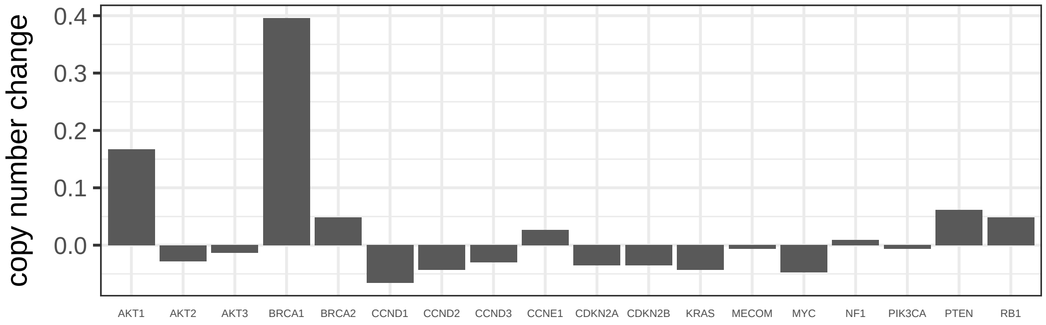

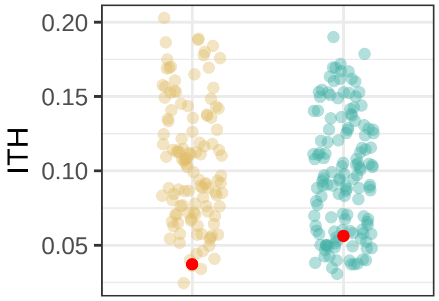

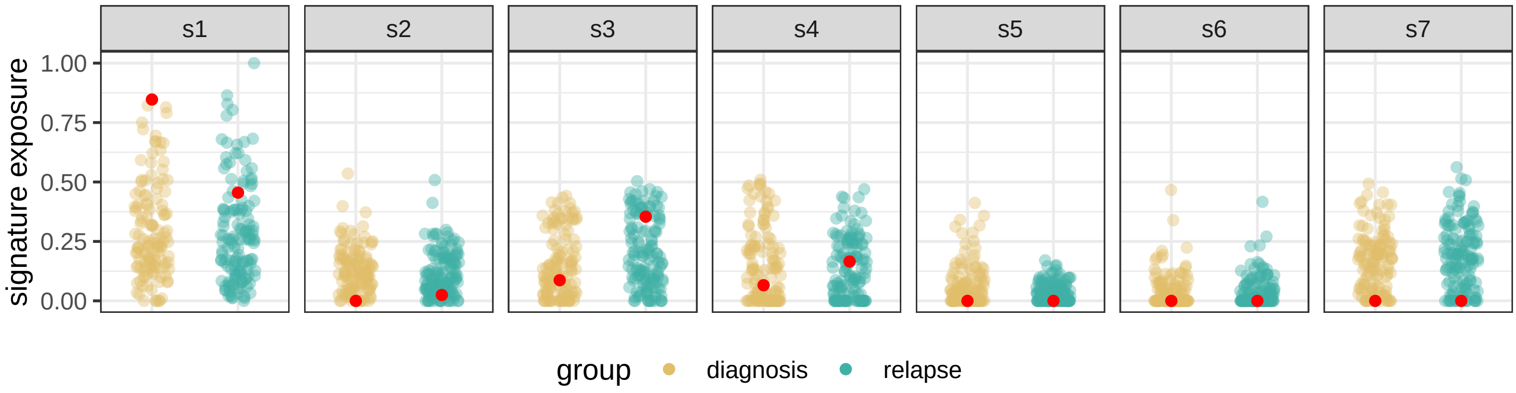
